# Label-free, low-cost AI ranking for systematic review screening

**DOI:** 10.64898/2026.10.09.26365173

**Authors:** Taeho Jo, Donghyun Lee, Leonard Lee

## Abstract

Title and abstract screening is among the most labor-intensive steps of systematic reviews. Jev, a low-cost AI decision model, scores records against eligibility criteria without screening labels. We scored 169,982 records from 128 reviews and, after tuning on development reviews, evaluated 23 held-out reviews once and 31 Cochrane reviews post hoc. Jev outranked the best free ranker in all 23 held-out reviews (mean AUC 0.966 vs 0.831) and came within 0.011 AUC of Claude Opus 5.5 on 1999 records at about 1/700 of its list-price cost (US$0.02/1000 records). Combined with active learning, it raised work saved at 95% recall from 0.659 to 0.806. A fixed score threshold fell below 95% recall in 3 of 28 Cochrane reviews, whereas Jev ranking with statistical stopping (post hoc) reached 95% recall in all 51 reviews, reading 86–88% of records. Label-free ranking with Jev was accurate and inexpensive, but stopping safely required screening decisions.

## Introduction

Systematic reviews underpin clinical guidelines but are slow to produce; an analysis of registered reviews estimated a mean of 67 weeks from registration to publication,^1^ and title and abstract screening is among the most labor-intensive steps. For two decades, machine learning has ranked records so that reviewers see likely eligible studies first.^2,3^ Active-learning tools such as ASReview retrain a classifier as reviewers label records,^4,5^ but they need labeled examples before they rank well and give no direct signal of when screening can safely stop. Stopping methods range from heuristics such as the knee method^6^ and practical procedures such as SAFE^7^ to statistical criteria^8^ and methods based on counting or point processes.^9,10^ Yet an evaluation of 15 stopping methods found that almost all either failed to stop reliably or stopped too late to save much work,^11^ so knowing when to stop remains a practical problem.

Generative large language models (LLMs) can judge eligibility from written criteria without review-specific training,^12–14^ and hybrid approaches have combined LLM outputs with active learning, one using LLM-derived pseudo-labels and one using fine-tuning on model disagreements.^15,16^ These approaches call a model for every record, so their cost grows with the number of records. Rankers that need no labels from the review include lexical and embedding rankers and neural cross-encoders, but in one evaluation, neural cross-encoders applied zero-shot were not competitive, whereas neural rankers fine-tuned on screening labels of other reviews were effective.^17^ A fine-tuned biomedical language model ensemble, which is non-generative, outperformed active learning and chatbot LLMs, but it was trained on almost 20,000 completed reviews; it reported a median area under the receiver operating characteristic curve (AUC) of 0.974 on its own test set and, with a threshold fixed on its validation split, retained 99.8% of included studies in 22 external reviews.^18^ These values come from other collections and label levels and are not comparable with ours. Workflows that route uncertain records to humans have also been proposed.^19^ Few studies of LLM screening address the steps that determine how many records must be read: of 290 PubMed records on LLMs in systematic review screening, only 32 (11%) also mentioned active learning, prioritization, or stopping in their title or abstract (Methods).

Decision models, a class of models that answer typed questions about a text with probabilities instead of generating text,^20^ offer another route. Jev, the decision model evaluated here, is described by its vendor as inexpensive and as returning calibrated probabilities.^21^ Such a model could score every record of a search cheaply and rank the records before any has been labeled, so it could be a starting point for both prioritization and stopping. Two concurrent reports have evaluated Jev: one against generative LLMs on the records of two systematic reviews,^22^ and one with a fixed label-free exclusion threshold developed on SYNERGY+ and evaluated on CLEF collections.^23^ Neither compared Jev with free rankers, combined its scores with an active-learning tool, or applied a statistical stopping criterion to its ranking.

We evaluated Jev on 169,982 records from 128 systematic reviews. We asked whether it ranks records better than free rankers (criterion C1) and comparably with generative LLMs (C2), whether adding its scores to an active-learning ranking reduces the records to be read (C3), and whether a single score threshold can serve as a stopping rule without labels (C4); these four criteria were fixed in advance (Methods). Post hoc, we also evaluated its label-free ranking followed by a statistical stopping criterion that tests recall within each review (label-free ranking with statistical stopping).

## Results

### Study overview

Fig. 1 shows the study design and Table 1 the review collections. Settings were chosen on 73 development reviews and on a pilot review by our group (Methods), and the 23 held-out reviews were then evaluated once against the four decision criteria (Table 2). Jev was compared with four free rankers, three generative LLMs, and active learning, and its ranking was combined with several stopping rules (Table 3). The external validation in 31 Cochrane reviews of CLEF 2019 and all other analyses marked post hoc are exploratory (Methods).

**Fig. 1.**
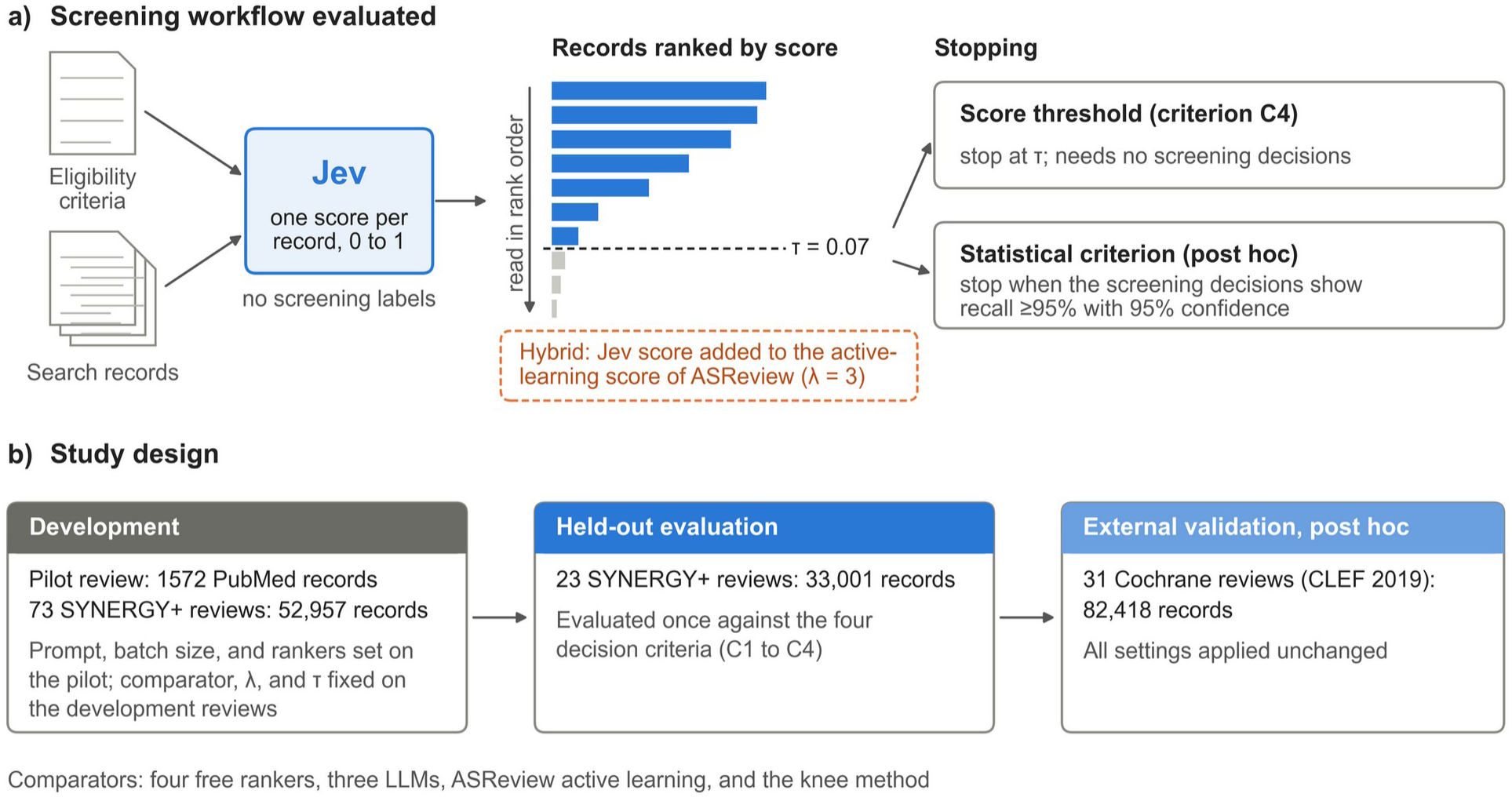
Screening workflow and study design. **a)** The workflow evaluated. Jev returns one score per record, between 0 and 1, for the question whether the record should be advanced to full-text screening, given the review title, research question, and eligibility criteria; ten records are sent per request. Records are ranked by the Jev score alone or by the Jev score added to the ranking score of the default ASReview model (hybrid, λ=3). Screening stops at a score threshold τ, which needs no screening decisions, or when the statistical criterion of Callaghan and Müller-Hansen, applied to the screening decisions, indicates recall of at least 95% with 95% confidence. In panel a, the ranked bars are illustrative; records scoring below τ (gray bars) are not read when screening stops at the threshold. **b)** Study design. The prompt, the batch size, and the shortlist of free rankers were set on a pilot review by our group, and the free-ranker comparator, λ, and τ on 73 development reviews of SYNERGY+; 23 held-out reviews were then evaluated once against four decision criteria (Methods, Study design). The external validation in Cochrane reviews and the statistical criterion are post hoc. SYNERGY+ counts refer to the subset exported by synergy-dataset 2.2 (open-access records with an abstract; Methods). LLM, large language model.

**Table 1.** Characteristics of the review collections.

| Collection | Reviews | Records | Positive records | Records per review, median (range) | Positive rate per review, median (range) | Use |
| --- | --- | --- | --- | --- | --- | --- |
| SYNERGY+ development | 73 | 52,957 | 2048 | 549 (54–1978) | 2.8% (0.5–23.8) | Choice of comparator, hybrid weight, and threshold |
| SYNERGY+ held-out | 23 | 33,001 | 597 | 840 (70–11,735) | 2.6% (0.3–12.9) | Evaluation against the decision criteria |
| CLEF 2019 Cochrane reviews | 31* | 82,418 | 882† | 1922 (61–9729) | 1.2% (0.0–19.0) | Post-hoc external validation |
| Alzheimer's disease genetics review (pilot) | 1 | 1572‡ | 143§ | .. | 9.1%§ | Prompt, batch size, and free-ranker shortlist; post-hoc analyses |

SYNERGY+ reviews span medicine, psychology, and computer science, and their positive records are finally included studies. The SYNERGY+ rows describe the subset exported by synergy-dataset 2.2, which keeps open-access records with an abstract (35.3% of the records and 41.2% of the included studies of the release for the 114 reviews; Methods). \*Eight diagnostic test accuracy, 20 intervention, one prognosis, and two qualitative reviews; three reviews had no content-relevant record among retrievable records and were used only with title and abstract labels; 6432 records (7.8%) had no abstract in PubMed and were scored from the title. †Records labeled relevant at the CLEF content (full-text) level. ‡PubMed arm of the search only, re-executed with the original search date as the end date (Methods). §Records passing title and abstract screening; 111 studies were finally included from all databases, 76 of them in the PubMed arm.

**Table 2.** Outcomes on the 23 held-out reviews against the decision criteria.

| Criterion | Comparison | Estimate (95% CI) | Reviews favoring Jev or the hybrid | Threshold | Met |
| --- | --- | --- | --- | --- | --- |
| C1 | AUC, Jev vs. bge-base (0.966 vs. 0.831) | 0.135 (0.098 to 0.173) | 23 of 23 | $\geq 0.03$, $P < 0.01$ | Yes |
| C2 | AUC, Jev vs. GPT-4o-mini (0.962 vs. 0.937) | 0.025 (0.010 to 0.040) | 16 of 23 (3 tied)<sup>¶</sup> | lower CI $> -0.02$ | Yes |
| C2 | AUC, Claude Opus 5.5 vs. Jev (0.973 vs. 0.962) | 0.011 (0.005 to 0.019) | 2 of 23 (7 tied)<sup>¶</sup> | $\leq 0.03$ | Yes |
| C3 | WSS@95, hybrid vs. ASReview (0.806 vs. 0.659) | 0.147 (0.089 to 0.213) | 21 of 23 | $\geq 0.05$, $P < 0.05$ | Yes |
| C3 | WSS@100, hybrid vs. ASReview (0.821 vs. 0.639) | 0.182 (0.115 to 0.258)<sup>¶</sup> | 21 of 23<sup>¶</sup> | $> 0$ | Yes |
| C4 | Reliability, Jev threshold vs. knee method as adapted (evaluated from 150 screened records) | 23 of 23 (85.7 to 100%)<sup>¶</sup> vs. 20 of 23 (67.9 to 95.5%)<sup>¶</sup> | .. | $\geq 90\%$ and ≥ knee | Yes |
| C4 | Records read, Jev threshold vs. knee method as adapted | 58.9% (49.0 to 69.0)<sup>¶</sup> vs. 94.0% (88.2 to 98.3)<sup>¶</sup> | 22 of 23<sup>¶</sup> | less than knee | Yes |
| Robustness (12 development reviews) | Within-review $\rho$ and AUC difference, two rephrased questions | $\rho$ 0.959 and 0.977; AUC difference 0.001 and 0.001 | .. | $\geq 0.90$ and AUC difference $\leq 0.02$ | Yes |

C2 was assessed on 1999 records scored by all models. The robustness check used 12 development reviews (5550 records), not the held-out reviews. CIs for differences and means are percentile bootstrap intervals over reviews, and CIs for reliability are two-sided Wilson intervals. Reliability in C4 was judged on the point estimate. C3 and C4 used the default ASReview model in place of the logistic-regression active learning named in the plan; with the planned comparators, the decisions were the same (Supplementary Note 5). The $P$ values of C1 and of the WSS@95 comparison of C3 are confirmatory; no test was specified for C2, C4, or the WSS@100 part of C3. <sup>¶</sup>Computed post hoc. AUC, area under the receiver operating characteristic curve; WSS@95 and WSS@100, work saved over sampling at 95% and 100% recall.

**Table 3.** Stopping options in the held-out and Cochrane reviews.

| Stopping option | Labels used | Statistical check of recall within the review | Held-out reviews: reliable; records read | Cochrane reviews: reliable; records read |
| --- | --- | --- | --- | --- |
| Threshold $\tau=0.07$ on the Jev score | No | No | 23 of 23; 58.9% | 25 of 28; 39.1% |
| Knee method as adapted (from 150 screened records), default ASReview ranking | Yes | No | 20 of 23; 94.0% | 26 of 28; 76.0% |
| Knee method with the published minimum of 1000 screened records, default ASReview ranking | Yes | No | 23 of 23; 97.4% | 28 of 28; 82.2% |
| Label-free ranking with statistical stopping (Jev ranking plus statistical criterion) | Yes | Yes | 23 of 23; 87.6% | 28 of 28; 86.1% |
| Default ASReview ranking plus statistical criterion | Yes | Yes | 23 of 23; 90.1% | 28 of 28; 86.9% |
| Threshold plus random-sample check below $\tau$ | Yes (records read at or above $\tau$ and the sample below $\tau$) | Yes | 23 of 23; 92.5% | 28 of 28; 90.6% |

### Zero-shot ranking

On the 23 held-out reviews, the mean AUC was 0.966 for Jev and 0.831 for bge-base, the best of four free rankers evaluated; MiniLM reached 0.821, TF-IDF 0.717, and BM25 0.712 (Table 2 and Fig. 2a). Jev was higher in all 23 reviews, with a difference of 0.135 (95% CI 0.098–0.173; *P*<0.0001), which met criterion C1. A biomedical cross-encoder evaluated post hoc reached a mean AUC of 0.709, below bge-base (Supplementary Note 6).

**Fig. 2.**
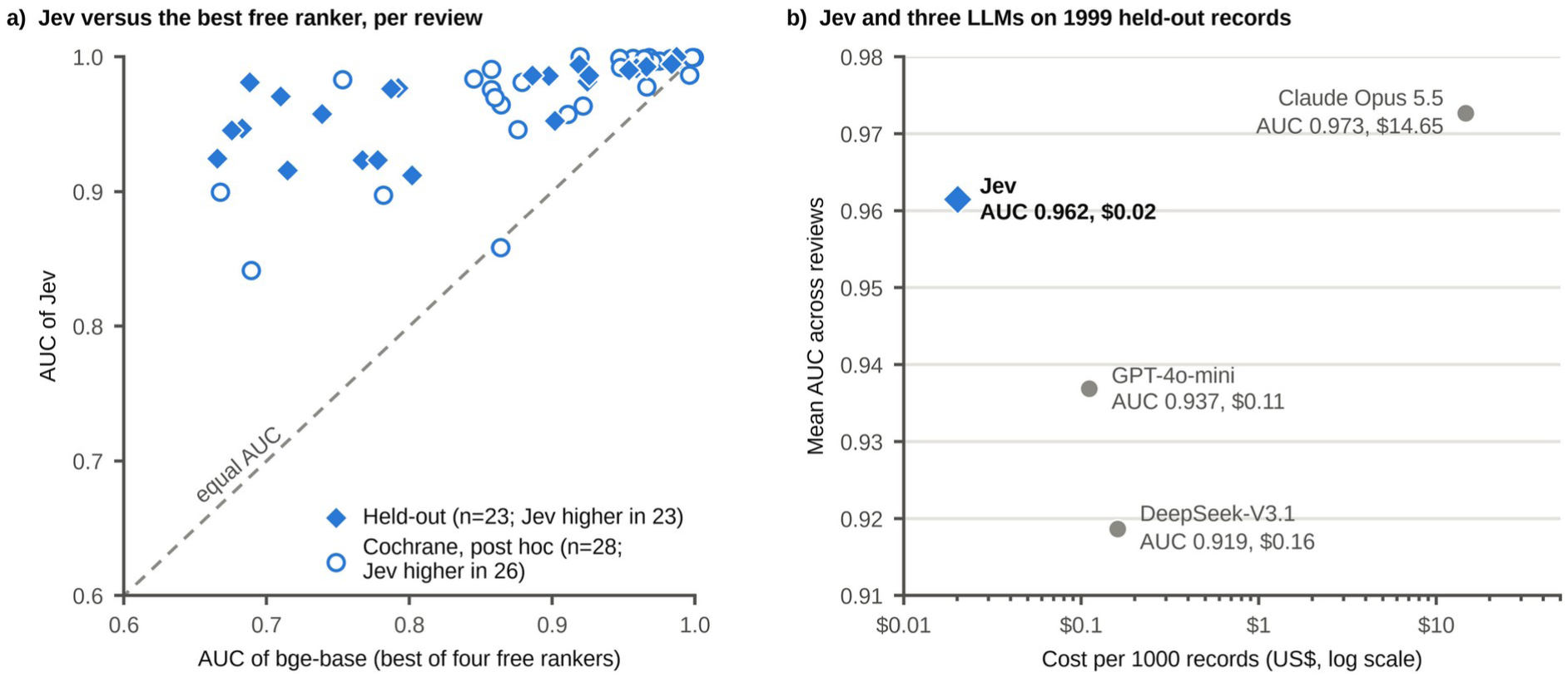
Discrimination and cost. **a)** Area under the receiver operating characteristic curve (AUC) of Jev against that of bge-base, the best of four free rankers evaluated, in each of the 23 held-out SYNERGY+ reviews (filled diamonds) and the 28 Cochrane reviews with at least one content-relevant record (open circles, post hoc). Points above the dashed line are reviews in which Jev ranked the records better. Mean differences and 95% CIs are given in the text and Table 2. **b)** Mean AUC across reviews and cost per 1000 records for Jev and three LLMs on the 1999 held-out records scored by all models. Costs are those reported by the gateway (for Claude Opus 5.5, the list-price equivalent reported by its command-line interface); the cost of Jev is for ten-record batches, and the costs of the LLMs are for single-record requests without batching or prompt caching. LLM, large language model.

### Comparison with generative LLMs

Of the 2002 held-out records selected for this comparison (Methods), 1999 were scored by all models; the requests to Claude Opus 5.5 for the other three were refused. The mean AUC was 0.962 for Jev, 0.973 for Claude Opus 5.5, 0.937 for GPT-4o-mini, and 0.919 for DeepSeek-V3.1 (Fig. 2b). Jev exceeded GPT-4o-mini by 0.025 (95% CI 0.010–0.040) and was 0.011 (0.005–0.019) below Claude Opus 5.5; these results met criterion C2. Post hoc, the deficit to Claude Opus 5.5 was also 0.011 with the three refused records scored 0 or 1, and 0.015 in the 16 reviews with at least 20 excluded records (Supplementary Note 15; Supplementary Tables 43 and 44).

Jev cost $0.020 per 1000 records in ten-record batches and $0.042 with single-record requests. With single-record requests, GPT-4o-mini cost $0.11, DeepSeek-V3.1 $0.16, and Claude Opus 5.5 $14.65 at list-price equivalent (Supplementary Note 2). Scoring all 33,001 held-out records with Jev cost $0.66, and each percentage point of them left unread corresponds to 330 records. At a reported rate of two abstracts per minute for an experienced reviewer,^24^ these 330 records would take about 2.75 h of one reviewer’s time.

### Prioritization and active learning

Work saved over sampling at 95% recall (WSS@95) is the proportion of reading saved, relative to random order, when 95% of the included studies have been found; WSS@100 is the same measure at 100% recall. The hybrid starts in Jev order and then adds the Jev score with weight λ=3 to ASReview’s ranking score. The mean WSS@95 was 0.659 for the default ASReview model and 0.806 for the hybrid, a gain of 0.147 (95% CI 0.089–0.213; *P*<0.0001), and the hybrid was higher in 21 of 23 reviews. With WSS@100 rising from 0.639 to 0.821, the hybrid met criterion C3 (Table 2 and Fig. 3a,c). In post-hoc analyses, the label-free Jev ranking alone reached 0.787, 0.128 (0.062– 0.200) above ASReview, and the hybrid added 0.019 (0.002–0.038) to it. With λ=2, the runner-up on the development reviews, this increment was 0.019 (-0.005 to 0.048; Supplementary Note 10). In the same analyses, the label-free Jev ranking also found included studies earlier than the default ASReview ranking, with an average time to discovery of 0.053 vs. 0.101 in held-out reviews (Fig. 3d; Supplementary Note 14). Post hoc, adding Jev scores at λ=3 to ASReview’s embedding-based elas_h3 model raised its WSS@95 from 0.724 to 0.806, a gain of 0.082 (95% CI 0.045–0.128), and weak supervision guided by Jev scores reached 0.746 (Supplementary Note 5).

**Fig. 3.**
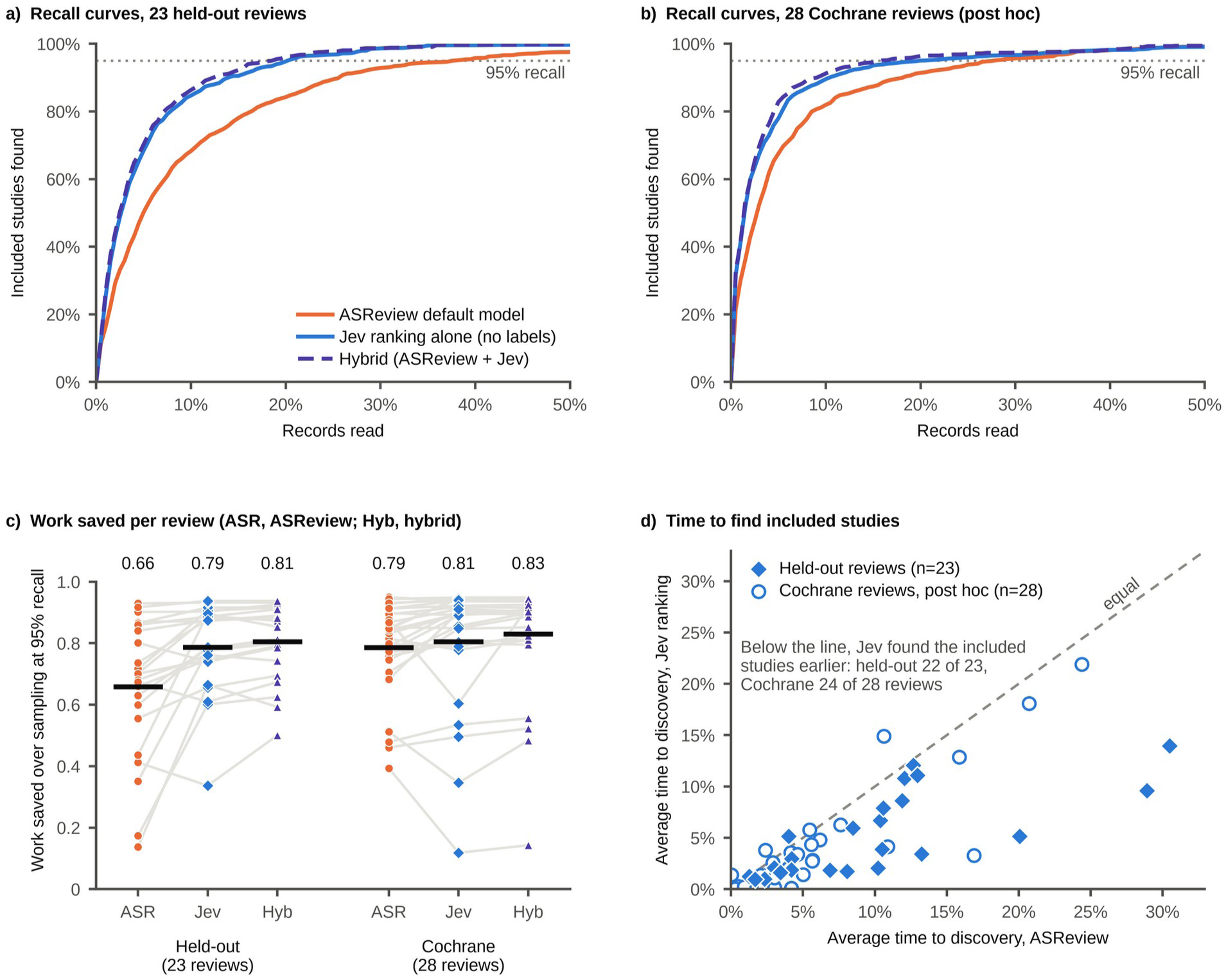
Prioritization. **a, b)** Proportion of included studies found, averaged over reviews, against the proportion of records read in the 23 held-out reviews (**a**) and the 28 Cochrane reviews (**b**, post hoc). The curves show the default ASReview model (mean of ten random starts), the label-free Jev ranking, and the hybrid; the horizontal axis ends at 50% of records. **c)** Work saved over sampling at 95% recall (WSS@95) per review; grey lines join the values of one review, and black bars and the numbers above them are means over reviews. **d)** Average time to discovery per review for the default ASReview model and the Jev ranking (post hoc). The average time to discovery is the mean, over included studies, of the proportion of records read when the study is found; points below the dashed line are reviews in which the Jev ranking found the included studies earlier. Paired differences with 95% CIs are given in the Results, Table 2, and Supplementary Notes 10 and 14. ASR, ASReview default model; Hyb, hybrid.

### Stopping

The label-free threshold τ=0.07 was chosen on the development reviews; records with a Jev score below τ are not read. With this threshold, recall was at least 95% in all 23 held-out reviews, and 58.9% of records were read (Fig. 4a). The knee method as adapted, evaluated from 150 screened records, reached 95% recall in 20 of 23 reviews and read 94.0% of records; against it, the threshold met criterion C4. The threshold read fewer records than the knee method in 22 of 23 reviews. Its reliability, the proportion of reviews with recall of at least 95%, was higher on the point estimate, but the paired difference was not distinguishable from zero (Tango 95% CI -0.031 to 0.321), and the study could not show a reliability of at least 90%: the one-sided exact lower 95% confidence limit was 87.8%. The knee method with the published minimum of 1000 screened records, applied post hoc, was reliable in every held-out review and read 38.5 percentage points (28.9–48.3) more records than the threshold (Table 3). It read every record in the 13 held-out reviews with fewer than 1000 records and 93.9% of records in the ten larger ones, against 60.1% for the threshold (Supplementary Table 24).

**Fig. 4.**
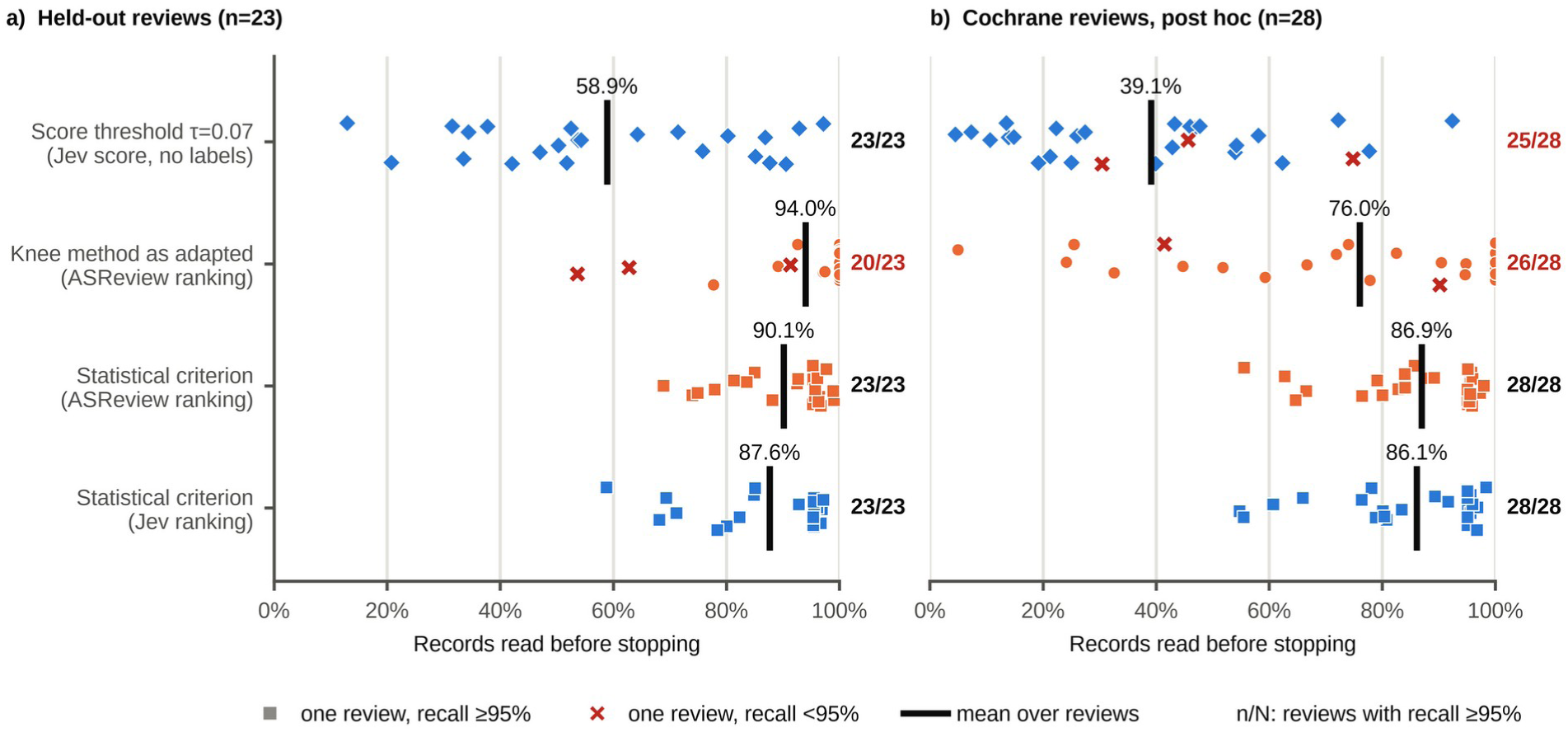
Stopping rules. Proportion of records read before stopping in each review, in the 23 held-out reviews (**a**) and the 28 Cochrane reviews (**b**, post hoc). The rules are the label-free score threshold (τ=0.07), the knee method as adapted (evaluated from 150 screened records) on the ASReview ranking, and the statistical criterion of Callaghan and Müller-Hansen (recall target 0.95, confidence 0.95) on the ASReview ranking and on the Jev ranking. The statistical criterion was evaluated post hoc in both collections. Each marker is one review; crosses mark reviews in which recall at the stopping point was below 95%. Rules applied to rankings with ten random starts or tie-breaking seeds are summarized by the mean over starts or seeds. Markers are spread vertically for legibility only. Black bars and the percentages above them are macro-averaged proportions read, and the numbers on the right are the reviews with recall of at least 95%. Confidence intervals are given in Supplementary Table 23.

Post hoc, in bootstrap resamples of the development reviews, the rule that selects τ (Methods) chose 0.07 in 84.4% of them. When τ was re-selected in each of a further 5000 resamples and applied to the development reviews left out of that resample, the mean reliability was 0.969, but it fell below 0.90 in 13.5% of resamples (Supplementary Note 12). Among the 2002 held-out records of the comparison with LLMs, single-record requests and ten-record batches formed from these records placed 143 of 1405 excluded records and one of 597 included studies on different sides of τ, although the numbers of excluded records at or above τ were similar, at 648 and 645. Recall at τ stayed at 95% or higher in all 23 reviews under both modes (Supplementary Table 35).

Post hoc, against title and abstract labels, the threshold kept recall at 95% or higher in seven of 12 held-out reviews (Fig. 5b). In these 12 reviews, the AUC of Jev was 0.903 against title and abstract labels and 0.962 against final inclusion (Supplementary Note 5). Of the 948 records that passed title and abstract screening in these reviews, 61 fell below τ, and one of them was finally included (Supplementary Table 38). Across development and held-out reviews, 12 of 2645 included studies (0.45%) fell below the threshold (Supplementary Note 7).

**Fig. 5.**
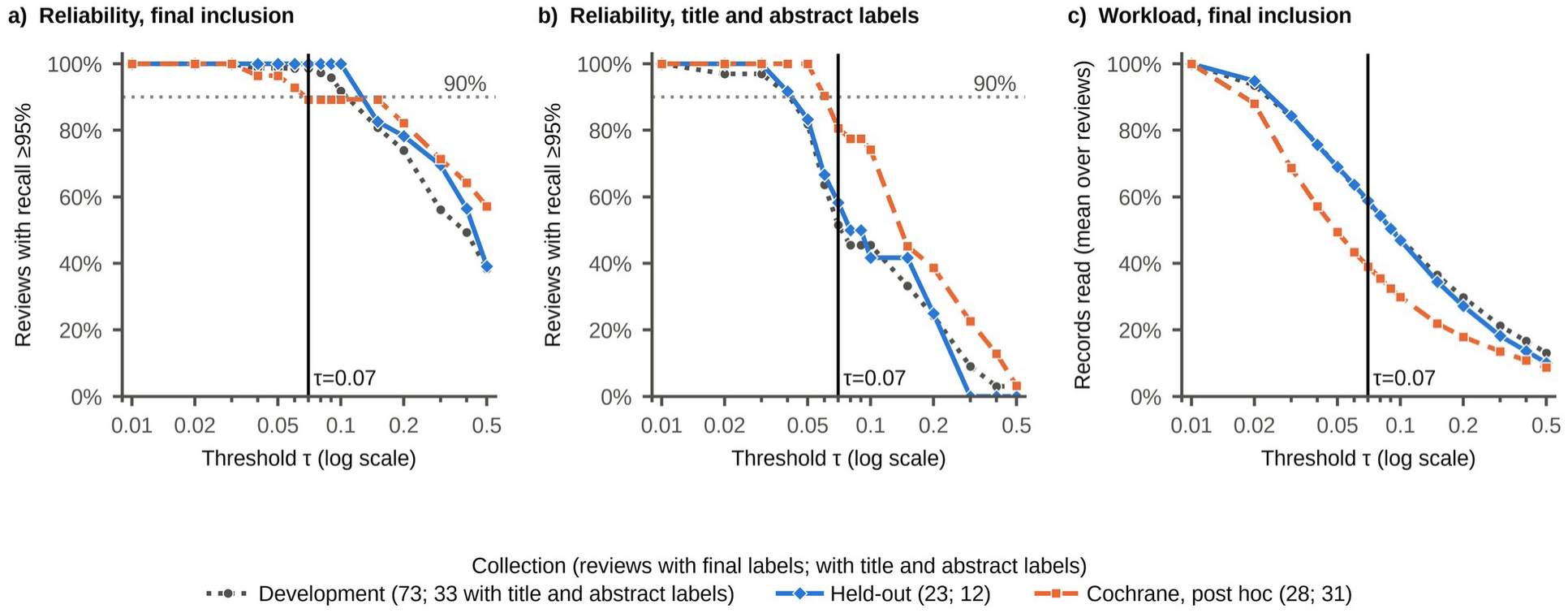
Threshold rule across thresholds (post hoc). **a, b)** Proportion of reviews in which the threshold rule kept recall at 95% or higher, against τ, for final inclusion (content-level labels in the Cochrane reviews; **a**) and for title and abstract labels (**b**), in the development, held-out, and Cochrane reviews. **c)** Macro-averaged proportion of records read against τ. The vertical line marks τ=0.07, chosen on the development reviews; the dotted horizontal line marks 90%. Wilson 95% CIs, pooled proportions read, and the lowest recall at each τ are shown in Supplementary Fig. 2.

### Calibration

Calibration of the scores was assessed post hoc and was poor (Supplementary Fig. 4). On the held-out reviews, the mean score was 0.140 and the observed inclusion rate 0.018, an observed-to-expected ratio of 0.13 (95% CI 0.08–0.22), with a recalibration slope of 1.30 (95% CI 1.11–1.65). The scores therefore overestimated the inclusion rate and were too narrowly spread on the logit scale. The ratio was 0.38 against title and abstract labels and 0.13 against the content-level labels of the Cochrane reviews (Supplementary Note 9).

### Robustness, controls, and other post-hoc analyses

On 12 development reviews, rephrasing the question changed the ranking little, with mean within-review Spearman ρ of 0.959 and 0.977, and changed the AUC by 0.001 with each rephrasing. With the review title alone, the AUC fell from 0.942 to 0.895, and with the criteria of a different review it fell to 0.601 (Supplementary Note 5). Post hoc, reviews that retained a larger share of their records in the open-access subset tended to have a larger proportion read at τ (ρ=0.40, *P*=0.058; in the 73 development reviews, ρ=0.32, *P*=0.0065) and a lower AUC of Jev (ρ=-0.36, *P*=0.096; development reviews, ρ=-0.22, *P*=0.066; Supplementary Table 8).

### Post-hoc external validation in Cochrane reviews

Of the 31 Cochrane reviews, 28 had at least one record judged relevant at the full-text level (content-relevant record; Methods). The mean AUC was 0.970 for Jev and 0.900 for bge-base, a difference of 0.070 (95% CI 0.047–0.095; Fig. 2a). WSS@95 was 0.786 for ASReview, 0.805 for the Jev ranking, and 0.830 for the hybrid (Fig. 3b,c). The difference between the Jev ranking and ASReview was 0.019 (95% CI -0.023 to 0.055; Supplementary Table 19).

The threshold kept recall at 95% or higher in 25 of 28 reviews, below the 90% target, and read 39.1% of records (Fig. 4b). Against title and abstract labels, it did so in 25 of 31 reviews (Wilson 95% CI 63.7–90.8%; Supplementary Table 36). The three reviews below 95% recall missed one of nine, three of 37, and one of ten content-relevant records. The knee method as adapted reached 95% recall in 26 of 28 reviews on the ASReview ranking, and the knee method with the published minimum in 28 of 28 (Table 3), so the reliability part of C4 was not reproduced; none of these differences was distinguishable from zero. Pooled over the held-out and Cochrane reviews, the threshold fell below 95% recall in 3 of 51 reviews (5.9%; exact 95% CI 1.2–16.2%). In CLEF, 43 content-relevant records had no abstract in PubMed and were scored from the title; none fell below 0.07.

### Comparison of stopping options

Table 3 compares the stopping options. At τ=0.07, the lowest recall was 0.958 in held-out reviews, 0.889 in Cochrane reviews, and 0.333 in one development review. Post hoc, for a single new review, the 95% prediction interval of the proportion read at τ was 18% to 93% in held-out reviews and 6% to 82% in Cochrane reviews (Supplementary Table 31). Label-free ranking with statistical stopping, evaluated post hoc, was reliable in every held-out and Cochrane review and read 87.6% and 86.1% of records, 28.8 and 47.0 percentage points more than the threshold (Fig. 4). Compared with the same stopping criterion on the default ASReview ranking, the Jev ranking reduced reading by 2.5 (95% CI 0.6–4.8) and 0.9 (-0.6 to 2.4) percentage points in held-out and Cochrane reviews, respectively.

### Pilot review

The screening decisions of the pilot review (Methods) were not public when the records were scored, so Jev could not have seen them in training. Post hoc, against final inclusion, with ten-record batches as in the benchmark, the AUC of Jev was 0.942, against means of 0.966 in the held-out reviews and 0.970 in the Cochrane reviews. Against title and abstract decisions, Jev ranked records better than the free rankers and slightly below Claude Opus 5.5: its AUC was 0.944 with single-record requests, against 0.838 for MiniLM, 0.829 for bge-base, and 0.951 for Claude Opus 5.5. Applied post hoc, the threshold would have read 63.2% of the PubMed records and retained all 111 included studies. In an illustrative conclusion-level analysis, the first included study was lost at τ=0.30, and domain-level conclusions were unchanged up to τ=0.50 (Supplementary Note 8).

## Discussion

Given only the eligibility criteria, Jev outranked the best free ranker in every held-out review and came within 0.011 AUC of Claude Opus 5.5 at about 1/700 of its list-price equivalent. Its accuracy held in a post-hoc external validation in 28 Cochrane reviews, with a mean AUC of 0.970. Combined post hoc with a statistical stopping criterion, its ranking reached 95% recall in all 51 held-out and Cochrane reviews, whereas a fixed score threshold fell short in three Cochrane reviews.

The ranking is useful in practice for two reasons. First, it needs no screening decisions: in post-hoc analyses of held-out reviews, the label-free Jev ranking alone provided 0.128 of the 0.147 gain in WSS@95 that the hybrid achieved over active learning (0.019 of 0.044 in Cochrane reviews) and found included studies earlier than the default ASReview ranking, so a team can read in a good order from the first record. Second, it is inexpensive; it was also more accurate and cheaper than GPT-4o-mini, although the saving, about $0.09 per 1000 records, is small and could narrow with prompt caching or batch pricing.

All benchmark labels were public before the model’s release, so the model might have encountered them in training. The mismatched-criteria control and the pilot review, whose screening decisions are unpublished, argue against simple recall of these labels, although the title-only control indicates that topic knowledge also contributes. The comparison with generative LLMs has two caveats. GPT-4o-mini gave token-level probabilities, and DeepSeek-V3.1 and Claude Opus 5.5 gave verbalized ones; the two kinds can differ in calibration.^25^ Because the verbalized scores of Claude Opus 5.5 produced more ties than those of Jev, its advantage might be larger under equivalent elicitation (Supplementary Note 15). Agreement between Jev and Claude Opus 5.5 is also not fully independent corroboration: a model from the same developer is one of the two references in the vendor’s evaluation, and our model inputs were drafted with that developer’s models (Methods).

Ranking alone does not tell a team when to stop. Because the scores were poorly calibrated, a threshold on them is an empirical cut-off. The fixed threshold reduced reading but missed one to three content-relevant records in three Cochrane reviews and gives no signal within a review when it fails. A concurrent evaluation in the CLEF 2019 collection kept every content-level include in its samples from 26 reviews, these three among them, but used a threshold of 0.06, single-record requests, and a reported model version.^23^ Criterion C4 showed lower workload than the knee method as adapted,^6,26^ not superior reliability, and for recall-critical health-care reviews the threshold is not supported as a stand-alone stopping rule. The statistical criterion was reliable in all reviews, in agreement with an evaluation of 15 stopping methods in which only the criterion of Callaghan and Müller-Hansen, the one used here, never stopped before the recall target.^11^ Because the same criterion was equally reliable on the default ASReview ranking, it was the screening decisions that protected recall; the Jev ranking reduced the records read by 2.5 percentage points in held-out reviews, but not distinguishably in Cochrane reviews. Reliability in all reviews does not establish a rate of at least 90% (Supplementary Table 23), but the criterion also tests recall within each review. For context, single human reviewers in a crowd-based trial had a sensitivity of 86.6% and pairs 97.5%, against a different reference standard.^27^

A team evaluating this workflow prospectively could take the eligibility criteria from its protocol, send records in ten-record requests in an order not sorted by relevance, screen in Jev order, and stop with a statistical criterion such as that of Callaghan and Müller-Hansen, with a recall target and a confidence level of 0.95. Abstract-less and non-English records would go to human screening. Because Jev reports no version, the team would need to archive scores and prompts and monitor drift by rescoring archived records, in line with guidance on AI use in evidence synthesis.^28^ Showing scores to screeners may lead to anchoring; safeguards include hiding the scores, dual screening of a random sample of unscreened records, and reporting the ordering method. Jev is not offered with zero data retention on the gateway used here, so teams should check confidentiality terms before sending unpublished material, and the product is in early access without independent documentation or audit, so its price and availability might change. We have implemented this workflow and these safeguards in ReviewFast (https://www.jolab.ai/reviewfast),^29^ an open-source application that was not used for the reported results.

This study has limitations. The analysis plan was not preregistered, and the change of active-learning comparator was recorded only after the held-out evaluation. Summary results of free rankers and three labels from the held-out reviews were displayed before the free-ranker comparator, λ, and τ were fixed; the held-out results of Jev were first viewed afterwards (Supplementary Note 1). Because the batch format, batch size, and free-ranker shortlist were set using the pilot labels, the pilot results are not an independent estimate. All criteria came from completed reviews (Supplementary Note 17), so performance with criteria written at the protocol stage is unknown. τ was derived on development reviews of at most 2000 records, although it kept 95% recall in 3 of 3 larger held-out and 12 of 13 larger Cochrane reviews; it was also derived under one unrecorded model version and one batching scheme and should not be applied in a new review without re-derivation on labeled reviews, and score stability on repeated scoring was not measured. The SYNERGY+ results describe open-access records with an abstract, and reviews that retained a larger share of their records tended to show a lower AUC and more reading at τ; the results may therefore be optimistic for complete searches. The CLEF content-level label may depart from the published reviews in either direction, and so may the Cochrane reliability estimate; the effect of missed studies on conclusions was not assessed in any meta-analysis. All evaluations were retrospective simulations, which assume consistent labeling and did not measure time savings. Records in languages other than English were not assessed. Jev was not compared with supervised ensembles,^18^ open-weight LLM rankers, or other decision models, and the generative LLMs were compared with it only on 1999 held-out records, so their workload is unknown. We did not evaluate settings that accept some loss of recall, such as rapid or scoping reviews, or stopping methods based on counting processes or practical procedures such as SAFE.^7,9,10^ Prospective studies should test the workflow with Jev and other decision models.

In conclusion, label-free ranking with Jev gave an accurate and inexpensive reading order from the first record, but stopping safely still required screening decisions.

## Methods

### Study design

The prompt, the batch size, and the shortlist of free rankers were fixed on the pilot review, and the free-ranker comparator, the hybrid weight λ, and the threshold τ on 73 development reviews of SYNERGY+. Four decision criteria (Table 2) were specified in an analysis plan before any metric on the 23 held-out reviews was computed; the plan is given in Supplementary Note 23. C1 required an AUC advantage over the strongest free ranker of at least 0.03 with a Wilcoxon *P*<0.01. C2 required non-inferiority to GPT-4o-mini, with the lower 95% confidence limit of the difference above -0.02, and an AUC deficit to Claude Opus 5.5 of at most 0.03 on the point estimate. C3 required a WSS@95 gain of the hybrid over active learning of at least 0.05 with *P*<0.05 and a gain in WSS@100. C4 required a threshold reliability of at least 90% with less work than the knee method at equal or higher reliability. Robustness to two rephrasings of the question was also required, with a mean within-review Spearman correlation of at least 0.90 and an AUC difference of at most 0.02. The margins and targets were set by judgment. The held-out reviews were then evaluated once.

Supplementary Note 1 lists the departures from the plan. The planned stopping rule, which estimated the number of remaining inclusions from calibrated probabilities, was replaced by the threshold rule during development, and the default ASReview model replaced the logistic-regression active learning named in the criteria; with the comparators of the plan, C3 and C4 led to the same decisions (Supplementary Note 5).

### Data sources

#### SYNERGY+

The primary data source was SYNERGY+ (version 3.0),^30^ an open collection of systematic reviews with search results, final inclusion labels, and eligibility criteria. It was downloaded with the synergy-dataset package (version 2.2), whose default export contains only open-access records with a valid abstract and omits reviews with fewer than three such included records. The export held 114 of the 119 reviews of the release, with 168,969 records and 4399 included studies, which are 35.3% of the records and 41.2% of the included studies of these reviews in the release; all results on SYNERGY+ refer to this subset. We used the official split of the package, which assigns 91 development and 23 held-out reviews. Development analyses used the 73 development reviews with at most 2000 records (Table 1), to limit the number of model requests. The primary label was final inclusion; 12 held-out reviews also report title and abstract decisions. The eligibility criteria were taken verbatim from the review metadata of SYNERGY+, which attributes them to the review publications (Supplementary Note 4).

#### CLEF 2019

The post-hoc external validation used the 31 Cochrane reviews of the CLEF 2019 Technology Assisted Reviews test set.^31^ Topics and relevance files were taken from the public repository of the task (task 2, test set), and 82,418 of 82,421 PubMed records were retrieved by PMID, of which 882 were relevant at the CLEF content (full-text) level. We call this label the CLEF content-level label; it was not checked against the included-study and excluded-study lists of the published reviews (Supplementary Note 7). Three reviews had no content-relevant record among the retrievable records and were analyzed only with title and abstract labels. The criteria were taken from the Objectives and Selection criteria sections of the abstract of each Cochrane review.

#### Pilot review

The pilot review is a systematic review by our group of genetic variants and domain-specific cognitive function in the Alzheimer’s disease continuum (D. Lee, J. P. Kim, K. Nho, A. J. Saykin, and T. Jo, manuscript under review). It searched PubMed, Embase, and Scopus on 18 June 2026. Two reviewers screened titles and abstracts in Rayyan and resolved disagreements by consensus; 216 records passed, and 111 of the 167 full texts assessed were included. The eligibility criteria were widened during full-text screening to include studies of mild cognitive impairment and of cognitively normal populations, so the title and abstract labels reflect the earlier criteria, whereas the criteria block given to the models was based on the revised criteria. Its PubMed search string was re-executed with the original search date as the end date (1572 records); records that had passed title and abstract screening were matched to PubMed records by title (143 of 216), and the 35 included studies absent from the PubMed arm were retrieved by DOI from PubMed or OpenAlex (33 with abstracts) and scored. A first matching by DOI had mislabeled three records, because some DOIs downloaded from PubMed belonged to cited references; the pilot results reported here use the corrected labels (Supplementary Note 1), and the included study that the first matching had assigned to another record was scored on 9 October 2026.

### Model, access, and provenance

Jev (TypeSafe AI, San Francisco, CA, USA) is a decision model. For a yes or no question about a block of text, it returns a score between 0 and 1 that the vendor describes as a calibrated probability;^21^ we used it as a zero-shot classifier. It was released in early access on 15 September 2026, and all scoring calls were made on 24 and 25 September 2026. Jev (typesafe-ai/jev), GPT-4o-mini (openai/gpt-4o-mini), and DeepSeek-V3.1 (deepseek/deepseek-v3.1) were called through the Vercel AI Gateway. Claude Opus 5.5 (claude-opus-5-5) was called through Anthropic’s command-line interface, Claude Code (version 2.1.281), with tools, settings files, and external servers disabled and a replaced system prompt. The responses of Jev contain no version or build string, so re-querying the model may not reproduce the scores; the stored per-record responses are the reproducible record of the primary analysis. The vendor does not disclose the model size, the training data, or a knowledge cut-off, and states that accuracy is lower in languages other than English. Its published evaluation uses the average predictions of two LLMs, GPT-6 Astra and Claude Fable 5.1, as reference probabilities and does not evaluate against observed outcomes of an external labeled task. Supplementary Note 1 gives further details of the access and the vendor documentation.

### Prompts and input formatting

All models received a criteria block followed by a record block, and no screening label was sent to any model. The criteria block was:

`Systematic review title: <TITLE>`

`Research question: <RESEARCH question>`

`Eligibility criteria reported by the review authors:`

`<ELIGIBILITY criteria>`

For the pilot review, the criteria block was a free-text summary of the eligibility criteria and did not use the title, research question, and criteria fields of this template. The record block was Title: <TITLE> and Abstract: <ABSHD>; when no abstract existed, (no abstract available; judge from the title).

Jev. Each request contained the criteria block, followed by “The following records are independent candidates retrieved by the search; judge each one on its own.” and ten record blocks labeled [Record R1] to [Record R10]. One probability question was asked per record: “Consider only Record Rk. Based on the title and abstract, should this record be advanced to full-text screening for this review? Give the probability that it meets the eligibility criteria.” The rephrasings for the robustness analysis were “Does this record potentially satisfy all of the eligibility criteria of this review, so that its full text should be retrieved and assessed?” and “Is this study likely to be included in this systematic review?” The model has no output-token limit and no temperature setting.

GPT-4o-mini (log-probability). System: “You are screening records for a systematic review. Answer with a single word: Yes or No.” User: criteria block, record block, and “Based on the title and abstract, should this record be advanced to full-text screening for this review? Answer Yes or No.” The settings were temperature 0, a maximum of 16 output tokens, the OpenAI provider only, and the top-20 log-probabilities of the first token; P(Yes)=p(yes)/(p(yes)+p(no)).

DeepSeek-V3.1 and Claude Opus 5.5 (verbalized probability). System: “You are screening records for a systematic review. Answer with a single JSON object and nothing else: {”p”: <ROBABILITY 0-1 that the record meets the eligibility criteria>}. A probability of 0.2 should mean about 20% of such records turn out eligible.” User: criteria block, record block, the question above, and “Return only the JSON object.” The settings were temperature 0 and a maximum of 60 output tokens for DeepSeek-V3.1, and the default temperature and no output-token limit for Claude Opus 5.5.

For Jev, the records of each review were sent in consecutive ten-record batches in the record order of the source datasets, without sorting by relevance or by any score. Single-record requests were used instead for the pilot-review comparisons with other models and with batched requests, for the 35 included studies of the pilot review from other databases, and for a post-hoc comparison with batched requests on the 2002 held-out records of the comparison with LLMs (Supplementary Note 3). The AUC of Jev in the comparison with LLMs came from the ten-record batches of all held-out records.

### Comparators

#### Free rankers

Free zero-shot rankers scored each record by its similarity to the criteria text with TF-IDF, BM25, and two sentence-embedding models (all-MiniLM-L6-v2 and bge-base-en-v1.5^32^). These four were retained from seven free rankers evaluated on the pilot review (Supplementary Note 6). The comparator for C1 was the free ranker with the highest mean AUC on the development reviews (bge-base).

#### LLMs

The generative LLMs, used as zero-shot classifiers, were GPT-4o-mini (OpenAI), DeepSeek-V3.1 (DeepSeek), and Claude Opus 5.5 (Anthropic). Because the gateway limited each chat model to five requests per minute, the LLMs were compared on 2002 held-out records comprising all 597 included studies and 1405 excluded records sampled at about 4.3% per review, drawn before the held-out evaluation (Supplementary Note 15).

#### Active learning

Active learning was simulated with the default model of ASReview^4^ (elas_u4): TF-IDF features of title and abstract, a linear support vector machine with class-balancing sample weights, and maximum-relevance querying of one record per iteration, with the classifier retrained after every labeled record (Supplementary Note 10). Priors were one included and one excluded record drawn with seeds 0–9 (ten random starts per review); they count toward the records read.

#### Hybrid

Records were ordered by Jev score (ties broken at random) and screened in that order until both classes had been seen; the ranking score for unscreened records was then z(SVM decision value)+λ×z(logit of Jev score), with z-scores computed over the unscreened pool. The weight λ=3 was chosen from {1, 2, 3} as the variant with the highest macro-averaged WSS@95 on development reviews, nearly tied with λ=2 (Supplementary Note 10).

Post hoc, we also evaluated weak supervision guided by Jev scores,^15^ the MedCPT cross-encoder^33^ as a further free ranker, and ASReview’s elas_h3 model, which uses sentence embeddings instead of TF-IDF features (Supplementary Notes 5 and 6).

### Stopping rules

#### Threshold

The label-free stopping rule reads only records with a Jev score of at least τ, set at the largest value of the grid {0.01, 0.02, 0.03, 0.05, 0.07, 0.10, 0.15, 0.20, 0.30} at which at least 95% of the 73 development reviews reached a recall of 95% or higher (τ=0.07).

#### Knee method

The comparator was a knee method on the ASReview ranking, adapted from Cormack and Grossman^6^ (the knee method as adapted). After at least 150 screened records and then every max(1, N/1000) records, it finds the knee of the gain curve and stops when the ratio of the slope before the knee to the slope after it is at least 156 minus the number of included records found (capped at 150). The published rule, as described by Yang and colleagues,^26^ also requires at least 1000 screened records and targets a recall of 0.7; the adaptation was judged against a recall target of 0.95. The rule was applied to each random start, and the reliability of a review was judged on the mean recall over the ten starts. Post hoc, the rule was also applied with the published minimum of 1000 screened records (Supplementary Note 11).

#### Statistical stopping criterion (post hoc)

The criterion of Callaghan and Müller-Hansen^8^ tests, at each point of screening, the null hypothesis that recall is below the target; screening stops when the smallest hypergeometric *P* value over all look-back windows is below 1 minus the confidence level. It was applied with a recall target of 0.95 and a confidence level of 0.95 to the ASReview, hybrid, and Jev rankings; label-free ranking with statistical stopping denotes this criterion applied to the Jev ranking. Unlike the threshold, the criterion uses the screening decisions of the records already read.

#### Random-sample check below the threshold (post hoc)

All records with a score of at least τ are read first. The records below τ are then drawn one at a time in random order. The hypergeometric probability of finding at most the observed number of positive records is computed under the smallest number of positive records below τ that would make total recall lower than 0.95. Screening stops when this probability is below 0.05 or when all records below τ have been read (Supplementary Note 14).

### Outcomes

The primary outcomes were the AUC, work saved over sampling at 95% recall (WSS@95),^2^ and, for stopping rules, reliability (the proportion of reviews with recall of at least 95%) and workload (the proportion of records read). AUC was computed per review with scikit-learn, with tied scores receiving mid-ranks. WSS@95=1-k95/N-0.05, where k95 is the rank of the ⌈0.95n⌉-th of the n included records, so that in reviews with at most 19 included records 95% recall means all included records. A secondary outcome was WSS@100=1-k100/N. Post hoc, we also computed the average time to discovery,^5^ the proportion of records screened when an included study is found, averaged across included studies. Mean average precision^13^ and the true negative rate at 95% recall,^34^ which are reported only in the Supplementary Information, are defined in Supplementary Note 19. Proportions read were macro-averaged across reviews.

### Statistical analysis

Metrics were computed within each review, averaged over seeds for simulations, and then averaged across reviews. The 95% CIs for means and paired differences are percentile bootstrap intervals from 5000 resamples of reviews unless stated otherwise; for proportions of reviews, we report Wilson intervals and, where stated, exact (Clopper-Pearson) limits. Paired comparisons used two-sided Wilcoxon signed-rank tests across reviews (SciPy, default settings). The *P* values of the two tests named in the decision criteria (C1 and the WSS@95 comparison of C3) are confirmatory; all other *P* values are exploratory, and no adjustment was made for multiplicity. The decision rule of the plan required that (C1 or C2) and (C3 or C4) hold on the held-out reviews and that the robustness check be passed; it does not control the overall type I error rate. Post hoc, paired reliability was also compared with exact conditional and mid-p McNemar tests^35^ and with Tango^36^ and Newcombe (method 10)^37^ score intervals for the difference of paired proportions. Zero differences were examined with Pratt’s method and a sign-flip permutation test, calibration was assessed with the observed-to-expected ratio and the slope of a logistic recalibration model,^38^ and prediction intervals for a new review were derived from a random-effects model.^39^ Supplementary Note 19 gives details of the tests and of the handling of reviews with tied values.

Software. Analyses used Python 3.12 (scikit-learn 1.9.1, NumPy 2.5.3, SciPy 1.18.1, pandas 2.3.3, ASReview 3.0.8) and Node.js 20.

### Post-hoc analyses

We label as post hoc all analyses other than the four decision criteria and the robustness check. They include the external validation in CLEF 2019 with all settings applied unchanged, weak supervision guided by Jev scores,^15^ single-record versus batched requests,^40^ title and abstract labels, ASReview’s elas_h3 model, a conclusion-level analysis in the pilot review,^41^ calibration, the statistical stopping criterion, and sensitivity to τ. Two negative controls, with the review title alone and with the criteria of a different review, used the 5550 records of the 12 development reviews of the robustness check. The CLEF validation, the comparison of single-record and batched requests, the mismatched-criteria control, and the conclusion-level analysis of the pilot review used Jev scores obtained in separate scoring after the held-out evaluation. Supplementary Note 1 gives when each analysis was planned and run.

### Literature search

PubMed was searched on 25 September 2026 for records on LLMs in systematic review screening published from 30 November 2022, the date of the public announcement of ChatGPT. A subset search added terms for active learning, prioritization, or stopping in the title or abstract (Supplementary Note 22).

### Provenance of model inputs and judgments

We used Claude Code (Anthropic), a command-line AI coding tool, to write the texts sent to the evaluated models: the criteria block of the pilot review, the Jev question, the batch preamble, and the rephrasings of the question; the prompt templates and system prompts of the LLMs; and the inputs of the two negative controls. The Jev question, the verbalized-probability system prompt of the LLMs, the criteria block of the pilot review, and the batch preamble were drafted with Claude Fable 5.1, one of the two reference models of the vendor’s evaluation, and the other texts with Claude Opus 5.5 (Supplementary Note 1). The tool was also used to write all analysis code, to run the analyses and the literature searches, to retrieve and check the references, to produce the figures, and to propose the judgments reported in Supplementary Notes 7 and 22.

### Use of AI tools

In addition to the uses described under Provenance of model inputs and judgments, T.J. used Claude Code as an assistant in preparing the study plan, the manuscript, and the Supplementary Information. The authors determined the content, revised all AI-assisted text, and take full responsibility for the manuscript.

### Ethics

Ethical approval was not required because the study used public bibliographic records, public screening labels, and the screening decisions and extraction data of a systematic review by our group, and involved no human participants or individual-level human data.

### Reporting guideline

The report follows the TRIPOD-LLM reporting guideline^42^ (Supplementary Note 24).

## Data Availability

SYNERGY+ and the CLEF 2019 collection are publicly available. The per-record model responses, the result files, and the analysis code are deposited at Zenodo (https://doi.org/10.5281/zenodo.23270999), except the result files of three post-hoc analyses that contain unpublished material. Records and screening decisions of the pilot review will be available from the corresponding author after the publication of that review. ReviewFast, the application described in the Discussion, is archived at Zenodo as version 0.3.1 (https://doi.org/10.5281/zenodo.23243074) and is available at https://www.jolab.ai/reviewfast.

## Data availability

SYNERGY+^30^ (Methods, Data sources) and the CLEF 2019 collection^31^ are publicly available. The raw per-record responses of every model, with batch membership, and the result files that underlie the tables and figures are deposited at Zenodo,^43^ except the result files of three post-hoc analyses that contain unpublished material; the deposit lists them with the reasons. Records and screening decisions of the pilot review will be available from the corresponding author after the publication of that review.

## Code availability

The analysis code and the prompts sent to the evaluated models, except the criteria block of the pilot review, are deposited with the data at Zenodo under the Apache License 2.0.^43^ ReviewFast, the application described in the Discussion, is archived at Zenodo as version 0.3.1 and is available at https://www.jolab.ai/reviewfast;^29^ the archive includes a script and the per-record data that reproduce the threshold and statistical-stopping results in Table 3.

## Acknowledgements

This study was funded by the Department of Radiology and Imaging Sciences, Indiana University School of Medicine, through start-up funds to T.J., and by Anthropic through Claude API credits from its AI for Science program. The funders and the vendor of Jev played no role in study design, data collection, analysis and interpretation of data, or the writing of this manuscript.

## Author contributions

T.J. conceived and designed the study, performed the analyses, and drafted the manuscript. D.L. and T.J. performed the title-and-abstract and full-text screening of the pilot review. D.L. and L.L. discussed the design and interpretation, verified the results, and reviewed and edited the manuscript. All authors read and approved the final manuscript.

## Competing interests

T.J. has received Claude API credits from Anthropic through its AI for Science program; Anthropic develops Claude Opus 5.5, one of the models compared, and Claude Code, the AI coding tool used in this study. T.J. is the author of Build with Claude Code (Packt, 2026). D.L. and L.L. declare no financial or non-financial competing interests.

## Notes

### Summary of Updates

Version 2 corrects an author affiliation, moves Figure 6 to the Supplementary Information as Supplementary Fig. 4, adds the web address of the ReviewFast application in the Discussion and in Code availability, and makes minor wording edits in the Introduction and in two headings. No result, number, table, or reference was changed.

