## Supplementary Information for "Label-free, low-cost AI ranking for systematic review screening"

The Methods of the main article describe the methods of the main analyses. The Supplementary Notes report the supplementary analyses, most of them post hoc, and describe how each was done; they also give per-review results and the reporting checklist. Supplementary Note 1 lists the post-hoc analyses with the times at which they were planned and run and records the steps that were executed before they were logged. "The plan" refers to the internal analysis plan described in the Methods (Study design), and P1 to P7 are the post-hoc analyses named in it and in a later memo (Supplementary Notes 1 and 23). The failure categories, the study-level reading of the CLEF content-level label in three Cochrane reviews, and the coding of prior studies were proposed with the AI coding tool used for the analyses (Methods, Provenance of model inputs and judgments). The main article does not use the classifications, the study-level reading of the CLEF content-level label in three Cochrane reviews, or the coding of prior studies. Reference numbers (ref.) refer to the reference list of the main article. The contents with page numbers follow the title page of the PDF version.

#### Supplementary Note 1. Record of the analysis plan, the settings, and the post-hoc analyses 1 to 52, with their planning and execution times

This note gives the times of the analysis plan, of the settings, and of the post-hoc analyses, from the creation and modification times of files and the times of commands in the project records (the session records of the AI coding tool used for the analyses and the files of the project; Methods, Provenance of model inputs and judgments), and lists the steps that were executed before they were logged or without a prior log entry. Times are US Eastern Time.

##### Plan file and held-out evaluation

The file, headed iteration 1, was created at 18:03:53 on 24 September 2026 and last modified at 13:14:44 on 25 September 2026. The data files of SYNERGY+ were created at 18:06, the first scoring log at 18:17:47, and the first model output on SYNERGY+ records at 18:18:15 on 24 September; no earlier run on SYNERGY+ exists in the project files. The scoring of the held-out records started at 00:48:48 on 25 September in an automated chain of commands, directly after the scoring of the development records and about 11 min before the entry of the plan file that fixed the free-ranker comparator, $\lambda$, and $\tau$ on the development reviews (01:00; the file of the development results was written at 00:59:23), and it was completed at 03:29:32. The scoring does not depend on $\tau$, $\lambda$, or the choice of comparator. The first active-learning runs, on the pilot review (18:31 to 18:45) and on 25 small development reviews (18:56 to 18:59) on 24 September, used logistic-regression and naive Bayes active learning; ASReview was installed from 19:00 to 19:02, the default ASReview model was first run at 19:02 (a test on one development review) and was used for all development runs from 19:06, and this change was recorded in the plan at 13:11 on 25 September, after the held-out evaluation. At 00:49:50 on 25 September, a status check displayed the summary line of the free rankers on the held-out reviews, cut after the macro AUCs of TF-IDF, BM25, and MiniLM, and one per-review line (Walker 2018) with the AUCs of all four free rankers, including bge-base (0.926); the free-ranker comparator, $\lambda$, and $\tau$ were fixed at 01:00. The ASReview baseline on the held-out reviews was simulated from 19:11 to 19:33 on 24 September with its summary output suppressed, so no held-out result was displayed. At 18:42:55 on 24 September, before the plan entry of 19:00 to 19:02, a functional test of the log-probability format of GPT-4o-mini scored three held-out records of one review and displayed their labels next to the probabilities; no metric was computed, and Jev was not involved. At 19:00 to 19:02 on 24 September, when results on the pilot review (including the simulations described below) and of 25 small development reviews, and no held-out metric, had been seen, an entry of the plan clarified the decision criteria and replaced the stopping rule named first, an estimator of the number of remaining inclusions that relied on calibrated probabilities, by the threshold rule. The file that holds the decision on the held-out reviews was created at 04:53:44 on 25 September, and the plan file states that the Jev results on the held-out reviews were first viewed at that time. Several time labels in the file were written as approximations; the last entry of the file verified them against the creation times of files produced by the same commands. The decision rule of the file planned a further iteration if the required criteria were not met, without stating whether the held-out reviews would be reused. Held-out reviews were evaluated once.

##### Settings taken from the pilot review

Of the settings taken from the pilot review (the prompt of Jev, the batch size, the free-ranker shortlist, and the log-probability elicitation of GPT-4o-mini), only the prompt of Jev (question wording and batch format) was fixed before the plan file was created. The first scoring of the pilot review ran from 09:19:35 to 10:55:14 on 24 September. The plan file as created names batches of five records; the pilot run with five records per request ended at 17:55:57, and the run with ten records per request ended at 18:08:12, after the plan file had been created. The change from five to ten records was appended to the plan file at 18:17:46 (labeled 18:40 in the file and verified as 18:17 by its last entry), 13 s after the output of the last evaluation and before the first model output on SYNERGY+ records (18:18:15); the batch size, but not the question wording, was therefore chosen after an evaluation on the pilot labels (AUC 0.941 with ten records and 0.943 with single requests; rank correlation 0.964; with five records, AUC 0.941 and rank correlation 0.970). These and the other pilot values in this note were computed with the labels in use before the correction of 9 October 2026 (Methods, Pilot review); Supplementary Note 5 gives the values with the corrected labels. The values for Jev with single-record requests and with five-record batches, for Claude Opus 5.5, and for DeepSeek-V3.1 and GPT-4o-mini on 544 records were known before the plan file was created; the value for ten-record batches and the values of the free rankers were obtained after the plan file had been created. The free rankers were evaluated on the pilot review from 18:17:49 to 18:46:55; the AUCs of six rankers were read at 18:28:58, and the development run of the four retained rankers was started at 18:46:12. The commands recorded from 17:25 to 18:46:55 contain no evaluation of results on SYNERGY+ records, apart from the functional test of 18:42:55 described above, which displayed three held-out labels without computing a metric; the first such command was issued at 18:56:22.

A post-hoc reconstruction from the project records and files (analysis 26 of Supplementary Table 2) found one question wording for Jev, one criteria text for the pilot review (1199 characters), and one input format with one record per request, all written at 09:18:47 on 24 September, and one system prompt and one user message for the three LLMs (verbalized probability), written at the same time. Seven evaluations against the pilot labels were run, by commands issued at 09:37:43 (partial output), 12:06:18, 14:03:11, 15:40:43, 17:34:07, 18:00:27, and 18:08:00; each output followed within 2 s, except that of 18:08:00, which came at 18:17:33. At 11:56:57, a further command listed pilot records with their labels (the six highest-scored records, the five highest-scored records that the review team had excluded, the four lowest-scored records that had passed title and abstract screening, and the four lowest-scored records), without computing a metric. The evaluation of 15:40:43 also covered the verbalized outputs of GPT-4o-mini (AUC 0.882, with eight distinct values) and DeepSeek-V3.1 (0.904) on a subset of 544 records. The question wording and the criteria text of the pilot review were not changed after any of the seven evaluations or after this listing, and the only change to the pilot scoring script (14:00:39) added an option to score a subset of records. The input format of Jev, the batch size, and the prompt of GPT-4o-mini were changed after evaluations against the pilot labels. First, at 17:31:36, after four evaluations, the batch format was added to the input of Jev (the preamble, the instruction "Consider only Record Rk.", and placeholders for a missing title or abstract), with the question and the system prompt of the LLMs unchanged. Second, batches of five and ten records were evaluated against the pilot labels in the last three evaluations. Third, the plan file lists the pilot AUC of GPT-4o-mini (0.882) among the results already seen and specifies token log-probabilities for GPT-4o-mini where possible; at 18:30:12, GPT-4o-mini was switched to the log-probability format, after two probe runs on a general question (six requests to GPT-4o-mini, DeepSeek-V3.1, and GPT-4o) and followed by a functional test on four pilot records in which no metric was computed. The log-probability format itself was not evaluated against the pilot labels. The only evaluation against the pilot labels between the first model output on SYNERGY+ records (18:18:15) and this change was that of the six free rankers (command 18:28:58, output 18:29:04), which involved no output of an LLM or of Jev. The runs of the LLMs on held-out records started at 18:30:37. The prompts of DeepSeek-V3.1 and Claude Opus 5.5 were not changed after the evaluations on the pilot review. The criteria-block template of the SYNERGY+ inputs (18:04:39) and the two rephrasings of the question (19:00:13) were not tried on the pilot review. The records do not show the reasons for the wording or for the change of the prompt of GPT-4o-mini beyond the commands and comments, any drafting outside the coding tool, or whether a person reviewed the prompts before the runs.

Active learning, hybrid variants, and stopping rules were also simulated on the pilot labels on 24 September, before the plan entry of 19:00 to 19:02. A simulation script written at 18:31:52 was first run at 18:31:59 and was stopped at 18:42:23 before it printed any result. A second run printed, at 18:44:08, WSS@95, WSS@100, and recall after reading 10% and 20% of records for logistic-regression active learning with the Jev score blended into the ranking score (weights 0.5, 1, and 2) or added as a feature (weights 1 and 3), logistic-regression active learning started in Jev order without blending, logistic-regression active learning without Jev (two kinds of starting records), naive

Bayes active learning, and the rankings by Jev, bge-base, and MiniLM alone. A third run, at 18:45:49, added, with the ten-record scores, the work and the recall against title and abstract labels of the label-free threshold rule at thresholds of 0.01, 0.02, 0.05, 0.1, 0.15, 0.2, and 0.3, of a knee rule, and of a target rule for three of these rankings. The earlier evaluations against the pilot labels at 12:06:18, 14:03:11, and 15:40:43 had printed, besides the AUC, the proportions of records read to reach 95%, 99%, and 100% recall and the work, recall, and precision of score thresholds at $p \geq 0.05$, 0.1, 0.2, 0.3, and 0.5 for Jev with single-record requests and for Claude Opus 5.5 (on all 1572 records at 12:06:18, with the observed share of records that passed title and abstract screening in tenths of the score; on the 544-record subset at 14:03:11 and 15:40:43, the latter also for GPT-4o-mini and DeepSeek-V3.1), as well as the AUC against final inclusion, the number of distinct score values, the mean scores of positive and negative records, rank correlations between models, and an ensemble AUC; the single-record rows of Supplementary Table 37 at $\tau=0.05$, 0.10, and 0.20 are values first displayed at 12:06:18, and the proportions read and the recall against title and abstract labels of its ten-record rows at $\tau=0.05$, 0.10, 0.15, and 0.20 are values first displayed at 18:45:49. These outputs were seen before the plan entry that replaced the stopping rule named first by the threshold rule and named the knee method as its comparator; $\lambda$ and $\tau$ were chosen afterwards on the development reviews. The outputs are kept with the revision records.

##### Analyses added after the held-out evaluation

We report as post hoc all analyses added after the held-out evaluation or not listed in the plan file. The plan file lists the post-hoc analyses P1 to P5: weak supervision guided by Jev scores (P1; ref. 15), single-record versus batched requests (P2; ref. 40), title and abstract labels (P3), ASReview's strongest preset (P4), and a classification of included studies below the threshold (P5). A separate memo file, created at 09:48:43 on 25 September 2026, after the held-out decision (04:53:44), holds two further entries: the CLEF validation (P6), with methods applied as fixed on SYNERGY+ (ten-record batches, $\lambda=3$, $\tau=0.07$) without re-tuning, and a conclusion-level analysis in the pilot review (P7; ref. 41). The CLEF records file was created at 10:00 on 25 September, the scoring of the CLEF records was completed at 17:03, and the CLEF results file was written at 17:21; the files of the conclusion-level analysis were written at 09:52 to 09:55 on 25 September; for this analysis, the 34 included studies of the pilot review that were absent from the PubMed arm were scored by Jev with single-record requests from 09:52:29 to 09:55:23, after the held-out decision. A 35th included study, which the first matching had assigned to another PubMed record, was scored in the same way at 16:06 on 9 October 2026. The two negative controls (review title only, and criteria of a different review) are recorded in neither file; their input files were created at 19:09:13 on 24 September, before any held-out metric, the title-only run was completed at 04:37:44 on 25 September, before the held-out decision file was created, and the run with mismatched criteria was completed at 05:01:25, after it. Both controls and the two rephrasings used 5550 records of 12 development reviews. The hybrid as fixed (the default ASReview model with $\lambda=3$) was applied to the pilot review at 04:56 on 25 September, after the held-out decision (logistic-regression hybrids had been simulated on the pilot labels on 24 September; see Settings taken from the pilot review), and the comparators named in the plan were computed on the held-out reviews at 13:37 on 25 September, after the correction entry of 13:11.

Analyses 1 to 21 (26 September). The plan of each analysis was written to an append-only log at 17:43:28, and no analysis was run before that time; details of the plans and changes to them were added to the log between 17:50 and 18:45. The following steps were executed before they were logged, or were not logged beforehand. Analysis 1: the fitting routine of the recalibration model was replaced after a first launch had been stopped because of its run time, and, after the results of the second launch had been inspected, the comparison of coefficients between scoring modes was restricted to reviews in which the fit had converged in both modes; both changes were logged afterwards (18:09:56), the model itself was unchanged, the other results of the second and third launches were identical, and the bootstrap CIs over reviews were an addition that the plan did not name. Analysis 19: the inclusion rates reported for the published ranker were compared with those of the two relevance files of CLEF 2019 at 17:53:44, and the search dates of the topic files were extracted at 17:55:08; the log entry that names this comparison as the method was written at 18:02:00 and was worded as an entry made before execution, and the script of the record was run at 18:04:18 and gave the same values. Analysis 21: the project records and the installation times of the command-line interface were inspected at 17:50:10 to 17:51:09, and a copy of the script for the batch check of the pilot review was re-run at 18:00:53, before the log entry of 18:02:00. Analysis 8: the cross-check against the buscarpy package (Supplementary Note 14) was made at 3148 evaluation points (the hybrid rankings of all 51 reviews) and not at every evaluation point; after the re-run simulations had been compared with the stored values, the script was changed at 18:48:44 to 18:49:24, before its first run at 18:56:17, so that the check of the saved labeling orders used the values of the same run and the analysis of the influence of single reviews used the stored values, and both points were recorded after the execution (19:03:50). Analyses 2, 4, 6, and 7: additions made while the scripts were written were not logged beforehand (the Jev ranking for the title and abstract labels of CLEF 2019 and two further comparisons in analysis 2; the knee method on four further rankings, results by review type, and intervals for the development reviews in analysis 4; five further values of $\tau$ and the held-out reviews at the level of final inclusion in analysis 6; and categories D and E with the counts at the level of studies in analysis 7). Analysis 3: the script also compared the hybrid at each weight and the Jev ranking with ASReview (36 rows of Supplementary Table 57), whereas the log entries made before the run (17:43:28 and 18:24:45) named only the differences from the hybrid with $\lambda=3$ and from the Jev ranking. Analysis 15: further sources were retrieved at 19:04:59 to 19:10:34 (the documentation site and the legal documents of the vendor, policy pages of the gateway, and archived copies of the vendor's pages in the Internet Archive) that neither the plan nor the detailed log entry of 18:45:25 named. Analyses 12, 17, 18, and 20: the retrieval of public documents began before the detailed log entries, and analysis 18 has no detailed log entry. Analyses 7, 8, 9, 16, and 17: first steps preceded the detailed log entries and the starts given in Supplementary Table 1, and the execution records did not name them until 27 September (analysis 9, a command at 17:49:38 that counted the refused records and the distinct score values of each model; analysis 16, a command at 17:52:03 that counted the studies and genes of the pilot review from its extraction sheet, whose script of record was first run at 18:04:45 and gave the same counts; analysis 17, commands at 17:48:30 and 17:51:57 that listed the metadata of the dataset by split and the files of its package; analysis 7, commands at 17:48:50 to 17:50:03 that tested the access to the full texts of the Cochrane reviews; analysis 8, the retrieval of the information on the package and of the publication of the criterion at 18:21:49 and 18:22:13, before the log entry of 18:24:45, and the creation of the separate environment at 18:25:24, before the log entry of 18:34:48). All these steps followed the plan of 17:43:28 and carried out methods that the plan names. The plans named the outcomes of each analysis; the result files hold further values per review, per random start, and per metric, and they are deposited with the code, with the exceptions stated under Data availability.

Checks of 27 September. Two further sets of checks were planned and run on 27 September 2026 and used no model call. The first recounted the reproduction of the simulations, recomputed from the per-review files the values that had been stored to four decimals, computed the throughput directly from the start and end times, recounted the reviews with title and abstract labels, and compared the stopping decisions of the statistical criterion with exact arithmetic; it was logged at 00:47:01 and run between 00:47:39 and 01:08:49. The second recounted in exact arithmetic the reviews of every paired comparison and repeated the tests, confirmed from file times and from the project record when the settings taken from the pilot review had been fixed, and confirmed the times of the first steps listed above; it was logged at 04:02:58 and 04:07:40 and run between 04:03:07 and 04:26:27, except that the project record of 24 September was first read at 04:04:20, before the log entry of 04:07:40 that names it. The five $P$ values for the comparisons of the threshold with the knee method in Supplementary Table 25 were computed in the second set of checks. The reported values are those obtained after both sets of checks.

Analyses 22 to 27 (27 September). These analyses were planned at 20:48:01 on 27 September 2026 and run between 20:52 and 21:22. They used the stored scores, the record files, the project records, and public documents, and no model call. Changes of plan were logged before the runs concerned (21:03:04, 21:05:13, 21:10:22, and 21:13:55). Technical corrections made during execution were recorded only afterwards, in the execution record of each analysis: the decoding of the compressed archived pages before their comparison (analysis 24; the first, void comparisons were kept separately), a widened text search and a corrected section label in the check of the sources, whose first output was overwritten (analysis 25), a corrected consistency check in the summary of analysis 26, and further comparison rows for the threshold and the knee method on the ASReview ranking (analysis 23). One connectivity check of the archive index (20:51:30) preceded the log entry that started the runs. The first attempts of analyses 24 and 25 failed on the verification of certificates and saved nothing but the request lines. For analysis 26, a first parser found no version of the scoring scripts, and the commands of the project records were then read on screen before the change of plan of 21:13:55; the second parser listed only outputs that print an AUC, WSS, or workload label and therefore missed the evaluation of the six free rankers against the pilot labels at 18:28:58, whose output printed the values without such a label, and this evaluation was confirmed from the project record on 28 September 2026. Analysis 25 is a verification of sources for the statistical methods and the references. A further query of the archive index at 22:30 was answered; no reported value depends on it.

Analyses 28 to 31 (28 September). These analyses were planned at 10:28:33 on 28 September 2026 and run between 10:36:27 and 10:43:04. They used the stored scores and record files, the saved copies of the vendor announcement, and public bibliographic records, and no model call. Supplements to the plans were logged at 10:34:49, before the runs. For analysis 29, descriptive fields (the numbers of partner records below and at or above $\tau$, and the index of each request) were added between two runs whose counts, expected values, and probabilities were identical (logged at 10:39:01). For analysis 31, a query of the DOI resolver was added before the requests (logged at 10:41:38). Analysis 30 made no network request, because its plan made the query of later captures conditional on the absence of the sentence from the saved copies. Analysis 31 is a check of bibliographic sources for the reference list.

Analyses 32 to 46 (28 September). The plans of these analyses carry the heading time 19:48:38 (the output of the date command) and were appended to the append-only log at 19:50:28 on 28 September 2026 (with a correction of the wording of analysis 42 at 19:50:36, before its run); no analysis step was run between these times, and details and changes were logged before the steps concerned at the times given in Supplementary Table 3; analysis 36 was run at 19:57:48, after its plan and before the detailed entry of 20:01:18, which did not concern it. The following steps were executed before they were logged, or were not logged beforehand. Analysis 32: a block of look-ups of stored values (the unrounded observed-to-expected ratio of the CLEF reviews and the macro-averaged WSS@95 at each $\lambda$) was added to the script without a prior log entry, and the key names of a stored result file were corrected in the script before its only run. Analysis 33: before its first run (20:06:30), the bootstrap draws were changed at 20:06:26 from one shared random-number generator to a new generator with the seed 20260928 in each summary; no output had been produced, and the change was recorded only in the execution record. Analysis 34: after the simulations had finished, a descriptive check of the starting sets of the seeds was run at 20:13:04 to 20:13:17, before the log entry of 20:13:42 that names it, and that entry is worded as if the check had not yet been run. Analysis 35: bge-base was ranked both with random ties and in file order, which the plan had not fixed. Analysis 39: the second and third runs of the summary script (starts 20:13:56 and 20:20:50) were not logged beforehand; the second changed only summary counts, the third added comparisons named in the plan, and the $P$ values of the earlier runs were unchanged. Analysis 44: one request that checked that the model repository could be reached (20:03:23) preceded the detailed log entry of 20:29:06, and a check of the environment against the example of the model card (20:29:32 to 20:30:40) was not named in the log. Analysis 45: one request to Crossref (the DOI of ref. 7, status code only) at 19:56:55 preceded the detailed log entry of 19:59:58, and six steps that the log entry of 20:07:51 introduced were taken before that entry (the programming interface of the publisher Elsevier at 20:03:43, the arXiv abstract page at 20:04:17 and the PDF at 20:04:26, text extraction from the PDF at 20:05:48, an arXiv author page at 20:07:06, which was not found, and arXiv title searches at 20:07:13 to 20:07:27); the execution record had stated that the log entries preceded every request, and a correction was appended to it. Analysis 46:

the folders and files named as protocols in the CLEF 2019 repository were listed at 19:42:42 and 19:42:47, and two of the three files were read in part at 19:42:55 (the first 1500 bytes of CD012164.xml with a count of its XML tags, and the first 800 bytes of the 2019 file of CD011686; CD011571 was listed only), before the plan of 19:48:38 that excluded them; the first attempt stopped with an interpreter error before any request, the second failed on the verification of certificates, and the third was stopped after a parsing error; manual queries at 23:08:49 to 23:10 and at 23:31 were not logged beforehand, and the log entries of 23:10:50, 23:21:19, and 23:31:58 recorded the changes before the next attempts. The failed attempts are kept with their outputs, and none was used for the decision on feasibility. No step of these analyses called an evaluated model.

Analyses 47 to 50 (29 September). The plans of these analyses carry the heading time 04:05:47 (the output of the date command) and were appended to the log at 04:06:41, with no analysis step run in between, with a correction of the path of an input file of analysis 49 at 04:06:49, and the analyses were run between 04:14:36 and 04:57:35 on 29 September 2026 (Supplementary Table 3). The following steps were executed before they were logged, or were not logged beforehand. Analysis 50: the text-mining address of the publisher named in the Crossref record of ref. 34 was requested at 04:16:10, before the log entry of 04:16:33 that added further sources, and returned bibliographic data only. Analysis 49: the check of the location of the 383 comparisons was corrected twice and re-run (starts 04:39:08 and 04:48:16), once for the format of the *P* values of the tables of the original analyses and of analyses 1 to 31 and once for the search window of the two tests on the development reviews; the results of the extension were identical in the three runs, and the outputs of the earlier runs are kept; the analysis also read the script that builds the tables of the original analyses and of analyses 1 to 31, which its plan did not name. Analysis 48 read the membership of the three sets of reviews from the per-review file of analysis 44, and its per-review check of the truncation counts against analysis 44 was run at 04:48:20, after its end file had been written (04:39:08). Analysis 47 read the source of the scoring script and of two scripts of the automated chain of commands, which its plan did not name, and used only the entries of 24 and 25 September 2026 of the project record that it shares with later work; while these entries were located, the outputs of four monitoring commands of 25 September 2026 were displayed, which contained aggregated results of the original analyses, and none of them was used. No step of these analyses called an evaluated model.

Analysis 51 (29 September). The plan of this analysis carries the heading time 06:53:39 (the output of the date command) and was appended to the log by the same command; the analysis was run from 06:58:54 to 07:03:06 on 29 September 2026 (Supplementary Table 3), and the two requests named in the plan were sent from 06:59:37 to 06:59:41. The comparison script was run three times. The first run (07:01:28, from the project records) stopped before writing any output because one quotation was interrupted by a margin line number of the extracted text; the second run (07:01:51) wrote two page references that did not agree with the pages found by the script itself. The change before the second run was logged at 07:01:44; one line that implemented it was added with the command that started the second run (07:01:51) and was recorded afterwards, in the entry of

07:02:15, which also logged the change before the third run; the output of the second run is kept, and the judgments and quotations of the second and third runs are identical. The comparison also checked the sentence of the Methods that cites the article and recorded the dates on its cover page, which the plan did not name; the comparison of the bibliographic data with the Crossref record was planned. No step of this analysis called an evaluated model.

Analysis 52 (29 September). The plan of this analysis carries the heading time 09:07:49 (the output of the date command) and was appended to the log by the same command; the analysis was run from 09:15:49 to 09:19:09 on 29 September 2026 (Supplementary Table 3). While the plan was prepared, the names of the columns and keys of the stored result files were read, and in doing so the stored summary values of the random-sample check at each label level (the mean over orders of the indicator of 95% recall, the lowest recall of a single order, and the reviews with an order below 95% recall) and the first two rows of three stored files were displayed; the plan entry lists the values of the first two files, and a later log entry (29 September, 11:45) records the third, the per-review file of analysis 42, whose two rows were development reviews, which analysis 52 does not use. Further consistency checks and a tabular copy of the results were logged at 09:17:39, before the only run of the main script (09:17:44). The comparison of the reviews not reliable on the mean recall with the reviews that had a run below 95% recall was logged at 09:18:08, after the results of the main script had been seen, and was run once at 09:18:28. No step of this analysis called an evaluated model.

##### **Archived copies of the vendor announcement**

The archived copy by which the vendor announcement is cited (Internet Archive capture 20260926091045, 09:10 UTC on 26 September 2026) has a text identical to the copy of the page saved on the analysis server on 26 September 2026. The earliest capture (20260915192610, 15 September 2026, 19:26 UTC) contains the same sentence on the reference models but shows the date "Sep 14, 2026", and its text differs from that of the later copy in 19 lines of the later copy and 17 lines of the earliest capture (title, date, navigation, headings, two footer links, and two typographical corrections in the body text), so the page was edited after its release (post hoc; analysis 24 of Supplementary Table 2).

##### **Model periods of the coding tool**

The session records carry a version field in every message entry and the model identifier in every response of the model. The main session was started at 04:53 on 24 September 2026 with version 2.1.281; after a restart at 19:41 on 26 September 2026, the entries carry version 2.1.282. The model was Claude Fable 5.1 from the start of the session until 17:36 on 24 September (the work on the pilot review), Claude Opus 5.5 from 17:54 on 24 September until 09:33 on 26 September (the scoring runs and original analyses of SYNERGY+ and the CLEF reviews, the classification of included studies below the threshold, and the original conclusion-level analysis), Claude Fable 5.1 from 15:45 on 26 September until 07:40 on 27 September (analyses 1 to 21), and Claude Opus 5.5 from 07:41 on 27 September until the last entry read (22:52 on 27 September; analyses 22 to 27 and the readings of the session records of 27 September, 09:42 to 22:52). The readings of the session record of 24 September reported under Settings taken from the pilot review were made on 28

September 2026, from 08:10 to 08:14 and from 08:58 to 08:59, and analyses 28 to 31 (run in a session whose entries span 10:33:32 to 10:47:30; Supplementary Table 2) and the check of a reference at 11:57 were run on 28 September, all in sessions of the same tool whose records carry only the model Claude Opus 5.5 and the version 2.1.282 (records read up to 08:58:58 for the first two sessions). Analyses 32 to 46 were planned (19:48:38) and run (19:57 to 23:42, Supplementary Table 3, with the earlier steps listed above) on 28 September 2026 in sessions of the same tool whose records carry the version 2.1.282 in every entry and the model Claude Opus 5.5 (claude-opus-5-5) in every response (records read on 29 September 2026). Analyses 47 to 50 were run (04:14:36 to 04:57:35 on 29 September 2026) in sessions of the same tool whose records carry the version 2.1.282 in every entry and the model Claude Opus 5.5 (claude-opus-5-5) in every response (records read on 29 September 2026), and so were analysis 51, which was planned and run from 06:53:39 to 07:03:06 on 29 September 2026, and analysis 52, which was planned and run from 09:07:49 to 09:19:09 on the same day. Of the texts sent to the evaluated models, the Jev question, the verbalized-probability system prompt of the LLMs with its sentence on the probability scale, and the criteria block of the pilot review were first written at 09:18:47 on 24 September, and the batch preamble at 17:31:36, all in the period of Claude Fable 5.1, one of the vendor's two reference models; the criteria-block template of the SYNERGY+ inputs (18:04:39), the log-probability prompt of GPT-4o-mini (18:30:12), the two rephrasings (19:00:13), the inputs of the negative controls (19:09:11), and the criteria sections of the CLEF reviews (09:49:41 on 25 September) were first written in the period of Claude Opus 5.5 (analyses 24 and 26 of Supplementary Table 2). Read again on 4 October 2026, the entries of the main session after 07:41 on 27 September 2026 carry only the model Claude Opus 5.5, up to the last entry read (16:02 on 4 October 2026); the readings of the session records of 24 and 25 September 2026 that added to this note the evaluations of 12:06:18, 14:03:11, and 15:40:43, the simulations of 18:31 to 18:45, and the functional test of 18:42:55 were made in this period.

**Supplementary Table 1. Further post-hoc analyses with their planning and execution times (26 September 2026)**

| Analysis | Purpose | Planned | Executed (start to end) | Reported in |
| --- | --- | --- | --- | --- |
| 1 | Calibration of Jev scores | 17:43:28; details 17:50:26 | 17:53:40 to 18:09:15 | Note 9 |
| 2 | Label-free Jev ranking and the increment of the hybrid | 17:43:28; details 17:50:27 | 17:55:34 to 17:59:48 | Note 10 |
| 3 | Larger values of $\lambda$ | 17:43:28; details 18:24:45 | 18:25:07 to 18:31:43 | Notes 10, 21 |
| 4 | Uncertainty, strata, and score bands for stopping rules | 17:43:28; details 17:50:27 | 18:00:54 to 18:01:01 | Note 11 |
| 5 | Grid, sensitivity, and re-selection of $\tau$ | 17:43:28; details 17:50:26 | 17:55:24 to 18:06:50 | Note 12 |
| 6 | Threshold against title and abstract labels; pilot review | 17:43:28 | 17:55:49 to 17:56:01 | Note 13 |
| 7 | CLEF 2019 reviews with recall below 95% | 17:43:28; details 17:50:26 | 17:51:32 to 18:03:53 | Note 7 |
| 8 | Statistical stopping criterion, label-free ranking with statistical stopping, time to discovery, recall curves, influence of single reviews | 17:43:28; details 18:24:45 and 18:34:48 | 18:35:25 to 19:04:19 | Note 14 |
| 9 | Composition of the subset for the comparison with LLMs | 17:43:28; details 17:59:01 | 18:04:11 to 18:04:33 | Note 15 |
| 10 | Publication year and label publicity | 17:43:28; details 17:59:01 | 18:04:33 to 18:04:42 | Note 16 |
| 11 | Provenance and length of the eligibility criteria | 17:43:28; details 17:59:01 | 18:04:42 to 18:04:45 | Methods; Notes 4, 17 |
| 12 | Cost, tokens, and throughput | 17:43:28; details 17:59:01 | 17:52:23 to 18:09:09 | Note 2 |
| 13 | Statistical procedures and exact *P* values | 17:43:28; details 17:50:27 | 17:56:01 to 18:00:22 | Methods; Note 19 |
| 14 | Simulation through the programming interface versus the command-line tool | 17:43:28; details 17:59:01 | 18:07:29 to 18:08:39 | Note 5 |
| 15 | Gateway response and vendor documentation | 17:43:28; details 18:24:45 and 18:45:25 | 19:00:39 to 19:11:16 | Methods |
| 16 | Study and gene counts in the pilot review | 17:43:28; details 17:59:01 | 18:04:45 to 18:10:44 | Note 8 |
| 17 | SYNERGY+ release and split | 17:43:28; details 17:59:01 | 17:52 to 18:04:51 | Methods; Note 4 |
| 18 | Verification of references | 17:43:28 | 17:55:33 to 18:10:06 | Note 22 |
| 19 | Label level of the published LLM-guided ranker; search dates of CLEF 2019 | 17:43:28; details 18:02:00 | 17:52:07 to 18:10:28 | Notes 18, 24 |
| 20 | Literature searches (Introduction and Methods) | 17:43:28; details 18:02:00 | 17:58:30 to 18:10:55 | Introduction; Methods; Note 22 |
| 21 | Provenance of the plan, timing, tool versions, and scoring modes | 17:43:28; details 18:02:00 | 17:49:11 to 18:11:21 | Methods; Note 1 |

Times are those of the log, of the execution records of each analysis, and of the files. For analyses 7, 12, 17, 18, 19, 20, and 21, the start is the time of the first retrieval of a public document or of the first file inspection that the execution record names; for analyses 12, 17, 18, 19, 20, and 21, this start precedes the detailed log entry. For analyses 7, 8, 9, 16, and 17, earlier first steps that the execution records did not name are listed above (17:48:50, 18:21:49, 17:49:38, 17:52:03, and 17:48:30).

Analyses 1, 2, 13, and 16 were run more than once because of errors in the scripts or added output fields, and the start is that of the first run (for analyses 2, 13, and 16, the creation time of the file that holds the start time); the final runs started at 18:04:58, 17:59:11, 17:59:48, and 18:10:42. Analyses 7 and 12 were also run more than once after failed attempts that produced no result: in analysis 7, the first run of the listing script failed at 17:56 with a certificate error and was re-run at about 17:57, and the compiling step failed on a key name and was re-run at 18:03:53; in analysis 12, the first attempt at 18:06 failed because a file was not found, and the successful run started at 18:09:06. The figures of analysis 8 were first drawn at 18:59:33 and redrawn at 19:04:11 after a layout fix, with no change to any number. The end is that of the last step, which for analyses 1, 5, and 8 was the redrawing of figures.

**Supplementary Table 2. Further post-hoc analyses with their planning and execution times (27 September 2026, analyses 22 to 27; 28 September 2026, analyses 28 to 31)**

| Analysis | Purpose | Planned | Executed (start to end) | Reported in |
| --- | --- | --- | --- | --- |
| 22 | Order of records in the data files and composition of the ten-record requests | 20:48:01; change 21:10:22 | 21:11:27 to 21:12:52 | Note 3 |
| 23 | McNemar-type tests and score intervals for paired differences in reliability; knee method on the hybrid ranking by review size | 20:48:01; supplement 21:05:13 | 21:08:32 to 21:08:36 | Methods; Note 11 |
| 24 | Archived copies of the vendor announcement; model periods of the texts sent to the evaluated models | 20:48:01; change 21:03:04 | 20:52:45 to 21:22:21 | Methods; Note 1 |
| 25 | Verification of sources (knee rule, ref. 11, McNemar-type methods) | 20:48:01; change 21:03:04; supplement 21:05:13 | 20:53:29 to 21:18:03 | Methods; Note 22 |
| 26 | Prompt development on the pilot review | 20:48:01; change 21:13:55 | 21:12:58 to 21:16:19 | Note 1 |
| 27 | Inclusion rates of the CLEF 2019 relevance files by review type | 20:48:01 | 21:06:34 | Note 18 |
| 28 | Direction of the differences in file position; order of the data file among tied scores | 10:28:33; supplement 10:34:49 | 10:36:27 to 10:36:44 | Note 3 |
| 29 | Positive records below $\tau$ and request composition; stored score of Kennis 2004 | 10:28:33; supplements 10:34:49 and 10:39:01 | 10:38:03 to 10:39:16 | Notes 3, 12 |
| 30 | Vendor's statement on price in the saved copies of the vendor announcement | 10:28:33; supplement 10:34:49 | 10:40:28 to 10:40:49 | Note 2 |
| 31 | Bibliographic data of refs 26 and 18 | 10:28:33; supplement 10:34:49; addition 10:41:38 | 10:41:38 to 10:43:04 | Note 22 |

Times are those of the log, of the execution records of each analysis, and of the files (US Eastern Time). The end of analysis 24 is the time at which the output file of its last archive query was written. Analysis 29 was run twice (10:38:03 to 10:38:06 and 10:39:13 to 10:39:16), and the start is that of the first run.

**Supplementary Table 3. Post-hoc analyses 32 to 52 with their planning and execution times (28 September 2026, analyses 32 to 46; 29 September 2026, analyses 47 to 52)**

| Analysis | Purpose | Planned | Executed (start to end) | Reported in |
| --- | --- | --- | --- | --- |
| 32 | Pooled failures of the threshold, positive records below $\tau$ by label level, cost and records not read, resolution of the scores, intervals for reliability | 19:48:38 | 20:10:15 to 20:10:38 | Methods; Notes 11, 13, 2 |
| 33 | Knee method with the published minimum of 1000 screened records | 19:48:38; details 20:01:18 | 20:06:30 to 20:17:09 | Note 11 |
| 34 | Hybrid with ten tie-breaking seeds of the Jev starting order | 19:48:38; details 19:59:47; addition 20:13:42 | 20:00:00 to 20:26:54 | Note 10 |
| 35 | True negative rate at 95% recall; rounding of k95 | 19:48:38; details 20:27:56 | 20:27:56 to 20:28:26 | Methods; Note 10 |
| 36 | Records on different sides of $\tau$ under three scoring modes | 19:48:38 | 19:57:48 to 19:58:20 | Note 12 |
| 37 | Out-of-sample reliability of the selection of $\tau$; results by review size | 19:48:38; details 20:01:18 | 20:05:03 to 20:05:10 | Note 12 |
| 38 | Two-stage bootstrap, refused records, and calibration in the comparison with LLMs | 19:48:38; details 20:01:18 | 20:05:03 to 20:05:55 | Note 15 |
| 39 | Treatment of zero differences in paired tests; confirmatory and exploratory $P$ values | 19:48:38; details 20:01:18 | 20:08:49 to 20:21:17 | Methods; Note 19 |
| 40 | CIIs for the negative controls and the rephrasings | 19:48:38; details 20:01:18 | 20:01:41 to 20:05:50 | Note 5 |
| 41 | Share of the release retained in the SYNERGY+ export, per review | 19:48:38; details 20:01:18 | 20:08:28 to 20:08:39 | Note 4 |
| 42 | Threshold with a random-sample recall check below $\tau$ | 19:48:38; correction 19:50:36; details 20:01:18 | 20:08:00 to 20:09:24 | Methods; Note 14 |
| 43 | Prediction intervals for the proportion of records read | 19:48:38; details 20:01:18 | 20:08:00 to 20:08:23 | Methods; Note 11 |
| 44 | Biomedical cross-encoder (MedCPT) as a further free ranker | 19:48:38; details 20:29:06 | 20:29:06 to 23:07:38 | Methods; Note 6 |
| 45 | Verification of new references; search for a published per-record screening time | 19:48:38; details 19:59:58 and 20:07:51 | 20:00:05 to 20:11:20 | Note 22 |
| 46 | Retrieval of protocol-stage criteria for the Cochrane reviews (feasibility) | 19:48:38; details 23:08:24; changes 23:10:50, 23:21:19, and 23:31:58 | 23:08:24 to 23:42:29 | Discussion; Note 17 |
| 47 | Scoring times of the three scoring modes of the 2002-record subset; record of the provider | 04:05:47 | 04:14:36 to 04:20:45 | Notes 3, 12 |
| 48 | Part of the MedCPT query-document pairs shortened by truncation to 512 tokens | 04:05:47 | 04:21:30 to 04:48:20 (computation ended 04:32:53; per-review check 04:48:20) | Note 6 |
| 49 | Scope of the zero-difference check of analysis 39 and its extension to analyses 32 to 46 | 04:05:47; correction<br>04:06:49 | 04:22:54 to<br>04:57:35 | Methods;<br>Notes 10, 11,<br>19 |
| 50 | Verification of refs 34 and 24 and of the description of ref. 17 | 04:05:47; change<br>04:16:33 | 04:14:36 to<br>04:18:15 | Introduction;<br>Results;<br>Methods;<br>Notes 2, 10, 22 |
| 51 | Definitions of ref. 34 (95% recall point, TNR@95, fewer than 20 included records) compared with the Methods | 06:53:39; changes<br>07:01:44 and 07:02:15 | 06:58:54 to<br>07:03:06 | Methods;<br>Notes 10, 22 |
| 52 | Single-run reliability of the evaluated stopping options of Table 3 | 09:07:49; changes<br>09:17:39 and 09:18:08 | 09:15:49 to<br>09:19:09 | Table 3; Note<br>14 |

Times are US Eastern Time from the append-only analysis log, the start and end files written with the date command in each analysis folder, and the execution records. Analysis 33 was run twice (20:06:30 to 20:06:41 and 20:16:51 to 20:17:09; the second run added the ten-seed hybrid orders of analysis 34 and reproduced every value of the first). Analysis 34 finished its computation at 20:20:31; the end file was written at 20:26:54. Analysis 35 finished at 20:28:01; the end file was written at 20:28:26. Analysis 39 comprises a copy of the exact recount (20:08:49 to 20:09:22) and three runs of the summary script (20:12:12, 20:13:56, and 20:20:50 starts). In analysis 44, the scoring of 115,419 record pairs ran from 20:30:46 to 22:58:44. The planning time 19:48:38 is the output of the date command that gives the heading of the plan entry; the entry was appended to the log at 19:50:28, and no analysis step was run in between (project record of the planning step).

Analyses 47 to 50 were planned on 29 September 2026; 04:05:47 is the output of the date command that gives the heading of their plan entry, which was appended to the log at 04:06:41, and no analysis step was run in between (project record of the planning step). The path of an input file of analysis 49 was corrected at 04:06:49 and further sources of analysis 50 were added at 04:16:33, before the steps concerned. Analysis 48 finished its computation at 04:32:53; its end file was written at 04:39:08, when the finished run was checked, and a check of the truncation counts per review against analysis 44 was run at 04:48:20. Analysis 49 was run three times (04:28:55, 04:39:08, and 04:48:16 starts); the second and third runs corrected the check of the location of the 383 comparisons, and the results of the extension were identical in the three runs.

Analysis 51 was planned on 29 September 2026; 06:53:39 is the output of the date command that gives the heading of its plan entry, which was appended by the same command. Its comparison script was run three times (first run 07:01:28); the change before the second run was logged at 07:01:44, and one line that implemented it was added with the command that started the second run (07:01:51) and was recorded afterwards, in the entry of 07:02:15, which also logged the change before the third run.

Analysis 52 was planned on 29 September 2026; 09:07:49 is the output of the date command that gives the heading of its plan entry, which was appended by the same command. Further consistency checks were logged at 09:17:39, before the only run of its main script (09:17:44); a comparison added after the results of that run had been seen was logged at 09:18:08 and run once at 09:18:28.

Further post-hoc analyses (1 to 31) were planned and run on 26 to 28 September 2026 (Supplementary Tables 1 and 2, with the planning and execution time of each), including calibration (ref. 38), a statistical stopping criterion (ref. 8), sensitivity to $\tau$, and the order and composition of requests. They used the model scores and the record files of the original analyses. Analyses 3 and 8 ran new active-learning simulations on these scores (248 and 561 runs). The only new model call was one probe request to the Jev endpoint (analysis 15), which sent a public sentence and no study record and cost US\$0.00001218. The decision criteria, $\tau=0.07$, $\lambda=3$, and the 90% target were not changed.

Analyses 32 to 46 were planned at 19:48 on 28 September 2026 and run between 19:57 and 23:42 on the same day (Supplementary Table 3); two earlier steps, the listing and partial reading of files of the CLEF 2019 repository for analysis 46 (19:42) and one request to Crossref for analysis 45 (19:56), are listed above. They used the stored scores, labeling orders, and record files, the files of the dataset release, public bibliographic records and pages, and a model downloaded from a public repository. No model call was made to any evaluated model; the paid scoring planned in analysis 46 was not run because its condition of feasibility was not met. The decision criteria, $\tau=0.07$, $\lambda=3$, and the 90% target were not changed.

Analyses 47 to 50 were planned at 04:05 on 29 September 2026 and run between 04:14 and 04:57 on the same day (Supplementary Table 3). They used the stored record files and run logs, the times of the project files, the project records of 24 and 25 September 2026, the stored results of analyses 32 to 35, 39, and 44, the text and the record of printed values of an earlier draft of this article and its Supplementary Information, a tokenizer downloaded in analysis 44, and public bibliographic records and pages. No model call was made, and the decision criteria, $\tau=0.07$, $\lambda=3$, and the 90% target were not changed.

Analysis 51 was planned at 06:53 on 29 September 2026 and run between 06:58 and 07:03 on the same day (Supplementary Table 3). It sent two requests to a public repository and used no study data; no model call was made, and the decision criteria, $\tau=0.07$, $\lambda=3$, and the 90% target were not changed.

Analysis 52 was planned at 09:07 on 29 September 2026 and run between 09:15 and 09:19 on the same day (Supplementary Table 3). It used the stored per-run results of analyses 8, 33, and 42; no model call was made, and the decision criteria, $\tau=0.07$, $\lambda=3$, and the 90% target were not changed.

Analyses not performed. The title-only and mismatched-criteria controls were not run on the held-out reviews or on the pilot review, and the post-hoc analyses 1 to 52 made no new model call with the records of the pilot review (the only new model call was the probe request of analysis 15, which sent no study record); the post-hoc conclusion-level analysis of 25 September 2026 (P7) sent the records of 34 included studies of the pilot review to Jev, and one further included study was sent on 9 October 2026 (see above). No re-scoring was done of the 2002-record subset with a finer probability instrument or through the programming interface of Claude Opus 5.5 at temperature 0, of the 18

remaining development reviews, or of repeated scoring of the same records to measure the stability of the scores. These analyses need new model calls; the endpoint reports no version, so new scores might come from a changed model and could not be combined with the original scores. A paired re-scoring of the CLEF records with protocol-stage and abstract-stage criteria in one new run was planned but not run, because too few protocols could be retrieved (analysis 46). No open-weight LLM ranker was run, because the analysis server has no graphics processing unit.

Other post-hoc analyses. The subset for the comparison with LLMs was analyzed for refusals, for the number of distinct values that each model returned, for pooled AUCs, and in the reviews with at least 20 excluded records (analysis 9). Discrimination and threshold outcomes were stratified by publication year and correlated with it (Spearman; analysis 10). Held-out and development reviews were compared with Mann-Whitney tests (analysis 17). The published LLM-guided ranker LGAR (ref. 13) was compared descriptively and without pairing with the values reported in its publication at the same label level (analyses 19 and 27).

**Departures from the plan and early displays.** The settings and analyses departed from the plan in six respects. (1) The plan named a stopping rule S1 that estimated the number of remaining inclusions from calibrated Jev probabilities, with calibration on a random pilot sample of each review. At 19:00 to 19:02 on 24 September 2026, an entry of the plan replaced it by the label-free threshold rule judged under C4; by then, the simulations on the pilot labels described below and the results of 25 small development reviews had been seen, and no metric on the held-out reviews had been computed. The file gives no reason for the replacement. (2) The criteria as written named logistic-regression active learning as the comparator of C3 and C4. The first active-learning runs, on the pilot review and on 25 development reviews, used logistic-regression and naive Bayes active learning; the later development analyses and the held-out decision used the default ASReview model, and this change was recorded in the plan only after the held-out evaluation. With the comparators named in the plan, C3 and C4 led to the same decisions (Supplementary Note 5). (3) The plan described the Jev scores as a starting order and as a feature of active learning; the hybrid as fixed uses the starting order and a blended ranking score, and feature concatenation, evaluated on the development reviews, gave a lower WSS@95 (0.7488 vs. 0.7662). (4) Tuning used the 73 of the 91 development reviews with at most 2000 records, a restriction not in the plan. (5) The plan anticipated 57 reviews with title and abstract labels; the records of 53 SYNERGY+ reviews carry these labels (12 held-out and 41 development reviews; Supplementary Table 7), of which the 12 held-out reviews and the 33 development reviews with at most 2000 records were analyzed. (6) The plan listed cross-encoder rerankers among the free baselines (M2); two cross-encoder rerankers and a further embedding model (e5-base-v2) were evaluated only on the pilot review, after the plan file had been created, and were not run on SYNERGY+ (Methods, Comparators), so the comparator of C1 was chosen from the four free rankers retained, none of which is a cross-encoder; the result of one reranker (0.725) became available 43 s after the development run of these four had been started.

Summary AUCs of three free rankers on the held-out reviews (TF-IDF, BM25, and MiniLM) and the AUCs of all four free rankers in one held-out review were displayed before the free-ranker comparator, $\lambda$, and $\tau$ were fixed on the development results, and the scoring of the held-out records started about 11 min before that entry; the scores of Jev do not depend on these settings, and the results of Jev on the held-out reviews were first viewed after the entry. Of the settings taken from the pilot review, only the prompt of Jev was fixed before the plan file was created; the batch size and the free-ranker shortlist were chosen afterwards from results on the pilot labels, before any metric on SYNERGY+ records was computed. Score thresholds from 0.05 to 0.5 had been evaluated on the pilot labels before the plan file was created, and hybrid variants and stopping rules, including the threshold rule at thresholds from 0.01 to 0.3, were simulated there before the plan named the threshold rule; $\lambda$ and $\tau$ were chosen afterwards on the development reviews. Supplementary Note 1 gives the times of these steps.

**Model access and vendor documentation.** All scoring calls were made on 24 and 25 September 2026; one probe request to the Jev endpoint, without study records, was made on 26 September 2026 (analysis 15). Jev (typesafe-ai/jev), GPT-4o-mini (openai/gpt-4o-mini), and DeepSeek-V3.1 (deepseek/deepseek-v3.1) were called through the Vercel AI Gateway, Jev through the endpoint https://ai-gateway.vercel.sh/typesafe/v1/systemone. Claude Opus 5.5 (claude-opus-5-5) was called through Anthropic's command-line interface, Claude Code (version 2.1.281), with tools, settings files, and external servers disabled and a replaced system prompt; all calls to Claude Opus 5.5 were completed by 00:58 on 25 September 2026. The version is established from the installation times of its files (version 2.1.281 installed at 04:51 on 24 September; version 2.1.282 installed at 08:57 on 25 September) and from the version field of the project records. No transcript of these calls was kept, and the number of input tokens of the replaced system prompt was not measured in the screening runs.

In a probe request on 26 September 2026, which sent one public sentence and no study record, the response of the Jev endpoint contained the fields model, answers, usage, and provider\_metadata; the model field held the alias typesafe-ai/jev, and neither the body nor the headers contained a version or build string. The gateway lists two providers for the model, and the provider that served each request was not recorded. The vendor documentation lists a versioned identifier (jev-1.13.0) with two aliases for its own interface and states that an alias moves when a new release ships; this documentation was present in archived copies dated 21 to 25 September 2026. The version that served the requests of this study is not recorded in any output of the study, so re-querying the model may not reproduce the scores; the stored per-record responses are the reproducible record of the primary analysis. The scoring script stores the value returned by the endpoint without rounding, and every stored score is a multiple of 0.01 (93 to 98 distinct values per output file, and 64 in the mismatched-criteria control; post hoc, analysis 32 of Supplementary Table 3), so $\tau=0.07$ and the lowest score of an included study below it (0.03) are single output levels.

The vendor does not disclose the model size, the training data, or a knowledge cut-off. It states that English is the primary training language and that accuracy is lower in other languages, that the model is not fine-tuned with customer data, and that it is not trained on customer requests or responses; the vendor's legal documents give no retention period in days for inputs. The gateway documentation states that prompts and responses are not used for training. The vendor describes the model as calibrated ("higher confidence means higher accuracy") and defines calibration over groups of predictions. Its published evaluation uses the average predictions of two LLMs, GPT-6 Astra and Claude Fable 5.1, as reference probabilities ("We use the average of GPT-6 Astra and Fable 5.1 as the reference answer, which biases answers toward OpenAI and Anthropic's models.") and does not evaluate against observed outcomes of an external labeled task (post hoc, analysis 15). Claude Opus 5.5 has the same developer as Claude Fable 5.1. No provider of the model on the gateway offers zero data retention. The vendor announcement is cited with its archived copy of 26 September 2026, whose text is identical to the copy of the page saved on the analysis server on the same day; the page was edited after its release (post hoc, analysis 24; Supplementary Note 1).

**Procedures of selected post-hoc analyses.** The Supplementary Notes describe each post-hoc analysis with its results, and Supplementary Note 1 gives the times at which each was planned and run and lists the steps that were executed before they were logged or without a prior log entry. Scoring modes (analysis 36): in the 2002-record subset, records were cross-tabulated by their side of $\tau$ under single-record requests, ten-record batches formed within the subset, and the ten-record batches of the full run, and recall at $\tau$ was computed per review under each mode. Out-of-sample reliability of $\tau$ (analysis 37): in each of 5000 bootstrap resamples of the 73 development reviews, $\tau$ was selected with the rule of the plan and applied to the development reviews not drawn in that resample. Comparison with LLMs (analysis 38): a two-stage percentile bootstrap resampled reviews and then the excluded records within each drawn review, and in a sensitivity analysis the three records refused by Claude Opus 5.5 were given a score of 0 or 1. Single-run reliability (analysis 52): each run of a review (a random start, a tie-breaking seed, or a random order) was counted as reaching 95% recall when at least $\lceil 0.95n \rceil$ of its $n$ included records had been found; no interval or test was computed, because runs are nested within reviews. The post-hoc analyses 1 to 52 used the stored scores of the evaluated models (analysis 44 also scored the records with a locally run MedCPT cross-encoder) and made no new request to the evaluated models, apart from one probe request to the Jev endpoint that sent one public sentence and no study record (analysis 15). The CLEF validation, the comparison of scoring modes in the 2002-record subset, the mismatched-criteria control, and the conclusion-level analysis of the pilot review, for which 34 of the 35 included studies of the pilot review from other databases were scored with single-record requests from 09:52 to 09:55 (the 35th on 9 October 2026), used Jev scores from runs completed on 25 September 2026 after the held-out decision (Supplementary Note 1), on scoring occasions different from that of the held-out run; the model version and the provider that served these requests were not recorded.

**Provenance of the model inputs.** Claude Code (Anthropic), a command-line AI coding tool (versions 2.1.281 and 2.1.282, with the models Claude Fable 5.1, claude-fable-5-1, and Claude Opus 5.5, claude-opus-5-5), was used on the analysis server to write the texts sent to the evaluated models: the criteria block of the pilot review, from the eligibility criteria in supplementary file S2 of the submitted manuscript of that review; the Jev question, the batch preamble, and the two rephrasings of the question; the prompt templates and system prompts of the LLMs; and the inputs of the two negative controls. In the session records of the tool, each of these texts first appears in a tool call of the model, and no message typed by a person contains it (records read on 27 September 2026). The Jev question, the verbalized-probability system prompt of the LLMs, the criteria block of the pilot review, and the batch preamble were first written while the tool used Claude Table 5.1, one of the two reference models of the vendor's evaluation (Methods, Model, access, and provenance); the criteria-block template of the SYNERGY+ inputs, the log-probability prompt of GPT-4o-mini, the rephrasings, the inputs of the controls, and the criteria sections of the Cochrane reviews were first written while it used Claude Opus 5.5. Supplementary Note 1 gives the periods in which each model was used. Claude Opus 5.5 was also an evaluated comparator; its scores were obtained through separate calls of the command-line interface without tools.

The tool was also used to write all analysis code, including the scripts that took the criteria of the SYNERGY+ reviews from the dataset and those of the Cochrane reviews from their abstracts; to run the analyses, including the post-hoc analyses 1 to 52, and the literature searches; to retrieve and check the references; and to produce the figures. The failure categories and the study-level reading of the CLEF content-level label in three Cochrane reviews (Supplementary Note 7) and the coding of prior studies (Supplementary Note 22) were proposed with the tool and are reported only in the Supplementary Information. "Project records" in this Supplementary Information are the session records of the tool and the files of the project.

#### Supplementary Note 2. Cost and throughput (compiled post hoc)

**Supplementary Table 4. Cost of scoring, tokens, and list prices (compiled post hoc from the run logs)**

| Run | Records scored | Cost of the run (US\$) | Cost per 1000 records (US\$) | Mean input tokens per record | List price (US\$ per million tokens) |
| --- | --- | --- | --- | --- | --- |
| Jev, development reviews, ten-record batches | 52,957 | 1.1134 | 0.0210 | 500.6 | 0.042 input; output free |
| Jev, held-out reviews, ten-record batches | 33,001 | 0.6634 | 0.0201 | 478.6 | 0.042 input; output free |
| Jev, CLEF 2019 reviews, ten-record batches | 82,418 | 1.5442 | 0.0187 | 446.1 | 0.042 input; output free |
| Jev, 2002-record subset, ten-record batches | 2002 | 0.0404 | 0.0202 | 480.2 | 0.042 input; output free |
| Jev, 2002-record subset, single-record requests | 2002 | 0.0845 | 0.0422 | 1005.1 | 0.042 input; output free |
| GPT-4o-mini, 2002-record subset, single-record requests | 2002 | 0.2220* | 0.1109 | not recorded | 0.15 input; 0.60 output |
| DeepSeek-V3.1, 2002-record subset, single-record requests | 2002 | 0.3204 | 0.1600 | not recorded | 0.25 input; 0.95 output (varies by provider) |
| Claude Opus 5.5, 2002-record subset, command-line interface (2002 attempted, 1999 scored) | 1999 | 29.3337 | 14.6522 | not recorded | 4.00 input; 20.00 output |
| Claude Opus 5.5, pilot review (PubMed arm), command-line interface | 1572 | 22.6903 | 14.4340 | not recorded | 4.00 input; 20.00 output |

Costs are those reported by the gateway, except for Claude Opus 5.5, for which the cost is the list-price equivalent reported by the command-line interface under a subscription and is not an itemized invoice. For Claude Opus 5.5 on the 2002-record subset the cost per 1000 records is per 1000 attempted records. Token counts of the chat models were not recorded by the scoring script. List prices are from the public gateway listing retrieved on 26 September 2026. \*Sum of US\$0.2133 for the resumed run (1957 records; run log) and \$0.0087 for a first run that was interrupted after 50 attempted records, of which 45 were kept and five had failed. The cost of the first run is the value of one line of a progress record that was written every two minutes (19:00 on 24 September 2026); the run was restarted by 19:02, and any cost between the two times is not recorded. With the resumed run alone, the cost was \$0.1090 per 1000 records.

The cost of Jev per 1000 records was US\$0.021 in the development run, \$0.020 in the held-out run, and \$0.019 in the CLEF run. The batched cost divides the criteria block over ten records; with single-record requests the cost was \$0.042 per 1000 records. For the five Jev runs in Supplementary Table 4, the number of input tokens multiplied by the list price reproduces the logged cost. For the single-record scoring of the pilot review, the logged cost (US\$0.056 for 1572 records) is lower than the token count multiplied by the list price (\$0.064); the reason is not recorded, and we do not report a cost for that run. Relative to the batched cost of Jev in the full held-out run (US\$0.0201 per 1000 records), the costs of GPT-4o-mini, DeepSeek-V3.1, and Claude Opus 5.5 were higher by factors of 5.5, 8.0, and 729; relative to the batched cost of Jev in the 2002-record subset (\$0.0202), by factors of 5.5, 7.9, and 725; and relative to its single-record requests in the subset (\$0.0422), by factors of 2.6, 3.8, and 347 (post hoc, analysis 32). The cost of GPT-4o-mini in these factors is the sum of two runs, of which the first is known only from a progress record (footnote to Supplementary Table 4); with the fully recorded run alone the factors for GPT-4o-mini are 5.4 and 2.6. In absolute terms, GPT-4o-mini cost US\$0.0908 per 1000 records more than Jev in ten-record batches of the full held-out run and \$0.0687 more than Jev with single-record requests in the subset. The Claude Opus 5.5 run on the PubMed arm of the pilot review cost US\$22.69 by the report of the interface (\$14.43 per 1000 records). The price of Jev is a launch price in early access. The vendor's announcement states: "Cost per call: We make our pricing transparent. We can't prove it isn't subsidized; we'll need the long-term to prove the sustainability of our pricing (which we expect to go down, not up)." (vendor announcement, archived copy of 26 September 2026; the capture of 15 September 2026, contains the same sentence; post hoc, analysis 30 of Supplementary Table 2). The expected fall in prices is a statement of the vendor. The cost of the LLMs with ten-record batches, prompt caching, or batch pricing was not measured; the vendors offer prompt caching (GPT-4o-mini and Claude Opus 5.5) and a batch discount of 50% (Claude Opus 5.5), which could narrow the difference. The costs exclude retrieval of records, the computing time of the free rankers and simulations, and human time.

Cost and records not read (post hoc, analysis 32). Scoring all records with Jev cost US\$0.66 for the 33,001 held-out records, \$1.54 for the 82,418 CLEF records, and \$1.11 for the 52,957 development records. In the held-out reviews, one percentage point of the records corresponds to 330 records; the threshold read 16,831 records and the knee method as adapted 26,180 (mean over random starts), a difference of 9349 records, and under the statistical criterion the Jev ranking read 25,364 and the ASReview ranking 27,075 records, a difference of 1711. In the 28 CLEF reviews with content-level labels (77,611 records), the corresponding differences were 24,658 and 1021 records. No published per-record time of human title and abstract screening was found in a recorded search (Supplementary Note 22). As an illustration only, at the rate of two abstracts per minute that ref. 24 states for an experienced reviewer, without citing a source, and noting that abstracts on difficult topics may take several minutes each, one percentage point of the held-out records (330 records) corresponds to about 2.75 h of one reviewer's time (post hoc, analysis 50); the rate was not measured in this study.

**Supplementary Table 5. Throughput of the scoring runs (compiled post hoc from the run logs and file times)**

| Run | Records | Wall-clock time (h) | Records per minute |
| --- | --- | --- | --- |
| Jev, development reviews | 52,957 | 6.52 | 135 |
| Jev, held-out reviews | 33,001 | 2.68 | 205 |
| Jev, first rephrasing (12 development reviews) | 5550 | 0.39 | 238 |
| Jev, second rephrasing | 5550 | 0.36 | 256 |
| Jev, control with the review title only | 5550 | 0.39 | 239 |
| Jev, control with mismatched criteria | 5550 | 0.39 | 234 |
| Jev, CLEF 2019 reviews | 82,418 | 6.40 | 215 |
| Claude Opus 5.5, 2002-record subset | 2002 | 0.75 | 44.3 |
| DeepSeek-V3.1, 2002-record subset | 2002 | 5.02 | 6.6 |
| GPT-4o-mini, 2002-record subset (resumed run) | 1957 | 9.81 | 3.3 |
| Jev, 2002-record subset, ten-record batches (times not recorded) | 2002 | .. | .. |

Times and rates were computed from the recorded start and end times. End times are file times. Start times of the Jev runs are the times at which the chain of commands started each run; for two runs the log file was created 3 s and 19 s later. Start times of the three chat-model runs were read from a progress record written every one to two minutes and are uncertain by about 1 min (Claude Opus 5.5), up to 8 min (DeepSeek-V3.1), and about 2 min (GPT-4o-mini). These uncertainties correspond to 43 to 45 records per minute for Claude Opus 5.5 and to 6.5 to 6.8 for DeepSeek-V3.1, and they leave the value for GPT-4o-mini unchanged at the printed precision. Jev requests were paced at one request per 1.9 s with ten records per request; chat models were limited by the gateway to about five requests per minute per model. Times exclude retrieval, the free rankers, the simulations, and human time.

##### Supplementary Note 3. Order of records and composition of requests (post hoc)

This note reports analyses 22, 28, and 29 of Supplementary Table 2 (methods at the end of this note). The requests rebuilt from the output files equal the consecutive slices of the data files in all 5328 development, 3308 held-out, and 8257 CLEF requests. The number of requests with two or more positive records did not exceed its expectation under random order within reviews with one-sided $P < 0.05$ in any collection at either label level (Supplementary Table 6), which was the criterion for clustering specified for this analysis; in the CLEF reviews (content-level label) there were fewer such requests than expected (110 versus 124.7; two-sided $P = 0.041$). Pairs of positive records in the same request gave observed-to-expected ratios of 0.93 to 1.04. The position of included records in the data files did not differ from that of excluded records in most reviews (median AUC of file position 0.494, 0.492, and 0.489 in development, held-out, and CLEF reviews; $P < 0.05$ in four of 73, three of 23, and three of 28 reviews, without adjustment for multiplicity). The plan of this analysis also named a descriptive comparison of scores. Within each review, the mean Jev score of positive records that shared their request with another positive record minus that of positive records without such a partner was, averaged over reviews with both kinds, 0.003 in 52 development reviews, $-0.0001$ in 17 held-out reviews, and $-0.068$ in 14 CLEF reviews at the level of final inclusion (content-level label for CLEF), and $-0.001$, $-0.014$, and $-0.063$ in 30, ten, and 24 reviews against title and abstract labels. This comparison is confounded: positive records that share a request with another positive record may differ in topic or in the content of their abstracts from those that do not, and the comparison does not measure the effect of the composition of a request on the scores. That effect, and thus its effect on $\tau$, could be isolated only with new model calls in which the records that share a request vary within one scoring occasion; the two ten-record batched modes of the 2002-record subset (Supplementary Table 35) differ in these records but also in the scoring occasion and possibly the provider (analysis 47), so the effect was not isolated.

###### Supplementary Table 6. Requests with two or more positive records compared with random order of records within reviews (post hoc)

| Collection | Label | Reviews (with at least two positive records) | Records | Positive records | Requests | Requests with at least two positive records | Expected under random order | Ratio, observed to expected | $P$ for an excess | $P$, two-sided |
| --- | --- | --- | --- | --- | --- | --- | --- | --- | --- | --- |
| Development | Final inclusion | 73 (73) | 52,957 | 2048 | 5328 | 386 | 390.5 | 0.989 | 0.683 | 0.706 |
| Held-out | Final inclusion | 23 (23) | 33,001 | 597 | 3308 | 79 | 87.5 | 0.903 | 0.942 | 0.163 |
| CLEF 2019 | Content-level label | 31 (27) | 82,418 | 882 | 8257 | 110 | 124.7 | 0.882 | 0.984 | 0.041 |
| Development | Title and abstract | 33 (33) | 20,487 | 2484 | 2064 | 643 | 648.4 | 0.992 | 0.705 | 0.656 |
| Held-out | Title and abstract | 12 (12) | 20,366 | 948 | 2041 | 210 | 201.2 | 1.044 | 0.115 | 0.231 |
| CLEF 2019 | Title and abstract | 31 (31) | 82,418 | 1682 | 8257 | 268 | 285.4 | 0.939 | 0.969 | 0.078 |

Requests are the ten-record requests of the scoring runs, rebuilt from the output files; they equal the consecutive slices of ten records of each review in the order of the data file (the last slice of a review is shorter). The expected number assumes that the records of each review were in random order, with the sizes of the requests fixed (hypergeometric distribution). $P$ values (one-sided for an excess, and two-sided) come from 10,000 random permutations of the records within each review (seed 20260927). Positive records are included studies (SYNERGY+), content-relevant records (CLEF 2019), or records that passed title and abstract screening. No adjustment was made for multiplicity.

Direction of the differences in file position (analysis 28). In the ten reviews in which the position of included records in the data file differed from that of excluded records with $P < 0.05$, the included records lay earlier in all four development reviews (AUC of file position 0.139 in Deckers 2022, 0.335 in de Matteis 2024b, 0.347 in Filges 2018, and 0.411 in Fejza 2023), in one of three held-out reviews (0.279 in Eggmann 2023; later in Bosch 2021, 0.644, and Lewowski 2021, 0.726), and in two of three CLEF reviews (0.150 in CD012567 and 0.237 in CD004414; later in CD012661, 0.569).

Order of the data file among tied scores (analysis 28). The stored Jev scores have one or two decimals, so records of a review often share a score, and when ties are kept in the order of the data file, that order decides which record of a tie group is read first. Of the pairs of an included and an excluded record with the same score in the same review, the included record came first in the data file in 1231 of 2621 pairs in the held-out reviews (0.470), 4106 of 7709 in the development reviews (0.533), and 3748 of 7781 in the CLEF reviews (content-level label; 0.482), and in 15,755 of 32,446 and 12,730 of 28,146 pairs against title and abstract labels in the held-out and CLEF reviews (0.486 and 0.452). The order of the data file therefore favored included records among tied scores in some reviews and in the development reviews overall, but not in the held-out or CLEF reviews overall. With ties in the order of the data file, the WSS@95 of the Jev ranking exceeded that with random ties by 0.00145 in the held-out reviews (0.7882 versus 0.7868; Supplementary Note 5); the three held-out reviews with $P < 0.05$ contributed 0.0000025 of this difference, which came mainly from reviews in which file position did not differ at $P < 0.05$ (largest difference within a review 0.011, in Menon 2022), and the sign of the difference in a review did not follow the order of its tied pairs. The difference was 0.00198 in the development reviews, $-0.00015$ in the CLEF reviews (content-level label), and $-0.00357$ and $-0.00062$ against title and abstract labels in the held-out and CLEF reviews. Random tie-breaking removes the dependence of the ranking on the order of the data file; the main article reports the Jev ranking with random ties.

Positive records below $\tau$ (analysis 29). Of the eight positive records of the evaluation collections with a score below $\tau = 0.07$ (three in Supplementary Table 12 and five in Supplementary Table 13), four shared their ten-record request with another positive record, against 4.06 expected under random order within reviews (probability of four or more, 0.70): one of the three held-out records (1.34 expected) and three of the five CLEF records, all in CD011977 (2.72 expected). Every partner record had a score at or above $\tau$, and no two records below $\tau$ shared a request. Of the nine development records below $\tau$, five shared a request (4.42 expected). Of all positive records, 429 of 1479 in the evaluation collections (0.290) and 913 of 2048 in the development reviews (0.446) shared a request with another positive record, against expected proportions of 0.318 and 0.454. Eight records carry little information, and sharing a request is not a measure of the effect of request composition on scores.

Use of $\tau$. The effect of request composition on scores, and hence on $\tau$, was not isolated from differences between scoring occasions and providers (Supplementary Table 35); the difference of $-0.068$ in 14 CLEF reviews reported above is descriptive and confounded. The condition under which $\tau=0.07$ was evaluated is ten-record requests formed from the records of one review in an order that is not sorted by relevance or by any score. In this study, that order was the order of the data files, which did not cluster positive records beyond random order within reviews (Supplementary Table 6); how the source files themselves were ordered is not documented. The export order of a bibliographic database or a reference manager may follow a relevance ranking (for example, a default relevance sort of the database); such an order is a form of pre-ranking that was not evaluated and should not be used as it stands, and records should not be sorted or pre-ranked by relevance or by any score before requests are formed. Random shuffling of the records within a review would give an order without such clustering, but it was not itself evaluated.

Order of records and composition of requests (analyses 22, 28, and 29). The requests were rebuilt from the output files and compared with the consecutive slices of the data files. For each collection and label level, the number of requests with two or more positive records was compared with its expectation under random order of the records within reviews, with the sizes of the requests fixed (hypergeometric distribution), using 10,000 random permutations; the criterion for clustering specified for this analysis was an excess with one-sided $P<0.05$. The position of positive records in the data files was summarized by the AUC of file position within each review. The plan of this analysis also named a descriptive comparison of the mean Jev score of positive records that shared their request with another positive record and of those without such a partner; this comparison is confounded, because such records may differ in topic or in the content of their abstracts, and it does not measure the effect of the composition of a request on the scores. That effect, and thus its effect on $\tau$, could be isolated only with new model calls in which the records that share a request vary within one scoring occasion; the two ten-record batched modes of the 2002-record subset (analysis 36) differ in these records but also in the scoring occasion and possibly the provider (analysis 47), so the effect was not isolated. Analysis 28 examined the direction of the differences in file position and the order of the data file among records with tied scores, and analysis 29 the request composition of the positive records below $\tau$.

Scoring times (analysis 47). For each of the three scoring modes of the 2002-record subset, the start and end of scoring were taken from the creation and modification times of the output and log files, the header, progress, and completion lines of the logs, the scoring commands, and the project records of 24 and 25 September 2026; a time was reported as not recorded where none of these established it, and the output files and the scoring script were searched for a record of the provider. No score or label was read. On 25 September 2026, the full run was scored from 00:48:48 to no later than 03:29:32, the single-record requests from 09:15:48 to 10:28:38, and the subset batches from 10:28:38 to 10:39:29.

**Composition of the requests.** For Jev, the scoring script grouped the records of each review and cut them into consecutive slices of ten records in the order of the data file; the last slice of a review is shorter (a single record when one record remained), and records whose request failed were grouped again when the script was re-run. Within each review, the data files keep the record order of the source files: the row order of the SYNERGY+ source files in all 73 development and 23 held-out reviews, and the line order of the content-level relevance file of CLEF 2019 in all 31 reviews; in no review are the records sorted by their identifiers, and how the source files themselves were ordered is not documented. Ten-record batches were used for the development reviews (52,620 of 52,957 records in batches of exactly ten), the held-out reviews (32,880 of 33,001), the CLEF records (82,280 of 82,418), the rephrasings and controls (5490 of 5550), and the PubMed arm of the pilot review in the threshold and conclusion-level analyses (1570 of 1572). Apart from these remainders, single-record requests were used only for the first scoring of the pilot review, for the 35 included studies of the pilot review from other databases, and for the comparison of scoring modes in the 2002-record subset. The AUC of Jev in the comparison with LLMs used the batched scores of the full held-out run.

#### Supplementary Note 4. Development and held-out reviews of SYNERGY+

**Supplementary Table 7. Development and held-out reviews of SYNERGY+ 3.0**

| Set | Reviews | Records per review, median (range) | Inclusion rate, median (range) | Publication year, median (range) | Reviews with title and abstract labels | Reviews flagged in the metadata as having these labels | Length of criteria, median (characters) |
| --- | --- | --- | --- | --- | --- | --- | --- |
| Held-out reviews | 23 | 840 (70 to 11,735) | 2.6% (0.3 to 12.9) | 2022 (2011 to 2024) | 12 | 12 | 973 |
| Development reviews used for tuning | 73 | 549 (54 to 1978) | 2.8% (0.5 to 23.8) | 2022 (2015 to 2024) | 33 | 37 | 939 |
| All development reviews | 91 | 714 (54 to 11,440) | 2.6% (0.1 to 23.8) | 2022 (2015 to 2025) | 41 | 45 | 930 |

The column of reviews with title and abstract labels counts the reviews whose records carry these labels. The metadata of the dataset flags 57 reviews, which is the number anticipated in the plan. The records of five flagged reviews carry no such labels (Adamo 2021, Bos 2018, Rinne 2021, Sep 2021, and Wassenaar 2017), and the records of one review without the flag carry them (Walker 2018).

In comparisons of the held-out reviews with the 73 development reviews used for tuning, Mann-Whitney tests gave $P=0.13$ for the number of records, $P=0.26$ for the inclusion rate, $P=0.92$ for the publication year, and $P=0.51$ for the length of the criteria (post hoc). The 18 development reviews that were not scored had a median of 3271 records (range 2265 to 11,440) and a median inclusion rate of 1.1%. The reason they were not scored is given in the Methods (Data sources).

Share of the release in the exported data (post hoc, analysis 41). Supplementary Table 8 gives, for each held-out review, the records and included studies of the release and of the exported data, with recall and the proportion read at $\tau$.

**Supplementary Table 8. Share of the SYNERGY+ release retained in the exported data, per**

#### held-out review (post hoc, analysis 41)

| Review | Records, export | Records, release | Share of records | Included, export | Included, release | Share of included | Recall at $\tau$ | Records read at $\tau$ |
| --- | --- | --- | --- | --- | --- | --- | --- | --- |
| Abgaz 2023 | 288 | 733 | 39.3% | 11 | 33 | 33.3% | 1.000 | 80.2% |
| Attai 2022 | 1726 | 3798 | 45.4% | 135 | 235 | 57.4% | 0.985 | 47.0% |
| Boersma-van Dam 2024 | 578 | 1961 | 29.5% | 24 | 102 | 23.5% | 0.958 | 20.8% |
| Bos 2018 | 2241 | 5072 | 44.2% | 6 | 10 | 60.0% | 1.000 | 42.1% |
| Bosch 2021 | 1716 | 5299 | 32.4% | 22 | 65 | 33.8% | 1.000 | 52.5% |
| Chakraborty 2023 | 2233 | 10,771 | 20.7% | 7 | 28 | 25.0% | 1.000 | 97.1% |
| Chueca 2023 | 609 | 2763 | 22.0% | 16 | 90 | 17.8% | 1.000 | 86.9% |
| Donners 2021 | 119 | 260 | 45.8% | 14 | 15 | 93.3% | 1.000 | 71.4% |
| Eggmann 2023 | 266 | 828 | 32.1% | 9 | 23 | 39.1% | 1.000 | 53.8% |
| Hall 2011 | 340 | 8669 | 3.9% | 4 | 104 | 3.8% | 1.000 | 12.9% |
| Harms 2024 | 840 | 1548 | 54.3% | 48 | 71 | 67.6% | 1.000 | 92.9% |
| Kerschbaumer 2024b | 1146 | 1691 | 67.8% | 15 | 21 | 71.4% | 1.000 | 90.5% |
| Lewowski 2021 | 329 | 1642 | 20.0% | 9 | 45 | 20.0% | 1.000 | 37.7% |
| Menon 2022 | 485 | 950 | 51.1% | 35 | 74 | 47.3% | 1.000 | 87.6% |
| Moran 2020 | 1992 | 5244 | 38.0% | 39 | 142 | 27.5% | 1.000 | 34.3% |
| Parodis 2023b | 1679 | 3923 | 42.8% | 33 | 78 | 42.3% | 1.000 | 85.1% |
| Roberts 2021 | 883 | 4304 | 20.5% | 3 | 24 | 12.5% | 1.000 | 31.5% |
| Seghers 2022 | 1759 | 4522 | 38.9% | 16 | 28 | 57.1% | 1.000 | 54.3% |
| Theobald 2021 | 70 | 351 | 19.9% | 9 | 35 | 25.7% | 1.000 | 75.7% |
| Toffalini 2021 | 328 | 725 | 45.2% | 22 | 37 | 59.5% | 1.000 | 50.3% |
| Tumkaya 2018 | 481 | 721 | 66.7% | 39 | 52 | 75.0% | 1.000 | 51.8% |
| van der Waal 2022 | 1158 | 2148 | 53.9% | 17 | 33 | 51.5% | 1.000 | 64.2% |
| Walker 2018 | 11,735 | 47,131 | 24.9% | 64 | 148 | 43.2% | 1.000 | 33.5% |
| All 23 reviews | 33,001 | 115,054 | 28.7% | 597 | 1493 | 40.0% | .. | .. |

The share of records retained had a median of 38.9% (range 3.9% to 67.8%) and the share of included studies a median of 42.3% (3.8% to 93.3%). Spearman correlations over the 23 held-out reviews: share of records with recall at $\tau$ ($\rho=-0.01$, $P=0.949$), with the proportion of records read at $\tau$ ($\rho=0.40$, $P=0.058$), and with the AUC of Jev ($\rho=-0.36$, $P=0.096$); share of included studies with recall at $\tau$ ($\rho=0.06$, $P=0.791$), with the proportion read ($\rho=0.31$, $P=0.150$), and with the AUC ($\rho=-0.24$, $P=0.268$). Recall at $\tau$ was 1 in 21 of 23 reviews, so its correlations rest on little variation.

Development reviews (the secondary set of the plan of this analysis; 73 reviews with at most 2000 records, per-review values in the result files): the exported data held 52,957 of 166,391 records and 2048 of 5544 included studies of the release; the share of records retained had a median of 33.1% (range 15.3% to 70.8%) and the share of included studies a median of 36.4% (10.9% to 95.7%). Spearman correlations over the 73 development reviews: share of records with recall at $\tau$ ($\rho=0.03$, $P=0.785$), with the proportion of records read at $\tau$ ($\rho=0.32$, $P=0.0065$), and with the AUC of Jev ($\rho=-0.22$, $P=0.066$); share of included studies with recall at $\tau$ ($\rho=0.02$, $P=0.845$), with the proportion read ($\rho=0.17$, $P=0.153$), and with the AUC ($\rho=-0.03$, $P=0.824$). Recall at $\tau$ was below 1 in 5 and below 0.95 in one of these reviews. The correlation of the share of records with the proportion read at $\tau$ was 0.40 in held-out and 0.32 in development reviews; one of the twelve correlations of the two sets had $P<0.05$ (that of the development reviews for the share of records and the proportion read), and none was adjusted for multiplicity.

The counting reproduced the totals of the release for the 114 reviews (478,478 records and 10,679 included studies). The correlations are observational across reviews and do not show what the omitted records would have given. *P* values are exploratory.

Share of the release in the export (analysis 41). The records and included studies of each held-out and development review in the dataset release were counted from the files of the release, not through the dataset package, and compared with the exported data; Spearman correlations of the retained shares with recall and the proportion read at $\tau$ and with the AUC of Jev were computed in the held-out reviews and, as a secondary set, in the 73 development reviews.

SYNERGY+. The primary data source was SYNERGY+ (version 3.0, release synergy\_plus\_v3.0 of 27 August 2026) (ref. 30), an open collection of systematic reviews with search results, final inclusion labels, and eligibility criteria. It was downloaded with the synergy-dataset package (version 2.2), whose default export for SYNERGY+ contains only open-access records with a valid abstract and omits reviews with fewer than three such included records. The exported data held 114 of the 119 reviews of the release, with 168,969 records and 4399 included studies; for the same 114 reviews, the release holds 478,478 records and 10,679 included studies, so the exported data are 35.3% of the records and 41.2% of the included studies (held-out reviews: 33,001 of 115,054 records and 597 of 1493 included studies). All results on SYNERGY+ refer to this subset, in which no record lacked an abstract. The analysis code calls the dataset SYNERGY 3.0. The 2023 deposit of the first release (26 reviews) is a different release. This release corresponds to the DataverseNL record cited (version 3.0, released 27 August 2026; CC0 license). The official split of the package, a fixed list described there as the official split, assigns 91 development and 23 held-out reviews; neither the package, the repository, nor the release notes describe how the split was formed. Development analyses used the 73 development reviews with at most 2000 records (Table 1), to limit gateway usage; the 18 development reviews that were not scored were not scored later, because new scores from a possibly changed model version could not be combined with the original scores to re-select $\tau$. The primary label was final inclusion; 12 held-out reviews also report title and abstract decisions.

Eligibility criteria. The criteria were taken verbatim from the review metadata of SYNERGY+. The dataset stores the criteria with the publication record of each review, next to the DOI of the review paper, and its documentation does not state that the text was taken from protocols; the dataset therefore attributes the criteria to the review publications. The file of reviews of the dataset has a column named Protocol, without a definition in the documentation available to us, which equals 1 for eight of the 23 held-out reviews and 32 of the 73 development reviews used for tuning (39 of all 91 development reviews); because its meaning is not documented, whether any criteria text was drafted at the protocol stage cannot be determined. The median length of the criteria was 973 characters (range 220 to 2172) in the held-out reviews and 939 characters (148 to 2756) in the development reviews used for tuning.

#### Supplementary Note 5. Supplementary results from the original analyses

Development reviews (73). Jev macro-AUC 0.945 versus bge-base 0.847 (difference 0.099, 0.076–0.123; Jev higher in 66). Hybrid WSS@95 0.766 versus ASReview 0.607 (difference 0.159, 0.119–0.203; 65 of 73). Threshold $\tau=0.07$: reliability 0.986 (72 of 73), records read 58.3%.

Held-out reviews, all active-learning variants (WSS@95 / WSS@100). ASReview default 0.659 / 0.639; ASReview with random start 0.601 / 0.583; Jev starting order only 0.654 / 0.637; hybrid $\lambda=2$ 0.8055 / 0.817; hybrid $\lambda=3$ 0.8055 / 0.821 (0.80548 and 0.80551 for WSS@95 at five decimals); LLM-guided weak supervision 0.746 / 0.728 (post hoc); ASReview strongest preset 0.724 / 0.702 (post hoc); strongest preset plus Jev ($\lambda=3$) 0.806 / 0.817 (post hoc). Comparators named in the plan (logistic regression and naive Bayes active learning implemented as in the plan, retraining after every N/200 records, ten seeds): WSS@95 0.638 and 0.633, WSS@100 0.613 and 0.598. The hybrid exceeded them by 0.167 (0.105–0.235) and 0.172 (0.108–0.239), higher in 21 of 23 reviews ($P<0.0001$ for both); WSS@100 differences 0.208 and 0.224 (CIs in Supplementary Note 19). The knee method on these rankings reached 95% recall in 21 of 23 reviews (91.3%) while reading 96.1% and 96.3% of records. C3 and C4 were therefore met with the comparators named in the plan as well.

Label-free Jev ranking. With tied scores kept in the order of the data file, the ranking by Jev alone gave a WSS@95 of 0.788, a WSS@100 of 0.801, a mean average precision of 0.513, and a true negative rate at 95% recall of 0.869. With ties broken at random (seeds 0 to 9), the values are 0.7868 (range over seeds 0.7849 to 0.7877), 0.8005 (0.7973 to 0.8022), 0.5115 (0.5097 to 0.5125), and 0.8674 (0.8654 to 0.8685). We report the values with random ties (Supplementary Note 10).

Post-hoc comparisons on held-out reviews. Hybrid versus weak supervision, WSS@95 difference 0.059 (0.022–0.106), 21 of 23, $P=0.0002$; WSS@100 difference 0.093 (0.050–0.146). Weak supervision versus ASReview 0.087 (0.042–0.139). Strongest preset plus Jev versus strongest preset 0.082 (0.045–0.128), 21 of 23, $P=0.0001$; WSS@100 0.115 (0.061–0.175).

Batch effects (2002-record subset; post hoc). Macro-AUC 0.9601 for single-record requests, 0.9615 for ten-record batches from the full run (the inclusion rate of the full run was 1.8%), and 0.9625 for ten-record batches formed within the subset (inclusion rate 29.8%); single minus full-run batches $-0.0014$ ($-0.0049$ to $0.0023$; $P=0.30$); subset batches minus full-run batches $0.0010$ ($-0.0022$ to $0.0045$). Mean score for included records 0.595 (single), 0.632 (subset batches), and 0.652 (full-run batches); for excluded records 0.116, 0.122, and 0.130. The mean within-review Spearman correlation between single-record and full-run scores was 0.936. The threshold $\tau=0.07$ was derived from batched scores and should not be transferred to single-record scoring without re-derivation.

Title and abstract labels (post hoc). Development (33 reviews): Jev AUC 0.872 (0.944 for final inclusion in the same reviews), bge-base 0.789. Held-out (12 reviews): Jev 0.903 (0.962 for final inclusion), bge-base 0.796; hybrid versus ASReview WSS@95 0.557 versus 0.462, difference 0.095 (0.028–0.179), $P=0.034$; WSS@100 0.480 versus 0.336, difference 0.144 (0.056–0.243), $P=0.042$. Supplementary Table 19 gives CIs for the same two comparisons from a separate bootstrap with a different seed (0.0277 to 0.1788 and 0.0551 to 0.2378), so the limits differ in the third decimal.

Controls (12 development reviews, 5550 records; post hoc). Original criteria AUC 0.942; review title only 0.895 (mean within-review $\rho$ with original 0.887); criteria from a different review 0.601 ($\rho$ 0.326). Rephrased questions (named in the plan): 0.943 ($\rho$ 0.959) and 0.943 ($\rho$ 0.977). Supplementary Table 9 gives bootstrap CIs (post hoc, analysis 40): the AUC with the review title only was 0.047 (0.012 to 0.095) and that with mismatched criteria 0.341 (0.279 to 0.405) below the AUC with the original criteria.

**Supplementary Table 9. Rephrasings and negative controls on 12 development reviews (5550 records) with bootstrap CIs (post hoc, analysis 40)**

| Condition | Mean AUC (95% CI) | Difference from original criteria (95% CI) | Reviews lower / higher than original | Mean within-review $\rho$ with original (95% CI) |
| --- | --- | --- | --- | --- |
| Original criteria and question | 0.942 (0.914 to 0.967) | .. | .. | .. |
| Rephrased question 1 | 0.943 (0.917 to 0.967) | 0.001 (−0.003 to 0.006) | 7 / 5 | 0.959 (0.937 to 0.976) |
| Rephrased question 2 | 0.943 (0.916 to 0.967) | 0.001 (−0.001 to 0.004) | 5 / 7 | 0.977 (0.970 to 0.984) |
| Review title only (control) | 0.895 (0.840 to 0.941) | −0.047 (−0.095 to −0.012) | 10 / 2 | 0.887 (0.839 to 0.924) |
| Criteria of a different review (control) | 0.601 (0.544 to 0.663) | −0.341 (−0.405 to −0.279) | 12 / 0 | 0.326 (0.202 to 0.447) |

Percentile bootstrap over the 12 reviews (5000 resamples). The stored macro-averaged AUCs were reproduced. The controls were not run on the held-out reviews or on the pilot review.

CLEF 2019 (31 Cochrane reviews; post hoc). The settings fixed before the held-out evaluation (ten-record batches, $\lambda=3$, and $\tau=0.07$) were applied unchanged. Jev scoring of 82,418 records cost US\$1.54 (\$0.019 per 1000 records).

**Supplementary Table 10. Zero-shot ranking in the CLEF 2019 Cochrane reviews (post hoc)**

| Metric | Jev | bge-base |
| --- | --- | --- |
| AUC, content-level label (28 reviews) | 0.970 | 0.900 |
| Mean average precision, content-level label | 0.598 | 0.225 |
| True negative rate at 95% recall, content-level label | 0.878 | 0.725 |
| Recall at 10% read, content-level label | 0.896 | 0.686 |
| Recall at 20% read, content-level label | 0.950 | 0.822 |
| AUC, title and abstract label (31 reviews) | 0.947 | 0.868 |
| Mean average precision, title and abstract label | 0.541 | 0.302 |
| True negative rate at 95% recall, title and abstract label | 0.806 | 0.633 |
| Recall at 10% read, title and abstract label | 0.784 | 0.589 |
| Recall at 20% read, title and abstract label | 0.893 | 0.731 |

Jev scores are ranked in data-file order for tied scores in this table (the values stored by the original analysis). Values with ties broken at random are in Supplementary Table 18.

Jev versus bge-base AUC (content-level label): 0.070 (0.047–0.095), higher in 26 of 28 reviews, $P<0.0001$. By review type (content-level label): diagnostic accuracy 0.944, intervention 0.980, prognosis 0.970, qualitative 0.992.

Active learning (28 reviews, WSS@95 / WSS@100): ASReview default 0.786 / 0.761; LLM-guided weak supervision 0.810 / 0.815; hybrid 0.830 / 0.849. Hybrid versus ASReview 0.044 (0.013–0.072), higher in 24 of 28, $P=0.0002$; WSS@100 0.088 (0.046–0.129). Hybrid versus weak supervision 0.020 (–0.006 to 0.038); WSS@100 0.033 (0.003–0.059).

Stopping (28 reviews): Jev threshold $\tau=0.07$ reliable in 25 (89.3%; mean recall 0.990, minimum 0.889), 39.1% read; recall below 95% in CD012233 (diagnostic accuracy, 0.889), CD011977 (intervention, 0.919), and CD010558 (intervention, 0.900). Knee method on the ASReview ranking reliable in 26 (92.9%), 76.0% read; on the hybrid ranking in 27 (96.4%), 59.8% read; on the weak-supervision ranking in 27 (96.4%), 81.9% read. On SYNERGY+ held-out reviews, the knee method on the hybrid ranking was reliable in 23 of 23 (83.7% read).

Pilot review. With title and abstract decisions as the label, the AUC was 0.944 for Jev with single-record requests (0.942 with ten-record batches), 0.951 for Claude Opus 5.5, 0.838 for MiniLM, and 0.829 for bge-base. With final inclusion as the label, the AUC was 0.951 for Jev with single-record requests, 0.942 with ten-record batches, and 0.960 for Claude Opus 5.5 (post hoc; all values in this paragraph were recomputed on 9 October 2026 with the corrected labels, Methods; the values for single-record requests and for Claude Opus 5.5 had first been displayed, with the earlier labels, in the evaluations of 24 September, Supplementary Note 1). On the 544-record subset scored by four models, the AUCs were 0.943 (Jev), 0.951 (Claude Opus 5.5), 0.904 (DeepSeek-V3.1), and 0.886 (GPT-4o-mini, verbalized probability). The hybrid gave a WSS@95 of 0.689 versus 0.628 for ASReview and 0.648 for the Jev ranking, and a WSS@100 of 0.418 versus 0.558 and 0.308 (post hoc for the hybrid as fixed on the development reviews; logistic-regression hybrids, the Jev ranking, and a Jev-ordered start had been evaluated on the pilot labels before, Supplementary Note 1; Supplementary Table 18).

Post-hoc comparators. ASReview's strongest preset (elas\_h3) used mxbai-embed-large-v1 sentence embeddings with L2 normalization, a linear support vector machine ($C=0.067$, maximum 5000 iterations), and balancing ratio 9.724; embeddings were computed in 32-bit floating point because the library version loaded the model in 16-bit precision by default, which is slow on central processing units and differs from the precision used when the preset was tuned. In LLM-guided weak supervision (ref. 15), the top T% of records by Jev score were pseudo-labeled included and the bottom B% excluded; the first classifier was trained on the pseudo-labels, and human labels replaced pseudo-labels as records were screened; T and B were chosen on development reviews from $T \in \{1, 2, 5, 10, 15, 20, 30\}$ and $B \in \{50, 80\}$ (best $T=10$, $B=50$; $WSS@95$ 0.734 on the development reviews, against 0.766 for the hybrid with $\lambda=3$). The comparators named in the plan (logistic regression and naive Bayes active learning, retraining after every $N/200$ records, ten seeds) were computed on the held-out reviews as a sensitivity analysis. Post hoc (analysis 44), the MedCPT cross-encoder (ref. 33), a model trained on PubMed search logs and not on screening decisions, was fixed as a further free ranker before any run (ncbi/MedCPT-Cross-Encoder, revision 71caf65d4927987813984f54c284405a13fcca49); it scored each record zero-shot on central processing units with the criteria text of bge-base as the query and the title and abstract as the document, truncated to 512 tokens, and reviews were classed as biomedical or other by the OpenAlex domain of the review publication, a rule fixed before the run. The truncation rule of the tokenizer removes tokens from the end of the longer of the two texts first. Post hoc (analysis 48), 26,919 of 33,001 held-out pairs (81.6%) were truncated, and in 22,719 of them, in 14 of the 23 reviews, the query was shortened (79.7% of query tokens and 75.7% of document tokens kept over all held-out pairs); in the CLEF reviews, 25,790 of 77,611 content-level and 28,502 of 82,418 title and abstract pairs were truncated, mostly in the document, and the query was shortened in 3120 pairs of one review (CD009044).

Implementation checks. In a post-hoc check against the ASReview command-line tool with saved output (analysis 14 of Supplementary Table 1), for two held-out reviews (Theobald 2021, 70 records; Abgaz 2023, 288 records) and seeds 0, 1, and 2, the labeling order up to the last relevant record was identical at 377 of 377 positions, and the prior records were identical in all six cases. When the simulations were re-run on 26 September 2026 with the labeling orders saved (analysis 8), 555 of 561 runs reproduced the stored values exactly (251 of 253 on held-out reviews and 304 of 308 on CLEF reviews); six runs of the default ASReview model differed by one to 26 labeling positions, and the macro-averaged $WSS@95$ and $WSS@100$ changed by less than 0.0001. The hybrid runs and all knee values were identical. We report the stored values.

Controls (analysis 40). The macro-averaged AUCs of the rephrasings and controls and their differences from the original criteria were given percentile bootstrap intervals over the 12 reviews.

#### **Supplementary Note 6. Biomedical cross-encoder as a further free ranker (post hoc)**

The method is described in Supplementary Note 5 (analysis 44). The cross-encoder, which was trained on PubMed search logs, had a lower mean AUC than bge-base in every collection and in the biomedical held-out reviews, so it did not provide a stronger free comparator in this configuration. In this configuration, the 512-token limit shortened the query, which is the criteria text, in most truncated held-out pairs (post hoc, analysis 48; second part of Supplementary Table 11); whether a shorter query would do better was not tested, because choosing one now would be a choice made on the evaluation data. In the Cochrane reviews, in which the query was shortened in one review only, the cross-encoder was also weaker than bge-base (mean AUC 0.783 vs. 0.900; lower in 26 of

28 reviews), so the shortening of the query alone does not explain the difference. Criterion C1, whose comparator bge-base was fixed on development reviews, is not changed.

**Supplementary Table 11. Biomedical cross-encoder (MedCPT) as a further free zero-shot ranker (post hoc, analysis 44)**

| Collection | Mean AUC, MedCPT | Mean AUC, bge-base | Mean AUC, Jev | MedCPT minus bge-base (95% CI); higher / lower; *P* | Jev minus MedCPT (95% CI); higher / lower; *P* | TNR@95, MedCPT / bge-base / Jev |
| --- | --- | --- | --- | --- | --- | --- |
| Held-out (23), final inclusion | 0.709 | 0.831 | 0.966 | -0.122 (-0.192 to -0.060); 6 / 17; 0.0017 | 0.257 (0.189 to 0.332); 23 / 0; <0.0001 | 0.331 / 0.552 / 0.867 |
| CLEF 2019 (28), content-level label | 0.783 | 0.900 | 0.970 | -0.117 (-0.166 to -0.074); 2 / 26; <0.0001 | 0.186 (0.133 to 0.242); 27 / 1; <0.0001 | 0.466 / 0.725 / 0.878 |
| CLEF 2019 (31), title and abstract label | 0.736 | 0.868 | 0.947 | -0.133 (-0.180 to -0.093); 0 / 31; <0.0001 | 0.211 (0.158 to 0.273); 31 / 0; <0.0001 | 0.344 / 0.633 / 0.807 |
| Held-out, biomedical reviews (10) | 0.646 | 0.853 | 0.971 | -0.207 (-0.314 to -0.102); 1 / 9; 0.0059 | 0.325 (0.204 to 0.447); 10 / 0; 0.0020 | .. |
| Held-out, other reviews (13) | 0.757 | 0.813 | 0.962 | -0.056 (-0.122 to 0.000); 5 / 8; 0.168 | 0.205 (0.140 to 0.278); 13 / 0; 0.00024 | .. |

Model ncbi/MedCPT-Cross-Encoder (ref. 33), revision 71caf65d4927, fixed before any run and scored zero-shot on central processing units; query = the criteria text used by bge-base, document = title and abstract, pairs truncated to 512 tokens (26,919 of 33,001 held-out pairs and 28,502 of 82,418 CLEF pairs were truncated). Reviews were classed as biomedical (Health Sciences or Life Sciences) or other by the OpenAlex domain of the review publication, a rule fixed before the run. In the other reviews, the interval of MedCPT minus bge-base included zero. The AUC of MedCPT was below 0.5 in three held-out reviews. No other query form was tried. *P* values are from two-sided Wilcoxon signed-rank tests across reviews and are exploratory; the bootstrap intervals use 5000 resamples of reviews.

Truncation of the query-document pairs (post hoc, analysis 48).

| Collection | Pairs | Truncated to 512 tokens (share) | Only the document shortened | Only the query shortened | Both shortened | Reviews with the query shortened | Query tokens per review, median (range) | Tokens kept over all pairs, query / document |
| --- | --- | --- | --- | --- | --- | --- | --- | --- |
| Held-out (23), final inclusion | 33,001 | 26,919 (81.6%) | 4200 | 5884 | 16,835 | 14 of 23 | 303 (116 to 492) | 79.7% / 75.7% |
| CLEF 2019 (28), content-level label | 77,611 | 25,790 (33.2%) | 22,670 | 1376 | 1744 | 1 of 28 | 136.5 (80 to 462) | 95.5% / 89.1% |
| CLEF 2019 (31), title and abstract label | 82,418 | 28,502 (34.6%) | 25,382 | 1376 | 1744 | 1 of 31 | 139 (80 to 462) | 95.8% / 88.6% |

Tokenizer of the model (same revision; transformers 5.17.0): truncation=True selects the strategy longest\_first, which removes tokens from the end of the longer of the two texts first. Query tokens exclude the two special tokens. In the held-out pairs in which the query was shortened, a median of 103 query tokens (maximum 238) was removed; in the truncated held-out pairs, a median of 80.1% of the document tokens was kept. In the CLEF sets, the query was shortened only in CD009044 (3120 pairs). No query was longer than 509 tokens, so no pair was reduced to the query alone. The numbers of truncated pairs reproduce those of analysis 44 in every review.

Truncation of the MedCPT pairs (analysis 48). The query-document pairs of analysis 44 were rebuilt and encoded with the tokenizer of the model (same revision) without and with truncation to 512 tokens, as in analysis 44; for each pair, the tokens of the query and of the document kept after truncation were counted from the token type identifiers. The truncation strategy was read from the source of the tokenizer library and from the configuration of the model. No model inference was run.

**Free rankers evaluated on the pilot review.** Seven free rankers were evaluated on the pilot review, and four were retained; two cross-encoder rerankers (AUC 0.725 and 0.631) and e5-base-v2 (0.777) performed worse than bge-base and MiniLM and were not run on SYNERGY+.

##### Supplementary Note 7. Included studies below the threshold (post hoc)

The categories and the study-level readings in this note were proposed with the AI coding tool used for the analyses (Methods, Provenance of model inputs and judgments). Neither is used in the main article, which states only that the CLEF content-level label was not checked against the included-study lists of the published reviews and may depart from them in either direction. Supplementary Table 12 lists the included records of development and held-out reviews with a Jev score below 0.07 (12 of 2645), with the proposed categories (A, inclusion departs from the written criteria; B, eligibility information appears only in the full text; C, metadata error; method at the end of this note).

**Supplementary Table 12. Included studies of SYNERGY+ reviews with a Jev score below $\tau=0.07$ (post hoc; proposed categories)**

| Review | Split | Jev score | Title (truncated) | Proposed category |
| --- | --- | --- | --- | --- |
| Ferreira 2022 | dev | 0.03 | Anti-hyperglycemic activity of selenium nanoparticles in streptozotocin-induced diabetic rats | A |
| Ferreira 2022 | dev | 0.04 | Hypoglycemic potential of selenium nanoparticles capped with polyvinyl-pyrrolidone in streptozo | A |
| Maciel 2024 | dev | 0.05 | Turkish Version of the Psychosis Attachment Measure: A Reliability and Validity Analysis | B |
| van der Valk 2021 | dev | 0.06 | Comparison of hair cortisol concentrations between self- and professionally-collected hair samp | B |
| van der Valk 2021 | dev | 0.04 | Hair cortisol concentration as a biomarker of parental burnout | B |
| Fong 2021 | dev | 0.04 | Autonomous and controlled regulation of performance-approach goals: Their relations to perfecti | B |
| Fong 2021 | dev | 0.05 | Study persistence and academic achievement as a function of the type of competing tendencies | B |
| Endedijk 2021 | dev | 0.04 | Safe Learning Environment Perception Scale (SLEPS): A Validity and Reliability Study | B |
| Endedijk 2021 | dev | 0.05 | Immigration status and bullying victimization: Associations across national and school contexts | B |
| Attai 2022 | test | 0.05 | Dengue Fever Outbreak Prediction in Surabaya using A Geographically Weighted Regression | C |
| Attai 2022 | test | 0.06 | Predictive modeling targets thymidylate synthase ThyX in Mycobacterium tuberculosis | A |
| Boersma-van Dam 2024 | test | 0.04 | Hypertrophic Scar Severity at Autograft Sites Is Associated With Increased Pain and Itch After | B |

Supplementary Table 13 lists the content-relevant records below $\tau=0.07$ in the three CLEF 2019 reviews in which the threshold fell below 95% recall (categories D, secondary report of an included study whose other reports were scored above $\tau$, and E, the content-level label does not correspond to an included study of the published review, were added for two records; method at the end of this note).

**Supplementary Table 13. Content-relevant records of CLEF 2019 reviews with a Jev score below $\tau=0.07$ (post hoc; proposed categories and contributions)**

| Review (type; records; content-relevant records) | Recall and proportion read at $\tau$ | Record below $\tau$ (Jev score) | Status in the published review | Proposed category | Contribution to the published review |
| --- | --- | --- | --- | --- | --- |
| CD012233 (diagnostic accuracy; 472; 9) | 0.889; 74.8% | PMID 9362194, Chen 1997 (0.05) | Cited as a background reference; not an included study | E | None; the record appears in no data table |
| CD011977 (intervention; 195; 37) | 0.919; 45.6% | PMID 15682919, Kennis 2004 (0.03) | Included study | B | Primary outcome data presented separately from the pooled estimate, which comprised two other studies |
| CD011977 |  | PMID 19491981, Cui 2009 (0.03) | Included study | B | No data for the primary outcome or for a meta-analysis |
| CD011977 |  | PMID 19781467, Yamaguchi 2009 (0.05) | Included study | B | No data for the primary outcome or for a meta-analysis |
| CD010558 (intervention; 2815; 10) | 0.900; 30.4% | PMID 23851972, secondary report of the included study Wiles 2016 (0.06) | Report of an included study | D | No outcome data; five other reports of the same trial were scored 0.77 to 0.95 |

For category B in CD011977, the abstracts describe comparisons of aspheric with spherical intraocular lenses and do not state that the spherical comparator is a blue-light filtering lens, which is the eligibility information of the review. None of the five records entered a pooled estimate. One of them, Kennis 2004, an included study of CD011977, contributed primary-outcome data that the review presented outside its pooled estimate of two other studies; whether this affected the certainty rating of the primary outcome could not be determined from the text. The published reviews included 16 (CD012233), 51 (CD011977), and six (CD010558) studies, of which four, 37, and four were represented among the content-relevant records. At the level of studies, the threshold retained four of these four studies (CD012233), 34 of 37 (CD011977), and four of four (CD010558); the included studies without a content-relevant record were not assessed. The largest $\tau$ that would have kept recall at 95% or higher was 0.05, 0.03, and 0.06 in the three reviews.

In this reading, the content-level label does not coincide with the included studies of the published review in at least one review (CD012233): of its nine content-relevant records, four are reports of included studies, four are listed by the published review as excluded studies, and one is a background citation. In CD011977 and CD010558, all content-relevant records were reports of included studies. In all three reviews, some included studies of the published review had no content-relevant record (12 of 16, 14 of 51, and two of six), so in this reading the label departs from the published inclusions in both directions in CD012233 and in one direction in the other two; the remaining 28 reviews were not checked.

Included studies below the threshold (analyses P5 and 7). Included records with a Jev score below 0.07 in development and held-out reviews were assigned a proposed category: A, inclusion departs from the written criteria; B, eligibility information appears only in the full text; C, metadata error. For the three Cochrane reviews in which the threshold fell below 95% recall, the content-relevant records below $\tau$ were compared with the reference lists and the text of the published reviews (full texts retrieved from PubMed Central on 26 September 2026), and two categories were added because two records fitted none of A, B, and C: D, secondary report of an included study whose other reports were scored above $\tau$; E, the content-level label does not correspond to an included study of the published review. The classification was proposed by comparing the criteria with the abstracts and texts and is reported only in the Supplementary Information.

##### Supplementary Note 8. Conclusion-level analysis in the pilot review (post hoc)

The aggregated results in this note come from a systematic review by our group that is under review (D. Lee, J. P. Kim, K. Nho, A. J. Saykin, and T. Jo). No record-level data of the review are included.

The quantitative results of the review were recomputed from its extraction sheet. The review has 111 included studies, all of which were scored (lowest Jev score 0.20). One of them was scored on 9 October 2026, because the first matching had assigned it the score of another record; with either score (both above 0.5), the results in Supplementary Table 14 are the same.

###### Supplementary Table 14. Effect of excluding included studies of the pilot review with a Jev score below $\tau$ (post hoc)

| $\tau$ | PubMed records read | Studies missed (of 111) | Findings lost | Domains changing significance | Multi-study genes lost | Genes lost |
| --- | --- | --- | --- | --- | --- | --- |
| 0.07 | 63.2% | 0 | 0 | none | none | 0 |
| 0.10 | 55.7% | 0 | 0 | none | none | 0 |
| 0.20 | 39.5% | 0 | 0 | none | none | 0 |
| 0.30 | 32.2% | 1 | 9 | none | none | 0 |
| 0.50 | 24.2% | 5 | 23 | none | none | 6 |

The proportion of PubMed records read uses the scores from ten-record batches; the 35 included studies from other databases were scored with single-record requests. The domain-level conclusions were unchanged at every $\tau$ examined. At $\tau=0.30$ one study (nine findings) was lost, and at $\tau=0.50$ five studies (23 findings) and six genes with a single study were lost. The statement that the threshold changed no conclusion applies to this review only. The analysis is illustrative: the sign tests are descriptive because findings are nested within studies, the analysis cannot show an effect at $\tau=0.07$, at which no included study was lost, and it becomes informative only from $\tau=0.30$, at which the first study was lost. The effect of missed studies on conclusions was not assessed in any meta-analysis.

Conclusion-level analysis in the pilot review (P7 and analysis 16). The quantitative results of the review were recomputed from its extraction sheet, and the included studies that would have been missed at each $\tau$ were removed. Domain-level sign tests compare detrimental with protective findings (two-sided exact binomial test, Bonferroni correction for six domains); they replicate the method of the review and treat findings as independent, although findings are nested within studies. Gene names were harmonized to symbols before genes were counted. No record-level data of the review are reported. The analysis is illustrative: its sign tests are descriptive, and at $\tau=0.07$, at which no included study was lost, it cannot show an effect.

**Handling of the pilot data.** The zero-shot results of the pilot review are reported as pilot data. The original scoring runs sent the criteria block and the records of the pilot review to four external model services (Jev, GPT-4o-mini, and DeepSeek-V3.1 through the gateway, and Claude Opus 5.5

through its command-line interface), and its submitted manuscript, supplementary criteria file S2, extraction sheet, and records were processed by the AI coding tool and its hosted models (Claude Table 5.1 and Claude Opus 5.5).

#### Supplementary Note 9. Calibration (post hoc)

The measures and the rule recorded before this analysis are described at the end of this note. The rule (pooled recalibration slope between 0.8 and 1.2 and O:E ratio between 0.8 and 1.25 in both the held-out and the CLEF reviews) was met at neither label level.

**Supplementary Table 15. Calibration of Jev scores from ten-record batches: observed and predicted rates (post hoc)**

| Data set | Reviews | Records | Positive records | Observed rate | Mean predicted probability | O:E ratio (95% CI) | Pooled AUC |
| --- | --- | --- | --- | --- | --- | --- | --- |
| Held-out, final inclusion | 23 | 33,001 | 597 | 0.0181 | 0.1404 | 0.129 (0.084 to 0.219) | 0.950 |
| CLEF 2019, content-level label | 28 | 77,611 | 882 | 0.0114 | 0.0910 | 0.125 (0.078 to 0.183) | 0.977 |
| Development, final inclusion | 73 | 52,957 | 2048 | 0.0387 | 0.1929 | 0.200 (0.154 to 0.258) | 0.928 |
| Held-out, title and abstract label | 12 | 20,366 | 948 | 0.0465 | 0.1220 | 0.382 (0.238 to 0.847) | 0.884 |
| CLEF 2019, title and abstract label | 31 | 82,418 | 1682 | 0.0204 | 0.0919 | 0.222 (0.156 to 0.304) | 0.958 |
| Development, title and abstract label | 33 | 20,487 | 2484 | 0.1212 | 0.1648 | 0.736 (0.548 to 0.968) | 0.836 |
| Held-out, final inclusion, the 12 reviews with both labels | 12 | 20,366 | 291 | 0.0143 | 0.1220 | 0.117 (0.069 to 0.264) | 0.960 |

O:E=ratio of the observed rate to the mean predicted probability. CIs are percentile bootstrap intervals over reviews (1000 resamples). Rows without a CI were not bootstrapped.

**Supplementary Table 16. Calibration of Jev scores from ten-record batches: recalibration model and scores (post hoc)**

| Data set | Intercept with slope fixed at 1 | Intercept | Slope (95% CI) | Slope with review effects | Brier score | Scaled Brier score | ECE |
| --- | --- | --- | --- | --- | --- | --- | --- |
| Held-out, final inclusion | -2.78 | -2.85 | 1.30 (1.11 to 1.65) | 1.52 | 0.0475 | -1.68 | 0.122 |
| CLEF 2019, content-level label | -2.91 | -3.06 | 1.31 (1.20 to 1.54) | 1.53 | 0.0289 | -1.57 | 0.080 |
| Development, final inclusion | -2.50 | -2.57 | 1.13 (0.97 to 1.34) | 1.56 | 0.0772 | -1.08 | 0.154 |
| Held-out, title and abstract label | -1.36 | -1.34 | 1.03 (0.94 to 1.25) | 1.19 | 0.0476 | -0.07 | 0.075 |
| CLEF 2019, title and abstract label | -2.17 | -2.15 | 1.15 (1.03 to 1.35) | 1.32 | 0.0297 | -0.49 | 0.071 |
| Development, title and abstract label | -0.52 | -0.67 | 0.84 (0.64 to 1.10) | 1.28 | 0.0885 | 0.17 | 0.044 |
| Held-out, final inclusion, the 12 reviews with both labels | -2.81 | -2.86 | 1.38 (1.18 to 1.93) | 1.74 | 0.0382 | -1.71 | 0.108 |

Intercept and slope are from a logistic recalibration model (logit of the label on the logit of the score). A negative scaled Brier score means that the scores have a larger squared error than the observed rate used as a constant prediction. ECE=expected calibration error (ten bins).

Per review, the O:E ratio for final inclusion lay between 0.8 and 1.25 in none of the 23 held-out reviews (median 0.20, IQR 0.07 to 0.36) and in one of the 28 CLEF reviews (median 0.10; Supplementary Fig. 1). Among reviews with at least five positive records and a converged fit, the slope lay between 0.8 and 1.2 in two of 20 held-out reviews (median 1.82) and in three of 24 CLEF reviews (median 1.67). For title and abstract labels, the O:E ratio lay in the range in one of 12 held-out reviews (median 0.57) and two of 31 CLEF reviews (median 0.30). In the held-out reviews, the tenth of records with the highest scores (scores of 0.38 to 0.97, mean 0.610) had an observed inclusion rate of 0.147 (Wilson 95% CI 0.135 to 0.159).

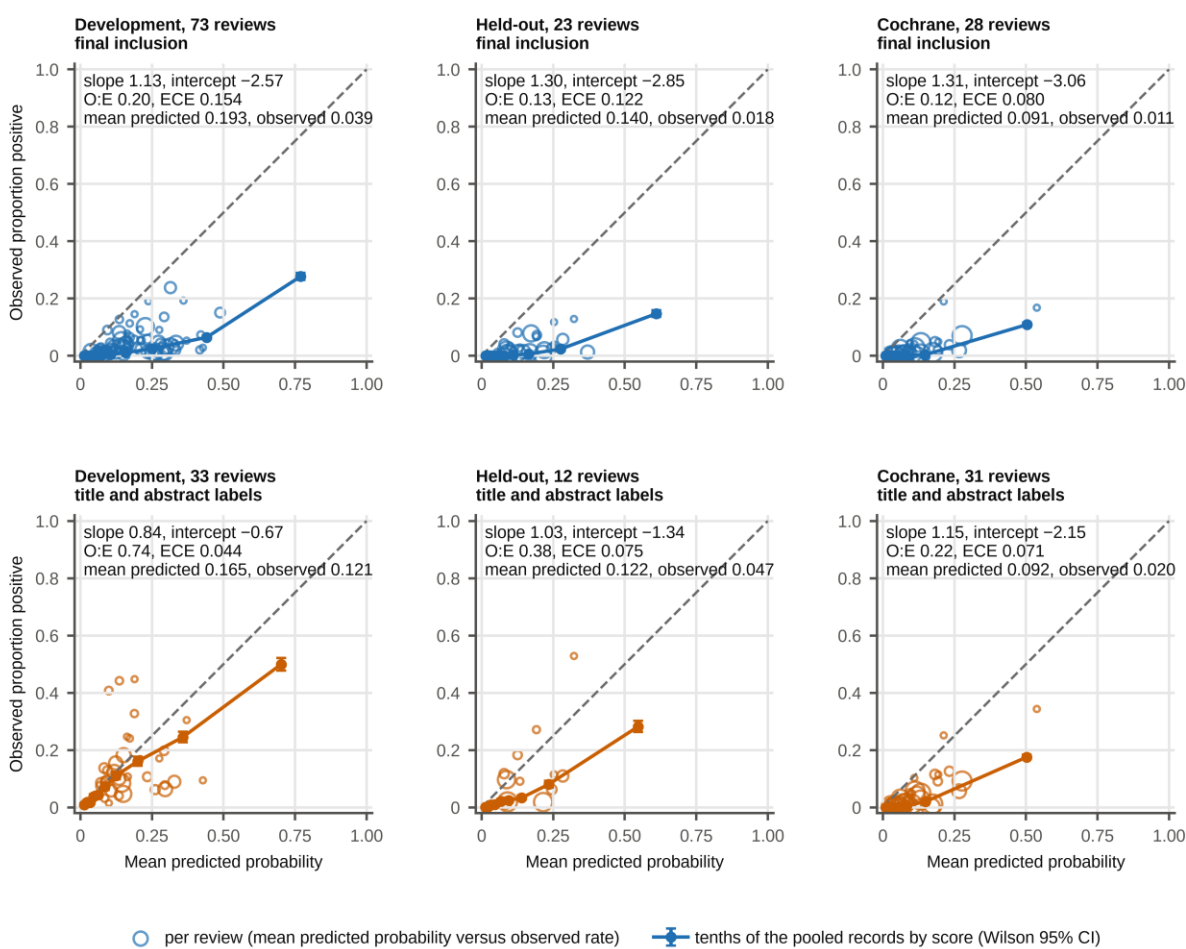

Supplementary Fig. 1 Calibration of Jev scores from ten-record batches (post hoc). Filled circles are tenths of the pooled records with Wilson 95% CIs; open circles are reviews (mean predicted probability against the observed rate; area proportional to the number of records). Panels labeled final inclusion for the Cochrane reviews use the CLEF content-level label.

##### Supplementary Table 17. Calibration by scoring mode in the 2002-record subset and in the pilot review (post hoc)

| Data set and scoring mode | Records | Positive | Observed rate | Mean predicted | O:E ratio (95% CI) | Slope (95% CI) | Brier score | ECE |
| --- | --- | --- | --- | --- | --- | --- | --- | --- |
| 2002-record subset, final inclusion, single-record requests | 2002 | 597 | 0.2982 | 0.2593 | 1.150 (0.917 to 1.437) | 1.47 (1.30 to 1.82) | 0.093 | 0.067 |
| 2002-record subset, final inclusion, batches formed within the subset | 2002 | 597 | 0.2982 | 0.2742 | 1.088 (0.870 to 1.361) | 1.47 (1.29 to 1.85) | 0.087 | 0.062 |
| 2002-record subset, final inclusion, batches of the full run | 2002 | 597 | 0.2982 | 0.2858 | 1.043 (0.839 to 1.294) | 1.48 (1.30 to 1.91) | 0.085 | 0.056 |
| Records of the subset in the 12 reviews with title and abstract labels, title and abstract label, single-record requests | 1160 | 319 | 0.2750 | 0.2188 | 1.257 (0.909 to 1.969) | 1.47 (1.17 to 2.10) | 0.095 | 0.074 |
| Same records, title and abstract label, batches formed within the subset | 1160 | 319 | 0.2750 | 0.2392 | 1.150 (0.854 to 1.686) | 1.55 (1.27 to 2.05) | 0.083 | 0.069 |
| Same records, title and abstract label, batches of the full run | 1160 | 319 | 0.2750 | 0.2503 | 1.099 (0.827 to 1.577) | 1.61 (1.36 to 2.12) | 0.077 | 0.063 |
| Pilot review, title and abstract label, ten-record batches | 1572 | 143 | 0.0910 | 0.2849 | 0.319 | 1.24 | 0.123 | 0.194 |
| Pilot review, final inclusion, ten-record batches | 1572 | 76 | 0.0483 | 0.2849 | 0.170 | 1.39 | 0.147 | 0.237 |
| Pilot review, title and abstract label, single-record requests | 1572 | 143 | 0.0910 | 0.2632 | 0.346 | 1.26 | 0.109 | 0.172 |
| Pilot review, final inclusion, single-record requests | 1572 | 76 | 0.0483 | 0.2632 | 0.184 | 1.45 | 0.130 | 0.215 |

The subset contains all included studies and a sample of excluded records (inclusion rate 29.8%), so its O:E ratio does not describe calibration at the inclusion rate of a full search. CIs are percentile bootstrap intervals over reviews (1000 resamples). Pilot review values are aggregated over its 1572 PubMed records.

Supplementary Table 17 gives the calibration by scoring mode for both label levels; the rows for title and abstract labels use the 1160 records of the subset that belong to the 12 held-out reviews with these labels (319 positive records). In the 2002-record subset, the mean predicted probability was higher with batched scoring than with single-record scoring in 21 of 23 reviews (mean difference 0.040). Per review and for final inclusion, the means of the calibration measures with ten-record batches of the full run and with single-record requests were as follows: mean predicted probability 0.349 with batches and 0.310 with single-record requests (mean difference 0.040; higher with batches in 21 of 23 reviews); O:E ratio 1.06 with batches and 1.25 with single-record requests (mean difference -0.19; higher with batches in two of 23 reviews); expected calibration error 0.159 with batches and 0.178 with single-record requests (mean difference -0.019; higher with batches in ten of 23 reviews); Brier score 0.088 with batches and 0.106 with single-record requests (mean difference -0.017; higher with batches in nine of 23 reviews); recalibration slope 2.17 with batches and 2.20 with single-record requests (mean difference -0.03; higher with batches in nine of 16 reviews); recalibration intercept 0.12 with batches and 0.94 with single-record requests (mean difference -0.82; higher with batches in three of 16 reviews). Slopes and intercepts were compared in the 16 reviews in which the recalibration model converged in both modes.

Calibration (analysis 1). Calibration was assessed with the ratio of the observed rate to the mean predicted probability (O:E), the intercept and slope of a logistic recalibration model (logit of the label on the logit of the score), and calibration plots (ref. 38), and we also report the Brier score and the expected calibration error (the absolute difference between the observed rate and the mean score in ten bins of equal width, averaged with the proportion of records in each bin as the weight). Before this analysis was run, the following rule was recorded in the log (26 September 2026); the mean scores of included and excluded records were already known at that time, so the rule is not a prespecification. The scores would be described as calibrated at a label level only if, at that level, the pooled recalibration slope lay between 0.8 and 1.2 and the O:E ratio between 0.8 and 1.25 in both the held-out and the CLEF reviews.

#### Supplementary Note 10. Label-free Jev ranking, hybrid, and the weight $\lambda$ (post hoc)

**Supplementary Table 18. Work saved and recall by ranking method (the Jev ranking is post hoc)**

| Collection | Method | WSS@95 | WSS@100 | Recall at 10% read | Recall at 20% read | Recall at 30% read |
| --- | --- | --- | --- | --- | --- | --- |
| Held-out, final inclusion (23 reviews) | ASReview default | 0.6587 | 0.6390 | 0.683 | 0.842 | 0.928 |
| Held-out, final inclusion (23 reviews) | ASReview started in Jev order, no blending | 0.6541 | 0.6366 | 0.706 | 0.840 | 0.923 |
| Held-out, final inclusion (23 reviews) | Weak supervision | 0.7460 | 0.7280 | 0.850 | 0.928 | 0.979 |
| Held-out, final inclusion (23 reviews) | Jev ranking, ties at random | 0.7868 | 0.8005 | 0.847 | 0.948 | 0.986 |
| Held-out, final inclusion (23 reviews) | Jev ranking, ties in file order | 0.7882 | 0.8012 | 0.845 | 0.951 | 0.986 |
| Held-out, final inclusion (23 reviews) | Hybrid ($\lambda=3$) | 0.8055 | 0.8212 | 0.863 | 0.963 | 0.987 |
| Held-out, title and abstract label (12 reviews) | ASReview default | 0.4623 | 0.3360 | 0.425 | 0.670 | 0.802 |
| Held-out, title and abstract label (12 reviews) | Jev ranking, ties at random | 0.4978 | 0.4067 | 0.521 | 0.719 | 0.837 |
| Held-out, title and abstract label (12 reviews) | Jev ranking, ties in file order | 0.4942 | 0.4002 | 0.521 | 0.717 | 0.834 |
| Held-out, title and abstract label (12 reviews) | Hybrid ($\lambda=3$) | 0.5575 | 0.4801 | 0.556 | 0.762 | 0.883 |
| CLEF 2019, content-level label (28 reviews) | ASReview default | 0.7863 | 0.7612 | 0.819 | 0.914 | 0.957 |
| CLEF 2019, content-level label (28 reviews) | Weak supervision | 0.8102 | 0.8154 | 0.898 | 0.954 | 0.972 |
| CLEF 2019, content-level label (28 reviews) | Jev ranking, ties at random | 0.8055 | 0.8249 | 0.896 | 0.950 | 0.967 |
| CLEF 2019, content-level label (28 reviews) | Jev ranking, ties in file order | 0.8053 | 0.8248 | 0.896 | 0.950 | 0.967 |
| CLEF 2019, content-level label (28 reviews) | Hybrid ($\lambda=3$) | 0.8304 | 0.8488 | 0.912 | 0.964 | 0.974 |
| CLEF 2019, title and abstract label (31 reviews) | Jev ranking, ties at random | 0.7196 | 0.6384 | 0.780 | 0.894 | 0.931 |
| CLEF 2019, title and abstract label (31 reviews) | Jev ranking, ties in file order | 0.7190 | 0.6395 | 0.781 | 0.893 | 0.930 |
| Pilot review, title and abstract label (one review) | ASReview default | 0.6279 | 0.5581 | 0.520 | 0.790 | 0.934 |
| Pilot review, title and abstract label (one review) | ASReview started in Jev order, no blending | 0.6262 | 0.5585 | 0.552 | 0.783 | 0.930 |
| Pilot review, title and abstract label (one review) | Jev ranking, ties at random | 0.6478 | 0.3084 | 0.631 | 0.895 | 0.944 |
| Pilot review, title and abstract label (one review) | Jev ranking, ties in file order | 0.6472 | 0.3142 | 0.636 | 0.895 | 0.944 |
| Pilot review, title and abstract label (one review) | Hybrid ($\lambda=3$) | 0.6892 | 0.4179 | 0.678 | 0.916 | 0.972 |

Values are macro-averages over reviews. ASReview values are means of ten random starts; the hybrid and weak supervision are single stored runs; the Jev ranking with ties at random is the mean of ten seeds. The rows of the pilot review apply the settings fixed on the development reviews ($\lambda=3$); logistic-regression hybrids with other weights, the Jev ranking, and a Jev-ordered start without blending had been evaluated on the pilot labels on 24 September, before the hybrid was defined (Supplementary Note 1).

**Supplementary Table 19. Paired differences between ranking methods (post hoc)**

| Collection | Comparison | Metric | Mean difference (95% CI) | Reviews higher / lower / tied | P value (Wilcoxon) |
| --- | --- | --- | --- | --- | --- |
| Held-out, final inclusion | Hybrid minus Jev ranking | WSS@95 | 0.0187 (0.0023 to 0.0378) | 16 / 4 / 3 | 0.025 (asymptotic) |
| Held-out, final inclusion | Hybrid minus Jev ranking | WSS@100 | 0.0207 (0.0030 to 0.0402) | 14 / 5 / 4 | 0.033 (asymptotic) |
| Held-out, final inclusion | Hybrid minus Jev ranking | Recall at 10% read | 0.0163 (0.0006 to 0.0336) | 8 / 1 / 14 | 0.051 (asymptotic) |
| Held-out, final inclusion | Hybrid minus Jev ranking | Recall at 20% read | 0.0148 (0.0022 to 0.0294) | 7 / 1 / 15 | 0.036 (asymptotic) |
| Held-out, final inclusion | Hybrid minus Jev ranking | Recall at 30% read | 0.0006 (−0.0119 to 0.0095) | 4 / 1 / 18 | 0.50 (asymptotic) |
| Held-out, final inclusion | Jev ranking minus ASReview | WSS@95 | 0.1281 (0.0616 to 0.1998) | 20 / 3 / 0 | 0.00048 (exact) |
| Held-out, final inclusion | Jev ranking minus ASReview | WSS@100 | 0.1615 (0.0831 to 0.2460) | 20 / 3 / 0 | 0.00025 (exact) |
| Held-out, final inclusion | Jev ranking minus ASReview | Recall at 10% read | 0.1642 (0.0968 to 0.2349) | 18 / 2 / 3 | 0.00039 (asymptotic) |
| Held-out, final inclusion | Jev ranking minus ASReview | Recall at 20% read | 0.1057 (0.0531 to 0.1622) | 16 / 1 / 6 | 0.00085 (asymptotic) |
| Held-out, final inclusion | Jev ranking minus ASReview | Recall at 30% read | 0.0580 (0.0113 to 0.1183) | 9 / 3 / 11 | 0.019 (asymptotic) |
| Held-out, final inclusion | Hybrid minus ASReview | WSS@95 | 0.1468 (0.0892 to 0.2114) | 21 / 2 / 0 | <0.0001 (exact) |
| Held-out, final inclusion | Hybrid minus ASReview | WSS@100 | 0.1822 (0.1158 to 0.2569) | 21 / 2 / 0 | <0.0001 (exact) |
| Held-out, final inclusion | Hybrid minus ASReview | Recall at 10% read | 0.1805 (0.1175 to 0.2467) | 20 / 0 / 3 | <0.0001 (asymptotic) |
| Held-out, final inclusion | Hybrid minus ASReview | Recall at 20% read | 0.1204 (0.0701 to 0.1753) | 17 / 0 / 6 | 0.00029 (asymptotic) |
| Held-out, final inclusion | Hybrid minus ASReview | Recall at 30% read | 0.0585 (0.0167 to 0.1101) | 10 / 1 / 12 | 0.0076 (asymptotic) |
| Held-out, final inclusion | ASReview started in Jev order minus Jev ranking | WSS@95 | −0.1327 (−0.2073 to −0.0640) | 3 / 19 / 1 | 0.0012 (asymptotic) |
| Held-out, final inclusion | ASReview started in Jev order minus Jev ranking | WSS@100 | -0.1639 (-0.2499 to -0.0845) | 3 / 19 / 1 | 0.00061 (asymptotic) |
| Held-out, final inclusion | ASReview started in Jev order minus Jev ranking | Recall at 10% read | -0.1412 (-0.2146 to -0.0710) | 2 / 14 / 7 | 0.0013 (asymptotic) |
| Held-out, final inclusion | ASReview started in Jev order minus Jev ranking | Recall at 20% read | -0.1074 (-0.1692 to -0.0515) | 2 / 15 / 6 | 0.0014 (asymptotic) |
| Held-out, final inclusion | ASReview started in Jev order minus Jev ranking | Recall at 30% read | -0.0635 (-0.1302 to -0.0126) | 3 / 9 / 11 | 0.019 (asymptotic) |
| Held-out, final inclusion | Weak supervision minus Jev ranking | WSS@95 | -0.0408 (-0.1014 to 0.0090) | 7 / 16 / 0 | 0.048 (exact) |
| Held-out, final inclusion | Weak supervision minus Jev ranking | WSS@100 | -0.0725 (-0.1353 to -0.0174) | 6 / 17 / 0 | 0.020 (exact) |
| Held-out, final inclusion | Weak supervision minus Jev ranking | Recall at 10% read | 0.0031 (-0.0012 to 0.0096) | 4 / 1 / 18 | 0.35 (asymptotic) |
| Held-out, final inclusion | Weak supervision minus Jev ranking | Recall at 20% read | -0.0203 (-0.0585 to 0.0093) | 6 / 6 / 11 | 0.39 (asymptotic) |
| Held-out, final inclusion | Weak supervision minus Jev ranking | Recall at 30% read | -0.0078 (-0.0221 to 0.0043) | 4 / 6 / 13 | 0.33 (asymptotic) |
| Held-out, final inclusion | Hybrid minus Jev ranking (ties in file order) | WSS@95 | 0.0173 (0.0016 to 0.0352) | 16 / 4 / 3 | 0.028 (asymptotic) |
| Held-out, final inclusion | Hybrid minus Jev ranking (ties in file order) | WSS@100 | 0.0200 (0.0029 to 0.0385) | 13 / 6 / 4 | 0.036 (asymptotic) |
| Held-out, final inclusion | Hybrid minus Jev ranking (ties in file order) | Recall at 10% read | 0.0186 (0.0016 to 0.0380) | 8 / 1 / 14 | 0.051 (asymptotic) |
| Held-out, final inclusion | Hybrid minus Jev ranking (ties in file order) | Recall at 20% read | 0.0121 (0.0000 to 0.0262) | 5 / 1 / 17 | 0.075 (asymptotic) |
| Held-out, final inclusion | Hybrid minus Jev ranking (ties in file order) | Recall at 30% read | 0.0007 (-0.0119 to 0.0099) | 4 / 1 / 18 | 0.50 (asymptotic) |
| Held-out, title and abstract label | Hybrid minus Jev ranking | WSS@95 | 0.0597 (0.0327 to 0.0873) | 10 / 2 / 0 | 0.0024 (exact) |
| Held-out, title and abstract label | Hybrid minus Jev ranking | WSS@100 | 0.0734 (0.0354 to 0.1140) | 10 / 2 / 0 | 0.0034 (exact) |
| Held-out, title and abstract label | Hybrid minus Jev ranking | Recall at 10% read | 0.0356 (0.0034 to 0.0732) | 7 / 2 / 3 | 0.074 (exact) |
| Held-out, title and abstract label | Hybrid minus Jev ranking | Recall at 20% read | 0.0432 (0.0166 to 0.0721) | 8 / 2 / 2 | 0.014 (exact) |
| Held-out, title and abstract label | Hybrid minus Jev ranking | Recall at 30% read | 0.0458 (0.0225 to 0.0765) | 10 / 0 / 2 | 0.0020 (exact) |
| Held-out, title and abstract label | Jev ranking minus ASReview | WSS@95 | 0.0355 (-0.0440 to 0.1355) | 7 / 5 / 0 | 0.62 (exact) |
| Held-out, title and abstract label | Jev ranking minus ASReview | WSS@100 | 0.0707 (-0.0216 to 0.1793) | 6 / 6 / 0 | 0.38 (exact) |
| Held-out, title and abstract label | Jev ranking minus ASReview | Recall at 10% read | 0.0956 (0.0288 to 0.1629) | 9 / 3 / 0 | 0.034 (exact) |
| Held-out, title and abstract label | Jev ranking minus ASReview | Recall at 20% read | 0.0491 (-0.0109 to 0.1112) | 8 / 4 / 0 | 0.20 (exact) |
| Held-out, title and abstract label | Jev ranking minus ASReview | Recall at 30% read | 0.0348 (-0.0252 to 0.0927) | 7 / 5 / 0 | 0.34 (exact) |
| Held-out, title and abstract label | Hybrid minus ASReview | WSS@95 | 0.0952 (0.0277 to 0.1788) | 9 / 3 / 0 | 0.034 (exact) |
| Held-out, title and abstract label | Hybrid minus ASReview | WSS@100 | 0.1441 (0.0551 to 0.2378) | 8 / 4 / 0 | 0.042 (exact) |
| Held-out, title and abstract label | Hybrid minus ASReview | Recall at 10% read | 0.1313 (0.0703 to 0.1948) | 11 / 1 / 0 | 0.0024 (exact) |
| Held-out, title and abstract label | Hybrid minus ASReview | Recall at 20% read | 0.0923 (0.0451 to 0.1432) | 10 / 2 / 0 | 0.0034 (exact) |
| Held-out, title and abstract label | Hybrid minus ASReview | Recall at 30% read | 0.0806 (0.0362 to 0.1250) | 9 / 2 / 1 | 0.019 (exact) |
| Held-out, title and abstract label | Hybrid minus Jev ranking (ties in file order) | WSS@95 | 0.0632 (0.0380 to 0.0897) | 11 / 1 / 0 | 0.00098 (exact) |
| Held-out, title and abstract label | Hybrid minus Jev ranking (ties in file order) | WSS@100 | 0.0799 (0.0395 to 0.1282) | 11 / 1 / 0 | 0.0024 (exact) |
| Held-out, title and abstract label | Hybrid minus Jev ranking (ties in file order) | Recall at 10% read | 0.0355 (0.0031 to 0.0732) | 6 / 2 / 4 | 0.078 (exact) |
| Held-out, title and abstract label | Hybrid minus Jev ranking (ties in file order) | Recall at 20% read | 0.0447 (0.0175 to 0.0737) | 8 / 2 / 2 | 0.014 (exact) |
| Held-out, title and abstract label | Hybrid minus Jev ranking (ties in file order) | Recall at 30% read | 0.0488 (0.0253 to 0.0800) | 10 / 0 / 2 | 0.0020 (exact) |
| CLEF 2019, content-level label | Hybrid minus Jev ranking | WSS@95 | 0.0249 (0.0071 to 0.0522) | 17 / 9 / 2 | 0.0043 (asymptotic) |
| CLEF 2019, content-level label | Hybrid minus Jev ranking | WSS@100 | 0.0239 (0.0042 to 0.0514) | 15 / 11 / 2 | 0.058 (asymptotic) |
| CLEF 2019, content-level label | Hybrid minus Jev ranking | Recall at 10% read | 0.0167 (0.0067 to 0.0285) | 10 / 1 / 17 | 0.0099 (asymptotic) |
| CLEF 2019, content-level label | Hybrid minus Jev ranking | Recall at 20% read | 0.0137 (0.0027 to 0.0269) | 6 / 1 / 21 | 0.063 (asymptotic) |
| CLEF 2019, content-level label | Hybrid minus Jev ranking | Recall at 30% read | 0.0073 (−0.0023 to 0.0192) | 3 / 2 / 23 | 0.22 (asymptotic) |
| CLEF 2019, content-level label | Jev ranking minus ASReview | WSS@95 | 0.0191 (−0.0227 to 0.0551) | 20 / 8 / 0 | 0.032 (exact) |
| CLEF 2019, content-level label | Jev ranking minus ASReview | WSS@100 | 0.0638 (0.0103 to 0.1129) | 22 / 6 / 0 | 0.0071 (exact) |
| CLEF 2019, content-level label | Jev ranking minus ASReview | Recall at 10% read | 0.0762 (0.0321 to 0.1288) | 17 / 4 / 7 | 0.0026 (asymptotic) |
| CLEF 2019, content-level label | Jev ranking minus ASReview | Recall at 20% read | 0.0365 (0.0028 to 0.0784) | 11 / 4 / 13 | 0.074 (asymptotic) |
| CLEF 2019, content-level label | Jev ranking minus ASReview | Recall at 30% read | 0.0097 (−0.0088 to 0.0307) | 9 / 2 / 17 | 0.15 (asymptotic) |
| CLEF 2019, content-level label | Hybrid minus ASReview | WSS@95 | 0.0440 (0.0142 to 0.0712) | 24 / 4 / 0 | 0.00015 (exact) |
| CLEF 2019, content-level label | Hybrid minus ASReview | WSS@100 | 0.0877 (0.0457 to 0.1285) | 25 / 3 / 0 | <0.0001 (exact) |
| CLEF 2019, content-level label | Hybrid minus ASReview | Recall at 10% read | 0.0929 (0.0513 to 0.1435) | 18 / 2 / 8 | 0.00014 (asymptotic) |
| CLEF 2019, content-level label | Hybrid minus ASReview | Recall at 20% read | 0.0502 (0.0181 to 0.0902) | 12 / 0 / 16 | 0.0022 (asymptotic) |
| CLEF 2019, content-level label | Hybrid minus ASReview | Recall at 30% read | 0.0170 (0.0046 to 0.0341) | 8 / 0 / 20 | 0.012 (asymptotic) |
| CLEF 2019, content-level label | Weak supervision minus Jev ranking | WSS@95 | 0.0047 (−0.0222 to 0.0392) | 9 / 18 / 1 | 0.12 (asymptotic) |
| CLEF 2019, content-level label | Weak supervision minus Jev ranking | WSS@100 | -0.0095 (-0.0427 to 0.0304) | 10 / 17 / 1 | 0.088 (asymptotic) |
| CLEF 2019, content-level label | Weak supervision minus Jev ranking | Recall at 10% read | 0.0029 (-0.0010 to 0.0067) | 5 / 1 / 22 | 0.25 (asymptotic) |
| CLEF 2019, content-level label | Weak supervision minus Jev ranking | Recall at 20% read | 0.0034 (-0.0072 to 0.0166) | 4 / 5 / 19 | 0.86 (asymptotic) |
| CLEF 2019, content-level label | Weak supervision minus Jev ranking | Recall at 30% read | 0.0055 (-0.0066 to 0.0187) | 3 / 4 / 21 | 0.61 (asymptotic) |
| CLEF 2019, content-level label | Hybrid minus Jev ranking (ties in file order) | WSS@95 | 0.0251 (0.0065 to 0.0543) | 17 / 9 / 2 | 0.0074 (asymptotic) |
| CLEF 2019, content-level label | Hybrid minus Jev ranking (ties in file order) | WSS@100 | 0.0241 (0.0033 to 0.0540) | 15 / 11 / 2 | 0.091 (asymptotic) |
| CLEF 2019, content-level label | Hybrid minus Jev ranking (ties in file order) | Recall at 10% read | 0.0158 (0.0058 to 0.0277) | 9 / 1 / 18 | 0.013 (asymptotic) |
| CLEF 2019, content-level label | Hybrid minus Jev ranking (ties in file order) | Recall at 20% read | 0.0135 (0.0026 to 0.0268) | 6 / 1 / 21 | 0.063 (asymptotic) |
| CLEF 2019, content-level label | Hybrid minus Jev ranking (ties in file order) | Recall at 30% read | 0.0073 (-0.0023 to 0.0192) | 3 / 2 / 23 | 0.22 (asymptotic) |

CIs are percentile bootstrap intervals (5000 resamples of reviews, seed 1002). The CI of the comparison of the hybrid with ASReview differs in the third or fourth decimal from the stored value of criterion C3 (0.0888 to 0.2127) because the bootstrap seed differs. The numbers of reviews and the tests were computed from exact differences (Supplementary Note 19). The table gives every comparison that the analysis computed. Exact=exact distribution, by enumeration when zero or tied differences were present among at most 13 reviews; asymptotic=normal approximation.

On the held-out reviews, of the gain of the hybrid over ASReview in WSS@95 (0.147), 0.128 was obtained by the Jev ranking without labels, and the hybrid added 0.019 (95% CI 0.002 to 0.038). The increment was not distinguishable from zero for recall after reading 30% of records. ASReview started in the Jev order without blending gave a WSS@95 of 0.654, which was 0.133 lower than the Jev ranking itself, so this value does not show the contribution of the ranking. On title and abstract labels, the Jev ranking did not differ from ASReview (0.035, -0.044 to 0.135), and the hybrid added 0.060 (0.033 to 0.087).

**Supplementary Table 20. Sensitivity of the hybrid to the weight $\lambda$ ($\lambda=5$ and $\lambda=10$ are post hoc)**

| Collection | Metric | $\lambda=1$ | $\lambda=2$ | $\lambda=3$ (primary) | $\lambda=5$ | $\lambda=10$ | Jev ranking | ASReview |
| --- | --- | --- | --- | --- | --- | --- | --- | --- |
| Development (73 reviews) | WSS@95 | 0.7309 | 0.7657 | 0.7662 | 0.7634 | 0.7538 | 0.7376 | 0.6071 |
| Development (73 reviews) | WSS@100 | 0.7372 | 0.7658 | 0.7657 | 0.7593 | 0.7502 | 0.7325 | 0.5973 |
| Held-out (23 reviews) | WSS@95 | not run | 0.8055 | 0.8055 | 0.8000 | 0.7951 | 0.7868 | 0.6587 |
| Held-out (23 reviews) | WSS@100 | not run | 0.8168 | 0.8212 | 0.8186 | 0.8141 | 0.8005 | 0.6390 |
| CLEF 2019 (28 reviews) | WSS@95 | not run | not run | 0.8304 | 0.8252 | 0.8162 | 0.8055 | 0.7863 |
| CLEF 2019 (28 reviews) | WSS@100 | not run | not run | 0.8488 | 0.8453 | 0.8392 | 0.8249 | 0.7612 |

The selection rule of the plan (largest macro-averaged WSS@95 on development reviews) selects $\lambda=3$ on the extended grid as on the original grid. On the held-out reviews, the WSS@95 was 0.80548

with $\lambda=2$ and 0.80551 with $\lambda=3$ (difference below 0.0001), so both round to 0.8055 at four decimals.

The WSS@95 of the hybrid declined with larger weights toward the value of the Jev ranking. The increment of the hybrid over the Jev ranking remained positive at $\lambda=10$ (held-out 0.008, 95% CI 0.003 to 0.014; CLEF 0.011, 0.003 to 0.024). On held-out reviews the increment was 0.019 (−0.005 to 0.048; $P=0.099$) at $\lambda=2$ and 0.013 (0.004 to 0.022) at $\lambda=5$. Supplementary Table 57 (Supplementary Note 21) gives every paired difference that was computed between weights, with CIs and tests.

Ten tie-breaking seeds (post hoc, analysis 34). The hybrid starts in the Jev order, whose ties were broken at random with one seed in the stored run. Supplementary Table 21 repeats the paired comparisons with the mean over ten seeds; on the held-out reviews the ten-seed values equal the stored run, and the increment of the hybrid over the Jev ranking was 0.0187 (0.0027 to 0.0374).

**Supplementary Table 21. Hybrid with ten tie-breaking seeds of the Jev starting order (post hoc, analysis 34)**

| Collection | Comparison | Mean difference (95% CI) | Higher / lower / tied | *P* |
| --- | --- | --- | --- | --- |
| Held-out (23) | Hybrid (ten-seed mean) minus ASReview, WSS@95 | 0.1468 (0.0899 to 0.2101) | 21 / 2 / 0 | <0.0001 |
| Held-out (23) | Hybrid (ten-seed mean) minus ASReview, WSS@100 | 0.1822 (0.1151 to 0.2555) | 21 / 2 / 0 | <0.0001 |
| Held-out (23) | Hybrid (ten-seed mean) minus Jev ranking, WSS@95 | 0.0187 (0.0027 to 0.0374) | 16 / 4 / 3 | 0.025 |
| Held-out (23) | Hybrid (ten-seed mean) minus Jev ranking, WSS@100 | 0.0207 (0.0027 to 0.0395) | 14 / 5 / 4 | 0.033 |
| Held-out (23) | Hybrid (ten-seed mean) minus Jev ranking, recall at 10% read | 0.0163 (−0.0003 to 0.0335) | 8 / 1 / 14 | 0.051 |
| Held-out (23) | Hybrid (ten-seed mean) minus Jev ranking, recall at 20% read | 0.0148 (0.0020 to 0.0292) | 7 / 1 / 15 | 0.036 |
| Held-out (23) | Jev ranking minus ASReview, WSS@95 | 0.1281 (0.0631 to 0.2002) | 20 / 3 / 0 | 0.00048 |
| CLEF 2019 (28) | Hybrid (ten-seed mean) minus ASReview, WSS@95 | 0.0440 (0.0120 to 0.0723) | 24 / 4 / 0 | 0.00015 |
| CLEF 2019 (28) | Hybrid (ten-seed mean) minus ASReview, WSS@100 | 0.0876 (0.0445 to 0.1281) | 25 / 3 / 0 | <0.0001 |
| CLEF 2019 (28) | Hybrid (ten-seed mean) minus Jev ranking, WSS@95 | 0.0249 (0.0065 to 0.0506) | 18 / 8 / 2 | 0.0034 |
| CLEF 2019 (28) | Hybrid (ten-seed mean) minus Jev ranking, WSS@100 | 0.0239 (0.0032 to 0.0506) | 16 / 10 / 2 | 0.049 |
| CLEF 2019 (28) | Hybrid (ten-seed mean) minus Jev ranking, recall at 10% read | 0.0164 (0.0056 to 0.0279) | 10 / 1 / 17 | 0.016 |
| CLEF 2019 (28) | Hybrid (ten-seed mean) minus Jev ranking, recall at 20% read | 0.0137 (0.0024 to 0.0268) | 6 / 1 / 21 | 0.063 |
| CLEF 2019 (28) | Jev ranking minus ASReview, WSS@95 | 0.0191 (−0.0226 to 0.0566) | 20 / 8 / 0 | 0.032 |

Held-out reviews: the labeling orders differed between seeds in 17 of 23 reviews but the positions at which 95% and 100% recall were reached did not differ in any review, so the macro-averaged

WSS@95 was 0.8055 for every seed, equal to the stored run. CLEF reviews: the positions differed in four reviews, and the macro-averaged WSS@95 ranged from 0.8301 to 0.8305 over seeds (WSS@100 0.8485 to 0.8489). Seed 0 reproduced every stored metric of the hybrid; its labeling order differed from the saved order of the earlier re-run in three reviews (Walker 2018, CD012069, and CD012080) at positions before the last included record, with the same metrics. The C3 comparison (0.05 or more in WSS@95 with $P < 0.05$) was met by each of the ten seeds on the held-out reviews. Criterion C3 is judged on the stored run. The bootstrap interval of the hybrid minus the Jev ranking on held-out reviews differs from that in the main article (0.002 to 0.038) only through the bootstrap seed. $P$ values are exploratory.

True negative rate at 95% recall (post hoc, analysis 35). Supplementary Table 22 gives TNR@95 beside WSS@95 for the rankings. The ordering of the methods was the same as for WSS@95.

**Supplementary Table 22. True negative rate at 95% recall (TNR@95) of the rankings (post hoc, analysis 35)**

| Collection | Ranking or comparison | TNR@95 or mean difference (95% CI) | Higher / lower / tied | $P$ |
| --- | --- | --- | --- | --- |
| Held-out (23) | ASReview default | 0.732 (0.637 to 0.814) | .. | .. |
| Held-out (23) | Jev ranking (ties at random) | 0.867 (0.808 to 0.916) | .. | .. |
| Held-out (23) | Jev ranking (ties in data-file order) | 0.869 (0.811 to 0.917) | .. | .. |
| Held-out (23) | Hybrid (ten-seed mean) | 0.887 (0.843 to 0.927) | .. | .. |
| Held-out (23) | bge-base (ties at random) | 0.552 (0.449 to 0.654) | .. | .. |
| Held-out (23) | Hybrid minus ASReview | 0.155 (0.094 to 0.222) | 21 / 2 / 0 | <0.0001 |
| Held-out (23) | Jev ranking minus ASReview | 0.135 (0.065 to 0.213) | 20 / 3 / 0 | 0.00048 |
| Held-out (23) | Hybrid minus Jev ranking | 0.020 (0.002 to 0.040) | 16 / 4 / 3 | 0.025 |
| Held-out (23) | Jev ranking minus bge-base | 0.315 (0.235 to 0.395) | 23 / 0 / 0 | <0.0001 |
| CLEF 2019 (28) | ASReview default | 0.859 (0.807 to 0.907) | .. | .. |
| CLEF 2019 (28) | Jev ranking (ties at random) | 0.878 (0.801 to 0.941) | .. | .. |
| CLEF 2019 (28) | Jev ranking (ties in data-file order) | 0.878 (0.802 to 0.940) | .. | .. |
| CLEF 2019 (28) | Hybrid (ten-seed mean) | 0.905 (0.834 to 0.958) | .. | .. |
| CLEF 2019 (28) | bge-base (ties at random) | 0.725 (0.635 to 0.813) | .. | .. |
| CLEF 2019 (28) | Hybrid minus ASReview | 0.045 (0.012 to 0.075) | 24 / 4 / 0 | 0.00015 |
| CLEF 2019 (28) | Jev ranking minus ASReview | 0.019 (-0.025 to 0.058) | 20 / 8 / 0 | 0.030 |
| CLEF 2019 (28) | Hybrid minus Jev ranking | 0.027 (0.007 to 0.053) | 18 / 8 / 2 | 0.0034 |
| CLEF 2019 (28) | Jev ranking minus bge-base | 0.153 (0.082 to 0.225) | 26 / 2 / 0 | <0.0001 |

TNR@95 is the proportion of excluded records not yet read when the $[0.95n]$-th of the $n$ included records is found. Because $[0.95n]=n$ for every $n$ of at most 19, 95% recall means all included records in 13 of 23 held-out reviews and 15 of 28 CLEF reviews. The comparisons use the Jev ranking with ties at random; with ties in the order of the data file, its TNR@95 is given in the row above, and bge-base gave the same value with either treatment of ties (0.552 held-out and 0.725 CLEF). The implementation reproduced the stored TNR@95 of the Jev ranking (0.8674 on held-out reviews with random ties; 0.8781 on CLEF reviews in file order) and of bge-base on CLEF reviews in file order (0.7251). Normalized WSS has been shown to be equivalent to the true negative rate, and

TNR at 95% recall has been proposed as a measure (ref. 34); a comparison with its journal pre-proof is reported in Supplementary Note 22 (post hoc, analyses 50 and 51). In the CLEF reviews, the $P$ value of the Jev ranking minus ASReview was not robust to the choice of test (sign-flip permutation test $P=0.387$; post hoc, analysis 49; Supplementary Table 52), as for WSS@95 of the same comparison. $P$ values are exploratory.

**Active learning.** Active learning was simulated with the default model of ASReview (ref. 4) (version 3.0.8; elas\_u4): TF-IDF of title and abstract (word 1–2-grams, sublinear term frequency, maximum document frequency 0.95), a linear support vector machine ($C=0.11$, squared hinge loss), class-balancing sample weights (ratio 9.8), and maximum-relevance querying of one record per iteration, with the classifier retrained after every labeled record. The simulations called the ASReview Simulate class with precomputed features. Priors were one included and one excluded record drawn with seeds 0–9 (ten random starts per review). The reading order starts with the prior records, which therefore count toward the records read, toward WSS, and toward the 150 records of the knee method.

**Hybrid.** Records were ordered by Jev score (ties broken at random) and screened in that order until both classes had been seen; the ranking score for unscreened records was then $z(\text{SVM decision value}) + \lambda \times z(\text{logit of Jev score})$, with $z$-scores computed over the unscreened pool, using the same classifier and retraining schedule. The hybrid thus modifies the ranking score of the default ASReview model. The weight $\lambda=3$ was chosen from $\{1, 2, 3\}$ as the single variant with the highest macro-averaged WSS@95 on development reviews, at the edge of the grid and nearly tied with $\lambda=2$ (WSS@95 0.7662 vs. 0.7657); feature concatenation of the Jev score, also evaluated on the development reviews, gave a lower WSS@95 (0.7488). Post hoc (analysis 34), the hybrid was re-run on the held-out and Cochrane reviews with ten seeds (0 to 9) for breaking ties in the Jev starting order; seed 0 reproduced every stored metric.

#### Supplementary Note 11. Stopping rules: uncertainty, paired differences, strata, and score bands (post hoc)

**Supplementary Table 23. Stopping rules: reliability and workload with 95% CIs (CIs and the statistical criterion are post hoc)**

| Collection (reviews) | Rule | Reviews with recall $\geq 0.95$ (Wilson 95% CI, %) | Exact 95% CI (%) | Records read, macro mean (95% CI) | Records read, median (IQR; range) | Read, pooled | Mean recall | Lowest recall |
| --- | --- | --- | --- | --- | --- | --- | --- | --- |
| Held-out (23) | Threshold $\tau=0.07$ (no labels) | 23 of 23 (85.7 to 100.0%) | 85.2 to 100.0% | 58.9% (49.0 to 69.0) | 53.8% (39.9 to 82.7; 12.9 to 97.1) | 51.0% | 0.998 | 0.958 |
| Held-out (23) | Knee, ASReview ranking | 20 of 23 (67.9 to 95.5%) | 66.4 to 97.2% | 94.0% (88.2 to 98.3) | 100.0% (94.8 to 100.0; 53.6 to 100.0) | 79.3% | 0.977 | 0.657 |
| Held-out (23) | Knee, hybrid ranking | 23 of 23 (85.7 to 100.0%) | 85.2 to 100.0% | 83.7% (75.7 to 91.2) | 88.8% (68.8 to 100.0; 33.1 to 100.0) | 64.6% | 1.000 | 1.000 |
| Held-out (23) | Knee, weak supervision ranking | 23 of 23 (85.7 to 100.0%) | 85.2 to 100.0% | 93.6% (86.2 to 98.6) | 100.0% (94.2 to 100.0; 26.2 to 100.0) | 70.3% | 1.000 | 1.000 |
| Held-out (23) | Knee, strongest preset ranking | 23 of 23 (85.7 to 100.0%) | 85.2 to 100.0% | 96.9% (93.7 to 99.6) | 100.0% (99.7 to 100.0; 74.8 to 100.0) | 90.2% | 1.000 | 0.999 |
| Held-out (23) | Knee, logistic regression active learning (named in the plan) | 21 of 23 (73.2 to 97.6%) | 72.0 to 98.9% | 96.1% (92.2 to 99.1) | 100.0% (98.8 to 100.0; 65.2 to 100.0) | 84.7% | 0.988 | 0.829 |
| Held-out (23) | Knee, naive Bayes active learning (named in the plan) | 21 of 23 (73.2 to 97.6%) | 72.0 to 98.9% | 96.3% (92.6 to 99.1) | 100.0% (98.4 to 100.0; 66.7 to 100.0) | 85.2% | 0.988 | 0.829 |
| Held-out (23) | Knee, Jev ranking | 23 of 23 (85.7 to 100.0%) | 85.2 to 100.0% | 85.4% (77.9 to 92.5) | 100.0% (67.8 to 100.0; 49.4 to 100.0) | 73.0% | 1.000 | 1.000 |
| Held-out (23) | Statistical criterion, ASReview ranking | 23 of 23 (85.7 to 100.0%) | 85.2 to 100.0% | 90.1% (86.3 to 93.6) | 95.3% (84.3 to 96.2; 68.8 to 99.0) | 82.0% | 1.000 | 0.993 |
| Held-out (23) | Statistical criterion, hybrid ranking | 23 of 23 (85.7 to 100.0%) | 85.2 to 100.0% | 87.0% (81.8 to 91.6) | 95.1% (80.9 to 95.5; 56.0 to 97.1) | 75.4% | 1.000 | 1.000 |
| Held-out (23) | Statistical criterion, Jev ranking | 23 of 23 (85.7 to 100.0%) | 85.2 to 100.0% | 87.6% (82.7 to 91.9) | 95.2% (81.2 to 95.5; 58.8 to 97.1) | 76.9% | 1.000 | 1.000 |
| Held-out (23) | Threshold, then statistical criterion, Jev ranking | 23 of 23 (85.7 to 100.0%) | 85.2 to 100.0% | 88.7% (83.9 to 92.7) | 95.3% (83.6 to 95.5; 58.8 to 97.1) | 77.5% | 1.000 | 1.000 |
| CLEF 2019 (28) | Threshold $\tau=0.07$ (no labels) | 25 of 28 (72.8 to 96.3%) | 71.8 to 97.7% | 39.1% (30.7 to 47.6) | 41.4% (20.7 to 54.0; 4.5 to 92.4) | 30.0% | 0.990 | 0.889 |
| CLEF 2019 (28) | Knee, ASReview ranking | 26 of 28 (77.4 to 98.0%) | 76.5 to 99.1% | 76.0% (65.0 to 85.9) | 90.3% (57.4 to 100.0; 4.9 to 100.0) | 61.8% | 0.979 | 0.550 |
| CLEF 2019 (28) | Knee, hybrid ranking | 27 of 28 (82.3 to 99.4%) | 81.7 to 99.9% | 59.8% (46.5 to 72.8) | 63.4% (18.1 to 100.0; 5.4 to 100.0) | 36.0% | 0.998 | 0.943 |
| CLEF 2019 (28) | Knee, weak supervision ranking | 27 of 28 (82.3 to 99.4%) | 81.7 to 99.9% | 81.9% (71.8 to 90.9) | 100.0% (67.5 to 100.0; 9.8 to 100.0) | 76.7% | 0.997 | 0.927 |
| CLEF 2019 (28) | Knee, Jev ranking | 27 of 28 (82.3 to 99.4%) | 81.7 to 99.9% | 64.2% (50.5 to 77.6) | 75.4% (22.5 to 100.0; 6.7 to 100.0) | 36.2% | 0.995 | 0.900 |
| CLEF 2019 (28) | Statistical criterion, ASReview ranking | 28 of 28 (87.9 to 100.0%) | 87.7 to 100.0% | 86.9% (82.3 to 90.9) | 95.0% (82.1 to 95.5; 55.5 to 97.9) | 84.9% | 0.999 | 0.984 |
| CLEF 2019 (28) | Statistical criterion, hybrid ranking | 28 of 28 (87.9 to 100.0%) | 87.7 to 100.0% | 85.7% (80.6 to 90.1) | 95.0% (79.1 to 95.2; 52.7 to 97.9) | 83.4% | 1.000 | 1.000 |
| CLEF 2019 (28) | Statistical criterion, Jev ranking | 28 of 28 (87.9 to 100.0%) | 87.7 to 100.0% | 86.1% (81.1 to 90.5) | 95.0% (79.7 to 95.3; 54.7 to 98.3) | 83.6% | 1.000 | 1.000 |
| CLEF 2019 (28) | Threshold, then statistical criterion, Jev ranking | 28 of 28 (87.9 to 100.0%) | 87.7 to 100.0% | 86.2% (81.3 to 90.7) | 95.0% (79.7 to 95.3; 54.7 to 98.3) | 83.7% | 1.000 | 1.000 |
| Development (73) | Threshold $\tau=0.07$ (no labels) | 72 of 73 (92.6 to 99.8%) | 92.6 to 100.0% | 58.3% (52.9 to 63.6) | 55.6% (42.5 to 80.1; 12.4 to 100.0) | 59.0% | 0.989 | 0.333 |
| Development (73) | Knee, ASReview ranking | 70 of 73 (88.6 to 98.6%) | 88.5 to 99.1% | 94.8% (91.4 to 97.6) | 100.0% (100.0 to 100.0; 30.9 to 100.0) | 90.0% | 0.992 | 0.818 |
| Development (73) | Knee, hybrid ranking | 73 of 73 (95.0 to 100.0%) | 95.1 to 100.0% | 88.5% (83.9 to 92.5) | 100.0% (80.8 to 100.0; 29.3 to 100.0) | 82.1% | 1.000 | 0.995 |

Reliability of rules applied to rankings with ten random starts is judged on the mean recall over starts, as in the original analysis. Exact CIs are Clopper-Pearson intervals. CIs of the macro mean are percentile bootstrap intervals over reviews (5000 resamples; seed 1004 for the threshold and knee rows from the analysis of uncertainty, seed 1008 for the rows from the analysis of the statistical criterion). The statistical criterion is that of Callaghan and Müller-Hansen (recall target 0.95, confidence 0.95).

Macro-averaged and pooled proportions. The macro-average is the mean of the proportions of the single reviews and describes what a team can expect in one review; the pooled proportion is the number of records read in all reviews divided by the number of all records, so that large reviews dominate it, and it describes the labor over a set of reviews. Under the threshold, the pooled proportion read was lower than the macro-average in the held-out reviews (51.0% versus 58.9%) and in the CLEF reviews (30.0% versus 39.1%), and the two were close in the development reviews (59.0% and 58.3%); the threshold therefore left a smaller share of records to read in the larger reviews of the two evaluation collections. The criteria of the plan used the macro-average, which the main article reports; Supplementary Tables 23 and 24 also give the pooled proportions. For planning a single review, we regard the macro-average together with the range over reviews as the relevant quantity (12.9% to 97.1% read in the held-out reviews and 4.5% to 92.4% in the CLEF reviews); for the total labor of a program of reviews, the pooled proportion is the relevant quantity.

When reliability was computed as the mean over random starts of the indicator of recall of at least 95%, the knee method on the ASReview ranking had a reliability of 0.974 in the held-out reviews (six of 230 runs below 95% recall), 0.975 in the CLEF reviews (seven of 280 runs), and 0.989 in the development reviews. In the three held-out reviews in which the mean recall of the knee method was below 95%, some random starts stopped very early. Criterion C4 was met under either definition.

Knee method with the published minimum (post hoc, analysis 33). The knee method as adapted is evaluated from 150 screened records, whereas the published rule is applied after 1000 screened records. Supplementary Table 24 gives both versions. With the published minimum, the knee method on the ASReview ranking was reliable in all held-out and all CLEF reviews; the threshold read fewer records in every held-out review and in 26 of 28 CLEF reviews, but in the CLEF reviews it was less reliable than this comparator.

**Supplementary Table 24. Threshold and knee method with the minimum of 150 screened records (as adapted) and 1000 screened records (published rule) (post hoc, analysis 33)**

| Collection | Rule and ranking | Reviews with recall $\geq 0.95$ (Wilson 95% CI) | Records read, macro (95% CI) | Records read, pooled |
| --- | --- | --- | --- | --- |
| Held-out (23) | Threshold $\tau=0.07$ (no ranking) | 23 of 23 (85.7 to 100.0%) | 58.9% (49.1 to 68.4) | 51.0% |
| Held-out (23) | Knee method as adapted (150), ASReview ranking | 20 of 23 (67.9 to 95.5%) | 94.0% (88.3 to 98.4) | 79.3% |
| Held-out (23) | Knee method, published minimum (1000), ASReview ranking | 23 of 23 (85.7 to 100.0%) | 97.4% (93.4 to 99.9) | 84.8% |
| Held-out (23) | Knee method as adapted (150), hybrid ranking, ten seeds | 23 of 23 (85.7 to 100.0%) | 83.7% (75.8 to 91.0) | 64.6% |
| Held-out (23) | Knee method, published minimum (1000), hybrid ranking, ten seeds | 23 of 23 (85.7 to 100.0%) | 89.8% (82.4 to 96.1) | 66.5% |
| CLEF 2019 (28) | Threshold $\tau=0.07$ (no ranking) | 25 of 28 (72.8 to 96.3%) | 39.1% (30.5 to 47.5) | 30.0% |
| CLEF 2019 (28) | Knee method as adapted (150), ASReview ranking | 26 of 28 (77.4 to 98.0%) | 76.0% (64.7 to 86.2) | 61.8% |

| Collection | Rule and ranking | Reviews with recall<br>≥0.95 (Wilson 95% CI) | Records<br>read, macro<br>(95% CI) | Records<br>read, pooled |
| --- | --- | --- | --- | --- |
| CLEF 2019 (28) | Knee method, published minimum (1000), ASReview ranking | 28 of 28 (87.9 to 100.0%) | 82.2% (72.5 to 91.0) | 70.2% |
| CLEF 2019 (28) | Knee method as adapted (150), hybrid ranking, ten seeds | 27 of 28 (82.3 to 99.4%) | 59.8% (46.2 to 73.0) | 36.0% |
| CLEF 2019 (28) | Knee method, published minimum (1000), hybrid ranking, ten seeds | 27 of 28 (82.3 to 99.4%) | 70.6% (57.6 to 82.7) | 42.0% |

| Collection | Comparison | Reliability<br>difference<br>(Tango 95% CI) | P, asymptotic /<br>exact / mid-p | Difference in<br>records read,<br>macro (95% CI) | Reviews with<br>fewer / more /<br>equal records<br>read | P (records read) |
| --- | --- | --- | --- | --- | --- | --- |
| Held-out | Threshold minus<br>knee as adapted<br>(150), ASReview | 0.130 (−0.031 to<br>0.321) | 0.083 / 0.250 /<br>0.125 | −0.351 (−0.454 to<br>−0.243) | 22 / 1 / 0 | <0.0001 |
| Held-out | Threshold minus<br>knee (1000),<br>ASReview | 0.000 (−0.143 to<br>0.143) | no test / no test /<br>no test | −0.385 (−0.483 to<br>−0.289) | 23 / 0 / 0 | <0.0001 |
| Held-out | Knee (1000)<br>minus knee as<br>adapted (150),<br>ASReview | 0.130 (−0.031 to<br>0.321) | 0.083 / 0.250 /<br>0.125 | 0.034 (0.004 to<br>0.074) | 0 / 5 / 18 | 0.043 |
| CLEF 2019 | Threshold minus<br>knee as adapted<br>(150), ASReview | −0.036 (−0.219 to<br>0.145) | 0.655 / 1.000 /<br>0.688 | −0.369 (−0.469 to<br>−0.269) | 25 / 3 / 0 | <0.0001 |
| CLEF 2019 | Threshold minus<br>knee (1000),<br>ASReview | −0.107 (−0.272 to<br>0.026) | 0.083 / 0.250 /<br>0.125 | −0.432 (−0.532 to<br>−0.329) | 26 / 2 / 0 | <0.0001 |
| CLEF 2019 | Knee (1000)<br>minus knee as<br>adapted (150),<br>ASReview | 0.071 (−0.058 to<br>0.226) | 0.157 / 0.500 /<br>0.250 | 0.063 (0.020 to<br>0.119) | 2 / 12 / 14 | 0.0029 |

| Collection and review<br>size (records) | Threshold<br>τ=0.07: reliable /<br>records read,<br>macro | Knee as adapted<br>(150), ASReview:<br>reliable / records<br>read, macro | Knee, published<br>minimum (1000),<br>ASReview:<br>reliable / records<br>read, macro | Knee as adapted<br>(150), hybrid, ten<br>seeds: reliable /<br>records read,<br>macro | Knee, published<br>minimum (1000),<br>hybrid, ten<br>seeds: reliable /<br>records read,<br>macro |
| --- | --- | --- | --- | --- | --- |
| Held-out, fewer than<br>1000 | 13 of 13 / 58.0% | 13 of 13 / 98.1% | 13 of 13 / 100.0% | 13 of 13 / 89.3% | 13 of 13 / 100.0% |
| Held-out, 1000 or more | 10 of 10 / 60.1% | 7 of 10 / 88.7% | 10 of 10 / 93.9% | 10 of 10 / 76.5% | 10 of 10 / 76.5% |
| Held-out, at most 2000 | 20 of 20 / 59.1% | 19 of 20 / 97.3% | 20 of 20 / 98.9% | 20 of 20 / 87.7% | 20 of 20 / 94.7% |
| Held-out, more than<br>2000 | 3 of 3 / 57.6% | 1 of 3 / 72.1% | 3 of 3 / 86.8% | 3 of 3 / 57.2% | 3 of 3 / 57.2% |
| CLEF 2019, fewer than<br>1000 | 9 of 11 / 47.8% | 11 of 11 / 95.6% | 11 of 11 / 100.0% | 11 of 11 / 85.5% | 11 of 11 / 100.0% |
| CLEF 2019, 1000 or<br>more | 16 of 17 / 33.4% | 15 of 17 / 63.3% | 17 of 17 / 70.7% | 16 of 17 / 43.2% | 16 of 17 / 51.5% |
| CLEF 2019, at most<br>2000 | 13 of 15 / 42.7% | 15 of 15 / 89.5% | 15 of 15 / 94.7% | 15 of 15 / 78.2% | 15 of 15 / 94.3% |
| CLEF 2019, more than<br>2000 | 12 of 13 / 34.8% | 11 of 13 / 60.4% | 13 of 13 / 67.8% | 12 of 13 / 38.7% | 12 of 13 / 43.2% |

| Collection, review size (records), and comparison | Reliability difference (Tango 95% CI) | P, asymptotic / exact / mid-p | Difference in records read, macro (95% CI) | Reviews with fewer / more / equal records read | P (records read) |
| --- | --- | --- | --- | --- | --- |
| Held-out, fewer than 1000, threshold minus knee as adapted (150) | 0.000 (-0.228 to 0.228) | no test / no test / no test | -0.401 (-0.526 to -0.276) | 13 / 0 / 0 | 0.00024 |
| Held-out, fewer than 1000, threshold minus knee (1000) | 0.000 (-0.228 to 0.228) | no test / no test / no test | -0.420 (-0.564 to -0.284) | 13 / 0 / 0 | 0.00024 |
| Held-out, fewer than 1000, knee (1000) minus knee as adapted (150) | 0.000 (-0.228 to 0.228) | no test / no test / no test | 0.019 (0.000 to 0.054) | 0 / 2 / 11 | 0.500 |
| Held-out, 1000 or more, threshold minus knee as adapted (150) | 0.300 (-0.061 to 0.603) | 0.083 / 0.250 / 0.125 | -0.286 (-0.451 to -0.108) | 9 / 1 / 0 | 0.020 |
| Held-out, 1000 or more, threshold minus knee (1000) | 0.000 (-0.278 to 0.278) | no test / no test / no test | -0.338 (-0.467 to -0.206) | 10 / 0 / 0 | 0.0020 |
| Held-out, 1000 or more, knee (1000) minus knee as adapted (150) | 0.300 (-0.061 to 0.603) | 0.083 / 0.250 / 0.125 | 0.053 (0.000 to 0.129) | 0 / 3 / 7 | 0.250 |
| Held-out, at most 2000, threshold minus knee as adapted (150) | 0.050 (-0.119 to 0.236) | 0.317 / 1.000 / 0.500 | -0.382 (-0.481 to -0.286) | 20 / 0 / 0 | <0.0001 |
| Held-out, at most 2000, threshold minus knee (1000) | 0.000 (-0.161 to 0.161) | no test / no test / no test | -0.399 (-0.504 to -0.297) | 20 / 0 / 0 | <0.0001 |
| Held-out, at most 2000, knee (1000) minus knee as adapted (150) | 0.050 (-0.119 to 0.236) | 0.317 / 1.000 / 0.500 | 0.017 (0.000 to 0.042) | 0 / 3 / 17 | 0.109 |
| Held-out, more than 2000, threshold minus knee as adapted (150) | 0.667 (-0.269 to 0.939) | 0.157 / 0.500 / 0.250 | -0.146 (-0.579 to 0.343) | 2 / 1 / 0 | 0.750 |
| Held-out, more than 2000, threshold minus knee (1000) | 0.000 (-0.561 to 0.561) | no test / no test / no test | -0.292 (-0.579 to -0.029) | 3 / 0 / 0 | 0.250 |
| Held-out, more than 2000, knee (1000) minus knee as adapted (150) | 0.667 (-0.269 to 0.939) | 0.157 / 0.500 / 0.250 | 0.146 (0.000 to 0.372) | 0 / 2 / 1 | 0.500 |
| CLEF 2019, fewer than 1000, threshold minus knee as adapted (150) | -0.182 (-0.477 to 0.124) | 0.157 / 0.500 / 0.250 | -0.477 (-0.605 to -0.338) | 11 / 0 / 0 | 0.00098 |
| CLEF 2019, fewer than 1000, threshold minus knee (1000) | -0.182 (-0.477 to 0.124) | 0.157 / 0.500 / 0.250 | -0.522 (-0.651 to -0.383) | 11 / 0 / 0 | 0.00098 |
| CLEF 2019, fewer than 1000, knee (1000) minus knee as adapted (150) | 0.000 (-0.259 to 0.259) | no test / no test / no test | 0.044 (0.000 to 0.100) | 0 / 3 / 8 | 0.250 |
| CLEF 2019, 1000 or more, threshold minus knee as adapted (150) | 0.059 (-0.185 to 0.303) | 0.564 / 1.000 / 0.625 | -0.299 (-0.431 to -0.167) | 14 / 3 / 0 | 0.00084 |
| CLEF 2019, 1000 or more, threshold minus knee (1000) | -0.059 (-0.270 to 0.136) | 0.317 / 1.000 / 0.500 | -0.374 (-0.514 to -0.230) | 15 / 2 / 0 | 0.00021 |
| CLEF 2019, 1000 or more, knee (1000) minus knee as adapted (150) | 0.118 (-0.088 to 0.343) | 0.157 / 0.500 / 0.250 | 0.074 (0.012 to 0.155) | 2 / 9 / 6 | 0.013 |
| CLEF 2019, at most 2000, threshold minus knee as adapted (150) | -0.133 (-0.379 to 0.098) | 0.157 / 0.500 / 0.250 | -0.467 (-0.584 to -0.349) | 15 / 0 / 0 | <0.0001 |
| CLEF 2019, at most 2000, threshold minus knee (1000) | -0.133 (-0.379 to 0.098) | 0.157 / 0.500 / 0.250 | -0.520 (-0.627 to -0.408) | 15 / 0 / 0 | <0.0001 |
| CLEF 2019, at most 2000, knee (1000) minus knee as adapted (150) | 0.000 (-0.204 to 0.204) | no test / no test / no test | 0.053 (0.008 to 0.106) | 0 / 5 / 10 | 0.043 |
| CLEF 2019, more than 2000, threshold minus knee as adapted (150) | 0.077 (-0.229 to 0.374) | 0.564 / 1.000 / 0.625 | -0.256 (-0.403 to -0.106) | 10 / 3 / 0 | 0.0081 |
| CLEF 2019, more than 2000, threshold minus knee (1000) | -0.077 (-0.333 to 0.169) | 0.317 / 1.000 / 0.500 | -0.330 (-0.504 to -0.160) | 11 / 2 / 0 | 0.0034 |
| CLEF 2019, more than 2000, knee (1000) minus knee as adapted (150) | 0.154 (-0.109 to 0.422) | 0.157 / 0.500 / 0.250 | 0.074 (0.002 to 0.176) | 2 / 7 / 4 | 0.039 |

The knee method was evaluated after at least 150 (as adapted) or 1000 (published rule) screened records and then every $\max(1, N/1000)$ records, with the stopping condition unchanged; in reviews with fewer than 1000 records the published rule reads all records (13 held-out and 11 CLEF reviews), so its reliability there is that of reading everything. In the reviews with 1000 or more records, the published rule was reliable in 10 of 10 held-out reviews with 93.9% of records read, against 60.1% for the threshold, and in 17 of 17 CLEF reviews with 70.7% read, against 16 of 17

and 33.4% for the threshold. Reliability was judged on the mean recall over the ten random starts. The bootstrap intervals of this table (seed 20260928) differ slightly from those of Table 2 for the same quantities. The saved labeling orders reproduced the stored knee values in 561 of 561 runs. With the published rule as the comparator, criterion C4 as written would be met on the held-out reviews (equal reliability and fewer records read). In the CLEF reviews, for which C4 was not a criterion, the threshold was less reliable than this comparator. C4 as reported is not changed. The hybrid ranking with seed 0 alone (the stored ranking) gave the same numbers of reliable reviews as the ten-seed mean in every collection and size stratum, and macro proportions read that differed by at most 0.13 percentage points. The published rule on the ASReview ranking stopped before the end of the ranking in 13.5% of the held-out and 40.7% of the CLEF random starts, and its mean reliability per random start was 1.0 in both collections. Under the alternative treatments of zero differences (post hoc, analysis 49; Supplementary Table 52), the conclusion at the 0.05 level of the comparison of the knee method with the published minimum and as adapted in the CLEF reviews changed in both size strata: with at most 2000 records, $P=0.0431$ (zeros discarded), 0.0262 (Pratt), and 0.0625 (sign flip); with more than 2000 records, 0.0391, 0.0547, and 0.0273. $P$ values are exploratory.

**Supplementary Table 25. Paired differences between stopping rules (post hoc)**

| Collection | Comparison (first minus second) | Difference in reliability (95% CI; $P$ value) | Reviews in which only the first / only the second rule was reliable | Difference in proportion read (95% CI) | Reviews in which the first rule read fewer / more / the same number of records | $P$ value for the proportion read (Wilcoxon) |
| --- | --- | --- | --- | --- | --- | --- |
| Held-out | Threshold minus knee on the ASReview ranking | 0.130 (0.000 to 0.304; tests in Supplementary Table 26) | 3 / 0 | -0.351 (-0.455 to -0.240) | 22 / 1 / 0 | <0.0001‡ |
| Held-out | Threshold minus knee on the hybrid ranking | 0.000 (0.000 to 0.000; no review differed) | 0 / 0 | -0.249 (-0.338 to -0.165) | 21 / 2 / 0 | <0.0001‡ |
| Held-out | Statistical criterion on the Jev ranking minus threshold | 0.000 (0.000 to 0.000; no review differed) | 0 / 0 | 0.288 (0.194 to 0.385) | 3 / 20 / 0 | <0.0001 |
| Held-out | Statistical criterion minus knee, ASReview ranking | 0.130 (0.000 to 0.261; $P=0.083$) | 3 / 0 | -0.039 (-0.089 to 0.013) | 19 / 4 / 0 | 0.042 |
| Held-out | Statistical criterion, Jev ranking minus ASReview ranking | 0.000 (0.000 to 0.000; no review differed) | 0 / 0 | -0.025 (-0.048 to -0.006) | 17 / 4 / 2 | 0.0051 |
| Held-out | Threshold then statistical criterion minus statistical criterion alone, Jev ranking | 23 of 23 versus 23 of 23 (no CI computed) | .. | 0.010 (0.000 to 0.030) | 0 / 3 / 20 | 0.11 |
| Held-out | Statistical criterion, hybrid ranking minus ASReview ranking | 23 of 23 versus 23 of 23 (no CI computed) | .. | -0.031 (-0.054 to -0.013) | 19 / 2 / 2 | 0.00054 |
| Held-out | Statistical criterion minus knee, hybrid ranking | 23 of 23 versus 23 of 23 (no CI computed) | .. | 0.032 (-0.048 to 0.114) | 14 / 9 / 0 | 0.56 |
| Held-out | Threshold minus statistical criterion on the ASReview ranking | 0.000 (0.000 to 0.000; no review differed) | 0 / 0 | -0.312 (-0.406 to -0.223) | 20 / 3 / 0 | <0.0001 |
| Held-out | Threshold then statistical criterion on the Jev ranking minus threshold | 0.000 (0.000 to 0.000; no review differed) | 0 / 0 | 0.298 (0.210 to 0.390) | 0 / 21 / 2 | <0.0001 |

| Collection | Comparison (first minus second) | Difference in reliability (95% CI; *P* value) | Reviews in which only the first / only the second rule was reliable | Difference in proportion read (95% CI) | Reviews in which the first rule read fewer / more / the same number of records | *P* value for the proportion read (Wilcoxon) |
| --- | --- | --- | --- | --- | --- | --- |
| CLEF 2019 | Threshold minus knee on the ASReview ranking | −0.036 (−0.179 to 0.107; tests in Supplementary Table 26) | 2 / 3 | −0.369 (−0.467 to −0.268) | 25 / 3 / 0 | <0.0001‡ |
| CLEF 2019 | Threshold minus knee on the hybrid ranking | −0.071 (−0.214 to 0.071; tests in Supplementary Table 26) | 1 / 3 | −0.207 (−0.327 to −0.089) | 20 / 8 / 0 | 0.0032‡ |
| CLEF 2019 | Statistical criterion on the Jev ranking minus threshold | 0.107 (0.000 to 0.214; *P*=0.083) | 3 / 0 | 0.470 (0.362 to 0.573) | 2 / 26 / 0 | <0.0001 |
| CLEF 2019 | Statistical criterion minus knee, ASReview ranking | 0.071 (0.000 to 0.179; *P*=0.16) | 2 / 0 | 0.110 (0.005 to 0.223) | 13 / 15 / 0 | 0.25 |
| CLEF 2019 | Statistical criterion, Jev ranking minus ASReview ranking | 0.000 (0.000 to 0.000; no review differed) | 0 / 0 | −0.009 (−0.024 to 0.006) | 17 / 8 / 3 | 0.14 |
| CLEF 2019 | Threshold then statistical criterion minus statistical criterion alone, Jev ranking | 28 of 28 versus 28 of 28 (no CI computed) | .. | 0.002 (0.000 to 0.004) | 0 / 2 / 26 | 0.18 |
| CLEF 2019 | Statistical criterion, hybrid ranking minus ASReview ranking | 28 of 28 versus 28 of 28 (no CI computed) | .. | −0.013 (−0.027 to 0.001) | 20 / 4 / 4 | 0.0086 |
| CLEF 2019 | Statistical criterion minus knee, hybrid ranking | 28 of 28 versus 27 of 28 (no CI computed) | .. | 0.259 (0.126 to 0.399) | 10 / 18 / 0 | 0.0076 |
| CLEF 2019 | Threshold minus statistical criterion on the ASReview ranking | −0.107 (−0.214 to 0.000; *P*=0.083) | 0 / 3 | −0.479 (−0.583 to −0.369) | 26 / 2 / 0 | <0.0001 |
| CLEF 2019 | Threshold then statistical criterion on the Jev ranking minus threshold | 0.107 (0.000 to 0.214; *P*=0.083) | 3 / 0 | 0.472 (0.364 to 0.574) | 0 / 26 / 2 | <0.0001 |
| Development | Threshold minus knee on the ASReview ranking | 0.027 (−0.027 to 0.082; tests in Supplementary Table 26) | 3 / 1 | −0.365 (−0.428 to −0.301) | 65 / 7 / 1 | <0.0001‡ |

CIs are percentile bootstrap intervals over reviews (5000 resamples). The table gives every comparison of proportions read that was computed; a difference in reliability is given where it was computed, with the *P* value of the Wilcoxon signed-rank test on the paired indicators of reliability where a test was stored; this test equals the asymptotic McNemar test on the discordant reviews (columns of reviews in which only one rule was reliable), and Supplementary Table 26 gives exact conditional and mid-p McNemar tests and score intervals for every comparison of reliability. Percentile bootstrap intervals for differences in reliability are degenerate when few or no reviews are discordant (a limit fixed at zero, or a single point when no review differed); for these differences the score intervals of Supplementary Table 26 (Tango and Newcombe) are preferred, and the tests of all comparisons of reliability are in Supplementary Table 26. The numbers of reviews and the tests were computed from exact differences (Supplementary Note 19). ‡These *P* values were not part of the analysis of uncertainty; they were computed on 27 September 2026, together with the recount in exact arithmetic. The tests are descriptive, and no adjustment for multiplicity was made.

Post hoc (analysis 23 of Supplementary Table 2), Supplementary Table 26 gives McNemar-type tests and score intervals for every paired comparison of reliability; no interval excluded zero.

**Supplementary Table 26. Paired comparisons of reliability between stopping rules: McNemar-type tests and score intervals (post hoc)**

| Collection | Comparison (first minus second) | Reviews with recall $\geq 0.95$, first / second | b / c | Difference | P, asymptotic | P, exact | Mid-p | Tango 95% CI | Newcombe 95% CI |
| --- | --- | --- | --- | --- | --- | --- | --- | --- | --- |
| Held-out | Threshold minus knee on the ASReview ranking | 23 / 20 of 23 | 3 / 0 | 0.130 | 0.083 | 0.250 | 0.125 | -0.031 to 0.321 | -0.036 to 0.321 |
| Held-out | Threshold minus knee on the hybrid ranking | 23 / 23 of 23 | 0 / 0 | 0.000 | no test | no test | no test | -0.143 to 0.143 | -0.143 to 0.143 |
| Held-out | Statistical criterion on the Jev ranking minus threshold | 23 / 23 of 23 | 0 / 0 | 0.000 | no test | no test | no test | -0.143 to 0.143 | -0.143 to 0.143 |
| Held-out | Statistical criterion minus knee, ASReview ranking | 23 / 20 of 23 | 3 / 0 | 0.130 | 0.083 | 0.250 | 0.125 | -0.031 to 0.321 | -0.036 to 0.321 |
| Held-out | Statistical criterion, Jev ranking minus ASReview ranking | 23 / 23 of 23 | 0 / 0 | 0.000 | no test | no test | no test | -0.143 to 0.143 | -0.143 to 0.143 |
| Held-out | Threshold then statistical criterion minus statistical criterion alone, Jev ranking | 23 / 23 of 23 | 0 / 0 | 0.000 | no test | no test | no test | -0.143 to 0.143 | -0.143 to 0.143 |
| Held-out | Statistical criterion, hybrid ranking minus ASReview ranking | 23 / 23 of 23 | 0 / 0 | 0.000 | no test | no test | no test | -0.143 to 0.143 | -0.143 to 0.143 |
| Held-out | Statistical criterion minus knee, hybrid ranking | 23 / 23 of 23 | 0 / 0 | 0.000 | no test | no test | no test | -0.143 to 0.143 | -0.143 to 0.143 |
| Held-out | Threshold minus statistical criterion on the ASReview ranking | 23 / 23 of 23 | 0 / 0 | 0.000 | no test | no test | no test | -0.143 to 0.143 | -0.143 to 0.143 |
| Held-out | Threshold then statistical criterion on the Jev ranking minus threshold | 23 / 23 of 23 | 0 / 0 | 0.000 | no test | no test | no test | -0.143 to 0.143 | -0.143 to 0.143 |
| CLEF 2019 | Threshold minus knee on the ASReview ranking | 25 / 26 of 28 | 2 / 3 | -0.036 | 0.655 | 1.000 | 0.688 | -0.219 to 0.145 | -0.213 to 0.140 |
| CLEF 2019 | Threshold minus knee on the hybrid ranking | 25 / 27 of 28 | 1 / 3 | -0.071 | 0.317 | 0.625 | 0.375 | -0.246 to 0.093 | -0.241 to 0.090 |
| CLEF 2019 | Statistical criterion on the Jev ranking minus threshold | 28 / 25 of 28 | 3 / 0 | 0.107 | 0.083 | 0.250 | 0.125 | -0.026 to 0.272 | -0.032 to 0.272 |
| CLEF 2019 | Statistical criterion minus knee, ASReview ranking | 28 / 26 of 28 | 2 / 0 | 0.071 | 0.157 | 0.500 | 0.250 | -0.058 to 0.226 | -0.060 to 0.226 |
| CLEF 2019 | Statistical criterion, Jev ranking minus ASReview ranking | 28 / 28 of 28 | 0 / 0 | 0.000 | no test | no test | no test | -0.121 to 0.121 | -0.121 to 0.121 |
| CLEF 2019 | Threshold then statistical criterion minus statistical criterion alone, Jev ranking | 28 / 28 of 28 | 0 / 0 | 0.000 | no test | no test | no test | -0.121 to 0.121 | -0.121 to 0.121 |
| CLEF 2019 | Statistical criterion, hybrid ranking minus ASReview ranking | 28 / 28 of 28 | 0 / 0 | 0.000 | no test | no test | no test | -0.121 to 0.121 | -0.121 to 0.121 |
| CLEF 2019 | Statistical criterion minus knee, hybrid ranking | 28 / 27 of 28 | 1 / 0 | 0.036 | 0.317 | 1.000 | 0.500 | -0.089 to 0.177 | -0.088 to 0.177 |
| CLEF 2019 | Threshold minus statistical criterion on the ASReview ranking | 25 / 28 of 28 | 0 / 3 | -0.107 | 0.083 | 0.250 | 0.125 | -0.272 to 0.026 | -0.272 to 0.032 |
| CLEF 2019 | Threshold then statistical criterion on the Jev ranking minus threshold | 28 / 25 of 28 | 3 / 0 | 0.107 | 0.083 | 0.250 | 0.125 | -0.026 to 0.272 | -0.032 to 0.272 |
| Development | Threshold minus knee on the ASReview ranking | 72 / 70 of 73 | 3 / 1 | 0.027 | 0.317 | 0.625 | 0.375 | -0.038 to 0.103 | -0.039 to 0.101 |

b / c=numbers of discordant reviews; b=reviews in which only the first rule reached recall of at least 0.95; c=reviews in which only the second did. The asymptotic McNemar test (without continuity correction) equals the Wilcoxon signed-rank test on the paired indicators of reliability whose *P* values are given in Supplementary Table 25. The exact test (column *P*, exact) is the exact conditional binomial test on the discordant reviews, and the mid-p test subtracts half the probability of the observed count. The Tango interval is the asymptotic score interval for the difference of paired proportions, and the Newcombe interval is method 10 of Newcombe (square-and-add of Wilson intervals with a correlation correction). Formulas follow the R package `contingencytables` 3.1.0; the implementation reproduced the 18 test values of the package. Rows without discordant reviews have no test. No interval excludes zero. The tests are descriptive, and no adjustment for multiplicity was made. Pairs of the evaluated options of Table 3 that are not listed here or in Supplementary Table 24 involve at least one option that was reliable in every review, so their discordant counts follow from the numbers of reliable reviews in Table 3, and their Tango intervals equal those of listed rows with the same counts, with the sign reversed when the order of the pair is reversed: without discordant reviews, -0.143 to 0.143 (held-out) and -0.121 to 0.121 (CLEF 2019); for an option reliable in every review minus the knee method as adapted, -0.031 to 0.321 (held-out, three discordant reviews) and -0.058 to 0.226 (CLEF 2019, two); and for an option reliable in every review minus the threshold, -0.026 to 0.272 (CLEF 2019, three). The only pair of evaluated options that were both below full reliability, the threshold and the knee method as adapted in the CLEF 2019 reviews (25 and 26 of 28 reviews), is listed above, with 2 and 3 discordant reviews (-0.219 to 0.145).

**Supplementary Table 27. Threshold rule and knee method by review size, inclusion rate, and review type (post hoc)**

| Collection | Stratum | Reviews | Records | Inclusion rate, range | T: reviews with recall $\geq 0.95$ (Wilson 95% CI, %) | T: read, macro | T: read, pooled | T: lowest recall | K: reviews with recall $\geq 0.95$ (Wilson 95% CI, %) | K: read, macro |
| --- | --- | --- | --- | --- | --- | --- | --- | --- | --- | --- |
| Held-out | At most 2000 records | 20 | 16,792 | 0.34 to 12.86% | 20 of 20 (83.9 to 100.0%) | 59.1% | 58.3% | 0.958 | 19 of 20 (76.4 to 99.1%) | 97.3% |
| Held-out | More than 2000 records | 3 | 16,209 | 0.27 to 0.55% | 3 of 3 (43.9 to 100.0%) | 57.6% | 43.5% | 1.000 | 1 of 3 (6.1 to 79.2%) | 72.1% |
| Held-out | Inclusion rate, highest third | 8 | 4627 | 4.15 to 12.86% | 8 of 8 (67.6 to 100.0%) | 62.2% | 58.1% | 0.958 | 8 of 8 (67.6 to 100.0%) | 99.1% |
| Held-out | Inclusion rate, lowest third | 8 | 22,053 | 0.27 to 1.31% | 8 of 8 (67.6 to 100.0%) | 51.8% | 46.5% | 1.000 | 6 of 8 (40.9 to 92.9%) | 86.4% |
| Held-out | Inclusion rate, middle third | 7 | 6321 | 1.47 to 3.82% | 7 of 7 (64.6 to 100.0%) | 63.2% | 61.4% | 1.000 | 6 of 7 (48.7 to 97.4%) | 96.8% |
| CLEF 2019 | At most 2000 records | 15 | 10,483 | 0.28 to 18.97% | 13 of 15 (62.1 to 96.3%) | 42.7% | 34.3% | 0.889 | 15 of 15 (79.6 to 100.0%) | 89.5% |
| CLEF 2019 | More than 2000 records | 13 | 67,128 | 0.03 to 7.04% | 12 of 13 (66.7 to 98.6%) | 34.8% | 29.4% | 0.900 | 11 of 13 (57.8 to 95.7%) | 60.4% |
| CLEF 2019 | Inclusion rate, highest third | 10 | 11,147 | 2.79 to 18.97% | 9 of 10 (59.6 to 98.2%) | 51.8% | 55.2% | 0.919 | 10 of 10 (72.2 to 100.0%) | 83.9% |
| CLEF 2019 | Inclusion rate, lowest third | 9 | 47,699 | 0.03 to 0.36% | 8 of 9 (56.5 to 98.0%) | 19.2% | 17.3% | 0.900 | 7 of 9 (45.3 to 93.7%) | 55.1% |
| CLEF 2019 | Inclusion rate, middle third | 9 | 18,765 | 0.64 to 2.60% | 8 of 9 (56.5 to 98.0%) | 44.8% | 47.5% | 0.889 | 9 of 9 (70.1 to 100.0%) | 87.9% |
| Development | At most 2000 records | 73 | 52,957 | 0.50 to 23.80% | 72 of 73 (92.6 to 99.8%) | 58.3% | 59.0% | 0.333 | 70 of 73 (88.6 to 98.6%) | 94.8% |
| Development | Inclusion rate, highest third | 25 | 13,273 | 4.53 to 23.80% | 25 of 25 (86.7 to 100.0%) | 65.7% | 65.2% | 0.955 | 25 of 25 (86.7 to 100.0%) | 94.7% |
| Development | Inclusion rate, lowest third | 25 | 21,162 | 0.50 to 2.06% | 24 of 25 (80.5 to 99.3%) | 51.6% | 54.2% | 0.333 | 22 of 25 (70.0 to 95.8%) | 94.7% |
| Development | Inclusion rate, middle third | 23 | 18,522 | 2.14 to 4.42% | 23 of 23 (85.7 to 100.0%) | 57.5% | 60.0% | 1.000 | 23 of 23 (85.7 to 100.0%) | 95.0% |
| CLEF 2019 | Review type: Diagnostic test accuracy | 8 | .. | .. | 7 of 8 (52.9 to 97.8%) | 46.6% | .. | .. | 7 of 8 (52.9 to 97.8%) | 65.7% |
| CLEF 2019 | Review type: Intervention | 18 | .. | .. | 16 of 18 (67.2 to 96.9%) | 35.0% | .. | .. | 17 of 18 (74.2 to 99.0%) | 80.7% |
| CLEF 2019 | Review type: Prognosis | 1 | .. | .. | 1 of 1 (20.7 to 100.0%) | 42.8% | .. | .. | 1 of 1 (20.7 to 100.0%) | 71.8% |
| CLEF 2019 | Review type: Qualitative | 1 | .. | .. | 1 of 1 (20.7 to 100.0%) | 47.7% | .. | .. | 1 of 1 (20.7 to 100.0%) | 77.8% |
| Held-out | Fewer than 150 records | 2 | .. | .. | 2 of 2 (34.2 to 100.0%) | 73.6% | 73.0% | .. | 2 of 2 (34.2 to 100.0%) | 100.0% |
| Held-out | At least 150 records | 21 | .. | .. | 21 of 21 (84.5 to 100.0%) | 57.5% | 50.9% | .. | 18 of 21 (65.4 to 95.0%) | 93.4% |
| Held-out | Fewer than 500 records | 9 | .. | .. | 9 of 9 (70.1 to 100.0%) | 57.9% | 56.1% | .. | 9 of 9 (70.1 to 100.0%) | 97.2% |
| Held-out | At least 500 records | 14 | .. | .. | 14 of 14 (78.5 to 100.0%) | 59.5% | 50.5% | .. | 11 of 14 (52.4 to 92.4%) | 91.9% |
| CLEF 2019 | Fewer than 150 records | 3 | .. | .. | 3 of 3 (43.9 to 100.0%) | 57.9% | 55.3% | .. | 3 of 3 (43.9 to 100.0%) | 100.0% |
| CLEF 2019 | At least 150 records | 25 | .. | .. | 22 of 25 (70.0 to 95.8%) | 36.8% | 29.9% | .. | 23 of 25 (75.0 to 97.8%) | 73.1% |
| CLEF 2019 | Fewer than 500 records | 8 | .. | .. | 6 of 8 (40.9 to 92.9%) | 45.9% | 45.3% | .. | 8 of 8 (67.6 to 100.0%) | 97.2% |
| CLEF 2019 | At least 500 records | 20 | .. | .. | 19 of 20 (76.4 to 99.1%) | 36.3% | 29.6% | .. | 18 of 20 (69.9 to 97.2%) | 67.5% |
| Development | Fewer than 150 records | 7 | .. | .. | 7 of 7 (64.6 to 100.0%) | 60.1% | 60.7% | .. | 7 of 7 (64.6 to 100.0%) | 100.0% |
| Development | At least 150 records | 66 | .. | .. | 65 of 66 (91.9 to 99.7%) | 58.1% | 59.0% | .. | 63 of 66 (87.5 to 98.4%) | 94.2% |

T=threshold rule ($\tau=0.07$). K=knee method on the ASReview ranking. Thirds of the inclusion rate were formed within each collection. The knee method is first applied after 150 screened records, so it reads every record in reviews with fewer than 150 records.

Supplementary Table 28 gives the knee method on the hybrid ranking in the same strata of review size (post hoc; analysis 23 of Supplementary Table 2). In the reviews with at least 150 records it was reliable in 21 of 21 held-out, 24 of 25 CLEF, and 66 of 66 development reviews.

###### Supplementary Table 28. Knee method on the hybrid ranking by review size (post hoc)

| Collection | Stratum | Reviews | Reviews with recall $\geq 0.95$ (Wilson 95% CI, %) | Read, macro | Read, pooled | Lowest recall |
| --- | --- | --- | --- | --- | --- | --- |
| Held-out | Fewer than 150 records | 2 | 2 of 2 (34.2 to 100.0%) | 100.0% | 100.0% | 1.000 |
| Held-out | At least 150 records | 21 | 21 of 21 (84.5 to 100.0%) | 82.2% | 64.4% | 1.000 |
| Held-out | Fewer than 500 records | 9 | 9 of 9 (70.1 to 100.0%) | 88.5% | 88.2% | 1.000 |
| Held-out | At least 500 records | 14 | 14 of 14 (78.5 to 100.0%) | 80.7% | 62.5% | 1.000 |

| Collection | Stratum | Reviews | Reviews with recall $\geq 0.95$<br>(Wilson 95% CI, %) | Read, macro | Read,<br>pooled | Lowest recall |
| --- | --- | --- | --- | --- | --- | --- |
| CLEF 2019 | Fewer than 150 records | 3 | 3 of 3 (43.9 to 100.0%) | 100.0% | 100.0% | 1.000 |
| CLEF 2019 | At least 150 records | 25 | 24 of 25 (80.5 to 99.3%) | 55.0% | 35.7% | 0.943 |
| CLEF 2019 | Fewer than 500 records | 8 | 8 of 8 (67.6 to 100.0%) | 91.8% | 90.5% | 1.000 |
| CLEF 2019 | At least 500 records | 20 | 19 of 20 (76.4 to 99.1%) | 47.0% | 34.7% | 0.943 |
| Development | Fewer than 150 records | 7 | 7 of 7 (64.6 to 100.0%) | 100.0% | 100.0% | 1.000 |
| Development | At least 150 records | 66 | 66 of 66 (94.5 to 100.0%) | 87.3% | 81.8% | 0.995 |
| Development | Fewer than 500 records | 35 | 35 of 35 (90.1 to 100.0%) | 95.6% | 93.9% | 1.000 |
| Development | At least 500 records | 38 | 38 of 38 (90.8 to 100.0%) | 82.0% | 79.4% | 0.995 |

Knee method as in Supplementary Table 27 (first applied after 150 screened records, so every record is read in reviews with fewer than 150 records), applied to the hybrid ranking. For the held-out and CLEF 2019 reviews the numbers of reliable reviews and the proportions read equal the stored values of the analysis of uncertainty; the values for the development reviews were computed in this analysis. The threshold and the knee method on the ASReview ranking in the same strata are in Supplementary Table 27.

**Supplementary Table 29. Records and included studies with Jev scores just below $\tau=0.07$ (post hoc)**

| Collection | Score band | Records in band | All records | Records in band, pooled | Records in band, macro | Included studies in band | All included studies | Included studies in band, pooled | Reviews with an included study in band |
| --- | --- | --- | --- | --- | --- | --- | --- | --- | --- |
| Held-out | 0.03 to $<0.07$ | 9722 | 33,001 | 29.5% | 25.5% | 3 | 597 | 0.50% | 2 |
| Held-out | 0.01 to $<0.07$ | 16,170 | 33,001 | 49.0% | 41.1% | 3 | 597 | 0.50% | 2 |
| CLEF 2019 | 0.03 to $<0.07$ | 21,737 | 77,611 | 28.0% | 29.6% | 5 | 882 | 0.57% | 3 |
| CLEF 2019 | 0.01 to $<0.07$ | 54,252 | 77,611 | 69.9% | 60.9% | 5 | 882 | 0.57% | 3 |
| Development | 0.03 to $<0.07$ | 13,611 | 52,957 | 25.7% | 25.7% | 9 | 2048 | 0.44% | 5 |
| Development | 0.01 to $<0.07$ | 21,661 | 52,957 | 40.9% | 41.6% | 9 | 2048 | 0.44% | 5 |
| Held-out and CLEF 2019 combined | 0.03 to $<0.07$ | 31,459 | 110,612 | 28.4% | 27.8% | 8 | 1479 | 0.54% | 5 |
| Held-out and CLEF 2019 combined | 0.01 to $<0.07$ | 70,422 | 110,612 | 63.7% | 52.0% | 8 | 1479 | 0.54% | 5 |

All included studies below $\tau$ had scores of at least 0.03, so the counts of included studies are the same in the two bands.

Failures of the threshold pooled over the evaluation collections (post hoc, analysis 32).

Supplementary Table 30 gives the reviews in which the threshold fell below 95% recall.

**Supplementary Table 30. Reviews in which the threshold fell below 95% recall in the**

#### evaluation collections (post hoc, analysis 32)

| Collection | Reviews below 95% recall | Proportion (Wilson 95% CI) | Exact 95% CI | One-sided exact upper 95% limit |
| --- | --- | --- | --- | --- |
| Held-out SYNERGY+ (final inclusion) and CLEF 2019 (content-level label), pooled | 3 of 51 | 5.9% (2.0 to 15.9%) | 1.2 to 16.2% | 14.5% |

| Failing review | Records | Content-relevant records | Missed below $\tau$ | Recall | Records read |
| --- | --- | --- | --- | --- | --- |
| CD012233 | 472 | 9 | 1 | 0.889 | 74.8% |
| CD011977 | 195 | 37 | 3 | 0.919 | 45.6% |
| CD010558 | 2815 | 10 | 1 | 0.900 | 30.4% |

No held-out review fell below 95% recall. The threshold gives no signal within a review that it has fallen below the target.

Proportion read in a new review (post hoc, analysis 43). Supplementary Table 31 gives random-effects estimates and 95% prediction intervals for the proportion of records read. The intervals are wide, so the proportion read in a single new review cannot be predicted closely from these collections.

##### Supplementary Table 31. Proportion of records read in a new review: random-effects estimates and 95% prediction intervals, and recall per review (post hoc, analysis 43)

| Collection | Rule | Pooled estimate (95% CI) | 95% prediction interval | Empirical 5th to 95th percentile | Range over reviews |
| --- | --- | --- | --- | --- | --- |
| Held-out (23) | Threshold $\tau=0.07$ | 62.8% (53.3 to 71.4) | 18.4 to 92.7% | 21.8 to 92.6% | 12.9 to 97.1% |
| Held-out (23) | Jev ranking plus statistical criterion | 90.9% (86.6 to 93.9) | 52.9 to 98.9% | 68.2 to 96.9% | 58.8 to 97.1% |
| CLEF 2019 (28) | Threshold $\tau=0.07$ | 36.1% (27.9 to 45.1) | 6.4 to 82.2% | 8.4 to 76.7% | 4.5 to 92.4% |
| CLEF 2019 (28) | Jev ranking plus statistical criterion | 90.0% (85.9 to 93.0) | 50.7 to 98.7% | 57.3 to 96.7% | 54.7 to 98.3% |

| Collection | Rule | Recall per review: mean | Empirical 5th percentile | Lowest | Reviews with recall 1 |
| --- | --- | --- | --- | --- | --- |
| Held-out (23) | Threshold $\tau=0.07$ | 0.998 | 0.987 | 0.958 | 21 of 23 |
| Held-out (23) | Jev ranking plus statistical criterion | 1.000 | 1.000 | 1.000 | 23 of 23 |
| CLEF 2019 (28) | Threshold $\tau=0.07$ | 0.990 | 0.907 | 0.889 | 25 of 28 |
| CLEF 2019 (28) | Jev ranking plus statistical criterion | 1.000 | 1.000 | 1.000 | 28 of 28 |

Random-effects meta-analysis of the logit of the proportion read per review (DerSimonian-Laird), with the prediction interval of Higgins, Thompson, and Spiegelhalter (t distribution with $k-2$ degrees of freedom; ref. 39), back-transformed. The pooled estimate weights reviews by the inverse of their variance and therefore differs from the macro-average. Between-review heterogeneity was large ($I^2$

above 0.99 in every row). At a fixed $\tau$, the proportion read in a review is determined by the stored scores, so the binomial within-review variance has no sampling interpretation; with $I^2$ above 0.99 it has no practical effect, and the prediction interval reflects the variation between reviews, which the empirical 5th and 95th percentiles describe directly. The random-effects center differs from the macro-average because of the logit scale and the weighting. Recall was summarized by its empirical distribution over reviews only; most reviews had a recall of 1, so no prediction interval was computed for it.

Derived quantities (analysis 32). From the stored scores and labels, the following were derived: the number of evaluation reviews in which the threshold fell below 95% recall, pooled over the held-out and Cochrane reviews, with Wilson and exact (Clopper-Pearson) two-sided intervals and the one-sided exact upper limit; the records missed in each failing review; the cross-tabulation of positive records below and at or above $\tau$ at the two label levels; the cost of scoring each collection and the numbers of records that correspond to the reported differences in the proportion read; and the resolution of the stored scores (Supplementary Note 1).

Prediction intervals (analysis 43). The logit of the proportion of records read per review was pooled in a random-effects meta-analysis (within-review variance from the binomial approximation, DerSimonian-Laird between-review variance), with a 95% prediction interval based on the $t$ distribution with $k-2$ degrees of freedom (ref. 39), back-transformed; the empirical 5th and 95th percentiles over reviews are also given. At a fixed $\tau$, the proportion read in a review is determined by the stored scores, so the binomial within-review variance has no sampling interpretation; with $I^2$ above 0.99 it has no practical effect, and the interval reflects the variation between reviews. The random-effects center differs from the macro-average because of the logit scale and the weighting.

Knee method. The comparator was a knee method on the ASReview ranking, adapted from Cormack and Grossman (ref. 6) (the knee method as adapted). The implementation evaluates the rule after at least 150 screened records and then every $\max(1, N/1000)$ records (integer division): it finds the knee of the gain curve, computes the ratio $\rho$ of the slope before the knee to the slope after the knee, and stops when $\rho$ is at least 156 minus the number of included records found (capped at 150). The full text of the original publication could not be retrieved on the analysis server (HTTP 403), so the parameters were checked in the description by Yang and colleagues (ref. 26) (arXiv version, sections 3.5 and 6; analysis 25 of Supplementary Table 2): the published rule stops at the first point $s$ at which $\rho(s)$ is at least 156 minus $\min(\text{relevant records found}, 150)$ and at least 1000 records have been screened, and "The Knee Method is targeted at a recall goal of 0.7". The implementation of this study applies the rule after 150 screened records and was judged against a recall target of 0.95, so the comparator is an adaptation of a rule designed for a lower recall goal; its evaluation interval was not checked against the original description. In reviews with fewer than 150 records the rule reads all records. The rule was applied to each random start, and the reliability of a review was judged on the mean recall over the ten starts; post hoc (analysis 52), the proportion of single runs that reached 95% recall is also reported for every evaluated stopping option (Supplementary Table 41, cited in the Table 3 footnote). Post hoc (analysis 33), the rule was also applied with the published minimum of 1000 screened records to the saved labeling orders of the ASReview ranking (ten random starts) and of the hybrid ranking (ten tie-breaking seeds), with the evaluation interval and the stopping condition unchanged; in reviews with fewer than 1000 records it then reads all records, and the saved orders reproduced the stored values of the rule as adapted in 561 of 561 runs. An evaluation of 15 stopping methods (full text read in PubMed Central) reported that almost all either failed to stop reliably or left work savings unused, that "BATCHPRECISION and KNEE have no clear point where they stop, far too early or late, and are spread across the entire range, independent of the hyperparameter settings", and that "Only one method, CMH, never stops before the set recall target" (ref. 11); CMH is a statistical stopping criterion of the kind evaluated post hoc here.

#### Supplementary Note 12. Sensitivity to $\tau$ (post hoc)

On the grid searched by the original analysis (Methods, Stopping rules), the reliability on the development reviews was 1.000 at 0.01 to 0.03, 0.986 at 0.05 and 0.07, 0.918 at 0.10, 0.808 at 0.15, 0.740 at 0.20, and 0.562 at 0.30 (Supplementary Fig. 2). On a grid in steps of 0.01 the same rule selects 0.09 (reliability 0.973 at 0.08 and 0.959 at 0.09). One development review lost 95% recall between 0.07 and 0.08.

**Supplementary Table 32. Reliability, workload, and recall of the threshold rule as a function of $\tau$ (post hoc)**

| Data set | $\tau$ | Reviews with recall $\geq 0.95$<br>(Wilson 95% CI, %) | Records<br>read, macro | Records<br>read, pooled | Recall,<br>mean | Recall,<br>minimum |
| --- | --- | --- | --- | --- | --- | --- |
| Development, final inclusion | 0.01 | 73 of 73 (95.0 to 100.0%) | 99.9% | 99.9% | 1.000 | 1.000 |
| Development, final inclusion | 0.02 | 73 of 73 (95.0 to 100.0%) | 93.6% | 93.9% | 1.000 | 1.000 |
| Development, final inclusion | 0.03 | 73 of 73 (95.0 to 100.0%) | 84.0% | 84.7% | 1.000 | 1.000 |
| Development, final inclusion | 0.05 | 72 of 73 (92.6 to 99.8%) | 69.0% | 69.9% | 0.990 | 0.333 |
| Development, final inclusion | 0.07 | 72 of 73 (92.6 to 99.8%) | 58.3% | 59.0% | 0.989 | 0.333 |
| Development, final inclusion | 0.08 | 71 of 73 (90.5 to 99.2%) | 54.2% | 54.8% | 0.989 | 0.333 |
| Development, final inclusion | 0.09 | 70 of 73 (88.6 to 98.6%) | 50.5% | 50.9% | 0.987 | 0.333 |
| Development, final inclusion | 0.10 | 67 of 73 (83.2 to 96.2%) | 47.4% | 47.7% | 0.978 | 0.333 |
| Development, final inclusion | 0.15 | 59 of 73 (70.3 to 88.2%) | 36.5% | 36.7% | 0.963 | 0.333 |
| Development, final inclusion | 0.20 | 54 of 73 (62.9 to 82.7%) | 29.8% | 30.2% | 0.952 | 0.333 |
| Development, final inclusion | 0.30 | 41 of 73 (44.8 to 67.0%) | 21.3% | 21.6% | 0.889 | 0.000 |
| Development, final inclusion | 0.50 | 28 of 73 (28.1 to 49.8%) | 13.1% | 12.9% | 0.795 | 0.000 |
| Held-out, final inclusion | 0.01 | 23 of 23 (85.7 to 100.0%) | 100.0% | 100.0% | 1.000 | 1.000 |
| Held-out, final inclusion | 0.02 | 23 of 23 (85.7 to 100.0%) | 94.9% | 94.9% | 1.000 | 1.000 |
| Held-out, final inclusion | 0.03 | 23 of 23 (85.7 to 100.0%) | 84.4% | 80.5% | 1.000 | 1.000 |
| Held-out, final inclusion | 0.05 | 23 of 23 (85.7 to 100.0%) | 69.0% | 62.1% | 0.998 | 0.958 |
| Held-out, final inclusion | 0.07 | 23 of 23 (85.7 to 100.0%) | 58.9% | 51.0% | 0.998 | 0.958 |
| Held-out, final inclusion | 0.08 | 23 of 23 (85.7 to 100.0%) | 54.4% | 46.6% | 0.998 | 0.958 |
| Held-out, final inclusion | 0.09 | 23 of 23 (85.7 to 100.0%) | 50.4% | 42.8% | 0.998 | 0.958 |
| Held-out, final inclusion | 0.10 | 23 of 23 (85.7 to 100.0%) | 46.9% | 39.2% | 0.996 | 0.958 |
| Held-out, final inclusion | 0.15 | 19 of 23 (62.9 to 93.0%) | 34.6% | 27.5% | 0.985 | 0.875 |
| Held-out, final inclusion | 0.20 | 18 of 23 (58.1 to 90.3%) | 27.2% | 21.0% | 0.974 | 0.846 |
| Held-out, final inclusion | 0.30 | 16 of 23 (49.1 to 84.4%) | 18.2% | 13.5% | 0.942 | 0.667 |
| Held-out, final inclusion | 0.50 | 9 of 23 (22.2 to 59.2%) | 10.1% | 6.9% | 0.834 | 0.154 |
| CLEF 2019, content-level label | 0.01 | 28 of 28 (87.9 to 100.0%) | 100.0% | 99.9% | 1.000 | 1.000 |
| CLEF 2019, content-level label | 0.02 | 28 of 28 (87.9 to 100.0%) | 88.0% | 78.9% | 1.000 | 1.000 |
| CLEF 2019, content-level label | 0.03 | 28 of 28 (87.9 to 100.0%) | 68.7% | 58.0% | 1.000 | 1.000 |
| CLEF 2019, content-level label | 0.05 | 27 of 28 (82.3 to 99.4%) | 49.4% | 39.8% | 0.998 | 0.946 |
| CLEF 2019, content-level label | 0.07 | 25 of 28 (72.8 to 96.3%) | 39.1% | 30.0% | 0.990 | 0.889 |
| CLEF 2019, content-level label | 0.08 | 25 of 28 (72.8 to 96.3%) | 35.6% | 26.7% | 0.989 | 0.889 |
| CLEF 2019, content-level label | 0.09 | 25 of 28 (72.8 to 96.3%) | 32.4% | 23.7% | 0.987 | 0.838 |
| CLEF 2019, content-level label | 0.10 | 25 of 28 (72.8 to 96.3%) | 30.0% | 21.4% | 0.986 | 0.838 |
| CLEF 2019, content-level label | 0.15 | 25 of 28 (72.8 to 96.3%) | 21.9% | 14.5% | 0.981 | 0.784 |
| CLEF 2019, content-level label | 0.20 | 23 of 28 (64.4 to 92.1%) | 17.9% | 11.3% | 0.975 | 0.778 |
| CLEF 2019, content-level label | 0.30 | 20 of 28 (52.9 to 84.7%) | 13.6% | 7.7% | 0.959 | 0.778 |
| CLEF 2019, content-level label | 0.50 | 16 of 28 (39.1 to 73.5%) | 8.7% | 4.4% | 0.880 | 0.000 |
| Held-out, title and abstract label | 0.01 | 12 of 12 (75.8 to 100.0%) | 100.0% | 100.0% | 1.000 | 1.000 |
| Held-out, title and abstract label | 0.02 | 12 of 12 (75.8 to 100.0%) | 92.7% | 93.1% | 1.000 | 1.000 |
| Held-out, title and abstract label | 0.03 | 12 of 12 (75.8 to 100.0%) | 80.1% | 75.0% | 0.999 | 0.985 |
| Held-out, title and abstract label | 0.05 | 10 of 12 (55.2 to 95.3%) | 66.0% | 55.8% | 0.959 | 0.750 |
| Held-out, title and abstract label | 0.07 | 7 of 12 (32.0 to 80.7%) | 57.7% | 45.7% | 0.934 | 0.647 |
| Held-out, title and abstract label | 0.08 | 6 of 12 (25.4 to 74.6%) | 53.9% | 41.7% | 0.926 | 0.632 |
| Held-out, title and abstract label | 0.09 | 6 of 12 (25.4 to 74.6%) | 50.3% | 38.2% | 0.912 | 0.618 |
| Held-out, title and abstract label | 0.10 | 5 of 12 (19.3 to 68.0%) | 47.2% | 34.8% | 0.902 | 0.618 |
| Held-out, title and abstract label | 0.15 | 5 of 12 (19.3 to 68.0%) | 35.7% | 23.8% | 0.864 | 0.559 |
| Held-out, title and abstract label | 0.20 | 3 of 12 (8.9 to 53.2%) | 28.8% | 17.9% | 0.800 | 0.529 |
| Held-out, title and abstract label | 0.30 | 0 of 12 (0.0 to 24.2%) | 19.3% | 11.0% | 0.684 | 0.426 |
| Held-out, title and abstract label | 0.50 | 0 of 12 (0.0 to 24.2%) | 10.7% | 5.3% | 0.494 | 0.114 |
| CLEF 2019, title and abstract label | 0.01 | 31 of 31 (89.0 to 100.0%) | 100.0% | 99.9% | 1.000 | 1.000 |
| CLEF 2019, title and abstract label | 0.02 | 31 of 31 (89.0 to 100.0%) | 88.7% | 80.0% | 1.000 | 1.000 |
| CLEF 2019, title and abstract label | 0.03 | 31 of 31 (89.0 to 100.0%) | 68.9% | 59.4% | 0.999 | 0.984 |
| CLEF 2019, title and abstract label | 0.05 | 31 of 31 (89.0 to 100.0%) | 49.0% | 40.8% | 0.995 | 0.953 |
| CLEF 2019, title and abstract label | 0.07 | 25 of 31 (63.7 to 90.8%) | 38.3% | 30.7% | 0.982 | 0.889 |
| CLEF 2019, title and abstract label | 0.08 | 24 of 31 (60.2 to 88.6%) | 34.8% | 27.2% | 0.976 | 0.875 |
| CLEF 2019, title and abstract label | 0.09 | 24 of 31 (60.2 to 88.6%) | 31.6% | 24.2% | 0.970 | 0.797 |
| CLEF 2019, title and abstract label | 0.10 | 23 of 31 (56.8 to 86.3%) | 29.1% | 21.7% | 0.964 | 0.719 |
| CLEF 2019, title and abstract label | 0.15 | 14 of 31 (29.2 to 62.2%) | 21.1% | 14.7% | 0.914 | 0.594 |
| CLEF 2019, title and abstract label | 0.20 | 12 of 31 (23.7 to 56.2%) | 17.2% | 11.3% | 0.853 | 0.333 |
| CLEF 2019, title and abstract label | 0.30 | 7 of 31 (11.4 to 39.8%) | 12.9% | 7.7% | 0.795 | 0.167 |
| CLEF 2019, title and abstract label | 0.50 | 1 of 31 (0.6 to 16.2%) | 8.3% | 4.4% | 0.662 | 0.094 |

The value $\tau=0.07$ is the one fixed before the held-out evaluation. The values 0.08 and 0.09 were not part of the grid searched.

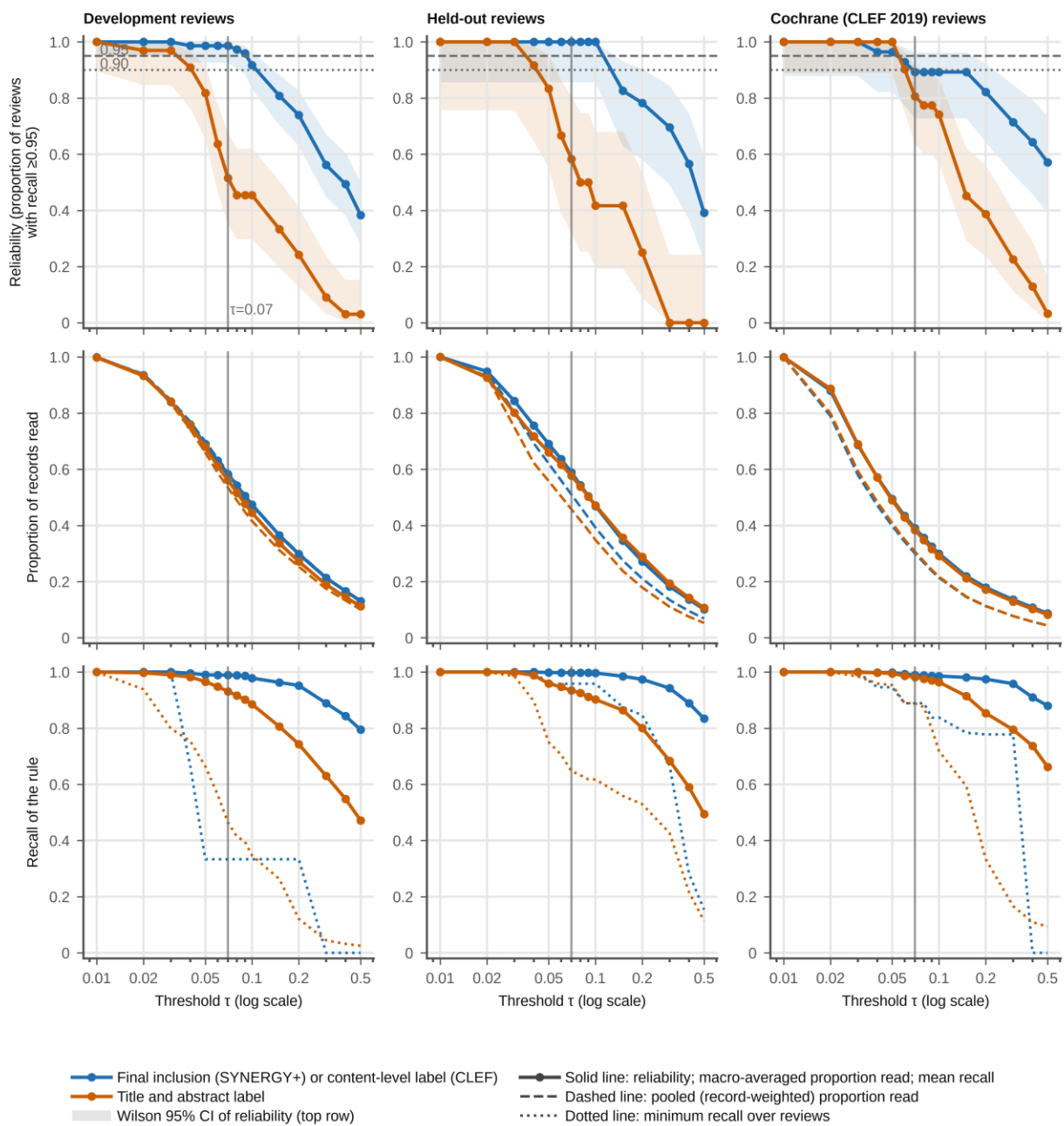

Supplementary Fig. 2 Reliability, workload, and recall of the threshold rule as a function of $\tau$ (post hoc). Columns are collections (development reviews, $n=73$, of which 33 have title and abstract labels; held-out reviews, $n=23$ and 12; Cochrane reviews of CLEF 2019, $n=28$ and 31). The vertical line marks $\tau=0.07$; the horizontal lines in the top row mark 0.95 and 0.90.

**Supplementary Table 33. Re-selection of $\tau$ in 5000 bootstrap resamples of the 73 development reviews (post hoc)**

| Grid | Selected $\tau$ | Resamples | Proportion of resamples | Held-out reviews with recall $\geq 0.95$ at this $\tau$ | Held-out records read, macro | CLEF 2019 reviews with recall $\geq 0.95$ at this $\tau$ | CLEF 2019 records read, macro |
| --- | --- | --- | --- | --- | --- | --- | --- |
| Grid searched | 0.03 | 84 | 1.7% | 23 of 23 | 84.4% | 28 of 28 | 68.7% |
| Grid searched | 0.07 | 4219 | 84.4% | 23 of 23 | 58.9% | 25 of 28 | 39.1% |
| Grid searched | 0.10 | 697 | 13.9% | 23 of 23 | 46.9% | 25 of 28 | 30.0% |
| Grid in steps of 0.01 up to 0.10 | 0.03 | 84 | 1.7% | 23 of 23 | 84.4% | 28 of 28 | 68.7% |
| Grid in steps of 0.01 up to 0.10 | 0.07 | 603 | 12.1% | 23 of 23 | 58.9% | 25 of 28 | 39.1% |
| Grid in steps of 0.01 up to 0.10 | 0.08 | 1029 | 20.6% | 23 of 23 | 54.4% | 25 of 28 | 35.6% |
| Grid in steps of 0.01 up to 0.10 | 0.09 | 2587 | 51.7% | 23 of 23 | 50.4% | 25 of 28 | 32.4% |
| Grid in steps of 0.01 up to 0.10 | 0.10 | 697 | 13.9% | 23 of 23 | 46.9% | 25 of 28 | 30.0% |

Leave-one-review-out re-selection gave $\tau=0.07$ in all 73 cases on the grid searched and 0.09 in all 73 cases on the finer grid.

In the bootstrap resamples, the re-selected values ranged from 0.03 to 0.10 on either grid. The values 0.07 to 0.10 kept recall at 95% or higher in 23 of 23 held-out and 25 of 28 CLEF reviews, and 0.03, selected in 84 resamples (1.7%), kept it in 23 of 23 and 28 of 28. The two values selected on all 73 development reviews (0.07 on the grid searched and 0.09 on the grid in steps of 0.01) both kept recall at 95% or higher in 23 of 23 held-out and 25 of 28 CLEF reviews. In all three collections, every included study below $\tau=0.07$ had a score of at least 0.03 (Supplementary Table 29); this floor was observed after the evaluation and is not a guarantee for other reviews. The included study of CD011977 that, in the proposed reading of Supplementary Table 13, contributed primary-outcome data (Kennis 2004) had a stored score of 0.03, equal to this floor at the stored precision of two decimals; it would have been read when screening down to a score of 0.03, with no margin (post hoc; analysis 29 of Supplementary Table 2).

Margins per review. In the held-out reviews, two of 23 reviews had an included study with a score below 0.07 (both reviews kept a recall of at least 95%); the largest $\tau$ that kept recall at 95% or higher had a minimum of 0.10 and a median of 0.42 over reviews. In the CLEF reviews, three of 28 reviews had a content-relevant record below 0.07; the minimum was 0.03 and the median 0.55. For title and abstract labels, seven of 12 held-out reviews and 11 of 31 CLEF reviews had a positive record below 0.07, and the medians were 0.08 and 0.12. Supplementary Tables 54 and 56 give the values per review.

Out-of-sample reliability of the selection of $\tau$ (post hoc, analysis 37). Supplementary Table 34 applies the $\tau$ selected in each bootstrap resample to the development reviews left out of that resample. These resamples were drawn with a different seed from those of Supplementary Table 33, so the counts of the selected values differ slightly. Resolution of the scores: the stored scores are multiples of 0.01 (Supplementary Note 1), so the grid in steps of 0.01 is at the resolution limit of the scores.

**Supplementary Table 34. Out-of-sample reliability of the selection of $\tau$ in 5000 bootstrap resamples of the 73 development reviews (post hoc, analysis 37)**

| Quantity, out-of-bag development reviews | Mean | Median | 2.5th to 97.5th percentile | Resamples below 0.90 |
| --- | --- | --- | --- | --- |
| Reliability (reviews with recall $\geq 0.95$) | 0.969 | 1.000 | 0.833 to 1.000 | 13.5% |
| Mean recall | 0.986 | 0.997 | 0.954 to 1.000 | 0.0% |
| Lowest recall | 0.724 | 0.955 | 0.333 to 1.000 | 42.3% |
| Records read, macro | 0.571 | 0.576 | 0.431 to 0.674 | .. |

| Development review size (records) | Out-of-bag reviews pooled over resamples | Reliable | Proportion |
| --- | --- | --- | --- |
| At most 500 | 63,889 | 60,244 | 0.943 |
| 501 to 1000 | 30,939 | 30,939 | 1.000 |
| 1001 to 2000 | 38,421 | 37,981 | 0.989 |
| All | 133,249 | 129,164 | 0.969 |

| Evaluation collection at $\tau=0.07$, review size | Reviews with recall $\geq 0.95$ (Wilson 95% CI) | Records read, macro | Records read, pooled |
| --- | --- | --- | --- |
| Held-out, at most 2000 records | 20 of 20 (83.9 to 100.0%) | 59.1% | 58.3% |
| Held-out, more than 2000 records | 3 of 3 (43.9 to 100.0%) | 57.6% | 43.5% |
| CLEF 2019, at most 2000 records | 13 of 15 (62.1 to 96.3%) | 42.7% | 34.3% |
| CLEF 2019, more than 2000 records | 12 of 13 (66.7 to 98.6%) | 34.8% | 29.4% |

In each resample, $\tau$ was selected with the rule of the plan on the grid searched and applied to the development reviews not drawn in that resample (on average 26.6 reviews). The rule selected 0.07 in 4214, 0.10 in 699, and 0.03 in 87 of the 5000 resamples. The lowest out-of-bag recall of 0.333 is that of Ferreira 2022, the only development review below 95% recall at $\tau=0.07$. $\tau$ was selected on development reviews of at most 2000 records.

Records on either side of $\tau$ under three scoring modes (post hoc, analysis 36). Supplementary Table 35 gives the records of the 2002-record subset that fell on different sides of $\tau$ under single-record requests and under the two kinds of ten-record batches.

**Supplementary Table 35. Records of the 2002-record subset below and at or above $\tau=0.07$ under three scoring modes, at the level of final inclusion and of title and abstract decisions (post hoc, analysis 36)**

| Label level and scoring mode | Positive below $\tau$ | Positive at or above $\tau$ | Negative below $\tau$ | Negative at or above $\tau$ | Reviews with recall below 0.95 |
| --- | --- | --- | --- | --- | --- |
| Final inclusion (23 reviews, 2002 records), single-record requests | 4 | 593 | 757 | 648 | 0 |
| Final inclusion (23 reviews, 2002 records), ten-record batches formed within the subset | 3 | 594 | 760 | 645 | 0 |
| Final inclusion (23 reviews, 2002 records), ten-record batches of the full run | 3 | 594 | 712 | 693 | 0 |
| Title and abstract decisions (12 reviews, 1160 records), single-record requests | 8 | 311 | 514 | 327 | 2 (Boersma-van Dam 2024, Tumkaya 2018) |
| Title and abstract decisions (12 reviews, 1160 records), ten-record batches formed within the subset | 5 | 314 | 499 | 342 | 1 (Boersma-van Dam 2024) |
| Title and abstract decisions (12 reviews, 1160 records), ten-record batches of the full run | 7 | 312 | 478 | 363 | 1 (Boersma-van Dam 2024) |

| Label level and pair of modes | Positive records on different sides of $\tau$ | Negative records on different sides of $\tau$ |
| --- | --- | --- |
| Final inclusion, single versus subset batches | 1 of 597 | 143 of 1405 |
| Final inclusion, single versus full-run batches | 1 of 597 | 151 of 1405 |
| Final inclusion, subset batches versus full-run batches | 0 of 597 | 98 of 1405 |
| Title and abstract decisions, single versus subset batches | 3 of 319 | 75 of 841 |
| Title and abstract decisions, single versus full-run batches | 1 of 319 | 78 of 841 |
| Title and abstract decisions, subset batches versus full-run batches | 2 of 319 | 51 of 841 |

At the level of final inclusion, recall at $\tau$ within the subset was at least 0.95 in all 23 reviews under each mode (lowest 0.958). At the level of title and abstract decisions (the 12 held-out reviews with these labels), recall at $\tau$ within the subset was below 0.95 in two reviews with single-record requests and in one with either batched mode (lowest 0.889 in each mode). Positive and negative records are the included and excluded records at the level of final inclusion, and the records that did and did not pass title and abstract screening at that level; at the title and abstract level, the subset holds the final inclusions and the title and abstract positives among the sampled excluded records, so its recall is not that of a full review. The subset is enriched (29.8% included), so the proportions of records below $\tau$ do not describe workload. Test-retest variation of the scores and differences between the two providers that the gateway lists for the model were not measured. Scoring times (post hoc, analysis 47; 25 September 2026): single-record requests from 09:15:48 to 10:28:38, ten-record batches formed within the subset from 10:28:38 to 10:39:29 (the same chain of commands), and the ten-record batches of the full held-out run from 00:48:48 to no later than 03:29:32; the exact end of that run and the time at which the records of the subset were scored within it are not recorded, and the provider that served the requests was not recorded in any mode. The two batched modes therefore differ in the records that share a request and also in the scoring occasion, so the differences between them do not isolate the effect of request composition on the scores.

Sensitivity to $\tau$ (analysis 5). The reliability, workload, and recall of the threshold rule were computed over the grid searched and over a finer grid in steps of 0.01, and $\tau$ was re-selected with the rule of the plan in 5000 bootstrap resamples of the 73 development reviews and with each development review left out.

Out-of-sample reliability of the selection of $\tau$ (analysis 37). In each of 5000 bootstrap resamples of the 73 development reviews, $\tau$ was selected with the rule of the plan on the grid searched and applied to the development reviews not drawn in that resample; the reliability, recall, and proportion read of these out-of-bag reviews were summarized overall and by review size (at most 500, 501 to 1000, and 1001 to 2000 records), and the held-out and Cochrane results at $\tau=0.07$ were given by review size (at most 2000 and more than 2000 records).

Scoring modes and $\tau$ (analysis 36). In the 2002-record subset, records were cross-tabulated by their side of $\tau$ under single-record requests, ten-record batches formed within the subset, and the ten-record batches of the full run, and recall at $\tau$ was computed per review under each mode.

Threshold. The label-free stopping rule reads only records with a Jev score of at least $\tau$, set at the largest value of the grid {0.01, 0.02, 0.03, 0.05, 0.07, 0.10, 0.15, 0.20, 0.30} at which at least 95% of the 73 development reviews reached a recall of 95% or higher ($\tau=0.07$). It was derived from ten-record requests formed in the order of the data files, which was not sorted by relevance or by any score, is an empirical cut-off specific to the model version and to this batching, and should be re-derived on labeled reviews if either changes; because the endpoint reports no version, a change of the model cannot be detected directly and would have to be inferred from drift monitoring (Discussion); it should not be transferred to single-record scoring without re-derivation.

#### Supplementary Note 13. Threshold rule against title and abstract labels, and in the pilot review (post hoc)

**Supplementary Table 36. Threshold rule ($\tau=0.07$) by label level (post hoc)**

| Collection and label | Reviews with recall $\geq 0.95$ (Wilson 95% CI, %) | Mean recall | Lowest recall | Pooled recall | Positive records below $\tau$ | Records read, macro | Records read, pooled | Reviews with recall below 0.95 |
| --- | --- | --- | --- | --- | --- | --- | --- | --- |
| Held-out, title and abstract label (12 reviews) | 7 of 12 (32.0 to 80.7%) | 0.934 | 0.647 | 0.936 | 61 of 948 | 57.7% | 45.7% | Boersma-van Dam 2024, Moran 2020, Roberts 2021, Theobald 2021, Tumkaya 2018 |
| Held-out, final inclusion, same 12 reviews | 12 of 12 (75.8 to 100.0%) | 0.997 | 0.958 | 0.997 | 1 of 291 | 57.7% | 45.7% | none |
| Held-out, final inclusion (23 reviews) | 23 of 23 (85.7 to 100.0%) | 0.998 | 0.958 | 0.995 | 3 of 597 | 58.9% | 51.0% | none |
| CLEF 2019, title and abstract label (31 reviews) | 25 of 31 (63.7 to 90.8%) | 0.982 | 0.889 | 0.986 | 23 of 1682 | 38.3% | 30.7% | CD011686, CD011977, CD011571, CD010558, CD006468, CD000996 |
| CLEF 2019, content-level label (28 reviews) | 25 of 28 (72.8 to 96.3%) | 0.990 | 0.889 | 0.994 | 5 of 882 | 39.1% | 30.0% | CD012233, CD011977, CD010558 |

**Supplementary Table 37. Threshold rule in the PubMed arm of the pilot review (1572 records; post hoc at the $\tau$ fixed on the development reviews; thresholds had also been evaluated on**

the pilot labels before the threshold rule was chosen: the single-record rows at $\tau=0.05$, 0.10, and 0.20 (12:06:18) and the proportion read and the recall against title and abstract labels of the ten-record rows at $\tau=0.05$, 0.10, 0.15, and 0.20 (18:45:49) were first displayed on 24 September 2026, before the threshold rule was chosen, Supplementary Note 1; aggregated counts only)

| Scoring mode | $\tau$ | Records read | Proportion read | Records that passed title and abstract screening below $\tau$ | Finally included studies below $\tau$ | Recall, title and abstract label | Recall, final inclusion |
| --- | --- | --- | --- | --- | --- | --- | --- |
| Ten-record batches | 0.03 | 1334 | 84.9% | 0 of 143 | 0 of 76 | 1.000 | 1.000 |
| Ten-record batches | 0.05 | 1119 | 71.2% | 0 of 143 | 0 of 76 | 1.000 | 1.000 |
| Ten-record batches | 0.07 | 994 | 63.2% | 1 of 143 | 0 of 76 | 0.993 | 1.000 |
| Ten-record batches | 0.10 | 875 | 55.7% | 1 of 143 | 0 of 76 | 0.993 | 1.000 |
| Ten-record batches | 0.15 | 729 | 46.4% | 1 of 143 | 0 of 76 | 0.993 | 1.000 |
| Ten-record batches | 0.20 | 621 | 39.5% | 1 of 143 | 0 of 76 | 0.993 | 1.000 |
| Single-record requests | 0.03 | 1269 | 80.7% | 0 of 143 | 0 of 76 | 1.000 | 1.000 |
| Single-record requests | 0.05 | 1054 | 67.0% | 1 of 143 | 0 of 76 | 0.993 | 1.000 |
| Single-record requests | 0.07 | 929 | 59.1% | 1 of 143 | 0 of 76 | 0.993 | 1.000 |
| Single-record requests | 0.10 | 813 | 51.7% | 2 of 143 | 0 of 76 | 0.986 | 1.000 |
| Single-record requests | 0.15 | 669 | 42.6% | 2 of 143 | 0 of 76 | 0.986 | 1.000 |
| Single-record requests | 0.20 | 574 | 36.5% | 4 of 143 | 0 of 76 | 0.972 | 1.000 |

The smallest score among the 143 records that passed title and abstract screening was 0.05 with ten-record batches and 0.04 with single-record requests; among the 76 finally included studies it was 0.20 and 0.21. The value $\tau=0.07$ was derived from batched scores.

Positive records below $\tau$ at the two label levels (post hoc, analysis 32). Supplementary Table 38 shows that most title and abstract positives below $\tau$ were excluded at full text. The question sent to Jev asks whether a record should be advanced to full-text screening and for the probability that it meets the eligibility criteria, whereas $\tau$ was tuned on final inclusion.

**Supplementary Table 38. Positive records below $\tau=0.07$ at the two label levels (post hoc, analysis 32)**

| Collection | Title and abstract positives | Of these, below $\tau$ | Of those below $\tau$, finally included (content-relevant for CLEF) | Final inclusions (content-relevant for CLEF) | Of these, below $\tau$ |
| --- | --- | --- | --- | --- | --- |
| Held-out SYNERGY+, 12 reviews with both labels | 948 | 61 | 1 | 291 | 1 |
| CLEF 2019, 31 reviews | 1,682 | 23 | 5 | 882 | 5 |
| Pilot review, PubMed arm (aggregated) | 143 | 1 | 0 | 76 | 0 |

In both collections, every final inclusion (content-relevant record) was also a title and abstract positive. For CLEF 2019, records without a content-level label count as not content-relevant. Pilot review: aggregated counts only; no record-level data are reported.

##### Supplementary Note 14. Statistical stopping criterion, label-free ranking with statistical stopping, time to discovery, recall curves, and influence of single reviews (post hoc)

The statistical criterion is described in the Methods (Stopping rules), and its cross-check against the buscarpy package is described later in this note. Supplementary Table 23 gives its reliability and workload on the ASReview, hybrid, and Jev rankings, and Supplementary Table 25 the paired differences. Label-free ranking with statistical stopping (the Jev ranking followed by the statistical criterion) was reliable in all held-out and CLEF reviews and read 87.6% and 86.1% of records; it read 28.8 percentage points (held-out) and 47.0 percentage points (CLEF) more records than the threshold rule. The criterion triggered before the last record in 1069 of the 1071 rankings; in two ASReview rankings of the smallest held-out review (Theobald 2021, 70 records), it triggered at the last record. Differences in recall at the stop (mean difference, 95% CI): held-out reviews, statistical criterion on the Jev ranking minus threshold 0.002 (0.000 to 0.007; higher in two and lower in none of 23 reviews; $P=0.18$), statistical criterion minus knee method on the ASReview ranking 0.023 (0.000 to 0.057; higher in three and lower in one of 23 reviews; $P=0.14$); CLEF 2019 reviews, statistical criterion on the Jev ranking minus threshold 0.010 (0.000 to 0.022; higher in three and lower in none of 28 reviews; $P=0.11$), statistical criterion minus knee method on the ASReview ranking 0.020 (0.000 to 0.055; higher in four and lower in one of 28 reviews; $P=0.14$).

In the reviews with fewer than 150 records (two held-out and three CLEF reviews), the knee method read all records, the statistical criterion on the ASReview ranking read 96.6% and 98.9% (held-out) and 95.9%, 97.2%, and 89.2% (CLEF), and the threshold read 71.4% and 75.7% (held-out) and 19.2%, 62.3%, and 92.4% (CLEF), all with a recall of 1.0.

##### Supplementary Table 39. Average time to discovery and recall by proportion of records read (post hoc)

| Collection | Ranking | Average time to discovery, macro mean (95% CI) | Median | Range | Recall at 5% read | at 10% | at 20% | at 30% | at 40% | at 50% |
| --- | --- | --- | --- | --- | --- | --- | --- | --- | --- | --- |
| Held-out | ASReview | 0.1006 (0.0723 to 0.1328) | 0.102 | 0.013 to 0.305 | 0.501 | 0.683 | 0.842 | 0.928 | 0.958 | 0.976 |
| Held-out | Hybrid | 0.0486 (0.0340 to 0.0643) | 0.034 | 0.007 to 0.148 | 0.702 | 0.863 | 0.963 | 0.987 | 0.997 | 0.998 |
| Held-out | Jev ranking | 0.0529 (0.0375 to 0.0694) | 0.039 | 0.009 to 0.140 | 0.684 | 0.847 | 0.948 | 0.986 | 0.996 | 0.997 |
| CLEF 2019 | ASReview | 0.0631 (0.0423 to 0.0877) | 0.044 | 0.000 to 0.244 | 0.675 | 0.819 | 0.914 | 0.957 | 0.981 | 0.992 |
| CLEF 2019 | Hybrid | 0.0394 (0.0224 to 0.0598) | 0.023 | 0.000 to 0.203 | 0.828 | 0.912 | 0.964 | 0.974 | 0.983 | 0.994 |
| CLEF 2019 | Jev ranking | 0.0448 (0.0264 to 0.0666) | 0.028 | 0.001 to 0.219 | 0.780 | 0.896 | 0.950 | 0.967 | 0.982 | 0.991 |

Average time to discovery is the mean, over included studies, of the proportion of records screened when the study is found (Ferdinands and colleagues, ref. 5).

Paired differences in the average time to discovery (mean difference, 95% CI): held-out reviews, hybrid minus ASReview $-0.0520$ ($-0.0726$ to $-0.0334$), Jev ranking minus ASReview $-0.0478$ ($-0.0704$ to $-0.0272$), hybrid minus Jev ranking $-0.0043$ ($-0.0079$ to $-0.0009$); CLEF 2019 reviews, hybrid minus ASReview $-0.0237$ ($-0.0354$ to $-0.0145$), Jev ranking minus ASReview $-0.0183$ ($-0.0308$ to $-0.0083$), hybrid minus Jev ranking $-0.0054$ ($-0.0087$ to $-0.0025$). The mean recall curves of the three rankings are shown in Supplementary Fig. 3.

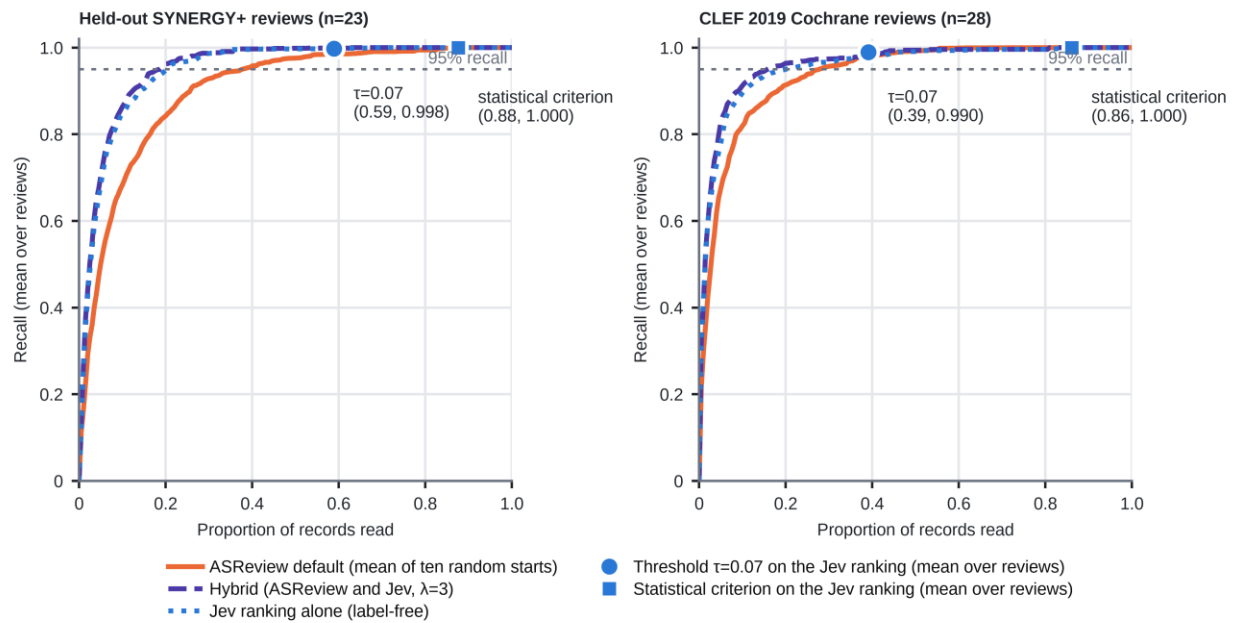

Supplementary Fig. 3 Recall by proportion of records read for the default model of ASReview, the hybrid, and the label-free Jev ranking (post hoc). Curves are means over reviews. The circle marks the threshold rule ($\tau=0.07$) and the square the statistical criterion on the Jev ranking, each at the mean proportion read and mean recall.

###### Supplementary Table 40. Influence of single reviews: each quantity recomputed with one review removed (post hoc)

| Quantity | All reviews | Smallest value with one review removed | Review removed | Largest value with one review removed | Review removed |
| --- | --- | --- | --- | --- | --- |
| C1: AUC, Jev minus bge-base (held-out) | 0.1351 | 0.1279 | Chakraborty 2023 | 0.1407 | Hall 2011 |
| C2: AUC, Jev minus GPT-4o-mini | 0.0246 | 0.0204 | Chakraborty 2023 | 0.0272 | Menon 2022 |
| C2: lower bootstrap limit of Jev minus GPT-4o-mini | 0.0101 | 0.0076 | Chakraborty 2023 | 0.0124 | Menon 2022 |
| C2: AUC, Claude Opus 5.5 minus Jev | 0.0112 | 0.0084 | Tumkaya 2018 | 0.0122 | Kerschbaumer 2024b |
| AUC, Jev minus DeepSeek-V3.1 | 0.0429 | 0.0381 | Tumkaya 2018 | 0.0470 | Menon 2022 |
| C3: WSS@95, hybrid minus ASReview | 0.1468 | 0.1328 | Theobald 2021 | 0.1542 | Parodis 2023b |
| C3: WSS@100, hybrid minus ASReview | 0.1822 | 0.1614 | Harms 2024 | 0.1923 | Tumkaya 2018 |
| WSS@95, hybrid minus Jev ranking | 0.0187 | 0.0121 | Menon 2022 | 0.0228 | Theobald 2021 |
| Wilcoxon *P* value, hybrid minus Jev ranking (WSS@95) | 0.0251 | 0.0055 | Theobald 2021 | 0.0442 | Menon 2022 |
| WSS@95, Jev ranking minus ASReview | 0.1281 | 0.1100 | Theobald 2021 | 0.1373 | Menon 2022 |
| C4: reliability of the threshold | 1.0000 | 1.0000 | Abgaz 2023 | 1.0000 | Abgaz 2023 |
| C4: proportion read, threshold | 0.5887 | 0.5713 | Chakraborty 2023 | 0.6096 | Hall 2011 |
| C4: reliability of the knee method | 0.8696 | 0.8636 | Abgaz 2023 | 0.9091 | Chakraborty 2023 |
| C4: proportion read, knee method | 0.9398 | 0.9371 | Abgaz 2023 | 0.9582 | Walker 2018 |
| Proportion read, statistical criterion on the ASReview ranking | 0.9012 | 0.8971 | Chueca 2023 | 0.9109 | Walker 2018 |
| Proportion read, statistical criterion on the Jev ranking | 0.8763 | 0.8720 | Theobald 2021 | 0.8895 | Walker 2018 |
| Proportion read, statistical criterion on the hybrid ranking | 0.8697 | 0.8651 | Theobald 2021 | 0.8838 | Walker 2018 |
| Minimum recall of the threshold | 0.9583 | 0.9583 | Abgaz 2023 | 0.9852 | Boersma-van Dam 2024 |
| CLEF 2019: WSS@95, hybrid minus ASReview | 0.0440 | 0.0376 | CD012164 | 0.0550 | CD012233 |
| CLEF 2019: WSS@100, hybrid minus ASReview | 0.0877 | 0.0799 | CD011787 | 0.1002 | CD012233 |
| CLEF 2019: AUC, Jev minus bge-base | 0.0697 | 0.0637 | CD012069 | 0.0726 | CD009044 |
| CLEF 2019: WSS@95, hybrid minus Jev ranking | 0.0249 | 0.0142 | CD010558 | 0.0265 | CD009069 |
| CLEF 2019: reliability of the threshold | 0.8929 | 0.8889 | CD000996 | 0.9259 | CD010558 |
| CLEF 2019: proportion read, threshold | 0.3905 | 0.3708 | CD012768 | 0.4033 | CD012455 |
| CLEF 2019: minimum recall of the threshold | 0.8889 | 0.8889 | CD000996 | 0.9000 | CD012233 |
| CLEF 2019: reliability of the knee method | 0.9286 | 0.9259 | CD000996 | 0.9630 | CD010038 |
| CLEF 2019: proportion read, knee method | 0.7596 | 0.7507 | CD004414 | 0.7859 | CD009044 |
| CLEF 2019: proportion read, statistical criterion on the Jev ranking | 0.8608 | 0.8563 | CD012233 | 0.8724 | CD008874 |

Criteria C1 to C4 were met with every single review removed (rows C1\_pass, C2\_pass, C3\_pass, and C4\_pass of the source file; for the tests of C1 and C3 also with exact differences). The statistical criterion was reliable in every review with every single review removed. The *P* values of the row for the hybrid minus the Jev ranking were computed from exact differences (Supplementary Note 19).

Random-sample check below $\tau$ (post hoc, analysis 42). Supplementary Table 41 compares the threshold alone, the threshold with a random-sample recall check below $\tau$ (Methods, Stopping rules), and label-free ranking with statistical stopping. The check restored reliability in every collection but read more records than label-free ranking with statistical stopping in almost every review. The second part of Supplementary Table 41 gives, for the evaluated stopping options of Table 3, how often a single run reached 95% recall (post hoc, analysis 52).

**Supplementary Table 41. Threshold with a random-sample recall check below $\tau$, compared with the threshold alone and label-free ranking with statistical stopping (post hoc, analysis 42), and single runs of the evaluated stopping options (post hoc, analysis 52)**

| Collection and label | Procedure | Reviews with recall $\geq 0.95$ (Wilson 95% CI) | Lowest recall | Records read, macro (95% CI) |
| --- | --- | --- | --- | --- |
| Development (73), final inclusion | Threshold alone | 72 of 73 (92.6 to 99.8%) | 0.333 | 58.3% (53.1 to 63.6) |
| Development (73), final inclusion | Threshold plus random-sample check | 73 of 73 (95.0 to 100.0%) | 0.984 | 93.8% (92.2 to 95.3) |
| Held-out (23), final inclusion | Threshold alone | 23 of 23 (85.7 to 100.0%) | 0.958 | 58.9% (49.1 to 68.4) |
| Held-out (23), final inclusion | Threshold plus random-sample check | 23 of 23 (85.7 to 100.0%) | 0.983 | 92.5% (88.1 to 96.1) |
| Held-out (23), final inclusion | Jev ranking plus statistical criterion | 23 of 23 (85.7 to 100.0%) | 1.000 | 87.6% (82.8 to 91.8) |
| Held-out (12), title and abstract label | Threshold alone | 7 of 12 (32.0 to 80.7%) | 0.647 | 57.7% (43.4 to 72.5) |
| Held-out (12), title and abstract label | Threshold plus random-sample check | 12 of 12 (75.8 to 100.0%) | 0.974 | 89.4% (82.9 to 94.8) |
| CLEF 2019 (28), content-level label | Threshold alone | 25 of 28 (72.8 to 96.3%) | 0.889 | 39.1% (30.5 to 47.5) |
| CLEF 2019 (28), content-level label | Threshold plus random-sample check | 28 of 28 (87.9 to 100.0%) | 0.992 | 90.6% (86.6 to 94.0) |
| CLEF 2019 (28), content-level label | Jev ranking plus statistical criterion | 28 of 28 (87.9 to 100.0%) | 1.000 | 86.1% (81.0 to 90.7) |
| CLEF 2019 (31), title and abstract label | Threshold alone | 25 of 31 (63.7 to 90.8%) | 0.889 | 38.3% (30.3 to 46.6) |
| CLEF 2019 (31), title and abstract label | Threshold plus random-sample check | 31 of 31 (89.0 to 100.0%) | 0.979 | 86.0% (81.2 to 90.1) |

Procedure: all records with a score of at least $\tau$ are read; records below $\tau$ are then drawn one at a time in random order, and after each draw the hypergeometric probability of finding at most the observed number of positive records in the sample, if enough positive records remained below $\tau$ for total recall to be below 0.95, is computed; screening stops when this probability is below 0.05 or when all records below $\tau$ have been read. Reliability is judged on the mean recall over ten random orders (seeds 0 to 9). The check read more records than label-free ranking with statistical stopping by 4.9 percentage points (95% CI 3.0 to 7.3) in held-out reviews (more in 22 of 23) and by 4.5 points (2.8 to 6.2) in CLEF reviews (more in 28 of 28). Against title and abstract labels, single random orders fell below 0.95 recall in some reviews (lowest 0.919 in held-out and 0.938 in CLEF reviews). Testing after every draw can inflate the error rate of the test. Label-free ranking with statistical stopping was not computed against title and abstract labels. The check stopped before all records below $\tau$ had been read in 95.9% of the review-seed runs of the development reviews, 95.7% of those of the held-out reviews, and in every run of the held-out reviews with title and abstract labels and of the CLEF reviews at either label level; it never stopped early in three development reviews and one held-out review, and in no review of the other collections. In the first part, the lowest recall of the random-sample check is the lowest of the per-review mean recalls over the ten orders.

Single runs of the evaluated stopping options of Table 3 (post hoc, analysis 52).

| Collection and label | Stopping option | Runs per review | Review-run pairs with recall $\geq 0.95$ | Reviews with a run below 0.95 | Lowest recall of a single run | Reviews reliable on the mean recall (Table 3) |
| --- | --- | --- | --- | --- | --- | --- |
| Held-out (23), final inclusion | Threshold $\tau=0.07$ on the Jev score | 1 | 23 of 23 (100.0%) | 0 | 0.958 (23 of 24; Boersma-van Dam 2024) | 23 of 23 |
| Held-out (23), final inclusion | Knee method as adapted (from 150 screened records), default ASReview ranking | 10 | 224 of 230 (97.4%) | 3 | 0.016 (1 of 64; Walker 2018) | 20 of 23 |
| Held-out (23), final inclusion | Knee method with the published minimum of 1000 screened records, default ASReview ranking | 10 | 230 of 230 (100.0%) | 0 | not stored | 23 of 23 |
| Held-out (23), final inclusion | Label-free ranking with statistical stopping (Jev ranking plus statistical criterion) | 10 | 230 of 230 (100.0%) | 0 | 1.000 | 23 of 23 |
| Held-out (23), final inclusion | Default ASReview ranking plus statistical criterion | 10 | 230 of 230 (100.0%) | 0 | 0.993 (134 of 135; Attai 2022) | 23 of 23 |
| Held-out (23), final inclusion | Threshold plus random-sample check below $\tau$ | 10 | 230 of 230 (100.0%) | 0 | 0.958 (23 of 24; Boersma-van Dam 2024) | 23 of 23 |
| CLEF 2019 (28), content-level label | Threshold $\tau=0.07$ on the Jev score | 1 | 25 of 28 (89.3%) | 3 | 0.889 (8 of 9; CD012233) | 25 of 28 |
| CLEF 2019 (28), content-level label | Knee method as adapted (from 150 screened records), default ASReview ranking | 10 | 273 of 280 (97.5%) | 2 | 0.111 (1 of 9; CD010038) | 26 of 28 |
| CLEF 2019 (28), content-level label | Knee method with the published minimum of 1000 screened records, default ASReview ranking | 10 | 280 of 280 (100.0%) | 0 | not stored | 28 of 28 |
| CLEF 2019 (28), content-level label | Label-free ranking with statistical stopping (Jev ranking plus statistical criterion) | 10 | 280 of 280 (100.0%) | 0 | 1.000 | 28 of 28 |
| CLEF 2019 (28), content-level label | Default ASReview ranking plus statistical criterion | 10 | 280 of 280 (100.0%) | 0 | 0.984 (241 of 245; CD012069) | 28 of 28 |
| CLEF 2019 (28), content-level label | Threshold plus random-sample check below $\tau$ | 10 | 280 of 280 (100.0%) | 0 | 0.973 (36 of 37; CD011977) | 28 of 28 |

A run is one screening of a review: the threshold is deterministic given the stored scores, with one run per review; the two knee methods and the statistical criterion on the ASReview ranking have ten random starts, label-free ranking with statistical stopping ten tie-breaking seeds of the Jev ranking, and the random-sample check ten random orders. Variation from re-scoring the records, which affects the threshold and the random-sample check directly through $\tau$ and label-free ranking with statistical stopping through the Jev ranking, is not represented in these runs, because repeated scoring of the same records under the same conditions was not measured (Supplementary Table 35). A run reaches 95% recall when at least $\lceil 0.95n \rceil$ of the $n$ included records are found. For the knee method with the published minimum, only the number of runs per review that reached 95% recall was stored. Runs below 95% recall: knee method as adapted, Chakraborty 2023 four, Walker 2018 one, van der Waal 2022 one, CD012567 six, and CD010038 one; threshold, one each in CD010558, CD011977, and CD012233. In every option and collection, the reviews not reliable on the mean recall were exactly the reviews with at least one run below 95% recall, and no review reliable on the mean recall had a run below 95% recall (comparison added after the first results had been seen). In the reviews not reliable on the mean, the knee method as adapted reached 95% recall in 24 of 30 runs in the held-out reviews (mean recall 0.657 in Chakraborty 2023, 0.902 in Walker 2018, and 0.906 in van der Waal 2022) and in 13 of 20 runs in the Cochrane reviews (0.550 in CD012567 and 0.911 in CD010038). A single run of this rule on the ASReview ranking is a simulation with random prior records. Against title and abstract labels, single orders of the random-sample check fell below 95% recall in some reviews that were reliable on the mean recall. Runs are nested within reviews, so no interval or test is given.

Single-run reliability (analysis 52). For the six evaluated stopping options of Table 3 in the held-out and Cochrane reviews, each run of a review (one run for the threshold, which is deterministic given the stored scores; ten random starts for the two knee methods and for the statistical criterion on the ASReview ranking; ten tie-breaking seeds for label-free ranking with statistical stopping; and ten random orders for the random-sample check) was counted as reaching 95% recall when at least $\lceil 0.95n \rceil$ of the $n$ included records had been found, in integer arithmetic. Variation from re-scoring the records, which affects the threshold and the random-sample check directly through $\tau$ and label-free ranking with statistical stopping through the Jev ranking, is not represented in these runs, because repeated scoring of the same records under the same conditions was not measured (Supplementary Table 35). The number and proportion of review-run pairs that reached 95% recall, the number of reviews with at least one run below 95% recall, and the lowest recall of a single run were taken from the stored per-run results; for the knee method with the published minimum and the random-sample check, the number of runs per review that reached 95% recall was stored, together with the lowest recall over runs for the random-sample check. The numbers of reviews reliable on the mean recall were checked against Table 3. After the first results had been seen, the reviews not reliable on the mean recall were compared with the reviews that had at least one run below 95% recall. No interval or test was computed, because runs are nested within reviews.

Statistical stopping criterion (post hoc). The criterion of Callaghan and Müller-Hansen (ref. 8) tests, at each point of screening, the null hypothesis that recall is below the target; screening stops when the smallest hypergeometric $P$ value over all look-back windows is below 1 minus the confidence level. It was applied with a recall target of 0.95 and a confidence level of 0.95 to the ASReview, hybrid, and Jev rankings, and label-free ranking with statistical stopping was defined as the Jev ranking, made without labels, followed by this criterion. It was implemented from the text of the publication and cross-checked against the buscarpy package (version 0.0.2, in a separate environment) at 3148 evaluation points on the hybrid rankings. At 23 of these points, at which all records had been read, the package returned no value and the implementation of this study returned $P=0$; at the other 3125 points, the largest absolute difference in $P$ values was $3.4 \times 10^{-11}$. The decision to stop differed at one point (Donners 2021, after 114 of 119 records), at which both implementations gave $P=0.0500$ and differed by $10^{-14}$. The criterion was evaluated after every record in reviews with at most 3999 records and every two to five records in the seven reviews with 4000 or more records (16 reviews had more than 2000 records). The comparison of $P$ with 0.05 was made in floating-point arithmetic. In nine of the 1071 rankings (seven ASReview rankings and two Jev rankings, in five reviews), the smallest $P$ value at the stopping point is exactly 0.05 in exact arithmetic and was computed as just below 0.05, so that screening stopped one record earlier than under a strict rule. With the strict rule, the macro-averaged proportions read change by less than 0.0001, no recall and no reliability changes, and one of the values printed per review changes (CD004414, ASReview ranking: 95.8% instead of 95.7% in Supplementary Table 56); we report the stored values. The criterion uses the labels of the screened records and therefore differs in kind from the label-free threshold.

Random-sample check below the threshold (post hoc, analysis 42). All records with a score of at least $\tau$ are read, and the records below $\tau$ are then drawn one at a time in random order. After each draw, with $k$ positive records found among the $n$ records drawn from the $M$ records below $\tau$, the hypergeometric probability of finding at most $k$ positive records is computed under the smallest number of positive records below $\tau$ that would make total recall lower than 0.95, a number that depends on the positive records found at or above $\tau$, so the stopping decision uses the screening decisions of all records read at or above $\tau$ as well as those of the sample; screening stops when this probability is below 0.05 or when all records below $\tau$ have been read. The procedure applies the test of Callaghan and Müller-Hansen to a simple random sample, without look-back windows; it was fixed before any run, reliability was judged on the mean recall over ten random orders, and testing after every draw can inflate its error rate.

#### Supplementary Note 15. Comparison with LLMs: subset, refusals, and resolution of the scores (post hoc)

The composition of the subset of 2002 records is described in the Methods (Comparators).

##### Supplementary Table 42. Composition of the subset used for the comparison with LLMs, and AUC per review on the records scored by all models (compiled post hoc)

| Review | Records in the review | Included studies | Records in the subset | Included in the subset | Excluded in the subset | AUC, Jev | AUC, Claude Opus 5.5 | AUC, GPT-4o-mini | AUC, DeepSeek-V3.1 |
| --- | --- | --- | --- | --- | --- | --- | --- | --- | --- |
| Abgaz 2023 | 288 | 11 | 23 | 11 | 12 | 1.000 | 1.000 | 0.985 | 0.985 |
| Attai 2022 | 1726 | 135 | 208 | 135 | 73 | 0.918 | 0.936 | 0.912 | 0.894 |
| Boersma-van Dam 2024 | 578 | 24 | 49 | 24 | 25 | 0.966 | 0.975 | 0.948 | 0.926 |
| Bos 2018 | 2241 | 6 | 101 | 6 | 95 | 0.982 | 0.986 | 0.975 | 0.941 |
| Bosch 2021 | 1716 | 22 | 95 | 22 | 73 | 0.981 | 0.989 | 0.925 | 0.922 |
| Chakraborty 2023 | 2233 | 7 | 102 | 7 | 95 | 0.986 | 0.985 | 0.869 | 0.850 |
| Chueca 2023 | 609 | 16 | 42 | 16 | 26 | 0.977 | 0.995 | 0.873 | 0.859 |
| Donners 2021 | 119 | 14 | 19 | 14 | 5 | 1.000 | 1.000 | 0.986 | 1.000 |
| Eggmann 2023 | 266 | 9 | 20 | 9 | 11 | 1.000 | 1.000 | 1.000 | 1.000 |
| Hall 2011 | 340 | 4 | 18 | 4 | 14 | 1.000 | 1.000 | 1.000 | 0.911 |
| Harms 2024 | 840 | 48 | 84 | 48 | 36 | 0.974 | 0.995 | 0.909 | 0.907 |
| Kerschbaumer 2024b | 1146 | 15 | 64 | 15 | 49 | 0.954 | 0.944 | 0.890 | 0.942 |
| Lewowski 2021 | 329 | 9 | 23 | 9 | 14 | 1.000 | 1.000 | 0.992 | 0.976 |
| Menon 2022 | 485 | 35 | 56 | 35 | 21 | 0.914 | 0.930 | 0.946 | 0.962 |
| Moran 2020 | 1992 | 39 | 124 | 39 | 85 | 0.954 | 0.996 | 0.890 | 0.865 |
| Parodis 2023b | 1679 | 33 | 104 | 33 | 71 | 0.966 | 0.988 | 0.972 | 0.967 |
| Roberts 2021 | 883 | 3 | 41 | 3 | 38 | 1.000 | 1.000 | 1.000 | 1.000 |
| Seghers 2022 | 1759 | 16 | 91 | 16 | 75 | 0.984 | 0.991 | 0.993 | 0.940 |
| Theobald 2021 | 70 | 9 | 14 | 9 | 5 | 0.800 | 0.811 | 0.756 | 0.778 |
| Toffalini 2021 | 328 | 22 | 36 | 22 | 14 | 1.000 | 1.000 | 0.984 | 0.935 |
| Tumkaya 2018 | 481 | 39 | 59 | 39 | 20 | 0.789 | 0.862 | 0.813 | 0.642 |
| Walker 2018 | 11,735 | 64 | 563 | 64 | 499 | 0.987 | 0.994 | 0.973 | 0.954 |
| van der Waal 2022 | 1158 | 17 | 66 | 17 | 49 | 0.983 | 0.996 | 0.959 | 0.973 |
| All 23 reviews | 33,001 | 597 | 2002 | 597 | 1405 | 0.962 | 0.973 | 0.937 | 0.919 |

AUCs were computed on the 1999 records scored by all four models (three records refused by the interface of Claude Opus 5.5 were excluded for all models). The last row gives the macro-averaged AUC. The subset was drawn before the held-out evaluation; the table was compiled post hoc.

##### Supplementary Table 43. Comparison with LLMs: sensitivity analyses (post hoc)

| Analysis | Reviews | Jev | Claude Opus 5.5 | GPT-4o-mini | DeepSeek-V3.1 | Claude Opus 5.5 minus Jev (95% CI) | Jev minus GPT-4o-mini (95% CI) | Jev minus DeepSeek-V3.1 (95% CI) |
| --- | --- | --- | --- | --- | --- | --- | --- | --- |
| Macro-average, 1999 common records (bootstrap of this analysis) | 23 | 0.9615 | 0.9727 | 0.9369 | 0.9187 | 0.0112 (0.0050 to 0.0190) | 0.0246 (0.0100 to 0.0409) | 0.0429 (0.0242 to 0.0629) |
| Macro-average, reviews with at least 20 excluded records in the subset | 16 | 0.9572 | 0.9726 | 0.9280 | 0.9090 | 0.0154 (0.0071 to 0.0257) | 0.0292 (0.0090 to 0.0511) | 0.0481 (0.0241 to 0.0735) |
| Macro-average, reviews with at least ten excluded records in the subset | 21 | 0.9674 | 0.9791 | 0.9433 | 0.9215 | 0.0117 (0.0049 to 0.0200) | 0.0241 (0.0083 to 0.0415) | 0.0459 (0.0260 to 0.0664) |
| Macro-average, all 2002 records, without Claude Opus 5.5 | 23 | 0.9615 | .. | 0.9369 | 0.9185 | .. | 0.0246 (0.0105 to 0.0406) | 0.0430 (0.0245 to 0.0637) |
| Pooled over the 1999 common records (CIs from a bootstrap of records, 2000 resamples) | .. | 0.9516 (0.9424 to 0.9596) | 0.9817 (0.9766 to 0.9857) | 0.9266 (0.9150 to 0.9379) | 0.9123 (0.8989 to 0.9243) | 0.0301 (0.0234 to 0.0372) | 0.0250 (0.0170 to 0.0337) | 0.0393 (0.0302 to 0.0488) |

CIs of the macro-averages are percentile bootstrap intervals from 5000 resamples of reviews, and CIs of the pooled values are percentile bootstrap intervals from 2000 resamples of records (seed 12345 for both). The CIs in the first row come from the bootstrap of this post-hoc analysis and differ in the third or fourth decimal from those stored with criterion C2 (Jev minus GPT-4o-mini 0.0102 to 0.0401; Claude Opus 5.5 minus Jev 0.0051 to 0.0191), which are the values reported in the main text. The pooled AUC is not the estimand named in the plan and weights large reviews (one review holds 562 of the 1999 records).

With records pooled over the 1999 common records, the deficit of Jev relative to Claude Opus 5.5 was 0.030 (0.023 to 0.037); this value also compares records between reviews, so differences between reviews in inclusion rates and score distributions enter it. In the 16 reviews with at least 20 excluded records in the subset, the macro-averaged deficit was 0.015 (0.007 to 0.026).

Refused records. The safeguards of the interface of Claude Opus 5.5 refused three records: two included records of a review of machine learning for the diagnosis of tropical diseases (Attai 2022) and one excluded record of an evidence map on the transgenerational inheritance of health effects (Walker 2018). The analysis script built the common set from the records scored by all four models, so the three records were dropped for every model. Jev scored the three records 0.22, 0.23, and 0.03. In practice, records that a model refuses should be routed to human screening.

Elicitation and resolution. The probabilities of the four models were elicited under conditions that were not equivalent (Methods, Prompts and input formatting). The models also differed in the number of distinct values that they returned on the common records: Jev (ten-record batches of the full run): 97 distinct values; tied pairs of an included and an excluded record 0.14% pooled, largest value in one review 0.7%; Claude Opus 5.5: 45 distinct values; tied pairs of an included and an excluded record 0.41% pooled, largest value in one review 6.7%; GPT-4o-mini: 1980 distinct values; tied pairs of an included and an excluded record 0.00% pooled, largest value in one review 0.0%; DeepSeek-V3.1: 13 distinct values; tied pairs of an included and an excluded record 8.80% pooled, largest value in one review 48.5%. Ties count one half in the AUC, so coarse scores lower the AUC that a model can reach. The observed difference between Claude Opus 5.5 and Jev could therefore be smaller than the difference that would be seen under equivalent conditions.

Uncertainty from the sampling of excluded records and refused records (post hoc, analysis 38). The excluded records of the subset were a sample of about 4.3% per review. Supplementary Table 44

adds this sampling to the bootstrap and scores the three refused records as 0 or 1 for Claude Opus 5.5; the interval of the deficit of Jev widened but still excluded zero.

**Supplementary Table 44. Comparison with LLMs on the 1999 common records: one-stage and two-stage bootstrap intervals and refused records (post hoc, analysis 38)**

| Quantity | Estimate | One-stage 95% CI (reviews) | Two-stage 95% CI (reviews, then excluded records within reviews) |
| --- | --- | --- | --- |
| Mean AUC, Jev | 0.962 | 0.936 to 0.982 | 0.928 to 0.984 |
| Mean AUC, GPT-4o-mini | 0.937 | 0.909 to 0.962 | 0.901 to 0.965 |
| Mean AUC, DeepSeek-V3.1 | 0.919 | 0.883 to 0.948 | 0.879 to 0.950 |
| Mean AUC, Claude Opus 5.5 | 0.973 | 0.951 to 0.990 | 0.944 to 0.991 |
| Claude Opus 5.5 minus Jev | 0.011 | 0.005 to 0.019 | 0.003 to 0.021 |
| Jev minus GPT-4o-mini | 0.025 | 0.010 to 0.041 | 0.008 to 0.043 |
| Jev minus DeepSeek-V3.1 | 0.043 | 0.024 to 0.062 | 0.023 to 0.065 |
| Claude Opus 5.5 minus Jev, all 2002 records, the three refused records given score 0 | 0.011 | 0.005 to 0.018 | .. |
| Claude Opus 5.5 minus Jev, all 2002 records, the three refused records given score 1 | 0.011 | 0.005 to 0.019 | .. |

The one-stage intervals use the same review draws as the two-stage intervals and differ slightly from those in the main article, which used a different bootstrap seed. In the two-stage bootstrap, 0.22% of the resamples gave a difference between Claude Opus 5.5 and Jev of zero or less. With the refused records scored 0 or 1, Claude Opus 5.5 was higher than Jev in 14, lower in two, and tied in seven reviews. 5000 resamples.

Calibration of the LLM scores (post hoc, analysis 38). Supplementary Table 45 describes the calibration of the four models on the enriched subset. Verbalized probabilities of language models tuned with human feedback have been reported to be better calibrated than their token probabilities (ref. 25); in this subset, 1663 of 1999 log-probability scores of GPT-4o-mini and 531 of 1999 verbalized scores of DeepSeek-V3.1 were at the clipping limits of the logit, so the recalibration slopes and intercepts of these two models depend on the clipping value and are not interpreted. No score of Jev or of Claude Opus 5.5 was at these limits; their recalibration slopes were 1.49 and 1.50.

**Supplementary Table 45. Calibration of the four models on the enriched 1999-record subset (post hoc, analysis 38)**

| Label level and model | Observed rate / mean predicted | O:E ratio (95% CI) | Recalibration slope (95% CI) | Expected calibration error | Brier score |
| --- | --- | --- | --- | --- | --- |
| Final inclusion, 23 reviews, Jev | 0.298 / 0.286 | 1.04 (0.82 to 1.27) | 1.49 (1.29 to 1.89) | 0.055 | 0.084 |
| Final inclusion, 23 reviews, GPT-4o-mini | 0.298 / 0.191 | 1.56 (1.24 to 2.22) | 0.27 (0.22 to 0.33) | 0.142 | 0.142 |
| Final inclusion, 23 reviews, DeepSeek-V3.1 | 0.298 / 0.245 | 1.21 (0.98 to 1.49) | 1.01 (0.86 to 1.23) | 0.056 | 0.101 |
| Final inclusion, 23 reviews, Claude Opus 5.5 | 0.298 / 0.198 | 1.50 (1.27 to 1.73) | 1.50 (1.37 to 1.75) | 0.099 | 0.080 |
| Title and abstract label, 12 reviews, Jev | 0.275 / 0.250 | 1.10 (0.83 to 1.56) | 1.61 (1.35 to 2.26) | 0.063 | 0.077 |
| Title and abstract label, 12 reviews, GPT-4o-mini | 0.275 / 0.141 | 1.96 (1.25 to 4.46) | 0.28 (0.19 to 0.65) | 0.158 | 0.157 |
| Title and abstract label, 12 reviews, DeepSeek-V3.1 | 0.275 / 0.193 | 1.42 (1.05 to 2.20) | 0.82 (0.63 to 1.24) | 0.084 | 0.118 |
| Title and abstract label, 12 reviews, Claude Opus 5.5 | 0.275 / 0.174 | 1.58 (1.33 to 1.87) | 1.48 (1.31 to 1.80) | 0.101 | 0.078 |

The subset is enriched with included studies (observed rate about 0.30 against 0.018 in the full held-out run), so these values describe calibration in this subset only and are not comparable with the calibration of Jev in the full held-out run (Supplementary Table 15). For GPT-4o-mini, 1663 of 1999 log-probability scores, and for DeepSeek-V3.1, 531 of 1999 verbalized scores, were at the clipping limits of the logit (analysis 38, below), so the recalibration slopes and intercepts of these two models depend on the clipping value and are not interpreted; no score of Jev or of Claude Opus 5.5 was at these limits. The effect of the clipping value was not analyzed. CIs are percentile intervals over 1000 bootstrap resamples of reviews. Probabilities were elicited under non-equivalent conditions (Methods).

Comparison with LLMs (analysis 38). A two-stage percentile bootstrap resampled reviews and then, within each drawn review, its excluded records (included records kept); in a sensitivity analysis on all 2002 records, the three records refused by Claude Opus 5.5 were given a score of 0 or 1; and the calibration of each model on the subset was described as for Jev (1000 resamples of reviews), although the subset is enriched with included studies; scores were clipped to [0.0001, 0.9999] before the logit of the recalibration model.

**Requests to the LLMs.** In the comparison subset, the number of excluded records per review ranged from five to 499 (median 36), and seven reviews had fewer than 20 excluded records. The analysis script built the common set from the records scored by all four models. The limit of about 30 requests per minute stated in the plan applied to the Jev endpoint of the gateway, and the limit of five requests per minute per model and provider applied to the chat models; the project records measured four to eight requests per minute for the chat models. In the final file of GPT-4o-mini, no record lacked both tokens, and the smallest sum of the two probabilities was above 0.9999. The log-probability prompt of GPT-4o-mini begins with the same first sentence as the verbalized prompt.

#### Supplementary Note 16. Publication year and label publicity (post hoc)

##### Supplementary Table 46. Discrimination and threshold rule by publication year of the review

(post hoc)

| Set | Publication year | Reviews | AUC, Jev | AUC, bge-base | Reviews with recall $\geq 0.95$ at $\tau=0.07$ | Minimum recall | Records read, macro |
| --- | --- | --- | --- | --- | --- | --- | --- |
| Held-out | 2011 to 2019 | 4 | 0.974 | 0.883 | 100.0% | 1.000 | 35.1% |
| Held-out | 2020 to 2021 | 7 | 0.969 | 0.824 | 100.0% | 1.000 | 50.5% |
| Held-out | 2022 | 4 | 0.953 | 0.852 | 100.0% | 0.985 | 63.3% |
| Held-out | 2023 | 5 | 0.968 | 0.805 | 100.0% | 1.000 | 80.6% |
| Held-out | 2024* | 3 | 0.959 | 0.791 | 100.0% | 0.958 | 68.0% |
| Development | 2011 to 2019 | 14 | 0.978 | 0.909 | 100.0% | 1.000 | 42.2% |
| Development | 2020 to 2021 | 21 | 0.947 | 0.835 | 100.0% | 0.955 | 56.7% |
| Development | 2022 | 12 | 0.918 | 0.859 | 91.7% | 0.333 | 60.9% |
| Development | 2023 | 12 | 0.949 | 0.855 | 100.0% | 1.000 | 60.5% |
| Development | 2024* | 14 | 0.929 | 0.783 | 100.0% | 0.968 | 72.7% |
| All 96 scored reviews | 2011 to 2019 | 18 | 0.977 | 0.904 | 100.0% | 1.000 | 40.6% |
| All 96 scored reviews | 2020 to 2021 | 28 | 0.953 | 0.832 | 100.0% | 0.955 | 55.1% |
| All 96 scored reviews | 2022 | 16 | 0.927 | 0.857 | 93.8% | 0.333 | 61.5% |
| All 96 scored reviews | 2023 | 17 | 0.955 | 0.841 | 100.0% | 1.000 | 66.4% |
| All 96 scored reviews | 2024* | 17 | 0.935 | 0.784 | 100.0% | 0.958 | 71.9% |

\*The band was defined as 2024 to 2025 before the analysis. All scored reviews of this band were published in 2024; the two reviews of the dataset that were published in 2025 are among the 18 development reviews that were not scored.

**Supplementary Table 47. Spearman rank correlation of publication year with discrimination and threshold outcomes (post hoc)**

| Set | Year and AUC of Jev | Year and difference in AUC between Jev and bge-base | Year and recall at $\tau=0.07$ | Year and proportion read at $\tau=0.07$ |
| --- | --- | --- | --- | --- |
| Held-out (23 reviews) | -0.26 ($P=0.22$) | 0.24 ($P=0.26$) | -0.28 ($P=0.19$) | 0.63 ($P=0.0014$) |
| Development (73 reviews) | -0.26 ($P=0.025$) | 0.15 ($P=0.22$) | -0.02 ($P=0.88$) | 0.38 ($P=0.0010$) |
| All 96 scored reviews | -0.25 ($P=0.014$) | 0.17 ($P=0.11$) | -0.08 ($P=0.46$) | 0.44 ($P<0.0001$) |

Discrimination was lower in reviews published more recently for Jev and for bge-base, and the difference between Jev and bge-base showed no decline with publication year. This pattern is compatible with an effect of period or topic and with exposure of the model to older reviews during training; the analysis cannot separate the two. All SYNERGY+ labels (release of 27 August 2026; first release 24 April 2023) and all CLEF 2019 labels were public before the release of the model on 15 September 2026, so only the pilot review lies outside the stratum of public labels. The mismatched-criteria control argues against a ranking that reflects recall of inclusion decisions independent of the criteria supplied; the control was run on 12 development reviews, and the AUC with mismatched criteria remained above 0.5 (0.601, 95% CI 0.544 to 0.663; post hoc,

Supplementary Table 9). The control does not exclude that the model knows which studies fit a topic. The title-only control shows that the topic alone carries most of the ranking information (AUC 0.895), which is compatible with topic knowledge and with memorization. Neither control was run on the held-out reviews or on the pilot review. No evaluation set was both released after the model and unused for development: the pilot review, whose labels are unpublished, set the prompt format, the batch size, and the shortlist of free rankers.

Same label level in three collections (title and abstract decisions): held-out SYNERGY+ reviews (12), AUC 0.903 for Jev and 0.796 for bge-base; CLEF 2019 reviews (31), 0.947 and 0.868; pilot review, 0.944 (single-record requests) or 0.942 (ten-record batches) for Jev and 0.829 for bge-base.

#### Supplementary Note 17. Protocol-stage criteria for the Cochrane reviews (post hoc)

All evaluations used eligibility criteria taken from completed reviews. Analysis 46 (methods at the end of this note) examined whether a paired comparison with criteria from the published protocols was feasible for the CLEF 2019 reviews. Too few protocols could be retrieved, so no such comparison was made, and the discrimination and the reliability of $\tau$ with protocol-stage criteria remain unknown. The files named as protocols in the CLEF 2019 repository were not used (Supplementary Table 48); with them, eight reviews would have had a text labeled as a protocol, the minimum fixed in the plan.

##### Supplementary Table 48. Retrieval of the protocol versions of the 28 Cochrane reviews with a content-relevant record (post hoc, analysis 46)

| Outcome | Reviews |
| --- | --- |
| Protocol version identified | 6 of 28 |
| Full text with the Objectives and the criteria sections retrieved | 5 of 28 (CD009069, CD012551, CD012567, CD012661, CD012669) |
| Protocol identified but full text not retrievable | 1 (CD012768; a challenge page was returned) |
| Minimum number fixed in the plan to run the paired rescoring | 8 |

The Cochrane Library and the publisher pages returned HTTP 412 and 403 for all 28 reviews. In two reviews (CD000996 and CD001261), the DOI without a version suffix belongs to the first published review, not to a protocol. The CLEF 2019 repository contains files named as protocols for three of the 28 reviews (CD011571, CD011686, and CD012164); the plan of this analysis excluded them as sources of protocol-stage criteria, because their text is written in the past tense and appears to be taken from the completed reviews. The folders and files had been listed at 19:42:42 and 19:42:47 on 28 September 2026, and two of the three files had been read in part at 19:42:55 (the first 1500 bytes of CD012164.xml with a count of its XML tags, and the first 800 bytes of the 2019 file of CD011686; CD011571 was listed only), before the plan was written and before any retrieval; had they been used, eight reviews would have had a text labeled as a protocol, the minimum fixed for the paired scoring. Because fewer than eight reviews had a usable protocol, the paired scoring with protocol-stage and abstract-stage criteria was not run, and no model call was made.

Protocol-stage criteria (analysis 46). For each of the 28 Cochrane reviews with a content-relevant record, the protocol version was searched for in the Cochrane Library, on the page of the publisher, in PubMed, in PubMed Central, and in Europe PMC, and its full text was retrieved where possible; only public identifiers were sent. The plan of this analysis fixed that the protocol criteria would be taken verbatim from the Objectives and the Criteria for considering studies for this review sections, with the template of the abstract condition, and that both criteria versions would be scored, alternating by review, in one new run only if at least eight reviews had a usable protocol; otherwise no model call would be made. The files named as protocols in the CLEF 2019 repository, which exist for three of the 28 reviews (CD011571, CD011686, and CD012164), were excluded by the plan because their text is written in the past tense and appears to be taken from the completed reviews; they had been listed, and two of them (CD011686 and CD012164) had been read in part, before the plan was written, and with them eight reviews would have had a text labeled as a protocol.

CLEF 2019. The post-hoc external validation used the 31 Cochrane reviews of the CLEF 2019 Technology Assisted Reviews test set (ref. 31). Topics and relevance files were taken from the public repository of the task (task 2, test set), and PubMed records were retrieved by PMID: of 82,421 records, 82,418 were retrieved, of which 882 were labeled relevant at the CLEF content (full-text) level. The task provides two relevance levels, an abstract level and a content level. The content-level label was not checked against the included-study and excluded-study lists of the published reviews and may depart from them in either direction; a comparison for three reviews is reported in Supplementary Note 7. We therefore describe the label as the CLEF content-level label. Three reviews (CD012342, CD011140, and CD011558) had no content-relevant record among the retrievable records and were analyzed only with title and abstract labels. The criteria were taken from the Objectives and Selection criteria sections of the abstract of each Cochrane review (latest version published by 2019, or the earliest available), which do not name the included studies; the abstracts were published in 2017 (three), 2018 (26), and 2019 (two), and the median length of the criteria was 348 characters (range 122 to 936). These sections were written after the reviews had been completed and could reflect refinements of scope made during the review. The CLEF records were not re-scored with criteria from the published protocols, because too few protocols could be retrieved from the analysis server: the Cochrane Library (HTTP 412) and the publisher's pages (HTTP 403) refused automated requests for all 28 reviews, and usable full texts were found through PubMed and PubMed Central for five (post hoc, analysis 46); the shortfall reflects access from the server, not the absence of protocols.

#### **Supplementary Note 18. Comparison with a published LLM-guided ranker (post hoc)**

**Supplementary Table 49. Jev, bge-base, and the values reported for the LGAR ranker on the 31 CLEF 2019 reviews, title and abstract labels (post hoc)**

| Metric | Jev | bge-base | LGAR (as reported in ref. 13) | Jev minus LGAR |
| --- | --- | --- | --- | --- |
| Mean average precision | 0.541 | 0.302 | 0.506 | 0.035 |
| True negative rate at 95% recall | 0.806 | 0.633 | 0.765 | 0.041 |
| Recall at 10% read | 0.784 | 0.589 | 0.767 | 0.017 |
| Recall at 20% read | 0.893 | 0.731 | 0.883 | 0.010 |

The values for LGAR are those of its publication (LLM grading with Llama-3.3-70B-Instruct followed by re-ranking). The two systems received different eligibility information, the runs are unpaired, and no test or CI is available for the difference.

**Supplementary Table 50. Inclusion rates of the two relevance files of CLEF 2019 by review type, with the rates reported for the published LLM-guided ranker (post hoc)**

| Review type | Relevance file | Topics | Records | Relevant records | Mean rate per review | Pooled rate | Rate in table 1 of the publication |
| --- | --- | --- | --- | --- | --- | --- | --- |
| Diagnostic test accuracy | full.test.dta.abs.2019.qrels | 8 | 30,521 | 440 | 7.05% | 1.44% | 7.05% |
| Diagnostic test accuracy | full.test.dta.content.2019.qrels | 8 | 30,521 | 250 | 3.31% | 0.82% | 7.05% |
| Intervention | full.test.intervention.abs.2019.qrels | 20 | 41,996 | 937 | 5.49% | 2.23% | 5.49% |
| Intervention | full.test.intervention.content.2019.qrels | 20 | 41,996 | 510 | 2.92% | 1.21% | 5.49% |
| Prognosis | full.test.prognosis.abs.2019.qrels | 1 | 3367 | 192 | 5.70% | 5.70% | 5.70% |
| Prognosis | full.test.prognosis.content.2019.qrels | 1 | 3367 | 94 | 2.79% | 2.79% | 5.70% |
| Qualitative | full.test.qualitative.abs.2019.qrels | 2 | 6537 | 113 | 1.32% | 1.73% | 1.32% |
| Qualitative | full.test.qualitative.content.2019.qrels | 2 | 6537 | 28 | 0.32% | 0.43% | 1.32% |

Files of task 2 (test set) of the public CLEF 2019 repository (commit dbc13d02bb3e); abs=abstract level, content=content level. The mean rate per review is the macro-averaged rate, and the pooled rate is the number of relevant records divided by the number of records. The rates of the publication are those of its table 1 for the four review types; its table 1 gives 6536 records for the qualitative reviews, and the relevance files hold 6537 lines without duplicates.

Label level. The limitations section of the publication describes the relevance labels of the collections as corresponding to the full-text screening stage and does not name the relevance file that was used. The inclusion rates that the publication reports for the four review types (7.05%, 5.49%, 1.32%, and 5.70%) equal the macro-averaged inclusion rates of the abstract-level relevance file of CLEF 2019 and differ from those of the content-level file (3.31%, 2.92%, 0.32%, and 2.79%; Supplementary Table 50, post hoc, analysis 27 of Supplementary Table 2). This agreement suggests that the published values refer to title and abstract labels, which is the label level of Supplementary Table 49. The remaining differences are the eligibility information (criteria and research questions extracted manually from the full texts of the reviews in the publication; two sections of the Cochrane abstract in this study) and the models. The comparison is unpaired and descriptive. No open-weight LLM ranker was run in this study.

#### Supplementary Note 19. Tests

**Supplementary Table 51. Two-sided Wilcoxon signed-rank tests across reviews (exact $P$ values and methods are post hoc)**

| Comparison | Reviews | Zero differences | Reviews higher / lower | Mean difference | $P$ value | Method | Stored in the original analysis |
| --- | --- | --- | --- | --- | --- | --- | --- |
| C1 AUC Jev vs bge-base, 23 held-out reviews | 23 | 0 | 23 / 0 | 0.1351 | <0.0001 | exact | yes |
| C2 AUC Jev vs GPT-4o-mini, 23 reviews, 1999 common records (descriptive; test added post hoc) | 23 | 3 | 16 / 4 | 0.0246 | 0.0064 | asymptotic | no (new) |
| C2 AUC Claude Opus vs Jev, 23 reviews (descriptive; test added post hoc) | 23 | 7 | 14 / 2 | 0.0112 | 0.0019 | asymptotic | no (new) |
| C2 AUC Jev vs DeepSeek, 23 reviews (descriptive; test added post hoc) | 23 | 3 | 18 / 2 | 0.0429 | 0.00059 | asymptotic | no (new) |
| C3 WSS@95 hybrid vs ASReview, 23 held-out reviews | 23 | 0 | 21 / 2 | 0.1468 | <0.0001 | exact | yes |
| C3 WSS@100 hybrid vs ASReview, 23 held-out reviews (test added post hoc) | 23 | 0 | 21 / 2 | 0.1822 | <0.0001 | exact | no (new) |
| Hybrid vs comparator named in the plan, logistic regression, WSS@95, 23 held-out reviews | 23 | 0 | 21 / 2 | 0.1675 | <0.0001 | exact | yes |
| Hybrid vs comparator named in the plan, logistic regression, WSS@100, 23 held-out reviews (test added post hoc) | 23 | 0 | 21 / 2 | 0.2083 | <0.0001 | exact | no (new) |
| Hybrid vs comparator named in the plan, naive Bayes, WSS@95, 23 held-out reviews | 23 | 0 | 21 / 2 | 0.1720 | <0.0001 | exact | yes |
| Hybrid vs comparator named in the plan, naive Bayes, WSS@100, 23 held-out reviews (test added post hoc) | 23 | 0 | 20 / 3 | 0.2236 | <0.0001 | exact | no (new) |
| CLEF 2019 AUC Jev vs bge-base, content-level label, 28 reviews | 28 | 0 | 26 / 2 | 0.0697 | <0.0001 | exact | yes |
| CLEF AUC Jev vs bge-base, title and abstract labels, 31 reviews | 31 | 0 | 27 / 4 | 0.0784 | <0.0001 | exact | yes |
| CLEF 2019 WSS@95 hybrid vs ASReview, 28 reviews | 28 | 0 | 24 / 4 | 0.0440 | 0.00015 | exact | yes |
| CLEF 2019 WSS@100 hybrid vs ASReview, 28 reviews | 28 | 0 | 25 / 3 | 0.0877 | <0.0001 | exact | yes |
| CLEF 2019 WSS@95 hybrid vs weak supervision, 28 reviews | 28 | 0 | 23 / 5 | 0.0202 | 0.00031 | exact | yes |
| CLEF 2019 WSS@100 hybrid vs weak supervision, 28 reviews | 28 | 0 | 22 / 6 | 0.0334 | 0.0011 | exact | yes |
| P1 WSS@95 hybrid vs weak supervision, 23 | 23 | 0 | 21 / 2 | 0.0595 | 0.00021 | exact | yes |
| P1 WSS@95 weak supervision vs ASReview, 23 | 23 | 0 | 18 / 5 | 0.0873 | 0.00064 | exact | yes |
| P1 WSS@100 hybrid vs weak supervision, 23 | 23 | 0 | 21 / 2 | 0.0932 | <0.0001 | exact | yes |
| P1 WSS@100 weak supervision vs ASReview, 23 | 23 | 0 | 17 / 6 | 0.0890 | 0.0024 | exact | yes |
| P4 WSS@95 strongest preset plus Jev vs strongest preset, 23 | 23 | 0 | 21 / 2 | 0.0822 | 0.00013 | exact | yes |
| P4 WSS@95 strongest preset vs default, 23 | 23 | 0 | 14 / 9 | 0.0655 | 0.080 | exact | yes |
| P4 WSS@100 strongest preset plus Jev vs strongest preset, 23 | 23 | 0 | 19 / 4 | 0.1147 | 0.00074 | exact | yes |
| P3 title and abstract labels WSS@95 hybrid vs ASReview, 12 | 12 | 0 | 9 / 3 | 0.0952 | 0.034 | exact | yes |

| Comparison | Reviews | Zero differences | Reviews higher / lower | Mean difference | *P* value | Method | Stored in the original analysis |
| --- | --- | --- | --- | --- | --- | --- | --- |
| P3 title and abstract labels WSS@100 hybrid vs ASReview, 12 | 12 | 0 | 8 / 4 | 0.1441 | 0.042 | exact | yes |
| P2 AUC single-record vs full-run batches, 23 | 23 | 7 | 4 / 12 | -0.0014 | 0.30 | asymptotic | yes |
| P2 AUC subset batches vs full-run batches, 23 | 23 | 9 | 9 / 5 | 0.0010 | 0.51 | asymptotic | yes§ |

SciPy 1.18.1 with default settings, applied to exact differences (described later in this note): zero differences are discarded; the exact distribution is used when there are no ties or zero differences (and at most 50 reviews), otherwise the normal approximation without continuity correction. Tests for criterion C2 are descriptive; the plan judged C2 on the bootstrap CI and on the point estimate. §The values stored by the original analysis for this comparison were computed from floating-point differences: $P=0.46$ with ten reviews higher, five lower, and eight zero differences.

All 22 bootstrap CIs and all 21 $P$ values stored by the original analysis were reproduced from the original scripts and seeds with floating-point differences. With exact differences (described later in this note), 20 of the 21 $P$ values were unchanged and one changed (last row of Supplementary Table 51). New CIs for the criteria: difference in WSS@100 between the hybrid and ASReview 0.182 (0.115 to 0.258; hybrid higher in 21 of 23 reviews). This interval continues the sequence of random numbers of the original script; as the log had announced, a second interval was computed with a separate seed and was 0.114 to 0.256. For the comparators named in the plan, the script and the seed of the stored intervals were not recorded, so new intervals were computed with the separate seed: differences in WSS@95 between the hybrid and logistic regression and naive Bayes active learning 0.167 (0.106 to 0.239) and 0.172 (0.110 to 0.244), against the stored intervals of 0.105 to 0.235 and 0.108 to 0.239 given in Supplementary Note 5; differences in WSS@100 0.208 (0.135 to 0.287; hybrid higher in 21 of 23 reviews) and 0.224 (0.141 to 0.310; 20 of 23). Further results. Per-review counts for criterion C2 on the 1999 common records: Jev higher than GPT-4o-mini in 16 reviews, lower in four, and tied in three; Claude Opus 5.5 higher than Jev in 14, lower in two, and tied in seven; Jev higher than DeepSeek-V3.1 in 18, lower in two, and tied in three.

Recount in exact arithmetic (post hoc, 27 September 2026; method described later in this note). In the original analyses, some reviews in which the two methods had exactly the same value were counted as higher or lower because the two means differed by a rounding residual of about $10^{-16}$, and the same residuals entered the tests as non-zero differences. Of the 372 comparisons stored by the analyses (a comparison that two analyses computed is counted twice), the numbers of reviews changed in 81 and the $P$ value changed at the printed precision in 61 (Supplementary Tables 19, 25, 40, 51, and 57). The tests of criteria C1 and C3 had no zero or tied differences and were unchanged. No floating-point difference lay between $10^{-12}$ and $10^{-7}$ in absolute value. A simpler rule that sets floating-point differences of at most $10^{-12}$ to zero gave the same numbers of reviews in all 372 comparisons; because the exact computation also keeps ties between absolute differences, its $P$ value differed from that of the simpler rule at the printed precision in seven comparisons on development reviews in Supplementary Table 57.

Zero differences and confirmatory tests (post hoc, analyses 39 and 49). Supplementary Table 52 repeats the paired comparisons of the main article with Pratt's treatment of zero differences and with a sign-flip permutation test that retains zeros, labels each $P$ value as confirmatory or exploratory, states where the 383 comparisons of analysis 39 are printed, and lists the 43 further printed comparisons of analyses 32 to 46 to which analysis 49 applied the same tests.

**Supplementary Table 52. Paired comparisons of the main article under three treatments of zero differences (post hoc, analysis 39)**

| Comparison | Where reported | Label | Zero differences | Wilcoxon, zeros discarded (reported) | Wilcoxon, Pratt | Sign-flip permutation of the mean |
| --- | --- | --- | --- | --- | --- | --- |
| Held-out, hybrid minus Jev ranking, WSS@95 | Results | exploratory | 3 of 23 | 0.025 | 0.017 | 0.039 |
| Held-out, Jev ranking minus ASReview, WSS@95 | Results | exploratory | 0 of 23 | 0.00048 | 0.00048 | 0.00039 |
| CLEF, hybrid minus Jev ranking, WSS@95 | Discussion | exploratory | 2 of 28 | 0.0043 | 0.0061 | 0.0011 |
| CLEF, Jev ranking minus ASReview, WSS@95 | Results | exploratory | 0 of 28 | 0.032 | 0.032 | 0.359 |
| Held-out, records read, statistical criterion on the Jev ranking minus threshold | Results | exploratory | 0 of 23 | <0.0001 | <0.0001 | <0.0001 |
| Held-out, records read, statistical criterion, Jev minus ASReview ranking | Discussion | exploratory | 2 of 23 | 0.0051 | 0.0044 | 0.015 |
| Held-out, average time to discovery, Jev ranking minus ASReview | Discussion | exploratory | 0 of 23 | <0.0001 | <0.0001 | <0.0001 |
| CLEF, records read, statistical criterion on the Jev ranking minus threshold | Results | exploratory | 0 of 28 | <0.0001 | <0.0001 | <0.0001 |
| CLEF, records read, statistical criterion, Jev minus ASReview ranking | Discussion | exploratory | 3 of 28 | 0.135 | 0.116 | 0.275 |
| Held-out, records read, threshold minus knee as adapted (C4) | Table 2, C4 records read | exploratory | 0 of 23 | <0.0001 | <0.0001 | <0.0001 |
| Held-out, AUC, Jev minus bge-base (C1) | Table 2, C1; Results | confirmatory | 0 of 23 | <0.0001 | <0.0001 | <0.0001 |
| AUC, Jev minus GPT-4o-mini (C2) | Table 2, C2 | exploratory | 3 of 23 | 0.0064 | 0.0056 | 0.0035 |
| AUC, Claude Opus 5.5 minus Jev (C2) | Table 2, C2 | exploratory | 7 of 23 | 0.0019 | 0.0016 | 0.00061 |
| Held-out, WSS@95, hybrid minus ASReview (C3) | Table 2, C3; Results | confirmatory | 0 of 23 | <0.0001 | <0.0001 | <0.0001 |
| Held-out, WSS@100, hybrid minus ASReview (C3) | Table 2, C3 | exploratory | 0 of 23 | <0.0001 | <0.0001 | <0.0001 |
| CLEF, AUC, Jev minus bge-base | Results | exploratory | 0 of 28 | <0.0001 | <0.0001 | <0.0001 |
| CLEF, WSS@95, hybrid minus ASReview | Results | exploratory | 0 of 28 | 0.00015 | 0.00015 | 0.0067 |
| Held-out, reliability, threshold minus knee as adapted (C4) | Table 2, C4 reliability | exploratory | 20 of 23 | 0.083 | 0.083 | 0.250 |
| CLEF, reliability, threshold minus knee as adapted | Results | exploratory | 23 of 28 | 0.655 | 0.655 | 1.000 |

The confirmatory $P$ values are those of the tests named in the decision criteria of the plan: C1 (Wilcoxon $P<0.01$) and the WSS@95 part of C3 ($P<0.05$); all other $P$ values are exploratory. The composite decision rule with fixed thresholds and disjunctions does not control the overall type I error rate. The sign-flip test permutes the signs of the per-review differences, zeros retained, by exact enumeration when at most 20 differences are non-zero and otherwise with 100,000 random sign patterns. Analysis 39 repeated 383 paired comparisons: those of Supplementary Tables 19

(75), 58 (222), 52 (27), and 26 (31), of the text of Supplementary Note 14 (10; the $P$ values of its six differences in the average time to discovery are not printed there), of analyses 33 (Supplementary Table 24; eight) and 34 (Supplementary Table 21; eight), and two tests on the development reviews whose $P$ values are not printed (post hoc, analysis 49); 284 had at least one zero difference, and the conclusion at the 0.05 level changed in 26 (Pratt in 10, sign flip in 17; seven of the 26 had no zero differences, so that the change reflects the test statistic). Among the comparisons of the main article and Table 2, it changed in one: the Jev ranking minus ASReview in CLEF reviews (WSS@95). The recount reproduced the counts and $P$ values of the 372 stored comparisons.

Further comparisons (post hoc, analysis 49). Of the 62 printed Wilcoxon comparisons of analyses 32 to 46, 19 were among the 383 and 43 were not; the same three tests were applied to these 43.

| Table | Comparison | Reviews | Zero differences | Wilcoxon, zeros discarded (reported) | Wilcoxon, Pratt | Sign-flip permutation of the mean | Conclusion at 0.05 changed |
| --- | --- | --- | --- | --- | --- | --- | --- |
| ST24 | Held-out, fewer than 1000 records, records read, threshold minus knee as adapted (150), ASReview | 13 | 0 | 0.00024 | 0.00024 | 0.00024 | No |
| ST24 | Held-out, fewer than 1000 records, records read, threshold minus knee (1000), ASReview | 13 | 0 | 0.00024 | 0.00024 | 0.00024 | No |
| ST24 | Held-out, fewer than 1000 records, records read, knee (1000) minus knee as adapted (150), ASReview | 13 | 11 | 0.500 | 0.500 | 0.500 | No |
| ST24 | Held-out, 1000 or more records, records read, threshold minus knee as adapted (150), ASReview | 10 | 0 | 0.020 | 0.020 | 0.021 | No |
| ST24 | Held-out, 1000 or more records, records read, threshold minus knee (1000), ASReview | 10 | 0 | 0.0020 | 0.0020 | 0.0020 | No |
| ST24 | Held-out, 1000 or more records, records read, knee (1000) minus knee as adapted (150), ASReview | 10 | 7 | 0.250 | 0.250 | 0.250 | No |
| ST24 | Held-out, at most 2000 records, records read, threshold minus knee as adapted (150), ASReview | 20 | 0 | <0.0001 | <0.0001 | <0.0001 | No |
| ST24 | Held-out, at most 2000 records, records read, threshold minus knee (1000), ASReview | 20 | 0 | <0.0001 | <0.0001 | <0.0001 | No |
| ST24 | Held-out, at most 2000 records, records read, knee (1000) minus knee as adapted (150), ASReview | 20 | 17 | 0.109 | 0.084 | 0.250 | No |
| ST24 | Held-out, more than 2000 records, records read, threshold minus knee as adapted (150), ASReview | 3 | 0 | 0.750 | 0.750 | 0.750 | No |
| ST24 | Held-out, more than 2000 records, records read, threshold minus knee (1000), ASReview | 3 | 0 | 0.250 | 0.250 | 0.250 | No |
| ST24 | Held-out, more than 2000 records, records read, knee (1000) minus knee as adapted (150), ASReview | 3 | 1 | 0.500 | 0.500 | 0.500 | No |
| ST24 | CLEF, fewer than 1000 records, records read, threshold minus knee as adapted (150), ASReview | 11 | 0 | 0.00098 | 0.00098 | 0.00098 | No |
| ST24 | CLEF, fewer than 1000 records, records read, threshold minus knee (1000), ASReview | 11 | 0 | 0.00098 | 0.00098 | 0.00098 | No |
| ST24 | CLEF, fewer than 1000 records, records read, knee (1000) minus knee as adapted (150), ASReview | 11 | 8 | 0.250 | 0.250 | 0.250 | No |
| ST24 | CLEF, 1000 or more records, records read, threshold minus knee as adapted (150), ASReview | 17 | 0 | 0.00084 | 0.00084 | 0.00079 | No |
| ST24 | CLEF, 1000 or more records, records read, threshold minus knee (1000), ASReview | 17 | 0 | 0.00021 | 0.00021 | 0.00023 | No |
| ST24 | CLEF, 1000 or more records, records read, knee (1000) minus knee as adapted (150), ASReview | 17 | 6 | 0.013 | 0.017 | 0.0068 | No |
| ST24 | CLEF, at most 2000 records, records read, threshold minus knee as adapted (150), ASReview | 15 | 0 | <0.0001 | <0.0001 | <0.0001 | No |
| ST24 | CLEF, at most 2000 records, records read, threshold minus knee (1000), ASReview | 15 | 0 | <0.0001 | <0.0001 | <0.0001 | No |
| ST24 | CLEF, at most 2000 records, records read, knee (1000) minus knee as adapted (150), ASReview | 15 | 10 | 0.043 | 0.026 | 0.062 | Yes |
| ST24 | CLEF, more than 2000 records, records read, threshold minus knee as adapted (150), ASReview | 13 | 0 | 0.0081 | 0.0081 | 0.0085 | No |
| ST24 | CLEF, more than 2000 records, records read, threshold minus knee (1000), ASReview | 13 | 0 | 0.0034 | 0.0034 | 0.0034 | No |
| ST24 | CLEF, more than 2000 records, records read, knee (1000) minus knee as adapted (150), ASReview | 13 | 4 | 0.039 | 0.055 | 0.027 | Yes |
| ST21 | CLEF, recall at 10% read, hybrid (ten-seed mean) minus Jev ranking | 28 | 17 | 0.016 | 0.0072 | 0.0068 | No |
| ST22 | Held-out, TNR@95, hybrid minus ASReview | 23 | 0 | <0.0001 | <0.0001 | <0.0001 | No |
| ST22 | Held-out, TNR@95, Jev ranking minus ASReview | 23 | 0 | 0.00048 | 0.00048 | 0.00041 | No |
| ST22 | Held-out, TNR@95, hybrid minus Jev ranking | 23 | 3 | 0.025 | 0.017 | 0.046 | No |
| ST22 | Held-out, TNR@95, Jev ranking minus bge-base | 23 | 0 | <0.0001 | <0.0001 | <0.0001 | No |
| ST22 | CLEF, TNR@95, hybrid minus ASReview | 28 | 0 | 0.00015 | 0.00015 | 0.0069 | No |
| ST22 | CLEF, TNR@95, Jev ranking minus ASReview | 28 | 0 | 0.030 | 0.030 | 0.387 | Yes |
| ST22 | CLEF, TNR@95, hybrid minus Jev ranking | 28 | 2 | 0.0034 | 0.0043 | 0.00099 | No |
| ST22 | CLEF, TNR@95, Jev ranking minus bge-base | 28 | 0 | <0.0001 | <0.0001 | 0.00015 | No |
| ST11 | Held-out, AUC, MedCPT minus bge-base | 23 | 0 | 0.0017 | 0.0017 | 0.00085 | No |
| ST11 | Held-out, AUC, Jev minus MedCPT | 23 | 0 | <0.0001 | <0.0001 | <0.0001 | No |
| ST11 | CLEF, content-level label, AUC, MedCPT minus bge-base | 28 | 0 | <0.0001 | <0.0001 | <0.0001 | No |
| ST11 | CLEF, content-level label, AUC, Jev minus MedCPT | 28 | 0 | <0.0001 | <0.0001 | <0.0001 | No |
| ST11 | CLEF, title and abstract label, AUC, MedCPT minus bge-base | 31 | 0 | <0.0001 | <0.0001 | <0.0001 | No |
| ST11 | CLEF, title and abstract label, AUC, Jev minus MedCPT | 31 | 0 | <0.0001 | <0.0001 | <0.0001 | No |
| ST11 | Held-out, biomedical reviews, AUC, MedCPT minus bge-base | 10 | 0 | 0.0059 | 0.0059 | 0.0059 | No |
| ST11 | Held-out, biomedical reviews, AUC, Jev minus MedCPT | 10 | 0 | 0.0020 | 0.0020 | 0.0020 | No |
| ST11 | Held-out, other reviews, AUC, MedCPT minus bge-base | 13 | 0 | 0.168 | 0.168 | 0.108 | No |
| ST11 | Held-out, other reviews, AUC, Jev minus MedCPT | 13 | 0 | 0.00024 | 0.00024 | 0.00024 | No |

ST, Supplementary Table. The recomputed default $P$ value equaled the stored and the printed value in 62 of the 62 printed comparisons. Of the 43 further comparisons, 11 had at least one zero difference, and the conclusion at the 0.05 level changed in three (Pratt in one, sign flip in two). With the extension, 426 comparisons were checked, 295 with at least one zero difference. The 21 McNemar comparisons of reliability in Supplementary Table 24 (63 printed $P$ values) are not Wilcoxon tests and were not repeated. The sign-flip test used exact enumeration when at most 20 differences were non-zero and otherwise 100,000 random sign patterns (seed 20260928); $P$ values are exploratory.

Scope of the zero-difference check (analysis 49). Each of the 383 comparisons of analysis 39 was assigned to the table or text in which it is printed, and its printed $P$ value was searched for there. Every printed Wilcoxon comparison of analyses 32 to 46 was listed from the record of the values printed in the tables; its default Wilcoxon $P$ value was recomputed from the stored per-review values and compared with the stored and printed values; a comparison with the same reviews and identical differences as one of the 383 was counted as covered, and the others were given the three tests of analysis 39 with the same code and seed.

**Outcome definitions.** The primary outcomes were the AUC, work saved over sampling at 95% recall (WSS@95) (ref. 2), and, for stopping rules, reliability (the proportion of reviews with recall of at least 95%) and workload (the proportion of records read). AUC was computed per review with scikit-learn; tied scores receive mid-ranks, so a tie between an included and an excluded record counts one half. $WSS@95 = 1 - k_{95}/N - 0.05$, where $k_{95}$ is the rank of the $[0.95n]$-th of the $n$ included records, so that in reviews with at most 19 included records 95% recall means all included records (13 of 23 held-out and 15 of 28 Cochrane reviews; post hoc, analysis 35). A secondary outcome was WSS at 100% recall ($WSS@100 = 1 - k_{100}/N$). Mean average precision follows the definition of the CLEF task as used by the LGAR study (ref. 13); the implementation was not verified line by line against that publication. The true negative rate at 95% recall (TNR@95) is the proportion of excluded records not yet read when the $[0.95n]$-th included record is found; post hoc (analysis 35), it was computed for all rankings. Normalized WSS has been shown to be equivalent to the true negative rate, and TNR at 95% recall has been proposed as a measure (ref. 34); no verification by the authors of its rounding of the 95% recall point against this definition is recorded, and a comparison with the journal pre-proof of that article is reported in Supplementary Note 22 (post hoc, analyses 50 and 51). The average time to discovery (post hoc, analysis 8) is the mean, over included studies, of the proportion of records screened when the study is found (ref. 5). Macro-averaged proportions read (the mean of the proportions of the single reviews) were used for the criteria; pooled proportions (records read in all reviews divided by all records) are reported as well.

**Intervals and tests.** Metrics were computed within each review, averaged over seeds for simulations, and then averaged across reviews (macro-averages). The 95% CIs for means and paired differences are percentile bootstrap intervals (2.5th and 97.5th percentiles of the mean over 5000 resamples of reviews with replacement) unless stated otherwise. Two sets of intervals from the post-hoc analyses used other settings: the CIs of the calibration measures are percentile bootstrap intervals from 1000 resamples of reviews, and the CIs of the pooled AUCs and of their differences (Supplementary Table 43) are percentile bootstrap intervals from 2000 resamples of records; these intervals were not recomputed with 5000 resamples of reviews. For proportions of reviews, we report Wilson intervals and, where stated, exact (Clopper-Pearson) limits. For 23 of 23 reviews, the two-sided 95% Wilson interval is 85.7% to 100% and the exact interval is 85.2% to 100%; the one-sided exact lower 95% confidence limit is 87.8%. The post-hoc analyses 32 to 46 used 5000 bootstrap resamples of reviews with the seed 20260928 unless stated, so their intervals for quantities that are also reported elsewhere can differ slightly.

Paired comparisons used two-sided Wilcoxon signed-rank tests across reviews (SciPy 1.18.1, default settings): zero differences are discarded; the exact distribution is used when there are no ties or zero differences and at most 50 reviews; when ties or zero differences are present, the $P$ value is obtained by enumeration for at most 13 reviews and by the normal approximation without continuity correction for more reviews. The tests and their $P$ values are given in Supplementary Tables 6, 8, 11, 19, 21, 22, 24 to 26, 40, 47, 51, 52, and 57 and in the text of Supplementary Notes 3, 4, 5, 10, and 14; Supplementary Table 51 gives the method of the Wilcoxon tests of the original analyses. The $P$ values of the two tests named in the decision criteria, C1 ($P < 0.01$) and the WSS@95 comparison of C3 ($P < 0.05$), are confirmatory in the sense of the plan; all other $P$ values, including those of all post-hoc analyses, are exploratory, and no adjustment was made for multiplicity. The decision rule of the plan required that (C1 or C2) and (C3 or C4) hold on the held-out reviews and that the robustness check be passed (Supplementary Note 23); it combined criteria with fixed thresholds and disjunctions and does not control the overall type I error rate. If a requirement was not met, the plan foresaw a further iteration, without stating whether the held-out reviews would be reused. All requirements were met (Table 2), so no further iteration was run, and the held-out reviews were not reused for one. For paired comparisons of reliability, the Wilcoxon signed-rank test on the paired indicators of reliability equals the asymptotic McNemar test without continuity correction. Post hoc (analysis 23 of Supplementary Table 2), we added the exact conditional and mid-p McNemar tests, which were compared by Fagerland, Lydersen, and Laake (ref. 35), who recommend the mid-p test and advise against the exact conditional test, and two score intervals for the difference of paired proportions (Tango (ref. 36) and Newcombe method 10 (ref. 37)). The formulas were taken from the R package *contingencytables* (version 3.1.0), and the implementation reproduced the 18 test values of the package. The full texts of the two papers of 1998 were not available, and the methods were checked against their abstracts and the package source. Post hoc (analysis 39), 383 paired comparisons were repeated with Pratt's treatment of zero differences and with a sign-flip permutation test of the mean difference that retains zeros (exact enumeration when at most 20 differences are non-zero, otherwise 100,000 random sign patterns): those of Supplementary Tables 19, 25, 51, and 57 and of the text of Supplementary Note 14, eight each of analyses 33 and 34 (Supplementary Tables 24 and 21), and two tests on the development reviews whose $P$ values are not printed. Post hoc (analysis 49), the same three tests were applied to the 43 other printed Wilcoxon comparisons of analyses 32 to 46 (Supplementary Tables 11, 21, 22, and 24), so that 426 comparisons were checked; the McNemar tests of reliability in Supplementary Table

24 were not repeated.

Reviews in which two methods had the same value. The original analyses counted the reviews in which one method was higher, lower, or tied from differences between floating-point means, in which rounding residuals made some equal values count as different. In a post-hoc recount on 27 September 2026, every metric was rebuilt as a fraction of integers (ranks, numbers of records, and numbers of included records; for the AUC, numbers of pairs of an included and an excluded record) and averaged over seeds as fractions, the paired differences were computed exactly, the reviews were counted by the sign of the exact difference, and the tests were applied to the exact differences. All numbers of reviews and all *P* values of paired comparisons in this article and its Supplementary Information come from this computation; mean differences and bootstrap CIs were not affected, and the tests of criteria C1 and C3 and the decisions on the four criteria were unchanged (Supplementary Note 19).

Post hoc, calibration was assessed with the observed-to-expected ratio and the slope of a logistic recalibration model (ref. 38), and prediction intervals for a new review were derived from a random-effects model (ref. 39).

Software. Analyses used Python 3.12 (scikit-learn 1.9.1, NumPy 2.5.3, SciPy 1.18.1, pandas 2.3.3, ASReview 3.0.8) and Node.js 20, on one server with two 12-core Intel Xeon Gold 6126 processors.

#### Supplementary Note 20. Results per review

**Supplementary Table 53. Held-out SYNERGY+ reviews: ranking outcomes per review (the Jev ranking is post hoc)**

| Review | Records | Included | Rate | AUC, Jev | AUC, bge-base | WSS@95, ASReview | WSS@95, Jev ranking | WSS@95, hybrid | WSS@100, ASReview | WSS@100, Jev ranking | WSS@100, hybrid |
| --- | --- | --- | --- | --- | --- | --- | --- | --- | --- | --- | --- |
| Abgaz 2023 | 288 | 11 | 3.82% | 0.986 | 0.898 | 0.701 | 0.887 | 0.874 | 0.751 | 0.937 | 0.924 |
| Attai 2022 | 1726 | 135 | 7.82% | 0.912 | 0.802 | 0.556 | 0.656 | 0.694 | 0.136 | 0.437 | 0.474 |
| Boersma-van Dam 2024 | 578 | 24 | 4.15% | 0.953 | 0.902 | 0.629 | 0.784 | 0.813 | 0.673 | 0.681 | 0.779 |
| Bos 2018 | 2241 | 6 | 0.27% | 0.992 | 0.959 | 0.902 | 0.903 | 0.926 | 0.952 | 0.953 | 0.976 |
| Bosch 2021 | 1716 | 22 | 1.28% | 0.977 | 0.792 | 0.869 | 0.872 | 0.890 | 0.736 | 0.893 | 0.886 |
| Chakraborty 2023 | 2233 | 7 | 0.31% | 0.981 | 0.688 | 0.717 | 0.906 | 0.916 | 0.767 | 0.956 | 0.966 |
| Chueca 2023 | 609 | 16 | 2.63% | 0.916 | 0.715 | 0.174 | 0.601 | 0.625 | 0.224 | 0.651 | 0.675 |
| Donners 2021 | 119 | 14 | 11.76% | 0.982 | 0.924 | 0.660 | 0.746 | 0.790 | 0.710 | 0.796 | 0.840 |
| Eggmann 2023 | 266 | 9 | 3.38% | 1.000 | 0.987 | 0.865 | 0.916 | 0.916 | 0.915 | 0.966 | 0.966 |
| Hall 2011 | 340 | 4 | 1.18% | 0.995 | 0.984 | 0.929 | 0.932 | 0.932 | 0.979 | 0.982 | 0.982 |
| Harms 2024 | 840 | 48 | 5.71% | 0.976 | 0.787 | 0.351 | 0.776 | 0.800 | 0.128 | 0.783 | 0.768 |
| Kerschbaumer 2024b | 1146 | 15 | 1.31% | 0.947 | 0.683 | 0.701 | 0.746 | 0.791 | 0.751 | 0.796 | 0.841 |
| Lewowski 2021 | 329 | 9 | 2.74% | 0.994 | 0.919 | 0.860 | 0.886 | 0.910 | 0.910 | 0.936 | 0.960 |
| Menon 2022 | 485 | 35 | 7.22% | 0.923 | 0.767 | 0.412 | 0.337 | 0.501 | 0.328 | 0.322 | 0.371 |
| Moran 2020 | 1992 | 39 | 1.96% | 0.971 | 0.710 | 0.599 | 0.787 | 0.798 | 0.600 | 0.833 | 0.834 |
| Parodis 2023b | 1679 | 33 | 1.97% | 0.958 | 0.739 | 0.801 | 0.740 | 0.786 | 0.837 | 0.669 | 0.823 |
| Roberts 2021 | 883 | 3 | 0.34% | 0.993 | 0.966 | 0.917 | 0.938 | 0.938 | 0.967 | 0.988 | 0.988 |
| Seghers 2022 | 1759 | 16 | 0.91% | 0.986 | 0.886 | 0.737 | 0.878 | 0.882 | 0.787 | 0.928 | 0.932 |
| Theobald 2021 | 70 | 9 | 12.86% | 0.923 | 0.778 | 0.137 | 0.664 | 0.593 | 0.187 | 0.714 | 0.643 |
| Toffalini 2021 | 328 | 22 | 6.71% | 0.945 | 0.676 | 0.680 | 0.762 | 0.743 | 0.532 | 0.787 | 0.787 |
| Tumkaya 2018 | 481 | 39 | 8.11% | 0.925 | 0.665 | 0.674 | 0.609 | 0.673 | 0.681 | 0.586 | 0.640 |
| van der Waal 2022 | 1158 | 17 | 1.47% | 0.990 | 0.954 | 0.436 | 0.896 | 0.853 | 0.486 | 0.946 | 0.903 |
| Walker 2018 | 11,735 | 64 | 0.55% | 0.986 | 0.926 | 0.841 | 0.874 | 0.882 | 0.659 | 0.872 | 0.930 |

ASReview values are means of ten random starts; the Jev ranking is the mean of ten random orders of tied scores; the hybrid is a single run.

**Supplementary Table 54. Held-out SYNERGY+ reviews: stopping outcomes per review (the statistical criterion and the margins are post hoc)**

| Review | Read at $\tau=0.07$ | Recall at $\tau=0.07$ | Lowest score of a positive record | Positive records below $\tau$ | Largest $\tau$ with recall $\geq 0.95$ | K: read | K: recall | SC on ASReview ranking: read | SC on Jev ranking: read | SC on Jev ranking: recall |
| --- | --- | --- | --- | --- | --- | --- | --- | --- | --- | --- |
| Abgaz 2023 | 80.2% | 1.000 | 0.85 | 0 | 0.85 | 100.0% | 1.000 | 96.4% | 95.5% | 1.000 |
| Attai 2022 | 47.0% | 0.985 | 0.05 | 2 | 0.16 | 92.6% | 1.000 | 73.9% | 68.1% | 1.000 |
| Boersma-van Dam 2024 | 20.8% | 0.958 | 0.04 | 1 | 0.10 | 100.0% | 1.000 | 85.0% | 84.8% | 1.000 |
| Bos 2018 | 42.1% | 1.000 | 0.32 | 0 | 0.32 | 100.0% | 1.000 | 95.3% | 95.3% | 1.000 |
| Bosch 2021 | 52.5% | 1.000 | 0.55 | 0 | 0.66 | 97.1% | 1.000 | 83.5% | 80.0% | 1.000 |
| Chakraborty 2023 | 97.1% | 1.000 | 0.52 | 0 | 0.52 | 62.8% | 0.657 | 96.2% | 95.2% | 1.000 |
| Chueca 2023 | 86.9% | 1.000 | 0.25 | 0 | 0.25 | 100.0% | 1.000 | 99.0% | 96.9% | 1.000 |
| Donners 2021 | 71.4% | 1.000 | 0.44 | 0 | 0.44 | 100.0% | 1.000 | 96.6% | 96.6% | 1.000 |
| Eggmann 2023 | 53.8% | 1.000 | 0.42 | 0 | 0.42 | 100.0% | 1.000 | 95.6% | 95.5% | 1.000 |
| Hall 2011 | 12.9% | 1.000 | 0.67 | 0 | 0.67 | 77.7% | 1.000 | 95.3% | 95.3% | 1.000 |
| Harms 2024 | 92.9% | 1.000 | 0.50 | 0 | 0.54 | 100.0% | 1.000 | 92.4% | 71.1% | 1.000 |
| Kerschbaumer 2024b | 90.5% | 1.000 | 0.62 | 0 | 0.62 | 100.0% | 1.000 | 96.3% | 96.1% | 1.000 |
| Lewowski 2021 | 37.7% | 1.000 | 0.53 | 0 | 0.53 | 97.4% | 1.000 | 95.7% | 95.4% | 1.000 |
| Menon 2022 | 87.6% | 1.000 | 0.10 | 0 | 0.11 | 100.0% | 1.000 | 92.7% | 92.8% | 1.000 |
| Moran 2020 | 34.3% | 1.000 | 0.13 | 0 | 0.13 | 100.0% | 1.000 | 77.9% | 69.3% | 1.000 |
| Parodis 2023b | 85.1% | 1.000 | 0.23 | 0 | 0.33 | 89.1% | 1.000 | 81.3% | 85.0% | 1.000 |
| Roberts 2021 | 31.5% | 1.000 | 0.62 | 0 | 0.62 | 100.0% | 1.000 | 95.2% | 95.1% | 1.000 |
| Seghers 2022 | 54.3% | 1.000 | 0.36 | 0 | 0.36 | 100.0% | 1.000 | 96.1% | 95.4% | 1.000 |
| Theobald 2021 | 75.7% | 1.000 | 0.60 | 0 | 0.60 | 100.0% | 1.000 | 98.9% | 97.1% | 1.000 |
| Toffalini 2021 | 50.3% | 1.000 | 0.35 | 0 | 0.41 | 100.0% | 1.000 | 88.1% | 82.3% | 1.000 |
| Tumkaya 2018 | 51.8% | 1.000 | 0.09 | 0 | 0.12 | 100.0% | 1.000 | 74.9% | 78.4% | 1.000 |
| van der Waal 2022 | 64.2% | 1.000 | 0.44 | 0 | 0.44 | 91.3% | 0.906 | 97.6% | 95.3% | 1.000 |
| Walker 2018 | 33.5% | 1.000 | 0.16 | 0 | 0.25 | 53.6% | 0.902 | 68.8% | 58.8% | 1.000 |

K=knee method on the ASReview ranking. SC=statistical criterion. Values on rankings with random starts or random tie orders are means over ten runs.

**Supplementary Table 55. CLEF 2019 Cochrane reviews with at least one content-relevant record: ranking outcomes per review (the Jev ranking is post hoc)**

| Review | Records | Relevant records | Rate | AUC, Jev | AUC, bge-base | WSS@95, ASReview | WSS@95, Jev ranking | WSS@95, hybrid | WSS@100, ASReview | WSS@100, Jev ranking | WSS@100, hybrid |
| --- | --- | --- | --- | --- | --- | --- | --- | --- | --- | --- | --- |
| CD000996 | 281 | 6 | 2.14% | 0.999 | 0.964 | 0.913 | 0.922 | 0.925 | 0.963 | 0.972 | 0.975 |
| CD001261 | 571 | 22 | 3.85% | 0.984 | 0.845 | 0.852 | 0.848 | 0.887 | 0.896 | 0.864 | 0.935 |
| CD004414 | 336 | 7 | 2.08% | 0.978 | 0.966 | 0.832 | 0.893 | 0.908 | 0.882 | 0.943 | 0.958 |
| CD006468 | 3874 | 12 | 0.31% | 0.999 | 0.947 | 0.930 | 0.940 | 0.944 | 0.980 | 0.990 | 0.994 |
| CD007867 | 943 | 9 | 0.95% | 0.991 | 0.857 | 0.773 | 0.891 | 0.910 | 0.823 | 0.941 | 0.960 |
| CD008874 | 2382 | 110 | 4.62% | 0.964 | 0.922 | 0.814 | 0.779 | 0.811 | 0.615 | 0.746 | 0.779 |
| CD009044 | 3169 | 1 | 0.03% | 0.986 | 0.996 | 0.950 | 0.936 | 0.936 | 1.000 | 0.986 | 0.986 |
| CD009069 | 1757 | 5 | 0.28% | 0.981 | 0.879 | 0.790 | 0.907 | 0.889 | 0.840 | 0.957 | 0.939 |
| CD009642 | 1922 | 50 | 2.60% | 0.997 | 0.975 | 0.894 | 0.917 | 0.916 | 0.640 | 0.925 | 0.907 |
| CD010038 | 8867 | 9 | 0.10% | 0.999 | 0.957 | 0.847 | 0.945 | 0.945 | 0.897 | 0.995 | 0.995 |
| CD010239 | 224 | 10 | 4.46% | 0.999 | 0.983 | 0.897 | 0.892 | 0.901 | 0.947 | 0.942 | 0.951 |
| CD010558 | 2815 | 10 | 0.36% | 0.964 | 0.864 | 0.876 | 0.604 | 0.919 | 0.926 | 0.654 | 0.969 |
| CD010753 | 2539 | 20 | 0.79% | 0.976 | 0.857 | 0.746 | 0.856 | 0.898 | 0.769 | 0.891 | 0.928 |
| CD011571 | 146 | 6 | 4.11% | 0.996 | 0.970 | 0.842 | 0.895 | 0.895 | 0.892 | 0.945 | 0.945 |
| CD011686 | 9729 | 3 | 0.03% | 1.000 | 0.999 | 0.944 | 0.949 | 0.949 | 0.994 | 0.999 | 0.999 |
| CD011768 | 9160 | 25 | 0.27% | 0.997 | 0.986 | 0.913 | 0.939 | 0.937 | 0.937 | 0.984 | 0.981 |
| CD011787 | 4369 | 28 | 0.64% | 0.992 | 0.948 | 0.868 | 0.911 | 0.903 | 0.642 | 0.949 | 0.938 |
| CD011977 | 195 | 37 | 18.97% | 0.897 | 0.782 | 0.512 | 0.346 | 0.483 | 0.531 | 0.375 | 0.467 |
| CD012069 | 3479 | 245 | 7.04% | 0.900 | 0.668 | 0.478 | 0.534 | 0.556 | 0.130 | 0.283 | 0.366 |
| CD012080 | 6643 | 77 | 1.16% | 0.957 | 0.911 | 0.771 | 0.799 | 0.796 | 0.645 | 0.811 | 0.806 |
| CD012164 | 61 | 3 | 4.92% | 1.000 | 0.920 | 0.683 | 0.901 | 0.901 | 0.733 | 0.951 | 0.951 |
| CD012233 | 472 | 9 | 1.91% | 0.858 | 0.864 | 0.394 | 0.118 | 0.143 | 0.444 | 0.168 | 0.193 |
| CD012455 | 1593 | 5 | 0.31% | 1.000 | 0.998 | 0.929 | 0.946 | 0.946 | 0.979 | 0.996 | 0.996 |
| CD012551 | 591 | 29 | 4.91% | 0.983 | 0.753 | 0.705 | 0.847 | 0.852 | 0.718 | 0.881 | 0.871 |
| CD012567 | 6735 | 4 | 0.06% | 0.999 | 0.968 | 0.848 | 0.948 | 0.946 | 0.898 | 0.998 | 0.996 |
| CD012661 | 3367 | 94 | 2.79% | 0.970 | 0.860 | 0.799 | 0.790 | 0.824 | 0.386 | 0.716 | 0.652 |
| CD012669 | 1260 | 24 | 1.90% | 0.946 | 0.876 | 0.758 | 0.803 | 0.811 | 0.717 | 0.742 | 0.766 |
| CD012768 | 131 | 22 | 16.79% | 0.841 | 0.689 | 0.461 | 0.496 | 0.523 | 0.489 | 0.493 | 0.565 |

ASReview values are means of ten random starts; the Jev ranking is the mean of ten random orders of tied scores; the hybrid is a single run.

**Supplementary Table 56. CLEF 2019 Cochrane reviews with at least one content-relevant record: stopping outcomes per review (the statistical criterion and the margins are post hoc)**

| Review | Read at $\tau=0.07$ | Recall at $\tau=0.07$ | Lowest score of a positive record | Positive records below $\tau$ | Largest $\tau$ with recall $\geq 0.95$ | K: read | K: recall | SC on ASReview ranking: read | SC on Jev ranking: read | SC on Jev ranking: recall |
| --- | --- | --- | --- | --- | --- | --- | --- | --- | --- | --- |
| CD000996 | 26.0% | 1.000 | 0.70 | 0 | 0.70 | 94.8% | 1.000 | 95.4% | 95.4% | 1.000 |
| CD001261 | 53.9% | 1.000 | 0.71 | 0 | 0.81 | 74.0% | 1.000 | 80.0% | 80.7% | 1.000 |
| CD004414 | 25.0% | 1.000 | 0.65 | 0 | 0.65 | 100.0% | 1.000 | 95.7% | 95.5% | 1.000 |
| CD006468 | 39.9% | 1.000 | 0.48 | 0 | 0.48 | 25.5% | 1.000 | 95.1% | 95.1% | 1.000 |
| CD007867 | 46.8% | 1.000 | 0.26 | 0 | 0.26 | 100.0% | 1.000 | 95.9% | 95.3% | 1.000 |
| CD008874 | 43.2% | 1.000 | 0.11 | 0 | 0.18 | 51.8% | 1.000 | 62.6% | 54.7% | 1.000 |
| CD009044 | 13.9% | 1.000 | 0.30 | 0 | 0.30 | 4.9% | 1.000 | 95.0% | 95.1% | 1.000 |
| CD009069 | 27.4% | 1.000 | 0.77 | 0 | 0.77 | 100.0% | 1.000 | 95.8% | 95.2% | 1.000 |
| CD009642 | 10.6% | 1.000 | 0.09 | 0 | 0.55 | 24.1% | 0.980 | 76.4% | 65.9% | 1.000 |
| CD010038 | 13.5% | 1.000 | 0.47 | 0 | 0.47 | 90.2% | 0.911 | 95.5% | 95.0% | 1.000 |
| CD010239 | 22.3% | 1.000 | 0.64 | 0 | 0.64 | 82.5% | 1.000 | 95.5% | 95.5% | 1.000 |
| CD010558 | 30.4% | 0.900 | 0.06 | 1 | 0.06 | 90.4% | 1.000 | 95.4% | 96.7% | 1.000 |
| CD010753 | 46.0% | 1.000 | 0.20 | 0 | 0.22 | 100.0% | 1.000 | 82.8% | 80.1% | 1.000 |
| CD011571 | 19.2% | 1.000 | 0.67 | 0 | 0.67 | 100.0% | 1.000 | 95.9% | 95.9% | 1.000 |
| CD011686 | 7.2% | 1.000 | 0.51 | 0 | 0.51 | 32.5% | 1.000 | 95.1% | 95.0% | 1.000 |
| CD011768 | 21.2% | 1.000 | 0.47 | 0 | 0.61 | 44.7% | 1.000 | 79.1% | 78.0% | 1.000 |
| CD011787 | 47.7% | 1.000 | 0.55 | 0 | 0.63 | 77.8% | 1.000 | 85.7% | 78.8% | 1.000 |
| CD011977 | 45.6% | 0.919 | 0.03 | 3 | 0.03 | 100.0% | 1.000 | 88.2% | 91.6% | 1.000 |
| CD012069 | 77.7% | 1.000 | 0.09 | 0 | 0.29 | 59.3% | 0.980 | 66.5% | 76.3% | 1.000 |
| CD012080 | 54.2% | 1.000 | 0.28 | 0 | 0.38 | 94.6% | 1.000 | 64.6% | 55.5% | 1.000 |
| CD012164 | 62.3% | 1.000 | 0.93 | 0 | 0.93 | 100.0% | 1.000 | 97.2% | 96.7% | 1.000 |
| CD012233 | 74.8% | 0.889 | 0.05 | 1 | 0.05 | 100.0% | 1.000 | 97.9% | 98.3% | 1.000 |
| CD012455 | 4.5% | 1.000 | 0.70 | 0 | 0.70 | 66.6% | 1.000 | 95.1% | 95.0% | 1.000 |
| CD012551 | 58.0% | 1.000 | 0.47 | 0 | 0.55 | 100.0% | 1.000 | 84.0% | 80.3% | 1.000 |
| CD012567 | 14.8% | 1.000 | 0.60 | 0 | 0.60 | 41.4% | 0.550 | 95.5% | 95.0% | 1.000 |
| CD012661 | 42.8% | 1.000 | 0.10 | 0 | 0.17 | 71.8% | 0.999 | 55.5% | 60.7% | 1.000 |
| CD012669 | 72.1% | 1.000 | 0.38 | 0 | 0.70 | 100.0% | 1.000 | 84.0% | 83.4% | 1.000 |
| CD012768 | 92.4% | 1.000 | 0.57 | 0 | 0.69 | 100.0% | 1.000 | 89.2% | 89.3% | 1.000 |

K=knee method on the ASReview ranking. SC=statistical criterion. Values on rankings with random starts or random tie orders are means over ten runs.

#### Supplementary Note 21. Further results of the analysis of the weight $\lambda$ (post hoc)

This note holds results of the analysis of the weight $\lambda$ (analysis 3 of Supplementary Table 1) that Supplementary Notes 10 and 19 do not give. The primary weight $\lambda=3$ and its selection on development reviews are unchanged.

##### Supplementary Table 57. Paired differences between the hybrid at each weight $\lambda$ and the Jev ranking, ASReview, and the hybrid with $\lambda=3$ (post hoc)

| Collection (reviews) | Comparison | Metric | $\lambda=1$ | $\lambda=2$ | $\lambda=3$ (primary) | $\lambda=5$ | $\lambda=10$ |
| --- | --- | --- | --- | --- | --- | --- | --- |
| Development (73) | Hybrid minus Jev ranking | WSS@95 | -0.0067 (-0.0297 to 0.0149); 37 / 33 / 3; $P=0.88$ | 0.0281 (0.0133 to 0.0427); 45 / 26 / 2; $P=0.0017$ | 0.0286 (0.0171 to 0.0404); 49 / 21 / 3; $P<0.0001$ | 0.0258 (0.0166 to 0.0355); 54 / 17 / 2; $P<0.0001$ | 0.0162 (0.0098 to 0.0235); 52 / 17 / 4; $P<0.0001$ |
| Development (73) | Hybrid minus Jev ranking | WSS@100 | 0.0046 (-0.0194 to 0.0269); 39 / 31 / 3; $P=0.38$ | 0.0333 (0.0156 to 0.0512); 47 / 24 / 2; $P=0.0015$ | 0.0332 (0.0198 to 0.0476); 49 / 20 / 4; $P<0.0001$ | 0.0268 (0.0158 to 0.0387); 51 / 20 / 2; $P<0.0001$ | 0.0176 (0.0104 to 0.0258); 52 / 18 / 3; $P<0.0001$ |
| Development (73) | Hybrid minus Jev ranking | Recall at 10% read | -0.0120 (-0.0358 to 0.0112); 22 / 31 / 20; $P=0.32$ | 0.0224 (0.0029 to 0.0421); 29 / 17 / 27; $P=0.032$ | 0.0268 (0.0103 to 0.0446); 33 / 11 / 29; $P=0.0010$ | 0.0275 (0.0138 to 0.0433); 32 / 9 / 32; $P=0.00016$ | 0.0195 (0.0080 to 0.0334); 24 / 6 / 43; $P=0.00061$ |
| Development (73) | Hybrid minus Jev ranking | Recall at 20% read | -0.0067 (-0.0266 to 0.0137); 13 / 21 / 39; $P=0.30$ | 0.0156 (-0.0004 to 0.0336); 19 / 12 / 42; $P=0.11$ | 0.0173 (0.0027 to 0.0340); 19 / 9 / 45; $P=0.028$ | 0.0188 (0.0062 to 0.0343); 20 / 5 / 48; $P=0.0063$ | 0.0071 (0.0003 to 0.0149); 15 / 5 / 53; $P=0.050$ |
| Development (73) | Hybrid minus Jev ranking | Recall at 30% read | 0.0013 (−0.0121 to 0.0163); 12 / 14 / 47; $P=0.85$ | 0.0132 (0.0015 to 0.0283); 14 / 6 / 53; $P=0.052$ | 0.0085 (−0.0007 to 0.0212); 13 / 5 / 55; $P=0.12$ | 0.0094 (0.0010 to 0.0214); 14 / 3 / 56; $P=0.0099$ | 0.0093 (0.0023 to 0.0207); 12 / 2 / 59; $P=0.0029$ |
| Development (73) | Hybrid minus ASReview | WSS@95 | 0.1238 (0.0937 to 0.1577); 67 / 6 / 0; $P<0.0001$ | 0.1586 (0.1200 to 0.2014); 65 / 8 / 0; $P<0.0001$ | 0.1591 (0.1191 to 0.2039); 65 / 8 / 0; $P<0.0001$ | 0.1563 (0.1163 to 0.2019); 65 / 8 / 0; $P<0.0001$ | 0.1467 (0.1059 to 0.1924); 62 / 11 / 0; $P<0.0001$ |
| Development (73) | Hybrid minus ASReview | WSS@100 | 0.1398 (0.1068 to 0.1767); 66 / 7 / 0; $P<0.0001$ | 0.1684 (0.1269 to 0.2149); 64 / 9 / 0; $P<0.0001$ | 0.1684 (0.1250 to 0.2168); 61 / 12 / 0; $P<0.0001$ | 0.1620 (0.1180 to 0.2111); 61 / 12 / 0; $P<0.0001$ | 0.1528 (0.1090 to 0.2028); 57 / 16 / 0; $P<0.0001$ |
| Development (73) | Hybrid minus ASReview | Recall at 10% read | 0.1265 (0.0962 to 0.1597); 59 / 5 / 9; $P<0.0001$ | 0.1608 (0.1287 to 0.1956); 60 / 5 / 8; $P<0.0001$ | 0.1652 (0.1311 to 0.2022); 59 / 6 / 8; $P<0.0001$ | 0.1660 (0.1315 to 0.2025); 59 / 6 / 8; $P<0.0001$ | 0.1579 (0.1226 to 0.1947); 55 / 10 / 8; $P<0.0001$ |
| Development (73) | Hybrid minus ASReview | Recall at 20% read | 0.0806 (0.0574 to 0.1072); 48 / 3 / 22; $P<0.0001$ | 0.1029 (0.0746 to 0.1344); 51 / 2 / 20; $P<0.0001$ | 0.1046 (0.0758 to 0.1368); 51 / 3 / 19; $P<0.0001$ | 0.1061 (0.0765 to 0.1395); 50 / 4 / 19; $P<0.0001$ | 0.0944 (0.0634 to 0.1284); 48 / 7 / 18; $P<0.0001$ |
| Development (73) | Hybrid minus ASReview | Recall at 30% read | 0.0685 (0.0433 to 0.0971); 35 / 3 / 35; $P<0.0001$ | 0.0804 (0.0519 to 0.1138); 40 / 3 / 30; $P<0.0001$ | 0.0757 (0.0445 to 0.1107); 38 / 5 / 30; $P<0.0001$ | 0.0766 (0.0451 to 0.1120); 37 / 5 / 31; $P<0.0001$ | 0.0764 (0.0446 to 0.1120); 37 / 5 / 31; $P<0.0001$ |
| Development (73) | Hybrid minus ASReview | Proportion read, knee method | −0.0363 (−0.0637 to −0.0116); 5 / 19 / 49; $P=0.0056$ | −0.0581 (−0.0893 to −0.0284); 6 / 24 / 43; $P=0.00039$ | −0.0629 (−0.1015 to −0.0271); 8 / 23 / 42; $P=0.0018$ | −0.0626 (−0.1028 to −0.0239); 8 / 22 / 43; $P=0.0030$ | −0.0666 (−0.1065 to −0.0294); 9 / 22 / 42; $P=0.0021$ |
| Development (73) | Hybrid minus ASReview | Recall at the stop, knee method | 0.0072 (0.0000 to 0.0168); 4 / 1 / 68; $P=0.080$ | 0.0073 (0.0001 to 0.0168); 4 / 1 / 68; $P=0.080$ | 0.0075 (0.0003 to 0.0170); 4 / 1 / 68; $P=0.080$ | 0.0075 (0.0003 to 0.0170); 4 / 1 / 68; $P=0.080$ | 0.0076 (0.0004 to 0.0170); 4 / 1 / 68; $P=0.080$ |
| Development (73) | Hybrid minus hybrid with $\lambda=3$ | WSS@95 | −0.0353 (−0.0542 to −0.0190); 15 / 51 / 7; $P<0.0001$ | −0.0005 (−0.0064 to 0.0053); 26 / 34 / 13; $P=0.62$ | reference | −0.0028 (−0.0071 to −0.0012); 26 / 32 / 15; $P=0.39$ | −0.0124 (−0.0192 to −0.0059); 20 / 44 / 9; $P=0.00069$ |
| Development (73) | Hybrid minus hybrid with $\lambda=3$ | WSS@100 | −0.0285 (−0.0454 to −0.0130); 21 / 45 / 7; $P=0.0027$ | 0.0001 (−0.0060 to 0.0061); 25 / 34 / 14; $P=0.82$ | reference | −0.0064 (−0.0107 to −0.0024); 21 / 37 / 15; $P=0.017$ | −0.0156 (−0.0237 to −0.0081); 20 / 43 / 10; $P=0.00073$ |
| Development (73) | Hybrid minus hybrid with $\lambda=3$ | Recall at 10% read | −0.0388 (−0.0546 to −0.0249); 6 / 37 / 30; $P<0.0001$ | −0.0044 (−0.0137 to 0.0035); 10 / 11 / 52; $P=0.59$ | reference | 0.0007 (−0.0065 to 0.0082); 12 / 11 / 50; $P=0.87$ | −0.0073 (−0.0185 to 0.0034); 11 / 20 / 42; $P=0.20$ |
| Development (73) | Hybrid minus hybrid with $\lambda=3$ | Recall at 20% read | −0.0241 (−0.0386 to −0.0104); 5 / 28 / 40; $P=0.00069$ | −0.0018 (−0.0072 to 0.0031); 8 / 8 / 57; $P=0.53$ | reference | 0.0014 (−0.0052 to 0.0087); 10 / 7 / 56; $P=0.69$ | −0.0102 (−0.0239 to 0.0012); 6 / 12 / 55; $P=0.11$ |
| Development (73) | Hybrid minus hybrid with $\lambda=3$ | Recall at 30% read | −0.0071 (−0.0179 to 0.0035); 5 / 15 / 53; $P=0.10$ | 0.0047 (−0.0012 to 0.0144); 10 / 4 / 59; $P=0.33$ | reference | 0.0009 (−0.0022 to 0.0042); 5 / 4 / 64; $P=0.37$ | 0.0008 (−0.0044 to 0.0058); 6 / 6 / 61; $P=0.69$ |
| Development (73) | Hybrid minus hybrid with $\lambda=3$ | Proportion read, knee method | 0.0266 (−0.0020 to 0.0578); 18 / 9 / 46; $P=0.11$ | 0.0048 (−0.0117 to 0.0223); 14 / 10 / 49; $P=0.65$ | reference | 0.0002 (−0.0088 to 0.0103); 12 / 12 / 49; $P=0.67$ | −0.0038 (−0.0246 to 0.0150); 16 / 12 / 45; $P=0.98$ |
| Development (73) | Hybrid minus hybrid with $\lambda=3$ | Recall at the stop, knee method | −0.0003 (−0.0008 to 0.0000); 0 / 1 / 72; $P=0.32$ | −0.0002 (−0.0006 to 0.0000); 0 / 1 / 72; $P=0.32$ | reference | 0.0000 (0.0000 to 0.0000); 0 / 0 / 73; no test | 0.0001 (0.0000 to 0.0002); 1 / 0 / 72; $P=0.32$ |
| Development (73) | Jev ranking minus ASReview (one value; no weight) | WSS@95 | 0.1305 (0.0872 to 0.1786); 60 / 13 / 0; $P<0.0001$ | .. | .. | .. | .. |
| Development (73) | Jev ranking minus ASReview (one value; no weight) | WSS@100 | 0.1352 (0.0891 to 0.1874); 53 / 20 / 0; $P<0.0001$ | .. | .. | .. | .. |
| Development (73) | Jev ranking minus ASReview (one value; no weight) | Recall at 10% read | 0.1384 (0.0984 to 0.1792); 53 / 12 / 8; $P<0.0001$ | .. | .. | .. | .. |
| Development (73) | Jev ranking minus ASReview (one value; no weight) | Recall at 20% read | 0.0873 (0.0534 to 0.1245); 45 / 11 / 17; $P<0.0001$ | .. | .. | .. | .. |
| Development (73) | Jev ranking minus ASReview (one value; no weight) | Recall at 30% read | 0.0672 (0.0344 to 0.1038); 36 / 9 / 28; $P<0.0001$ | .. | .. | .. | .. |
| Held-out (23) | Hybrid minus Jev ranking | WSS@95 | not run | 0.0187 (−0.0049 to 0.0480); 15 / 6 / 2; $P=0.099$ | 0.0187 (0.0025 to 0.0376); 16 / 4 / 3; $P=0.025$ | 0.0132 (0.0039 to 0.0221); 16 / 4 / 3; $P=0.019$ | 0.0083 (0.0032 to 0.0136); 17 / 4 / 2; $P=0.0057$ |
| Held-out (23) | Hybrid minus Jev ranking | WSS@100 | not run | 0.0162 (−0.0065 to 0.0405); 13 / 8 / 2; $P=0.20$ | 0.0207 (0.0028 to 0.0399); 14 / 5 / 4; $P=0.033$ | 0.0180 (0.0056 to 0.0310); 15 / 5 / 3; $P=0.017$ | 0.0136 (0.0057 to 0.0224); 16 / 5 / 2; $P=0.0057$ |
| Held-out (23) | Hybrid minus Jev ranking | Recall at 10% read | not run | 0.0111 (−0.0082 to 0.0317); 7 / 4 / 12; $P=0.25$ | 0.0163 (0.0004 to 0.0334); 8 / 1 / 14; $P=0.051$ | 0.0160 (0.0027 to 0.0310); 7 / 1 / 15; $P=0.050$ | 0.0109 (0.0017 to 0.0206); 8 / 1 / 14; $P=0.051$ |
| Held-out (23) | Hybrid minus Jev ranking | Recall at 20% read | not run | 0.0143 (0.0015 to 0.0299); 7 / 1 / 15; $P=0.050$ | 0.0148 (0.0022 to 0.0295); 7 / 1 / 15; $P=0.036$ | 0.0071 (−0.0078 to 0.0212); 6 / 1 / 16; $P=0.24$ | 0.0071 (−0.0072 to 0.0200); 7 / 1 / 15; $P=0.16$ |
| Held-out (23) | Hybrid minus Jev ranking | Recall at 30% read | not run | −0.0001 (−0.0125 to 0.0087); 4 / 1 / 18; $P=0.50$ | 0.0006 (−0.0119 to 0.0095); 4 / 1 / 18; $P=0.50$ | −0.0006 (−0.0130 to 0.0081); 4 / 1 / 18; $P=0.50$ | 0.0040 (0.0002 to 0.0083); 4 / 0 / 19; $P=0.068$ |
| Held-out (23) | Hybrid minus ASReview | WSS@95 | not run | 0.1468 (0.0926 to 0.2061); 23 / 0 / 0; $P<0.0001$ | 0.1468 (0.0893 to 0.2091); 21 / 2 / 0; $P<0.0001$ | 0.1413 (0.0801 to 0.2083); 20 / 3 / 0; $P<0.0001$ | 0.1364 (0.0720 to 0.2061); 20 / 3 / 0; $P=0.00025$ |
| Held-out (23) | Hybrid minus ASReview | WSS@100 | not run | 0.1778 (0.1151 to 0.2476); 23 / 0 / 0; $P<0.0001$ | 0.1822 (0.1155 to 0.2554); 21 / 2 / 0; $P<0.0001$ | 0.1795 (0.1087 to 0.2566); 21 / 2 / 0; $P<0.0001$ | 0.1751 (0.1012 to 0.2547); 21 / 2 / 0; $P<0.0001$ |
| Held-out (23) | Hybrid minus ASReview | Recall at 10% read | not run | 0.1753 (0.1134 to 0.2407); 20 / 0 / 3; $P<0.0001$ | 0.1805 (0.1176 to 0.2485); 20 / 0 / 3; $P<0.0001$ | 0.1802 (0.1151 to 0.2494); 19 / 1 / 3; $P=0.00014$ | 0.1751 (0.1082 to 0.2455); 18 / 2 / 3; $P=0.00025$ |
| Held-out (23) | Hybrid minus ASReview | Recall at 20% read | not run | 0.1200 (0.0708 to 0.1747); 17 / 0 / 6; $P=0.00029$ | 0.1204 (0.0713 to 0.1756); 17 / 0 / 6; $P=0.00029$ | 0.1128 (0.0669 to 0.1646); 17 / 1 / 5; $P=0.00028$ | 0.1128 (0.0662 to 0.1650); 17 / 1 / 5; $P=0.00028$ |
| Held-out (23) | Hybrid minus ASReview | Recall at 30% read | not run | 0.0579 (0.0155 to 0.1087); 10 / 1 / 12; $P=0.0076$ | 0.0585 (0.0163 to 0.1088); 10 / 1 / 12; $P=0.0076$ | 0.0574 (0.0150 to 0.1075); 9 / 2 / 12; $P=0.013$ | 0.0619 (0.0148 to 0.1197); 9 / 2 / 12; $P=0.013$ |
| Held-out (23) | Hybrid minus ASReview | Proportion read, knee method | not run | -0.0831 (-0.1524 to -0.0172); 3 / 9 / 11; $P=0.028$ | -0.1025 (-0.1660 to -0.0420); 2 / 11 / 10; $P=0.0071$ | -0.1003 (-0.1623 to -0.0404); 2 / 11 / 10; $P=0.0071$ | -0.1074 (-0.1776 to -0.0394); 2 / 12 / 9; $P=0.013$ |
| Held-out (23) | Hybrid minus ASReview | Recall at the stop, knee method | not run | 0.0233 (0.0000 to 0.0596); 3 / 0 / 20; $P=0.11$ | 0.0233 (0.0000 to 0.0596); 3 / 0 / 20; $P=0.11$ | 0.0233 (0.0000 to 0.0596); 3 / 0 / 20; $P=0.11$ | 0.0233 (0.0000 to 0.0596); 3 / 0 / 20; $P=0.11$ |
| Held-out (23) | Hybrid minus hybrid with $\lambda=3$ | WSS@95 | not run | 0.0000 (-0.0096 to 0.0121); 8 / 12 / 3; $P=0.28$ | reference | -0.0055 (-0.0168 to 0.0027); 7 / 12 / 4; $P=0.42$ | -0.0104 (-0.0281 to 0.0031); 6 / 14 / 3; $P=0.13$ |
| Held-out (23) | Hybrid minus hybrid with $\lambda=3$ | WSS@100 | not run | -0.0044 (-0.0118 to 0.0032); 6 / 14 / 3; $P=0.14$ | reference | -0.0026 (-0.0099 to 0.0037); 9 / 10 / 4; $P=0.75$ | -0.0071 (-0.0201 to 0.0045); 9 / 12 / 2; $P=0.29$ |
| Held-out (23) | Hybrid minus hybrid with $\lambda=3$ | Recall at 10% read | not run | -0.0052 (-0.0143 to 0.0033); 1 / 4 / 18; $P=0.22$ | reference | -0.0003 (-0.0097 to 0.0085); 2 / 2 / 19; $P=1.00$ | -0.0054 (-0.0176 to 0.0046); 3 / 4 / 16; $P=0.35$ |
| Held-out (23) | Hybrid minus hybrid with $\lambda=3$ | Recall at 20% read | not run | -0.0005 (-0.0033 to 0.0019); 1 / 1 / 21; $P=0.65$ | reference | -0.0077 (-0.0200 to 0.0020); 1 / 4 / 18; $P=0.22$ | -0.0076 (-0.0223 to 0.0060); 1 / 4 / 18; $P=0.35$ |
| Held-out (23) | Hybrid minus hybrid with $\lambda=3$ | Recall at 30% read | not run | -0.0006 (-0.0019 to 0.0000); 0 / 1 / 22; $P=0.32$ | reference | -0.0011 (-0.0033 to 0.0000); 0 / 1 / 22; $P=0.32$ | 0.0034 (-0.0037 to 0.0142); 1 / 2 / 20; $P=1.00$ |
| Held-out (23) | Hybrid minus hybrid with $\lambda=3$ | Proportion read, knee method | not run | 0.0194 (-0.0068 to 0.0480); 6 / 5 / 12; $P=0.25$ | reference | 0.0022 (-0.0288 to 0.0355); 4 / 4 / 15; $P=1.00$ | -0.0049 (-0.0510 to 0.0396); 5 / 5 / 13; $P=0.88$ |
| Held-out (23) | Hybrid minus hybrid with $\lambda=3$ | Recall at the stop, knee method | not run | 0.0000 (0.0000 to 0.0000); 0 / 0 / 23; no test | reference | 0.0000 (0.0000 to 0.0000); 0 / 0 / 23; no test | 0.0000 (0.0000 to 0.0000); 0 / 0 / 23; no test |
| Held-out (23) | Jev ranking minus ASReview (one value; no weight) | WSS@95 | 0.1281 (0.0633 to 0.1988); 20 / 3 / 0; $P=0.00048$ | .. | .. | .. | .. |
| Held-out (23) | Jev ranking minus ASReview (one value; no weight) | WSS@100 | 0.1615 (0.0838 to 0.2443); 20 / 3 / 0; $P=0.00025$ | .. | .. | .. | .. |
| Held-out (23) | Jev ranking minus ASReview (one value; no weight) | Recall at 10% read | 0.1642 (0.0971 to 0.2341); 18 / 2 / 3; $P=0.00039$ | .. | .. | .. | .. |
| Held-out (23) | Jev ranking minus ASReview (one value; no weight) | Recall at 20% read | 0.1057 (0.0538 to 0.1635); 16 / 1 / 6; $P=0.00085$ | .. | .. | .. | .. |
| Held-out (23) | Jev ranking minus ASReview (one value; no weight) | Recall at 30% read | 0.0580 (0.0106 to 0.1165); 9 / 3 / 11; $P=0.019$ | .. | .. | .. | .. |
| CLEF 2019 (28) | Hybrid minus Jev ranking | WSS@95 | not run | not run | 0.0249 (0.0068 to 0.0516); 17 / 9 / 2; $P=0.0043$ | 0.0197 (0.0053 to 0.0411); 19 / 7 / 2; $P=0.0020$ | 0.0108 (0.0026 to 0.0240); 17 / 7 / 4; $P=0.0056$ |
| CLEF 2019 (28) | Hybrid minus Jev ranking | WSS@100 | not run | not run | 0.0239 (0.0040 to 0.0515); 15 / 11 / 2; $P=0.058$ | 0.0203 (0.0050 to 0.0421); 17 / 9 / 2; $P=0.012$ | 0.0143 (0.0042 to 0.0282); 17 / 7 / 4; $P=0.0051$ |
| CLEF 2019 (28) | Hybrid minus Jev ranking | Recall at 10% read | not run | not run | 0.0167 (0.0064 to 0.0288); 10 / 1 / 17; $P=0.0099$ | 0.0142 (0.0043 to 0.0258); 9 / 1 / 18; $P=0.017$ | 0.0054 (-0.0007 to 0.0124); 5 / 2 / 21; $P=0.13$ |
| CLEF 2019 (28) | Hybrid minus Jev ranking | Recall at 20% read | not run | not run | 0.0137 (0.0024 to 0.0275); 6 / 1 / 21; $P=0.063$ | 0.0125 (0.0017 to 0.0257); 6 / 1 / 21; $P=0.063$ | 0.0057 (-0.0007 to 0.0148); 5 / 1 / 22; $P=0.17$ |
| CLEF 2019 (28) | Hybrid minus Jev ranking | Recall at 30% read | not run | not run | 0.0073 (-0.0026 to 0.0198); 3 / 2 / 23; $P=0.22$ | 0.0068 (-0.0027 to 0.0193); 3 / 2 / 23; $P=0.35$ | 0.0062 (-0.0032 to 0.0187); 3 / 2 / 23; $P=0.35$ |
| CLEF 2019 (28) | Hybrid minus ASReview | WSS@95 | not run | not run | 0.0440 (0.0121 to 0.0712); 24 / 4 / 0; $P=0.00015$ | 0.0388 (0.0057 to 0.0671); 23 / 5 / 0; $P=0.0011$ | 0.0299 (-0.0070 to 0.0616); 22 / 6 / 0; $P=0.010$ |
| CLEF 2019 (28) | Hybrid minus ASReview | WSS@100 | not run | not run | 0.0877 (0.0447 to 0.1288); 25 / 3 / 0; $P<0.0001$ | 0.0841 (0.0380 to 0.1277); 24 / 4 / 0; $P=0.00034$ | 0.0781 (0.0298 to 0.1243); 23 / 5 / 0; $P=0.00097$ |
| CLEF 2019 (28) | Hybrid minus ASReview | Recall at 10% read | not run | not run | 0.0929 (0.0519 to 0.1439); 18 / 2 / 8; $P=0.00014$ | 0.0904 (0.0496 to 0.1420); 18 / 2 / 8; $P=0.00016$ | 0.0816 (0.0388 to 0.1341); 18 / 3 / 7; $P=0.00084$ |
| CLEF 2019 (28) | Hybrid minus ASReview | Recall at 20% read | not run | not run | 0.0502 (0.0185 to 0.0916); 12 / 0 / 16; $P=0.0022$ | 0.0490 (0.0176 to 0.0901); 12 / 1 / 15; $P=0.0030$ | 0.0422 (0.0095 to 0.0835); 11 / 2 / 15; $P=0.016$ |
| CLEF 2019 (28) | Hybrid minus ASReview | Recall at 30% read | not run | not run | 0.0170 (0.0049 to 0.0342); 8 / 0 / 20; $P=0.012$ | 0.0165 (0.0047 to 0.0334); 8 / 0 / 20; $P=0.012$ | 0.0159 (0.0043 to 0.0325); 8 / 0 / 20; $P=0.012$ |
| CLEF 2019 (28) | Hybrid minus ASReview | Proportion read, knee method | not run | not run | -0.1618 (-0.2696 to -0.0384); 1 / 20 / 7; $P=0.0010$ | -0.1696 (-0.2795 to -0.0429); 3 / 17 / 8; $P=0.0022$ | -0.1545 (-0.2686 to -0.0276); 4 / 16 / 8; $P=0.0045$ |
| CLEF 2019 (28) | Hybrid minus ASReview | Recall at the stop, knee method | not run | not run | 0.0187 (-0.0019 to 0.0547); 4 / 1 / 23; $P=0.22$ | 0.0185 (-0.0022 to 0.0546); 4 / 1 / 23; $P=0.22$ | 0.0154 (-0.0082 to 0.0532); 4 / 2 / 22; $P=0.60$ |
| CLEF 2019 (28) | Hybrid minus hybrid with $\lambda=3$ | WSS@95 | not run | not run | reference | -0.0052 (-0.0107 to -0.0012); 6 / 14 / 8; $P=0.030$ | -0.0142 (-0.0285 to -0.0032); 7 / 14 / 7; $P=0.016$ |
| CLEF 2019 (28) | Hybrid minus hybrid with $\lambda=3$ | WSS@100 | not run | not run | reference | -0.0036 (-0.0100 to 0.0018); 10 / 11 / 7; $P=0.59$ | -0.0096 (-0.0235 to 0.0013); 9 / 12 / 7; $P=0.23$ |
| CLEF 2019 (28) | Hybrid minus hybrid with $\lambda=3$ | Recall at 10% read | not run | not run | reference | -0.0025 (-0.0067 to 0.0006); 1 / 4 / 23; $P=0.22$ | -0.0113 (-0.0212 to -0.0034); 1 / 8 / 19; $P=0.015$ |
| CLEF 2019 (28) | Hybrid minus hybrid with $\lambda=3$ | Recall at 20% read | not run | not run | reference | -0.0013 (-0.0034 to 0.0000); 0 / 2 / 26; $P=0.18$ | -0.0080 (-0.0179 to -0.0010); 0 / 5 / 23; $P=0.043$ |
| CLEF 2019 (28) | Hybrid minus hybrid with $\lambda=3$ | Recall at 30% read | not run | not run | reference | -0.0004 (-0.0013 to 0.0000); 0 / 1 / 27; $P=0.32$ | -0.0010 (-0.0031 to 0.0000); 0 / 1 / 27; $P=0.32$ |
| CLEF 2019 (28) | Hybrid minus hybrid with $\lambda=3$ | Proportion read, knee method | not run | not run | reference | -0.0078 (-0.0417 to 0.0215); 8 / 7 / 13; $P=0.91$ | 0.0073 (-0.0355 to 0.0489); 10 / 6 / 12; $P=0.57$ |
| CLEF 2019 (28) | Hybrid minus hybrid with $\lambda=3$ | Recall at the stop, knee method | not run | not run | reference | -0.0001 (-0.0004 to 0.0000); 0 / 1 / 27; $P=0.32$ | -0.0033 (-0.0107 to 0.0009); 1 / 1 / 26; $P=0.65$ |
| CLEF 2019 (28) | Jev ranking minus ASReview (one value; no weight) | WSS@95 | 0.0191 (-0.0230 to 0.0563); 20 / 8 / 0; $P=0.032$ | .. | .. | .. | .. |
| CLEF 2019 (28) | Jev ranking minus ASReview (one value; no weight) | WSS@100 | 0.0638 (0.0105 to 0.1147); 22 / 6 / 0; $P=0.0071$ | .. | .. | .. | .. |
| CLEF 2019 (28) | Jev ranking minus ASReview (one value; no weight) | Recall at 10% read | 0.0762 (0.0333 to 0.1295); 17 / 4 / 7; $P=0.0026$ | .. | .. | .. | .. |
| CLEF 2019 (28) | Jev ranking minus ASReview (one value; no weight) | Recall at 20% read | 0.0365 (0.0031 to 0.0793); 11 / 4 / 13; $P=0.074$ | .. | .. | .. | .. |
| CLEF 2019 (28) | Jev ranking minus ASReview (one value; no weight) | Recall at 30% read | 0.0097 (-0.0091 to 0.0310); 9 / 2 / 17; $P=0.15$ | .. | .. | .. | .. |

Each cell gives the mean difference over reviews (95% CI); the numbers of reviews in which the first method was higher, lower, and tied; and the $P$ value of the two-sided Wilcoxon signed-rank test (SciPy default settings). The numbers of reviews and the tests were computed from exact differences (Supplementary Note 19). CIs are percentile bootstrap intervals (5000 resamples of reviews, seed 1003), so they differ in the third or fourth decimal from the intervals of Supplementary Table 19 (seed 1002) and of criterion C3. The table gives every comparison that the analysis of weights computed. $\lambda=1$ was run on development reviews only, and $\lambda=2$ on development and held-out reviews. The values $\lambda=5$ and $\lambda=10$ were added post hoc; the tests are descriptive, and no adjustment for multiplicity was made. Not run=no simulation with this weight in this collection.

On held-out reviews, the differences between the hybrid with $\lambda=2$, $\lambda=5$, or $\lambda=10$ and the hybrid with $\lambda=3$ had CIs that included zero for WSS@95, and the increment of the hybrid over the Jev ranking had a CI that included zero at $\lambda=2$. In CLEF reviews, the CI of the difference in WSS@95 between the hybrid with $\lambda=10$ and ASReview included zero (0.030, -0.007 to 0.062).

#### Supplementary Note 22. Coding of prior studies and verification of references

The 32 records of the subset search (Methods, Literature search) were coded from their abstracts with the AI coding tool used for the analyses (Methods, Provenance of model inputs and judgments)

on 26 September 2026 (post hoc; analysis 20 of Supplementary Table 1). The coding is not used in the main article. The checks of the references reported in this note were made with the same tool.

**Supplementary Table 58. Characteristics of the 32 records of the subset search (coded post hoc from abstracts)**

| Characteristic | Records |
| --- | --- |
| Primary evaluation of screening performance | 10 of 32 |
| Applied case study without a labeled evaluation | 4 of 32 |
| Protocol for an evaluation | 1 of 32 |
| Topic review or unrelated protocol | 17 of 32 |
| Among the ten evaluations: retrospective simulation on completed reviews | 9 |
| Among the ten evaluations: labeled sample within one live review | 1 |
| Among the ten evaluations: held-out or independent set of reviews stated | 3 (one further study used the test set of a public benchmark; two stated none; four did not state) |
| Among the ten evaluations: prespecified analysis stated | 1 |
| Among the ten evaluations: cost reported in the abstract | 0 |
| Among the ten evaluations: generative LLM as a zero-shot screener | 5 |

On 26 September 2026 (post hoc; analyses 17 and 18 of Supplementary Table 1), all 23 references then cited were found in Crossref, PubMed, arXiv, medRxiv, DataCite, or on the page of the publisher, and the responses were saved. Reference 15: the title cited is that of version 2 of the preprint (posted 10 May 2026); version 1 (posted 26 August 2025) had a different title; the citation gives both dates. Reference 18: the journal article exists (published online 21 September 2026), and its abstract reports a median AUC of 0.974 across 3434 internal test reviews and 0.976 on 22 external reviews. The SYNERGY+ release is cited by its web address in the Methods (Data sources). Reference 26 was added on 27 September 2026; its arXiv version (2106.09871, version 1 of 18 June 2021) and the Crossref record of the published version (doi 10.1145/3469096.3469873) were retrieved, and the passages cited in the Methods (Stopping rules) were read in the full text; the passages of ref. 11 quoted in Supplementary Note 11 were read in its full text in PubMed Central (analysis 25 of Supplementary Table 2). The vendor announcement is cited by its web address in the Introduction and the Methods, and its archived copy is described in Supplementary Note 1. The three references on McNemar-type methods (35, 36, and 37) were retrieved in PubMed and Crossref in analysis 25. On 28 September 2026 (post hoc; analysis 31 of Supplementary Table 2), references 26 and 18 were checked again in Crossref and PubMed and through the DOI resolver. For reference 26, Crossref records the DOI 10.1145/3469096.3469873 and pages 1–10, and no article number; the page of the ACM Digital Library could not be read (HTTP 403), and the reference format printed in the arXiv version gives no article number. Reference 18 has six authors in PubMed and Crossref, so the reference list gives the first author followed by et al., as the journal style requires for more than five authors; it has the article number 112514 and no volume. Reference 32 is cited in its published form: on 28 September 2026, Crossref recorded the DOI

10.1145/3626772.3657878, the six authors, the title, the Proceedings of the 47th International ACM SIGIR Conference on Research and Development in Information Retrieval, and pages 641–649, and the arXiv record of the preprint (2309.07597) names SIGIR 2024 in its comment. On 28 September 2026 (post hoc; analysis 45 of Supplementary Table 3), nine further references were checked: refs 28, 17, 9, 10, 7, 25, 33, and 39 were found in Crossref, PubMed, arXiv, or the ACL Anthology, and the use for which each was to be cited was compared with its abstract or full text; ref. 27 was checked against its abstract only, which does not state whether the reference standard was title and abstract relevance or final inclusion. For ref. 18, the numbers on which the Introduction draws (training on 19,787 completed reviews, a median AUC of 0.974 across 3434 internal test reviews, and 99.8% of included studies retained in 22 external Cochrane reviews at a threshold fixed on the internal validation split) were found in its PubMed abstract (PMID 42767575); its full text could not be retrieved, so the set of reviews and the pooling behind the 99.8% could not be confirmed. The PubMed search for a published per-record time of human title and abstract screening used the string ("abstract screening"[tiab] OR "title and abstract screening"[tiab] OR "citation screening"[tiab] OR "screening of titles and abstracts"[tiab]) AND ("per abstract"[tiab] OR "per citation"[tiab] OR "per record"[tiab] OR "per reference"[tiab] OR "per article"[tiab] OR seconds[tiab]). PubMed dropped three phrases that are not in its phrase index ("screening of titles and abstracts", "per abstract", and "per citation") and returned nine records at 20:03:57 on 28 September 2026, none of which reported a measured per-record time of human title and abstract screening in its abstract. The search was narrow, and its result is not evidence that no such estimate exists. On 29 September 2026 (post hoc; analysis 50 of Supplementary Table 3), three further sources were checked. Ref. 24 was found in Crossref and PubMed, and its full text in PubMed Central states in the Background that "An experienced reviewer can screen an average of two abstracts per minute", without a citation, and that abstracts on difficult topics may take several minutes each; the rate is used only for the illustrative conversion in the Results and Supplementary Note 2. Ref. 34 was found in Crossref; its full text could not be retrieved from the page of the publisher, three institutional repositories, or the addresses listed by two bibliographic indexes, and its abstract, read in the repository of TU Wien, states that normalized WSS is equivalent to the true negative rate and gives benchmark scores with the $TNR@95\%$ recall measure; the detailed definition, including the rounding of the 95% recall point, could not be compared with those of the Methods (Outcomes) and Supplementary Note 19. Ref. 34 is a different article from ref. 41. Ref. 17 was read again in the saved full text of its arXiv version: the review title was the query, and the fine-tuned rankers were trained on the training topics of the CLEF TAR 2017, 2018, and 2019 collections and applied to their test topics, which supports the wording of the Introduction. On 29 September 2026 (post hoc; analysis 51 of Supplementary Table 3), the journal pre-proof of ref. 34 (26 pages; not the version of record) was obtained from the UCL Discovery repository at the address of its PDF file, which had returned HTTP 404 in analysis 50 for a reason that is not known; the record page of the repository returned HTTP 403. The title, authors, DOI, and article number of the pre-proof agree with the Crossref record. The comparison with the Methods (Outcomes) and Supplementary Note 19 found three agreements. First, the article sets the number of false negatives at $r\%$ recall to $\lfloor ||I| \cdot (1-r) \rfloor$ (Section 3.3), so that the recall point is the rank of the $[0.95n]$-th included record, as in the Methods ($n - [0.05n] = [0.95n]$ was checked exactly for $n$ from 1 to 100,000). Second, it defines normalized WSS at 95% recall as $TN@95\%/|E|$, the true negative rate at 95% recall (Equations 9 to 11), which is the $TNR@95$ defined in Supplementary Note 19 and supports the sentences that cite the article. Third, it states that $WSS@95\%$ equals $WSS@100\%$ minus 0.05 when fewer than 20 included records are evaluated (Section 3.5, Equation 6), in agreement with the statement that 95% recall means all included records in reviews with at most 19 ($[0.95n]=n$ for every $n$ from 1 to 19 and for no $n$ from 20 to 1000). Section 4.1 and a sentence of Section 3.3 ("Consequently, for a fixed level of recall, true positives (TP) are equal to $r \cdot |I|$") write the recall point and the true positives without the rounding (for example, $TP=r \cdot |I|$), which is exact only when 5% of the included records is a whole number (not for 950 of the values of $n$ from 1 to 1000); this is an inconsistency within the article and not a difference from the Methods. The pre-proof may differ from the version of record, which was not obtained.

Verification of sources (analysis 50). Two articles proposed as references and the description of one cited article were checked against their text in Crossref, PubMed, PubMed Central, three institutional repositories (text, an abstract, was obtained from one), the page of the publisher, two bibliographic indexes, and the saved full text of the arXiv version; only public identifiers and addresses were sent.

Definitions of ref. 34 (analysis 51). The PDF file and the record page of this article in an institutional repository (UCL Discovery) were requested, and the text of the PDF was extracted. Its version and bibliographic data were compared with the Crossref record, and the passages that define the 95% recall point, normalized WSS at 95% recall, and $WSS@95\%$ for fewer than 20 included records were compared with the definitions of the Outcomes and Supplementary Note 19; the arithmetic equivalence of the two roundings of the 95% recall point was checked exactly for 1 to 100,000 included records. Only public addresses were sent.

PubMed was searched through E-utilities on 25 September 2026 without language restrictions with the string ("large language model"[tiab] OR "LLM"[tiab] OR "LLMs"[tiab] OR "GPT-4"[tiab] OR "ChatGPT"[tiab] OR "generative artificial intelligence"[tiab]) AND ("systematic review"[tiab] OR "evidence synthesis"[tiab] OR "meta-analysis"[tiab]) AND ("screening"[tiab] OR "study selection"[tiab] OR "citation screening"[tiab] OR "abstract screening"[tiab]) AND ("2022/11/30"[dp] : "2026/09/25"[dp]), which returned 290 records (291 when the search was re-run on 26 September 2026). Without the date clause the search returned 292 records on 26 September 2026, of which one was dated before 30 November 2022. The subset search added AND ("active learning"[tiab] OR "prioriti\*" [tiab] OR "stopping"[tiab]) and returned 32 records on both dates, none dated before 30 November 2022. arXiv was searched through its programming interface on 25 September 2026, without a date limit, with abs:"systematic review" AND abs:screening AND abs:"language model", which returned 44 records on both dates; the arXiv interface does not accept the field syntax of PubMed, so this query is not equivalent to the PubMed search. The search window began on 30 November 2022, the date of the public announcement of ChatGPT, because the search targeted screening with LLMs; the earlier literature on machine learning for screening and on stopping rules is cited in the Introduction. One preprint (ref. 15) was identified outside these recorded searches. The file of the 32 records saved by the original analysis held 31 records, because the script kept journal articles only and omitted one book record (a rapid review and evidence map on machine learning tools for evidence synthesis); the count of 32 is that of the search. Post hoc (analysis 45), nine further references were verified in Crossref, PubMed, arXiv, the ACL Anthology, PubMed Central, and on the pages of the publishers, and the numbers quoted from ref. 18 were checked against its PubMed abstract (the full text could not be retrieved). One PubMed search, fixed before it was run, sought a published per-record time of human title and abstract screening; it was run on 28 September 2026 and returned nine records, none of which reported such a time. The rate used for the illustrative conversion of records not read into reviewer time in the Results was identified outside this search and checked in the full text of its source (ref. 24), which states it without a citation (post hoc, analysis 50).

##### **Supplementary Note 23. English translation of the analysis plan and of the memo of 25 September 2026**

The analysis plan (Methods, Study design) is an internal, time-stamped, unregistered plain-text file written in Korean (PREREG.md; created at 18:03:53 on 24 September 2026 and last modified at 13:14:44 on 25 September 2026). The memo of 25 September 2026 is a separate file (PREREG.md in a separate folder; created at 09:48:43 on 25 September 2026, after the held-out decision of 04:53:44). This note gives the complete English translation of both files, in the order of the files. The translation was made on 26 September 2026 with the AI coding tool used for the analyses (Methods, Provenance of model inputs and judgments); one phrase of the last entry was revised on 27 September 2026, and typography was later adapted to the journal style; the Korean originals are the authoritative text. The translation departs from the files only where personal data are withheld, each time marked in place: the e-mail address of the corresponding author (one occurrence in the plan file) and the personal name of a third party, by which the files refer to the pilot review (three occurrences in the plan file and one in the memo). Bold type of the original is not reproduced, and commands and file names are unchanged. The times in the entries are those written in the file; its last entry verified them against the creation times of files, and Supplementary Note 1 gives the verified times.

Sizes and hashes. The original plan file has 9875 bytes and the SHA-256 hash 7cd0d3105748a279d21aa6760b96de80efefbe9810828744c9ef15a8b3502164, and the original memo has 515 bytes and the hash d719cce85fc1ebba1c36c309d12bc8ddcc867418453f6e3580554dc7522059c1. The redacted copies, which are available from the corresponding author on request, differ from the originals only in the marked replacements: the plan file has 9983 bytes and the hash 438d9076e4edbbb9d7f7d4e7c816bfcb3e23257e2ec7ae7d50a7bc95a447da54, and the memo has 546 bytes and the hash 611969efd6a938e23e80ded844afe4616e3836a231a4ebcf49fa528ce6d9eb09. The hashes of the redacted copies were computed on 4 October 2026, and those of the original files were recomputed on the same day.

#### Plan file

Preregistration: Jev screening benchmark (iteration 1)

Written 2026-09-24 18:10 EDT. This document was written before any model result on SYNERGY records had been seen. Already seen: results on the 1572 records of the [personal name withheld pending consent] review (Jev AUC 0.943, Claude Opus 0.952, and, on 544 records, DeepSeek 0.904 and GPT-4o-mini 0.882), the batch-size check (batches of five: AUC 0.941, rank correlation with single requests 0.970), and SYNERGY metadata (114 reviews, split 91/23). When this document is modified, the date and the reason are recorded in the change log below. The decision criteria are not changed after results have been seen.

#### Data

• SYNERGY 3.0 (`python -m synergy_dataset get, CC0`). 114 reviews, 168,969 records, 4399 final inclusions.
• Label: `label_included` (final inclusion). Reviews with title and abstract stage labels (57) are for secondary analysis.
• Split: the official split of the package. Methods are decided only on the 91 development reviews (train), and the 23 evaluation reviews (test) are run only once after the methods have been fixed.
• Sent to the models: review title, research question, eligibility criteria (original text of the SYNERGY metadata), record title and abstract. Labels are not sent.

#### Methods (fixed)

• M1 Jev zero-shot: `run_screen.mjs`, batches of five, question "probability of advancing to full-text screening". The prompt uses the same template as for the [personal name withheld pending consent] review.
• M2 free zero-shot baselines: BM25 (criteria plus question as the query), dense-embedding cosine similarity, cross-encoder rerankers. All local, cost 0.
• M3 LLM zero-shot (subset): GPT-4o-mini (token log-probability $P(\text{yes})$ if possible, otherwise verbalized probability), DeepSeek V3.1, Claude Opus 5.5 (subscription).
• M4 active learning (ASReview style): TF-IDF plus logistic regression, certainty sampling, starting from one random included and one random excluded record. Mean of ten random starts.
• M5 hybrid: Jev scores are used as the starting order and as a feature of active learning. The detailed design is decided only on the development split.
• S1 stopping rule: the number of remaining inclusions is estimated from calibrated Jev probabilities. Calibration uses only a random pilot sample per review.

#### Metrics

• Per-review AUC; $\text{WSS@95} = 1 - (\text{number read to reach 95\% recall})/N - 0.05$; position of the last included record ($\text{WSS@100}$); recall at 10%, 20%, and 30% read.

• Macro-average over reviews. CIs by bootstrap resampling of reviews. Methods are compared with the paired Wilcoxon test over reviews.

##### Decision criteria (judged on the 23 reviews of the evaluation split)

• C1 zero-shot quality: the macro AUC of Jev is at least 0.03 higher than that of the best free zero-shot baseline, and Wilcoxon $P < 0.01$.
• C2 versus LLMs: on the common subset, the macro AUC of Jev is not lower than that of GPT-4o-mini (non-inferiority margin 0.02), and the difference from Claude Opus is within 0.03. Cost per 1000 records is presented.
• C3 hybrid: the WSS@95 of the hybrid is on average at least 5 percentage points higher than that of active learning alone, and Wilcoxon $P < 0.05$. The position of the last included record also improves.
• C4 stopping: the Jev stopping rule achieves recall of at least 0.95 in at least 90% of the evaluation reviews and does less work than a baseline stopping rule of the same reliability.
• Publication decision: if (C1 or C2) and (C3 or C4) hold on the evaluation split and the rank correlation is at least 0.9 for two rephrasings of the prompt, the result is regarded as "a solid result worth submitting to a journal with an impact factor of 5 to 6" and is reported to [e-mail address withheld]. If any of these is missing, the next iteration is planned.

##### Known limitations (written down in advance)

• The training cut-off of Jev is not disclosed. The possibility that it was trained on the inclusion lists of published reviews is stated as a limitation common to all LLM screening studies.
• Claude is called headless under a subscription, so the temperature cannot be set.
• Because of the gateway limit (about 30 requests per minute), the LLM comparison is made only on a subset.

##### Change log

• (none)
• 2026-09-24 18:40: the batch size of M1 was changed from 5 to 10. Reason: batches of ten also passed the prior criterion on the 1572 records of [personal name withheld pending consent] (AUC 0.941 versus 0.943 for single requests, rank correlation 0.964), and with half the number of requests the throughput under the gateway limit (about 30 requests per minute) is doubled. Decided while no SYNERGY result had yet been seen.
• 2026-09-24 19:05: ambiguous parts of the decision criteria were clarified. No result of the evaluation split has yet been seen (the runs of Claude, GPT-4o-mini, and DeepSeek on the evaluation subset are in progress, but their result files have not been opened). The results of 25 small reviews of the development split have been seen.
• (sub-item) The "best free baseline" of C1 is fixed in advance as the one free method with the highest macro AUC over the whole development split, and on the evaluation split the comparison is made with that one only.

• (sub-item) Non-inferiority in C2: the lower limit of the 95% CI (resampling of reviews) of the difference in macro AUC (Jev – GPT-4o-mini) $> -0.02$. "Within 0.03 of Claude": point estimate of (Claude – Jev) $\leq 0.03$. GPT-4o-mini is compared with the token log-probability P(Yes) method (a continuous score). Because the limit of the free team plan of the gateway is five requests per minute per model and provider, the comparison is made only on an evaluation subset of 2002 records (all 597 inclusions plus random exclusions in proportion to each review).
• (sub-item) The comparator of C3 is, as preregistered, LR active learning, starting from one random inclusion plus one exclusion, mean of ten seeds. The details of the hybrid method (the weight $\lambda$ and so on) are fixed as the single variant with the highest macro-average WSS@95 over the whole development split. NB active learning is reported as well.
• (sub-item) The Jev stopping rule of C4 is "read only the records with Jev probability $\geq \tau$ and stop" (no labels needed). $\tau$ is fixed as the largest value at which the proportion of reviews of the development split that achieve recall of at least 0.95 is at least 0.95. The baseline stopping rule is the knee method (Cormack & Grossman 2016) on the order of LR active learning. C4 holds = on the evaluation split, the reliability of the Jev rule (the proportion of reviews with recall $\geq 0.95$) $\geq 0.90$, and the reliability is at least that of the knee method while the mean workload is smaller than that of the knee method.
• (sub-item) Robustness: for each of the question wordings q1 and q2, on a development subset, the mean per-review Spearman correlation with q0 $\geq 0.90$, and the difference in macro AUC $\leq 0.02$.
• 2026-09-25 00:50: disclosure. When `zs_baselines.py test` finished, it printed to the log a summary of the macro AUCs of the free baselines on the evaluation split (tfidf 0.717, bm25 0.712, minilm 0.821, and so on) and one line for Walker\_2018, and these were seen. The results of Jev on the evaluation split have not been seen. The comparator of C1 has already been fixed as "the best free method on the development split", so the decision procedure does not change.
• 2026-09-25 01:00: methods fixed (before seeing the Jev results on the evaluation split). Based on the 73 reviews of the development split.
• (sub-item) C1 comparator = bge-base (development macro AUC 0.847, the highest among the free methods). Jev on development 0.945.
• (sub-item) C3 hybrid = `asr_jeblend_3` ($\lambda=3$). Development WSS@95: $\lambda=3$ 0.7662, $\lambda=2$ 0.7657, $\lambda=1$ 0.7309, feature concatenation 0.7488. By the rule, the highest value, $\lambda=3$.
• (sub-item) C4 $\tau = 0.07$ (proportion of development reviews with recall $\geq 0.95$: 0.986; $\tau=0.10$ gives 0.918, below the criterion).
• (sub-item) Decision command for the evaluation split: `final_eval.py test --best-free bge --lam 3 --tau 0.07`

##### Post-hoc analyses (started 2026-09-25, exploratory)

These analyses were added after the decision on the evaluation split (C1 to C4). The main conclusion is drawn only from the decision above, and the analyses below are marked as exploratory in the manuscript.

• P1: LLM pseudo-label active learning (the medRxiv 2025 method) re-implemented with the same Jev scores and compared with the hybrid ($\lambda=3$). T and B of the pseudo-labels are chosen only on the development split.
• P2 batch effect: on the 2002 records of the evaluation subset, comparison of single requests, batches of ten within the subset (prevalence about 30%), and the batches of ten of the original full run (prevalence about 1.8%).
• P3: re-analysis in the reviews that have title and abstract stage labels (`label_abstract_included`).
• P4: sensitivity analysis against a stronger ASReview model.
• P5: analysis of the failure types of included records that Jev scored low.
• P1 fixed (2026-09-25 10:10): on the grid of the development split (T 1 to 30%, B 50 and 80%), the best pseudo-label setting = T 10%, B 50% (WSS@95 0.734; on the same development split the hybrid $\lambda=3$ gives 0.766). Applied to the evaluation split as `asr_pseudo_10_50`.
• 2026-09-25 13:40: correction of an omission in the record (disclosure of a protocol deviation). The entry of 2026-09-24 19:05 named "LR active learning" as the comparator of C3, but from 19:06 the development split was run with the default model of ASReview 3.0.8 (elas\_u4: TF-IDF + LinearSVC) (`asr_dev.json` created 19:06:29), and the decision on the evaluation split was also made with this model (`asr_prior`). The decision command in the entry of 01:00 (methods fixed) points to this model, but the change of comparator itself was not stated. The Jev results of the evaluation split were first seen on 2026-09-25 at 04:53 (`final_test.json`), so the change preceded the results. The comparators as literally preregistered (LR and NB active learning, ten seeds) are also computed on the evaluation split and reported as a sensitivity analysis (`al_test_prereg.json`).
• 2026-09-25 13:13: the time "13:40" of the preceding entry was written in error. The actual time of writing, according to the file modification time, is 2026-09-25 13:11. The content remains valid.
• 2026-09-25 13:14: verification of the time labels (correction). Some of the times in the entries above were written as approximations by the writer of the entries, so they were verified against the creation times of files produced by the same command. First writing "18:10" → actually 18:03 (`PREREG.md` created 18:03:53). Batch change "18:40" → actually 18:17 (`run_jev_dev.log`, started by the same command, created 18:17:47). Clarification of the decision criteria "19:05" → actually 19:00 to 19:02 (the last line of the progress record in the output of the same command is 19:00). The disclosure "00:50" and the fixing of methods "01:00" agree with the supporting files (`asr_dev_full.log` created 00:50:40, completed 00:58:40). Pseudo-label fixing "10:10" → actually 10:06 (`asr_test_p1.log` created 10:06:45). The order of all entries is preserved. The first SYNERGY model output was at 2026-09-24 18:18:15 (`jev_dev2000.json` created), and the Jev results of the evaluation split were first viewed on 2026-09-25 at 04:53 (`final_test.json`).

#### Memo of 25 September 2026

• P6 external validation in clinical reviews: public data of the CLEF eHealth TAR task (Cochrane reviews). Eligibility criteria are taken only from the Objectives and Selection criteria sections (primary source) of the abstract of each Cochrane review. Methods are applied as fixed on SYNERGY (batches of ten, $\lambda=3$, $\tau=0.07$) without re-tuning.
• P7 conclusion-level impact: in the [personal name withheld pending consent] review, the included studies that would have been missed under the fixed rules ($\tau=0.07$, hybrid plus knee) are removed and the main quantitative results of the review (direction sign tests by domain, numbers of genes, and so on) are recomputed.

#### Supplementary Note 24. TRIPOD-LLM checklist

TRIPOD-LLM (ref. 42) items applicable to an evaluation study of existing models for a classification task. M=main article; SN=Supplementary Note; ST=Supplementary Table. The same checklist is supplied as a separate file.

| Item | Topic | Where reported |
| --- | --- | --- |
| 1 | Title identifies evaluation, task, and outcome | M, title |
| 2 | Abstract | M, Abstract |
| 3a | Context, rationale, existing approaches | M, Introduction (including the literature search summarized there and described in Methods, Literature search) |
| 3b | Target population and intended use | M, Introduction, Discussion, and Methods, Stopping rules (review teams; ordering of records within a human-supervised workflow; the threshold was evaluated as a stopping rule and is not supported as a stand-alone stopping rule for recall-critical reviews, a labeled stopping check is supported where recall is critical, and stopping at the threshold where some loss of recall is acceptable was not evaluated; all evaluations used criteria taken from completed reviews; $\tau$ was derived under one unrecorded model version and one batching scheme and should not be applied at the screening stage of a new review without re-derivation; $\tau$ was evaluated only for ten-record requests formed from the records of one review in an order not sorted by relevance or by any score, and it should not be applied to requests formed from a relevance-sorted export); SN3 (Use of $\tau$: in this study the order was that of the data files, which did not cluster positive records beyond random order within reviews, and how the source files were ordered is not documented; the export order of bibliographic databases or reference managers may follow a relevance ranking, which is a form of pre-ranking that was not evaluated; random shuffling within a review was not itself evaluated; the effect of request composition on scores was not isolated from differences between scoring occasions and providers) |
| 4 | Objectives (evaluation of existing models) | M, Introduction, final paragraph |
| 5a | Data sources for tuning and evaluation | M, Methods, Study design and Data sources (pilot review, on which the prompt formats, batch size, and free-ranker shortlist were set; development SYNERGY+ reviews, on which the free-ranker comparator, $\lambda$, and $\tau$ were fixed; held-out SYNERGY+ reviews; CLEF 2019 reviews used post hoc); SN1 (hybrid variants and stopping rules simulated on the pilot labels before the threshold rule was chosen) |
| 5b | Description of data | M, Table 1; M, Methods, Data sources (SYNERGY+ data are the subset exported by synergy-dataset 2.2: open-access records with an abstract, 35.3% of the records and 41.2% of the included studies of the release for the 114 reviews); SN4 (share retained per held-out review, ST8) |
| 5c | Dates of oldest and newest text | Evaluation texts: the scored SYNERGY+ reviews were published in 2011–24 (two reviews of the dataset published in 2025 were not scored; ST46). CLEF 2019 Cochrane reviews: the end date of the search of each review ranged from 24 February 2015 to 23 January 2019 (one review was limited to publication years 2013–16), and the Cochrane abstracts from which criteria were taken were published in 2017–19. Pilot review: search to 18 June 2026. The vendor of Jev does not disclose training data or a knowledge cut-off (M, Methods, Model, access, and provenance); the training cut-off dates of the three LLMs were not recorded |
| 5d | Preprocessing and quality checks | M, Methods, Data sources |
| 5e | Missing and imbalanced data | M, Table 1 (inclusion rates; records without an abstract scored from the title); M, Methods, Prompts and input formatting (record block for records without an abstract) and Data sources (the SYNERGY+ export contains only records with an abstract) |
| 6a | Model name, version, last training date | M, Methods, Model, access, and provenance (access dates, identifiers, release status, version that served the requests not recorded, the two reference models of the vendor's evaluation); SN1 (versioned identifier documented by the vendor) |
| 6b | Development process | Not applicable (existing models); developer information cited; M, Methods, Model, access, and provenance |
| 6c | Prompting and inference settings | M, Methods, Prompts and input formatting (full prompts, temperature, output-token limits, batching); Supplementary Notes 3 (composition of requests) and 15 (LLM subset) |
| 6d | Raw and processed outputs | M, Methods, Prompts and input formatting (probability-valued scores; log-probability conversion for GPT-4o-mini) |
| 6e | Classification thresholds | M, Methods, Stopping rules (grid and rule for $\tau$; $\tau$ fixed on development reviews); SN12 |
| 7a | Quality metrics for generative outputs | Not applicable (probabilities only) |
| 7b | Relevance of metrics to deployment | M, Methods, Outcomes (WSS, recall, workload) |
| 7c | Outcome definition, computation, inference dates | M, Methods, Outcomes, Statistical analysis, and Model, access, and provenance |
| 7d | Subjective outcome assessment | Labels from the original review teams. The failure classification (SN7) and the coding of prior evidence (SN22) were proposed with the AI coding tool used for the analyses (M, Methods, Provenance of model inputs and judgments) and are reported only in the Supplementary Information. The main article states that the CLEF content-level label was not checked against the included-study lists of the published reviews (M, Methods, Data sources, and Discussion); the study-level reading of the CLEF content-level label in three Cochrane reviews is reported in SN7 |
| 7e | Comparison with other models and benchmarks | M, Methods, Comparators; Table 2; Fig. 2; SN15 (non-equivalent elicitation; two-stage bootstrap and calibration of the LLMs, post hoc); SN6 (biomedical cross-encoder, post hoc); ST24 (knee method with the published minimum, post hoc); M, Table 3 (stopping options, post hoc except the threshold and the knee method as adapted on the held-out reviews) |
| 8a–8c | Annotation | No new annotation; labels from the original reviews (SYNERGY+, CLEF 2019, pilot review team) |
| 9a–9b | Prompt design and data used | M, Methods, Prompts and input formatting (full prompts, settings, and the two rephrasings) and Study design (prompt formats, batch size, and free-ranker shortlist set on the pilot review); SN1 (times and content of these steps; hybrid variants and stopping rules also simulated on the pilot labels before the threshold rule was chosen) |
| 10–11 | Summarization; instruction tuning | Not applicable |
| 12 | Compute | Costs in M, Results, and SN2; throughput in ST5; simulations on one server with two 12-core Intel Xeon Gold 6126 processors (SN19) |
| 13 | Ethical approval | M, Methods, Ethics: not required (public bibliographic records and public screening labels; screening decisions and extraction data of a systematic review by our group; no human participants or individual-level human data). |
| 14a–14b | Funding; conflicts of interest | M, Acknowledgements; Competing interests |
| 14c–14d | Protocol; registration | M, Methods, Study design (analysis plan; decision criteria fixed before the held-out evaluation; the order of the steps is stated there and timed in SN1); SN1 (numbered list of the departures from the plan); SN23 (internal, time-stamped plan file; complete English translation of the plan and of the memo of 25 September 2026, with the sizes and SHA-256 hashes of the original files and of the redacted copies); not registered in a public registry |
| 14e–14f | Data and code availability | M, Data availability and Code availability (re-querying the model may not reproduce the scores because Jev reports no version; the deposited per-record responses are the reproducible record of the primary analysis; Zenodo, ref. 43); M, Methods, Model, access, and provenance |
| 15 | Public involvement | No patient or public involvement |
| 16a–16d | Patient or EHR data | Not applicable |
| 17 | Performance on prespecified metrics | M, Results; Table 2; post-hoc results are marked as such, and *P* values other than those of C1 and of the WSS@95 comparison of C3 are exploratory (M, Methods, Statistical analysis) |
| 18 | Model updating | Not applicable |
| 19a–19b | Interpretation; limitations | M, Discussion (limitations in the Discussion prose, including criteria taken from completed reviews, the export restriction of SYNERGY+, and the reference standard of the Cochrane reviews) |
| 19c–19f | Usability in context | M, Methods, Stopping rules and Prompts and input formatting ($\tau$ derived from ten-record requests formed in the order of the data files, which was not sorted by relevance or by any score); SN12 (re-derivation of $\tau$ if the batching changes or if drift monitoring suggests that the model has changed, because no version is reported); M, Discussion (use within a human-supervised workflow; labeled stopping check where recall is critical; $\tau$ evaluated only for requests formed from the records of one review in an order not sorted by relevance or by any score, not for relevance-sorted exports; $\tau$ should not be applied in a new review without re-derivation on labeled reviews; archiving of scores and prompts and monitoring of drift by rescoring an archived set of records, because no model version can be reported; implementation risks of an early-access product); SN3 and ST35 (effect of request composition on scores not isolated from differences between scoring occasions and providers) |
| 19g | Next steps | M, Discussion (prospective studies) |
| Journal policy | Use of AI tools in the research process and in manuscript preparation | M, Methods, Provenance of model inputs and judgments (AI coding tool used for the model inputs, the analyses, and the judgments reported in the Supplementary Information); M, Methods, Use of AI tools |

### Supplementary Fig. 4 Distribution and calibration of Jev scores (post hoc).

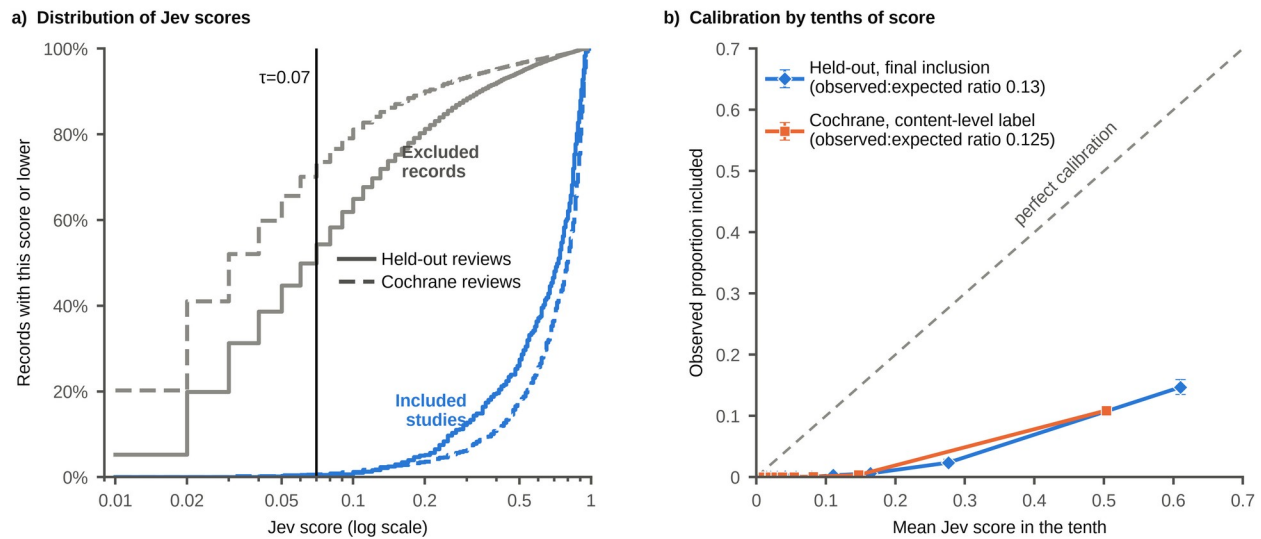

a) Cumulative distribution of Jev scores of included studies (blue) and excluded records (grey), pooled over the 23 held-out reviews (solid lines; 597 included studies and 32,404 excluded records) and the 31 Cochrane reviews (dashed lines; 882 content-relevant and 81,536 other records). Scores are reported in steps of 0.01; scores below 0.01 are counted at 0.01. The vertical line marks $\tau=0.07$. b) Observed proportion of included studies against the mean score in tenths of the pooled records ordered by score, with Wilson 95% CIs, for the 23 held-out reviews (final inclusion) and the 28 Cochrane reviews with a content-relevant record (content-level labels); the dashed line marks perfect calibration. The observed-to-expected ratios and their CIs are given in Supplementary Note 9. The Wilson intervals of the pooled tenths do not account for the clustering of records within reviews.

This figure was Figure 6 of the main text in an earlier version and is placed in the Supplementary Information to respect the limit on display items; it is cited in the Results (Calibration).
